# Time to Hyperkalemia and Other Outcomes Post-Implementation of the 2021 CKD-EPI Creatinine Equation: An Observational Cohort Study

**DOI:** 10.1101/2024.05.21.24307717

**Authors:** Charlotte Baker, Brianna M. Goodwin Cartwright, Samuel Gratzl, Duy Do, Patricia J. Rodriguez, Michael Simonov, Nicholas L Stucky

**Author notes:** Corresponding author: Nicholas Stucky, MD, PhD Truveta, Incorporated, 1745 114th Ave SE, Bellevue, WA 98004.

## Abstract

**Background:** Timely diagnosis can delay progression to poor clinical outcomes and biomarker endpoints in CKD. Our objective was to analyze how the implementation of the 2021 CKD-EPI Creatinine Equation (CKD-EPI 2021) affected time to the doubling of serum creatinine, prescription or dispense of potassium-lowering drugs, hyperkalemia, and creation of arteriovenous graft fistula for dialysis within one year of CKD diagnosis.

**Methods:** We used retrospective EHR data to create two cohorts of adult patients based on date of diagnosis. We followed patients from their CKD diagnosis to the occurrence of each outcome, their last medical encounter, or 1-year post diagnosis, whichever occurred first. The first cohort was diagnosed in 2021 when the CKD-EPI 2009 Creatinine Equation (pre-update cohort) was recommended. The second cohort was diagnosed in 2022 when CKD-EPI 2021 (post-update cohort) was recommended. Multivariable models for the time relationship between cohort and each outcome were adjusted for demographics, social determinants of health, and comorbidities. As CKD-EPI 2021 was race-free, we also considered the interaction between cohort and race.

**Results:** We found 261,774 patients with a first-time CKD diagnosis. After implementation of CKD-EPI 2021, patients were less likely to have a diagnosis of hyperkalemia, potassium-lowering drug prescription, a doubling of serum creatinine, or an arteriovenous graft fistula placement within a year of CKD diagnosis. Black patients in the post-update cohort were less likely to be diagnosed with hyperkalemia compared to Black patients in the pre-update cohort (AHR 0.83; CI 0.80, 0.86), but Black patients overall were significantly more likely than non-Black patients to have hyperkalemia.

**Conclusions:** Time to poor clinical outcomes and biomarkers differs by the date of CKD diagnosis. Future work should elucidate the mechanisms driving these differences, whether there have been significant changes in treatment practices since the CKD-EPI 2021 recommendation, and long-term effects of each outcome.

## Introduction

Chronic kidney disease (CKD) is a major public health issue. In the Americas, CKD was the 8^th^ leading cause of death and the 10^th^ leading cause of years of life lost as of 2019.(1) CKD affects 15% of the US population (37 million adults); 100.6 people per 1,000 population die annually.(1,2)

Timely diagnosis enables treatment and can delay progression. Suggested clinical guidelines recommend annual screening for people with risk factors for CKD such as hypertension, diabetes, or being over 50 years of age.(3) The National Kidney Foundation and the American Society of Nephrology Task Force on Reassessing the Inclusion of Race in Diagnosing Kidney Disease recommend estimating GFR using the race-free 2021 Chronic Kidney Disease Epidemiology Collaboration (CKD-EPI) Creatinine Equation (CKD-EPI 2021).(4)

Black or African-American patients (Black) are four times more likely to be diagnosed with end-stage kidney disease (ESKD) than white patients.(5) Hispanic patients are twice as likely to be diagnosed compared to non-Hispanic or Latino white patients. Like differences in diagnosis, ESKD treatment and outcomes vary by demographics and social determinants of health (SDOH) such as age, race, geography, and access to care.(6,7)

Sixty-six percent of hemodialysis patients have an arteriovenous graft fistula (AVF), the gold standard method for providing hemodialysis.(8–10) Yet, many patients have significant morbidity prior to needing dialysis including having an increased risk of hyperkalemia; medications to treat CKD and comorbidities (e.g. diabetes, hypertension) can induce this condition.(10–14) Potassium-lowering drugs such as patiromer can be used alongside other medicines to reduce blood potassium and lower cardiac risk.(10,15) A doubled serum creatinine value is an accepted biomarker for a significant negative change in GFR, a sign of worsening CKD.(16–18) This endpoint helps focus clinical trials and treatments for patients and those with comorbidities.

CKD-EPI 2021 uses age and sex to help estimate GFR.(4) Prior to this equation, race (dichotomized as Black and non-Black) was included as a marker for a presumed higher muscle mass in Black people.(19,20) The use of CKD-EPI 2021 has resulted in more severe especially for Black patients.(21) Although we know there were changes in time-to-diagnosis with the implementation of CKD-EPI 2021, it is unknown if implementation was also associated with a change in time from diagnosis to poor clinical outcomes and biomarker endpoints. Further, it is unknown if these differences are dependent on race.

This study examines the association between the implementation of the CKD-EPI 2021 and time from CKD diagnosis to each of the following outcomes: hyperkalemia, prescription or dispense of a potassium-lowering drug, doubling of serum creatinine, and AVF placement. Furthermore, the study investigates whether and how these associations vary by race.

## Methods

### Data source

This study used a subset of Truveta Data. Truveta provides access to continuously updated and linked electronic health record (EHR) from a collective of US health care systems, including structured information on demographics, encounters, diagnoses, vital signs (e.g., weight, BMI, blood pressure), medication requests (prescriptions), medication administration, medication dispense (fills), laboratory and diagnostic tests and results, and procedures. Updated EHR data are provided daily to Truveta by constituent health care systems. In addition to EHR data, SDOH information are made available through linked third-party data. Data are normalized into a common data model through syntactic and semantic normalization. Truveta Data are then de-identified by expert determination under the HIPAA Privacy Rule. In accordance with 45 C.F.R. § 46.101 Protection of Human Subjects, our study did not require Institutional Review Board approval because it used only deidentified medical records. Data for this study were accessed on February 6, 2024.

### Study population

We included patients first diagnosed with CKD at or after January 1, 2021 (using ICD-9, ICD-10, and SNOMED CT codes; see supplement). Patients were categorized into two mutually exclusive cohorts based on their first date of diagnosis. The first (pre-update cohort) was diagnosed between January 1, 2021 and December 31, 2021 (inclusive). These patients were diagnosed when the race-based 2009 CKD-EPI Creatinine Equation (CKD-EPI 2009) was recommended. The second cohort (post-update cohort) was diagnosed between January 1, 2022 and December 31, 2022 (inclusive), when CKD-EPI 2021 was recommended. Patients in both cohorts were followed from their index diagnosis of CKD until the first occurrence of the outcome of interest (each outcome was studied separately), the last medical encounter recorded in the data, or the end of the 1-year follow-up period, whichever occurred first.

### Inclusion/Exclusion

We included adult patients aged 18 years and older at the time of their index CKD diagnosis. Patients must have interacted with the health care system within 36 months prior to their CKD diagnosis. We dichotomized race (Black and non-Black) to correlate with CKD-EPI 2009 race categories. All white, Asian, Native Hawaiian and Pacific Islander, American Indian and Alaska Native, and ‘other’ patients were categorized as ‘Not Black or African-American’ (non-Black). Unknown race and ‘choose not to answer’ were excluded.

Three outcomes had specific exclusion criteria independent of all other outcomes. These included: patients with a history of hyperkalemia at or at least one day before CKD diagnosis (time to hyperkalemia); patients that had a prescription or dispense of a potassium lowering drug at or at least one day prior to CKD diagnosis (time to potassium lowering drugs); and evidence of a procedure or condition for AVF at or at least one day prior to CKD diagnosis (AVF placement).

Patients were excluded if they had a diagnosis of renal failure, polycystic kidney disease, or acute kidney injury (AKI) (defined using ICD-10 or SNOMED billing codes) at any time prior to their CKD diagnosis. AKI patients were excluded primarily due to 1) the potential for AKI to occur because of CKD or for CKD to occur because of AKI (high collinearity); and 2) the possibility of AKI patients to be sicker than other patients that develop CKD.(22–24) We were unable to discern these issues accurately in the data.

### Study Outcomes

We examined the time between the first CKD diagnosis and the first occurrence of four independent outcomes. For all outcomes, we calculated time to event by finding the difference between CKD diagnosis and the first time the outcome occurred. All ICD-9, ICD-10, SNOMED CT, RXNORM, CPT, and LOINC codes used to define outcomes are available in the supplement.

- Hyperkalemia

`o` We identified the first hyperkalemia diagnosis after CKD diagnosis using laboratory values (LOINC codes).
- Prescription of potassium-lowering drugs

`o` We identified the first date of either a prescription or dispense of a potassium binder (i.e., patiromer, sodium zirconium cyclosilicate, sodium polystyrene sulfonate, and calcium polystyrene sulfonate product) after diagnosis using RXNORM codes.
- Doubling of serum creatinine

`o` We identified the first non-hospitalization related serum-creatinine laboratory value for each patient at or after their CKD diagnosis using LOINC codes. We allowed serum creatinine values on the day of diagnosis to be included to have baseline value as close to diagnosis as possible. We identified the first date the serum creatinine value was exactly or more than twice the baseline.
- AVF placement

`o` To estimate time to dialysis, we identified the first AVF placement after CKD diagnosis using diagnosis (SNOMED CT) and procedure (CPT) codes.

### Statistics

Unadjusted Kaplan-Meier curves stratified by cohort and race were used to describe the probability of the outcome within one year of CKD diagnosis. We fit Cox-proportional hazards models assessing the relationship between cohort and time to event adjusting for demographics, SDOH, and comorbidities present at least one day prior to CKD diagnosis. We extended these models to include an interaction term between cohort and race to assess disparities in the time to outcome. A two-sided p-value < 0.05 was considered statistical significance. We used Python 3.10.13 to clean the data and R 4.2.3 for statistics.

### Covariates

We created binary variables (yes/no) for obesity (BMI≥30), type 2 diabetes, chronic hypertension, and hyperlipidemia. We used the Elixhauser Comorbidity Index with van Walraven weights as a summary measure of disease burden.(25,26) We excluded renal disease from the list of comorbidities included in index.(26) For the interpretation of the Elixhauser Comorbidity Index, weighted scores of less than 11 were categorized as “low comorbidity”, 11 to 15 were categorized as “medium comorbidity”, and greater than 15 were categorized as “high comorbidity”.(27) Comorbidities were defined using ICD-9, ICD-10, and SNOMED CT codes (supplement).

Demographic variables included age (18 – 44, 45 – 64, and 65+ years), sex (female and male), and ethnicity (Hispanic or Latino, not Hispanic or Latino). Four SDOH variables were included. Income impacts health processes and outcomes including access to care. Annual patient income was categorized into 6 groups: $0 – 30,000, 30,001 – 50,000, 50,001 – 80,000, 80,001 – 100,000, 100,001 – 120,000, and more than $120,000.

As a marker of housing stability, we included the number of address changes in the last year (0, 1, and 2 or more). We included two social and familial support determinants. First, we included distance to a family or other close connection; categories were < 25 miles away, 25 to 100 miles away, 100 to 500 miles away, or > 500 miles away. Patients with no information on the proximity of their closest tie were categorized as ‘no information’. Second, if a patient was divorced, widowed, in a domestic partnership or civil union, or married their marital status was categorized as ‘ever married or partnered’. Patients that were unmarried or never married were categorized as ‘unmarried or never married’; patients whose status was unknown or did not fit in these categories was categorized as ‘other’.

## Results

We identified 261,774 patients with a first-time CKD diagnosis. Black patients accounted for 15% of the population (Table 1). Within a year of diagnosis, 140,995 (54.0%) had a diagnosis of hyperkalemia, 5,283 (2.0%) patients had a prescription of a potassium-lowering drug, 2,446 (0.9%) had a doubling of serum creatinine, and 330 (0.1%) had an AVF.

**Table 1.** Descriptive statistics of patients with poor outcomes within 1 year of chronic kidney disease diagnosis by Recommended CKD-EPI Creatinine Equation.

| **Characteristic** | **Overall,**<br>**N = 261,774** | **CKD-EPI 2009 (Pre-**<br>**update),**<br>**N = 87,903** | **CKD-EPI 2021 (Post-**<br>**update),**<br>**N = 173,871** |
| --- | --- | --- | --- |
| Race, n (%) |  |  |  |
| Not Black* | 222,647 (85%) | 76,038 (87%) | 146,609 (84%) |
| Black* | 39,127 (15%) | 11,865 (13%) | 27,262 (16%) |
| Age Group, n (%) |  |  |  |
| 18-44 | 11,748 (4.5%) | 4,041 (4.6%) | 7,707 (4.4%) |
| 45-64 | 55,848 (21%) | 18,877 (21%) | 36,971 (21%) |
| 65+ | 194,178 (74%) | 64,985 (74%) | 129,193 (74%) |
| Sex, n (%) |  |  |  |
| Female | 135,909 (52%) | 45,327 (52%) | 90,582 (52%) |
| Male | 125,865 (48%) | 42,576 (48%) | 83,289 (48%) |
| Ethnicity, n (%) |  |  |  |
| Hispanic or Latino | 20,481 (7.8%) | 6,960 (7.9%) | 13,521 (7.8%) |
| Not Hispanic or Latino | 227,193 (87%) | 76,307 (87%) | 150,886 (87%) |
| Unknown | 14,100 (5.4%) | 4,636 (5.3%) | 9,464 (5.4%) |
| Marital Status, n (%) |  |  |  |
| Unmarried and Never Married | 43,449 (17%) | 14,115 (16%) | 29,334 (17%) |
| Ever Married or Partnered | 200,434 (77%) | 67,363 (77%) | 133,071 (77%) |
| Other | 1,546 (0.6%) | 457 (0.5%) | 1,089 (0.6%) |
| Unknown | 16,345 (6.2%) | 5,968 (6.8%) | 10,377 (6.0%) |
| Distance in Miles to Closest Ties, n (%) |  |  |  |
| Less than 25 miles | 204,375 (91%) | 70,511 (91%) | 133,864 (91%) |
| 25 - 100 miles | 3,949 (1.8%) | 1,420 (1.8%) | 2,529 (1.7%) |
| 100 - 500 miles | 3,636 (1.6%) | 1,239 (1.6%) | 2,397 (1.6%) |
| Over 500 miles | 4,411 (2.0%) | 1,559 (2.0%) | 2,852 (1.9%) |
| No Information | 7,789 (3.5%) | 2,894 (3.7%) | 4,895 (3.3%) |
| Estimated Annual Income Range, n (%) |  |  |  |
| 0 - 30,000 | 20,338 (9.1%) | 6,897 (9.0%) | 13,441 (9.2%) |
| 30,001 - 50,000 | 99,636 (45%) | 34,607 (45%) | 65,029 (45%) |
| 50,001 - 80,000 | 71,460 (32%) | 23,958 (31%) | 47,502 (33%) |
| 80,001 - 100,000 | 17,761 (8.0%) | 6,565 (8.5%) | 11,196 (7.7%) |
| 100,001 - 120,000 | 8,396 (3.8%) | 3,087 (4.0%) | 5,309 (3.6%) |
| More than 120K | 4,849 (2.2%) | 1,812 (2.4%) | 3,037 (2.1%) |
| Number of Address Changes in Last 12 Months, n (%) |  |  |  |
| 0 | 201,368 (90%) | 69,776 (90%) | 131,592 (90%) |
| 1 | 19,349 (8.6%) | 6,668 (8.6%) | 12,681 (8.7%) |
| 2 or More | 3,446 (1.5%) | 1,179 (1.5%) | 2,267 (1.5%) |
| Elixhauser Comorbidity Index Score**, n (%) |  |  |  |
| 0- < 11 | 186,444 (71%) | 62,350 (71%) | 124,094 (71%) |
| 11 - 15 | 29,832 (11%) | 10,046 (11%) | 19,786 (11%) |
| > 15 | 45,498 (17%) | 15,507 (18%) | 29,991 (17%) |
| Chronic Hypertension (Yes/No), n (%) |  |  |  |
| No | 99,532 (38%) | 33,323 (38%) | 66,209 (38%) |
| Yes | 162,242 (62%) | 54,580 (62%) | 107,662 (62%) |
| Type 2 Diabetes<br>(Yes/No), n (%) |  |  |  |
| No | 141,577 (54%) | 47,255 (54%) | 94,322 (54%) |
| Yes | 120,197 (46%) | 40,648 (46%) | 79,549 (46%) |
| Hyperlipidemia<br>(Yes/No), n (%) |  |  |  |
| No | 73,074 (28%) | 24,970 (28%) | 48,104 (28%) |
| Yes | 188,700 (72%) | 62,933 (72%) | 125,767 (72%) |
| Obesity<br>(Yes/No), n (%) |  |  |  |
| No | 128,164 (49%) | 42,481 (48%) | 85,683 (49%) |
| Yes | 133,610 (51%) | 45,422 (52%) | 88,188 (51%) |
| Hyperkalemia, n<br>(%) |  |  |  |
| No | 120,779 (46%) | 34,538 (39%) | 86,241 (50%) |
| Yes | 140,995 (54%) | 53,365 (61%) | 87,630 (50%) |
| Potassium Drug<br>Prescription<br>Request, n (%) |  |  |  |
| No | 256,491 (98%) | 85,907 (98%) | 170,584 (98%) |
| Yes | 5,283 (2.0%) | 1,996 (2.3%) | 3,287 (1.9%) |
| Doubling of<br>Serum<br>Creatinine, n (%) |  |  |  |
| No | 259,328 (99%) | 86,868 (99%) | 172,460 (99%) |
| Yes | 2,446 (0.9%) | 1,035 (1.2%) | 1,411 (0.8%) |
| Fistula<br>Placement, n (%) |  |  |  |
| No | 261,444 (100%) | 87,750 (100%) | 173,694 (100%) |
| Yes | 330 (0.1%) | 153 (0.2%) | 177 (0.1%) |

\*Black refers to Black or African American Individuals
\*\*Elixhauser Comorbidity Index computed using van Walraven weights. Categories correlate to low, medium, and high comorbidity.

In Figure 1 and Figure 2, the scale for the outcomes of prescription of potassium lowering drugs (B) has a y-axis of 0% to 5% while doubling of serum creatinine (C) and placement of an AVF (D) and have a y-axis of 0% to 2% due to small probabilities. The probability of all outcomes over time differed across cohorts (Figure 1) and across cohorts by race (Figure 2). In Figure 1, we see patients in the post-update cohort were significantly less likely than patients in the pre-update cohort to have any of the four outcomes. Black patients in the pre-update cohort have a higher risk of being prescribed or dispensed a potassium-lowering drug than Black patients in the post-update cohort (Figure 2) but Black patients in both cohorts have a greater probability than their non-Black counterparts, especially as time increases.

**Figure 1.**
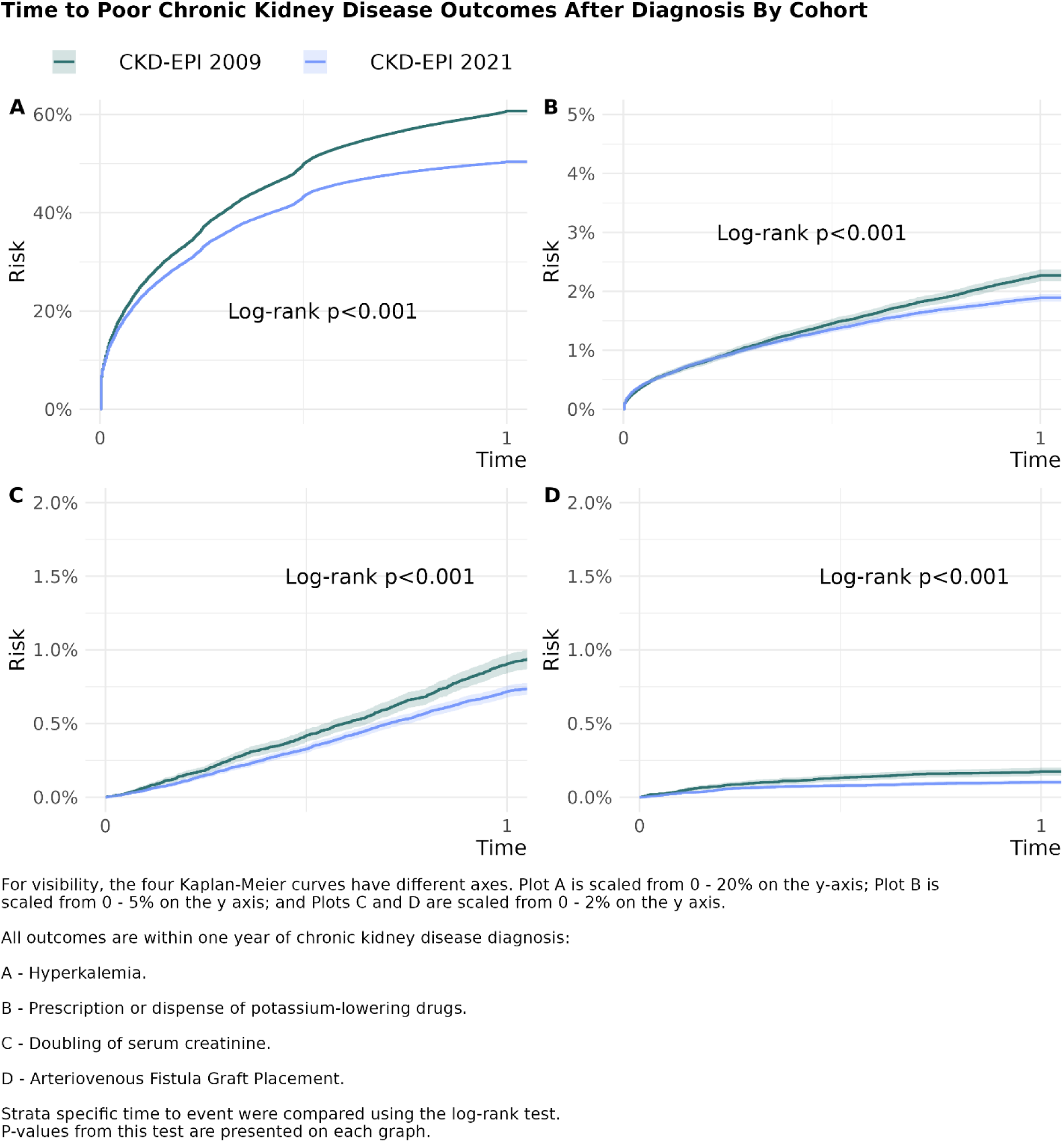
Time to Poor Chronic Kidney Disease Outcomes After Diagnosis by Cohort

**Figure 2.**
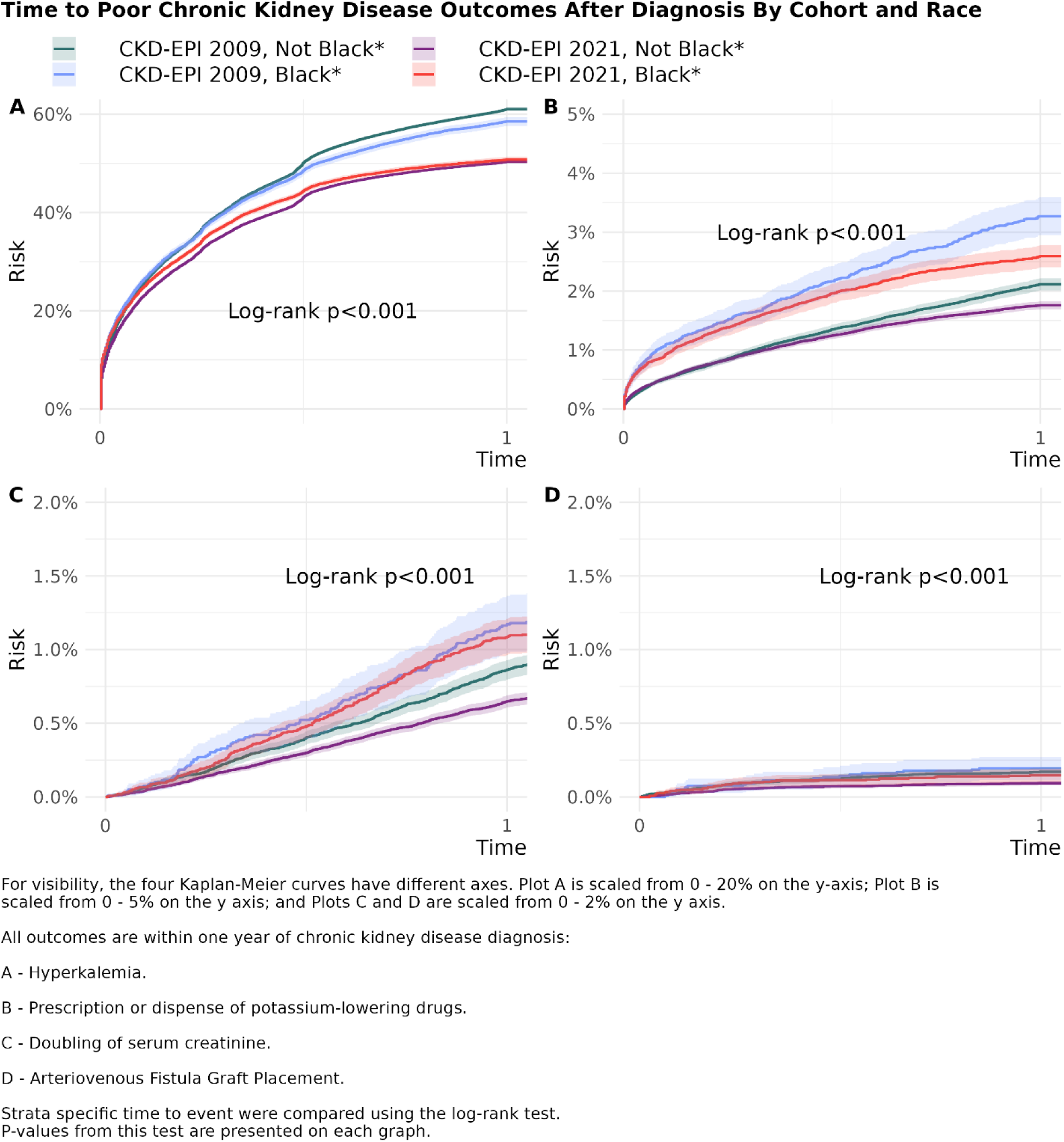
Time to Poor Chronic Kidney Disease Outcomes After Diagnosis by Cohort and Race

After controlling for comorbidities, SDOH, and demographics, we identified significant interaction between race and cohort in the time-to-hyperkalemia (Table 2). Overall, while both Black and Non-Black patients in the post-update cohort had reductions in the time-to-hyperkalemia compared to patients in the pre-update cohort, we found a greater reduction in the diagnosis of hyperkalemia within a year of CKD diagnosis for Non-Black patients compared to Black patients (Table 2, Table 3; p < 0.001). Non-Black patients in the post-update cohort were less likely than those in the pre-update cohort to be diagnosed with hyperkalemia within a year of CKD diagnosis (AHR 0.78; CI 0.77, 0.79). Black patients in the post-update cohort were less likely than those in the pre-update cohort to be diagnosed with hyperkalemia (AHR 0.83; CI 0.80, 0.86). Patients with hyperlipidemia (AHR 1.34; CI 1.23, 1.26) and patients with high comorbidity (AHR 1.29; CI 1.27, 1.31) were more likely to be diagnosed with hyperkalemia than patients without hyperlipidemia and patients with low comorbidity.

**Table 2.**
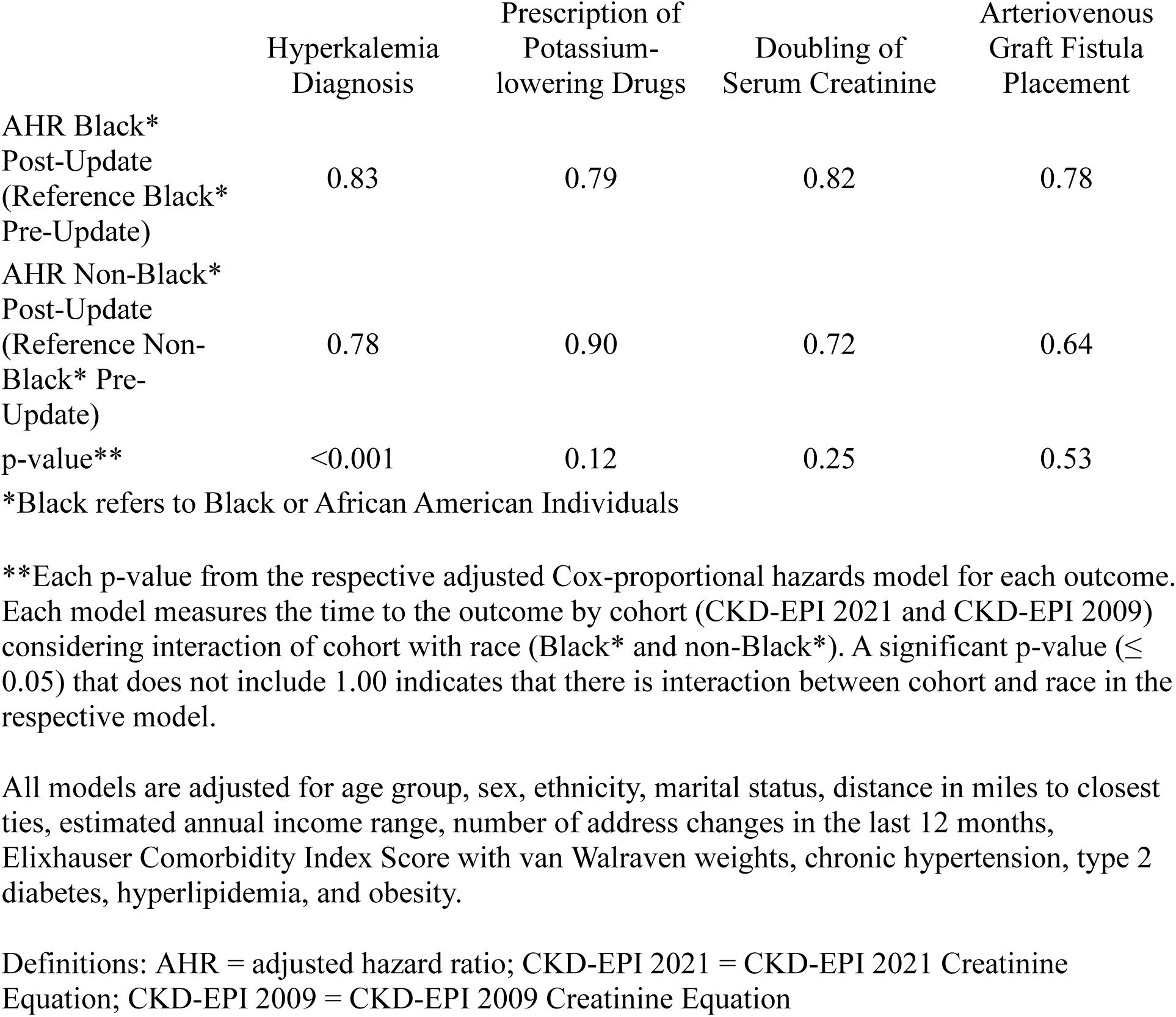
Comparison of adjusted cox-proportional hazard ratios by cohort and race for time to prescription of potassium-lowering drugs, arteriovenous graft fistula placement, hyperkalemia diagnosis, and doubling of serum creatinine after chronic kidney disease diagnosis

**Table 3.** Impact of Demographics, Social Determinants and Comorbidities on Hyperkalemia After Chronic Kidney Disease Diagnosis.

| Characteristic | HR<sup>I</sup> | 95% CI<sup>I</sup> | p-value |
| --- | --- | --- | --- |
| **CKD-EPI Serum Creatinine Formula** |  |  |  |
| CKD-EPI 2009 (Pre-update) | — | — |  |
| CKD-EPI 2021 (Post-update) | 0.78 | 0.77, 0.79 | <0.001 |
| **Race** |  |  |  |
| Not Black* | — | — |  |
| Black* | 1.02 | 0.99, 1.04 | 0.26 |
| **Age Group** |  |  |  |
| 18-44 | — | — |  |
| 45-64 | 1.03 | 1.00, 1.07 | 0.049 |
| 65+ | 1.03 | 1.0, 1.06 | 0.10 |
| **Sex** |  |  |  |
| Female | — | — |  |
| Male | 0.95 | 0.94, 0.96 | <0.001 |
| **Ethnicity** |  |  |  |
| Hispanic or Latino | — | — |  |
| Not Hispanic or Latino | 0.93 | 0.91, 0.95 | <0.001 |
| Unknown | 0.84 | 0.81, 0.87 | <0.001 |
| **Marital Status** |  |  |  |
| Unmarried and Never Married | — | — |  |
| Ever Married or Partnered | 1.08 | 1.06, 1.10 | <0.001 |
| Other | 1.06 | 0.98, 1.14 | 0.13 |
| Unknown | 1.18 | 1.15, 1.22 | <0.001 |
| **Distance in Miles to Closest Ties** |  |  |  |
| Less than 25 miles | — | — |  |
| 25 - 100 miles | 1.04 | 1.00, 1.09 | 0.050 |
| 100 - 500 miles | 1.04 | 0.99, 1.08 | 0.13 |
| Over 500 miles | 1.05 | 1.01, 1.09 | 0.019 |
| No Information | 1.04 | 1.01, 1.08 | 0.005 |
| **Estimated Annual Income Range (USD\$)** | | | |
| 0 - 30,000 | — | — |  |
| 30,001 - 50,000 | 1.03 | 1.00, 1.05 | 0.016 |
| 50,001 - 80,000 | 1.06 | 1.04, 1.08 | <0.001 |
| 80,001 - 100,000 | 1.08 | 1.05, 1.12 | <0.001 |
| 100,001 - 120,000 | 1.09 | 1.05, 1.13 | <0.001 |

| **Characteristic** | **HR<sup>1</sup>** | **95% CI<sup>1</sup>** | **p-value** |
| --- | --- | --- | --- |
| More than 120K | 1.05 | 1.00, 1.09 | 0.045 |
| **Number of Address Changes in Last 12 Months** |  |  |  |
| 0 | — | — |  |
| 1 | 1.02 | 1.00, 1.05 | 0.018 |
| 2 or More | 1.08 | 1.03, 1.13 | 0.001 |
| **Elixhauser Comorbidity Index Score\*\*** |  |  |  |
| 0- < 11 | — | — |  |
| 11 - 15 | 1.12 | 1.11, 1.14 | <0.001 |
| ≥ 15 | 1.29 | 1.27, 1.31 | <0.001 |
| **Chronic Hypertension (Yes/No)** |  |  |  |
| No | — | — |  |
| Yes | 1.00 | 0.99, 1.02 | 0.52 |
| **Type 2 Diabetes (Yes/No)** |  |  |  |
| No | — | — |  |
| Yes | 1.08 | 1.06, 1.09 | <0.001 |
| **Hyperlipidemia (Yes/No)** |  |  |  |
| No | — | — |  |
| Yes | 1.24 | 1.23, 1.26 | <0.001 |
| **Obesity** |  |  |  |
| No | — | — |  |
| Yes | 1.11 | 1.10, 1.12 | <0.001 |
| **CKD-EPI Serum Creatinine Formula \* Race** |  |  |  |
| CKD-EPI 2021 * Black* | 1.06 | 1.02, 1.09 | <0.001 |

\*Black refers to Black or African American Individuals
\*\*Elixhauser Comorbidity Index computed using van Walraven weights. Categories correlate to low, medium, and high comorbidity
<sup>1</sup>HR = Hazard Ratio, CI = Confidence Interval

We found a significant difference by cohort in time to prescription of potassium-lowering drugs (Table 4), doubling of serum creatinine (Table 5), and AVF placement (Table 6) after controlling for all covariates. However, changes in the chance of each outcome were not different for Black and non-Black patient in each cohort. (Table 2).

**Table 4.** Impact of Demographics, Social Determinants and Comorbidities on Prescription of Potassium-lowering Drugs After Chronic Kidney Disease Diagnosis.

| Characteristic | HR<sup>I</sup> | 95% CI<sup>I</sup> | p-value |
| --- | --- | --- | --- |
| **CKD-EPI Serum Creatinine Formula** |  |  |  |
| CKD-EPI 2009 (Pre-update) | — | — |  |
| CKD-EPI 2021 (Post-update) | 0.87 | 0.82, 0.93 | <0.001 |
| **Race** |  |  |  |
| Not Black* | — | — |  |
| Black* | 1.27 | 1.17, 1.38 | <0.001 |
| **Age Group** |  |  |  |
| 18-44 | — | — |  |
| 45-64 | 1.00 | 0.87, 1.14 | 0.96 |
| 65+ | 0.76 | 0.67, 0.87 | <0.001 |
| **Sex** |  |  |  |
| Female | — | — |  |
| Male | 1.33 | 1.25, 1.42 | <0.001 |
| **Ethnicity** |  |  |  |
| Hispanic or Latino | — | — |  |
| Not Hispanic or Latino | 0.57 | 0.52, 0.64 | <0.001 |
| Unknown | 0.72 | 0.61, 0.85 | <0.001 |
| **Marital Status** |  |  |  |
| Unmarried and Never Married | — | — |  |
| Ever Married or Partnered | 0.81 | 0.75, 0.89 | <0.001 |
| Other | 1.49 | 1.12, 1.98 | 0.007 |
| Unknown | 1.03 | 0.90, 1.19 | 0.66 |
| **Distance in Miles to Closest Ties** |  |  |  |
| Less than 25 miles | — | — |  |
| 25 - 100 miles | 1.30 | 1.06, 1.60 | 0.011 |
| 100 - 500 miles | 0.96 | 0.75, 1.24 | 0.78 |
| Over 500 miles | 1.17 | 0.95, 1.45 | 0.13 |
| No Information | 0.86 | 0.72, 1.03 | 0.11 |
| **Estimated Annual Income Range (USD\$)** | | | |
| 0 - 30,000 | — | — |  |
| 30,001 - 50,000 | 0.73 | 0.67, 0.81 | <0.001 |
| 50,001 - 80,000 | 0.64 | 0.58, 0.72 | <0.001 |
| 80,001 - 100,000 | 0.55 | 0.47, 0.65 | <0.001 |
| 100,001 - 120,000 | 0.57 | 0.46, 0.70 | <0.001 |

| **Characteristic** | **HR<sup>I</sup>** | **95% CI<sup>I</sup>** | **p-value** |
| --- | --- | --- | --- |
| More than 120K | 0.54 | 0.41, 0.71 | <0.001 |
| **Number of Address Changes in Last 12 Months** |  |  |  |
| 0 | — | — |  |
| 1 | 1.27 | 1.15, 1.40 | <0.001 |
| 2 or More | 1.32 | 1.07, 1.64 | 0.009 |
| **Elixhauser Comorbidity Index Score\*\*** |  |  |  |
| 0- < 11 | — | — |  |
| 11 - 15 | 1.02 | 0.91, 1.13 | 0.79 |
| ≥ 15 | 1.14 | 1.04, 1.24 | 0.006 |
| **Chronic Hypertension (Yes/No)** |  |  |  |
| No | — | — |  |
| Yes | 0.73 | 0.68, 0.78 | <0.001 |
| **Type 2 Diabetes (Yes/No)** |  |  |  |
| No | — | — |  |
| Yes | 1.82 | 1.70, 1.95 | <0.001 |
| **Hyperlipidemia (Yes/No)** |  |  |  |
| No | — | — |  |
| Yes | 0.72 | 0.67, 0.77 | <0.001 |
| **Obesity (Yes/No)** |  |  |  |
| No | — | — |  |
| Yes | 0.84 | 0.79, 0.90 | <0.001 |

\*Black refers to Black or African American Individuals
\*\*Elixhauser Comorbidity Index computed using van Walraven weights. Categories correlate to low, medium, and high comorbidity
<sup>I</sup>HR = Hazard Ratio, CI = Confidence Interval

**Table 5.** Impact of Demographics, Social Determinants and Comorbidities on Doubling of Serum Creatinine After Chronic Kidney Disease Diagnosis.

| Characteristic | HR<sup>I</sup> | 95% CI<sup>I</sup> | p-value |
| --- | --- | --- | --- |
| **CKD-EPI Serum Creatinine Formula** |  |  |  |
| CKD-EPI 2009 (Pre-update) | — | — |  |
| CKD-EPI 2021 (Post-update) | 0.74 | 0.68, 0.81 | <0.001 |
| **Race** |  |  |  |
| Not Black* | — | — |  |
| Black* | 1.33 | 1.19, 1.49 | <0.001 |
| **Age Group** |  |  |  |
| 18-44 | — | — |  |
| 45-64 | 0.81 | 0.67, 0.97 | 0.024 |
| 65+ | 0.60 | 0.50, 0.72 | <0.001 |
| **Sex** |  |  |  |
| Female | — | — |  |
| Male | 0.93 | 0.85, 1.02 | 0.12 |
| **Ethnicity** |  |  |  |
| Hispanic or Latino | — | — |  |
| Not Hispanic or Latino | 0.80 | 0.68, 0.93 | 0.005 |
| Unknown | 0.75 | 0.58, 0.97 | 0.026 |
| **Marital Status** |  |  |  |
| Unmarried and Never Married | — | — |  |
| Ever Married or Partnered | 0.95 | 0.84, 1.07 | 0.37 |
| Other | 0.98 | 0.60, 1.62 | 0.94 |
| Unknown | 1.17 | 0.96, 1.43 | 0.12 |
| **Distance in Miles to Closest Ties** |  |  |  |
| Less than 25 miles | — | — |  |
| 25 - 100 miles | 1.08 | 0.79, 1.47 | 0.64 |
| 100 - 500 miles | 0.95 | 0.67, 1.34 | 0.75 |
| Over 500 miles | 1.06 | 0.79, 1.44 | 0.69 |
| No Information | 0.86 | 0.67, 1.11 | 0.24 |
| **Estimated Annual Income Range (USD\$)** | | | |
| 0 - 30,000 | — | — |  |
| 30,001 - 50,000 | 0.71 | 0.63, 0.81 | <0.001 |
| 50,001 - 80,000 | 0.59 | 0.51, 0.68 | <0.001 |
| 80,001 - 100,000 | 0.65 | 0.53, 0.80 | <0.001 |
| 100,001 - 120,000 | 0.51 | 0.38, 0.69 | <0.001 |

| **Characteristic** | **HR<sup>1</sup>** | **95% CI<sup>1</sup>** | **p-value** |
| --- | --- | --- | --- |
| More than 120K | 0.60 | 0.42, 0.85 | 0.005 |
| **Number of Address Changes in Last 12 Months** |  |  |  |
| 0 | — | — |  |
| 1 | 1.16 | 1.01, 1.34 | 0.035 |
| 2 or More | 1.08 | 0.78, 1.50 | 0.63 |
| **Elixhauser Comorbidity Index Score\*\*** |  |  |  |
| 0- < 11 | — | — |  |
| 11 - 15 | 1.14 | 0.99, 1.31 | 0.066 |
| ≥ 15 | 1.46 | 1.30, 1.64 | <0.001 |
| **Chronic Hypertension (Yes/No)** |  |  |  |
| No | — | — |  |
| Yes | 0.74 | 0.67, 0.81 | <0.001 |
| **Type 2 Diabetes (Yes/No)** |  |  |  |
| No | — | — |  |
| Yes | 1.53 | 1.40, 1.68 | <0.001 |
| **Hyperlipidemia (Yes/No)** |  |  |  |
| No | — | — |  |
| Yes | 1.00 | 0.90, 1.11 | >0.99 |
| **Obesity (Yes/No)** |  |  |  |
| No | — | — |  |
| Yes | 1.07 | 0.98, 1.18 | 0.12 |

\*Black refers to Black or African American Individuals
\*\*Elixhauser Comorbidity Index computed using van Walraven weights. Categories correlate to low, medium, and high comorbidity
<sup>1</sup>HR = Hazard Ratio, CI = Confidence Interval

**Table 6.** Impact of Demographics, Social Determinants and Comorbidities on Fistula Placement After Chronic Kidney Disease Diagnosis.

| Characteristic | HR<sup>I</sup> | 95% CI<sup>I</sup> | p-value |
| --- | --- | --- | --- |
| **CKD-EPI Serum Creatinine Formula** |  |  |  |
| CKD-EPI 2009 (Pre-update) | — | — |  |
| CKD-EPI 2021 (Post-update) | 0.67 | 0.53, 0.85 | <0.001 |
| **Race** |  |  |  |
| Not Black* | — | — |  |
| Black* | 1.13 | 0.83, 1.55 | 0.44 |
| **Age Group** |  |  |  |
| 18-44 | — | — |  |
| 45-64 | 1.13 | 0.74, 1.73 | 0.56 |
| 65+ | 0.64 | 0.42, 1.00 | 0.048 |
| **Sex** |  |  |  |
| Female | — | — |  |
| Male | 1.36 | 1.07, 1.72 | 0.013 |
| **Ethnicity** |  |  |  |
| Hispanic or Latino | — | — |  |
| Not Hispanic or Latino | 0.43 | 0.31, 0.59 | <0.001 |
| Unknown | 0.35 | 0.18, 0.69 | 0.002 |
| **Marital Status** |  |  |  |
| Unmarried and Never Married | — | — |  |
| Ever Married or Partnered | 0.65 | 0.49, 0.87 | 0.004 |
| Other | 1.29 | 0.47, 3.53 | 0.62 |
| Unknown | 0.67 | 0.39, 1.16 | 0.15 |
| **Distance in Miles to Closest Ties** |  |  |  |
| Less than 25 miles | — | — |  |
| 25 - 100 miles | 1.44 | 0.71, 2.91 | 0.32 |
| 100 - 500 miles | 0.21 | 0.03, 1.47 | 0.12 |
| Over 500 miles | 0.92 | 0.38, 2.24 | 0.86 |
| No Information | 1.06 | 0.58, 1.96 | 0.84 |
| **Estimated Annual Income Range (USD\$)** | | | |
| 0 - 30,000 | — | — |  |
| 30,001 - 50,000 | 1.40 | 0.93, 2.11 | 0.10 |
| 50,001 - 80,000 | 1.27 | 0.81, 1.98 | 0.30 |
| 80,001 - 100,000 | 0.64 | 0.31, 1.35 | 0.24 |
| 100,001 - 120,000 | 1.24 | 0.57, 2.67 | 0.59 |

| **Characteristic** | **HR<sup>I</sup>** | **95% CI<sup>I</sup>** | **p-value** |
| --- | --- | --- | --- |
| More than 120K | 1.59 | 0.68, 3.69 | 0.29 |
| **Number of Address Changes in Last 12 Months** |  |  |  |
| 0 | — | — |  |
| 1 | 1.11 | 0.77, 1.61 | 0.58 |
| 2 or More | 0.69 | 0.26, 1.86 | 0.46 |
| **Elixhauser Comorbidity Index Score\*\*** |  |  |  |
| 0- < 11 | — | — |  |
| 11 - 15 | 0.40 | 0.21, 0.75 | 0.004 |
| ≥ 15 | 0.34 | 0.18, 0.63 | <0.001 |
| **Chronic Hypertension (Yes/No)** |  |  |  |
| No | — | — |  |
| Yes | 0.25 | 0.19, 0.33 | <0.001 |
| **Type 2 Diabetes (Yes/No)** |  |  |  |
| No | — | — |  |
| Yes | 1.66 | 1.30, 2.13 | <0.001 |
| **Hyperlipidemia (Yes/No)** |  |  |  |
| No | — | — |  |
| Yes | 0.64 | 0.49, 0.82 | <0.001 |
| **Obesity (Yes/No)** |  |  |  |
| No | — | — |  |
| Yes | 0.75 | 0.58, 0.96 | 0.022 |

\*Black refers to Black or African American Individuals
\*\*Elixhauser Comorbidity Index computed using van Walraven weights. Categories correlate to low, medium, and high comorbidity
<sup>I</sup>HR = Hazard Ratio, CI = Confidence Interval

Prescription of potassium-lowering drugs was less likely for patients in the post-update cohort than those in the pre-update cohort when considering covariates (AHR 0.87; CI 0.82, 0.93). Black patients were more likely (AHR 1.27; CI 1.17, 1.38) to obtain potassium lowering drugs than non-Black patients. Patients that were obese (AHR 0.84; CI 0.79, 0.90) and those that had chronic hypertension (AHR 0.73; CI 0.68, 0.78) were less likely to have a prescription or dispense of a potassium-lowering drug.

The probability of serum creatinine doubling within one year was lower for patients in the post-update cohort compared to patients in the pre-update cohort (AHR 0.74; CI 0.68, 0.81). This probability significantly decreased as income increased (Table 5). Patients with chronic hypertension experienced a lower probability of serum creatinine doubling than patients without chronic hypertension (AHR 0.74; CI 0.67, 0.81). Patients with Type 2 diabetes were more likely to have a doubled serum creatinine than patients without Type 2 diabetes (AHR 1.53; CI 1.40, 1.68).

The probability of having an AVF within a year after CKD diagnosis was 23% lower for patients in the post-update cohort than patients in the pre-update cohort (AHR 0.67; CI 0.53, 0.85). Non-Hispanic or Latino patients (AHR 0.43; CI 0.31, 0.59), patients that were ever married or partnered (AHR 0.65; CI 0.49, 0.87), and patients with chronic hypertension (AHR 0.25; CI 0.19, 0.33) were less likely to get an AVF within a year of diagnosis compared to Hispanic or Latino patients, patients that were never married or partnered, and patients without chronic hypertension respectively.

## Discussion

The purpose of this retrospective cohort study was to identify changes in time between first CKD diagnosis and four clinical endpoints tied to ESKD – hyperkalemia, prescription of potassium-lowering drugs, doubling of serum creatinine, and hemodialysis (as measured by AVF placement) – after the implementation of the race-free CKD-EPI 2021. Hyperkalemia was the most probable of the four outcomes investigated in this study of 261,774 patients within the first year after CKD diagnosis after controlling for comorbidities, demographics, and SDOH. Hyperkalemia was the only outcome with a significant change in probability of diagnosis when considering the interaction of the recommended equation (cohort) and race.

### Hyperkalemia disparities by race

Both non-Black and Black patients were less likely to be diagnosed with hyperkalemia after the CKD-EPI 2021 implementation than before the implementation. However, Black patients were still significantly more likely than Non-Black patients to be diagnosed with hyperkalemia within a year of diagnosis. This is consistent with literature suggesting Black patients have high levels of serum potassium and have a higher risk of hyperkalemia within one year of diagnosis compared to white patients.(28) The observed increased risk in this study for Black patients is potentially due to the CKD-EPI 2009 race-adjustment that led to late diagnosis and worse staging at diagnosis than non-Black patients; however, this question is outside the scope of this study.

### Prescription of potassium lowering drugs does not show the same trend

Hyperkalemia can be treated with potassium binders; newer medications include patiromer and sodium zirconium cyclosilicate. Longer term potassium-lowering drug use has been associated with lower mortality for patients with hyperkalemia.(32) These drugs can be expensive in the US, especially for patients that are underinsured or uninsured, which may drive the low utilization in this study and in others.(12)

Fifty-four percent of the present study’s patient population developed hyperkalemia but only 0.1% of patients obtained a prescription for potassium-lowering drugs (Table 1). The higher probability of prescription observed for Black patients could be due to earlier diagnosis, lower staging of disease at diagnosis, and earlier access to treatment since race was removed from the CKD-EPI creatinine equation.(21) The longer CKD-EPI 2021 is used, more data will be available to observe time based changes in treatment and diagnosis.

### Doubling of serum creatinine

Doubling of serum creatinine is accepted by the US Food and Drug Administration as a surrogate for kidney failure development and mortality in clinical trials.(16,18,38) Other studies have estimated that the doubling of serum creatinine is equal to a 30% to 50% decrease in a patient’s GFR.(18,39,40) The race-inclusive CKD-EPI 2009 equation and the race-free CKD-EPI 2021 equation provide different GFR estimations especially for Black patients, affecting both the initial GFR estimation and calculations of GFR change over time. Whereas the change in GFR is also dependent on other baseline patient characteristics, treatments, and comorbidities, the present study findings support including the CKD-EPI GFR estimation recommendation as a factor in clinical trials and longitudinal studies to reduce bias in study results.(16,17,38,39)

### Placement of an AVF

We found a difference in time to AVF placement based on which equation was recommended at time of CKD diagnosis. However, the results were non-informative as we did not have a large enough population of patients with an AVF placement within one year of CKD diagnosis. While disparities exist and are known to exist for the timing to start dialysis, we did not see a significant difference in placement of an AVF by race in this study.(41,42) A longer study period may improve our ability to examine this outcome.

### The impact of SDOH and comorbidities

SDOH and disease comorbidities are important for the development of CKD and disease progression.(5,21,28,43) These factors were significant contributors to potassium-lowering drug prescription and serum creatinine doubling within a year of CKD diagnosis. This may be due to a lack of consistent medical care due to frequent moves, poor access to care or medications, structural barriers leading to late diagnosis, or biased diagnostic guidelines such as CKD-EPI 2009.

Further statistical exploration of individual comorbidities, multimorbidity, and utilizing alternative SDOH variables could result in a stronger understanding of how patient and system factors contribute to these outcomes and when intervention may be needed.

### Calculating incidence as future work

Most available population estimates of CKD, including those from the CDC, Kidney Disease Surveillance System, and the United States Renal Data System, are prevalence estimates not incidence.(5,44–46) However, incidence numbers are readily available for ESKD.(44,45) The Kidney Disease Surveillance System’s published healthcare system estimate of CKD incidence uses Veterans Health Administration data, a population not representative of the adult US population.(47) Determining the incidence of CKD from prevalence is difficult as the average duration of the disease differs by stage and age.(48) While challenging, future work should target systematically identifying an accurate incidence for the US adult population.

### Strengths and limitations

#### Strengths

This study included 261,774 patients with CKD and had equal follow-up for patients diagnosed before and after the change in the CKD-EPI GFR estimation equation in 2021. The richness of our SDOH data allowed us to consider and adjust for individual level factors that are important for the occurrence of both the development of CKD and each outcome. Having this information at the patient level strengthened our belief in how race and ethnicity affected the time to event for CKD patient outcomes before and after the recommendations to adopt CKD-EPI 2021; it has highlighted additional areas for future research. Using EHR data allowed us to easily examine individual level time-based aspects surrounding the time between CKD diagnosis and each outcome of interest.

#### Limitations

Our study had several limitations. First, we used the placement of an AVF as opposed to a direct measure of dialysis treatment, which likely underestimated the true probability of receipt of dialysis within one year of diagnosis. Second, until 2021, primary diagnosis for CKD was based on equations that incorporated race and have demonstrated racial bias.(4,19,49) As we used the race categories that match CKD-EPI 2009 – Black and non-Black – we did not extend the study to examine a change in outcomes across more groups. Examining these outcomes across more diverse populations is necessary to improve clinical care.

EHR data is subject to a variety of limitations.(50) We are only able to identify events that are captured by the constituent health care systems that are a part of the Truveta member system. This means we will not capture disease diagnoses, medications, procedures, or laboratory values reported or diagnosed by a health care system that is not a part of the Truveta data. This limitation means we may have missed patients in the implementation of our inclusion and exclusion criteria.

## Conclusions

We found that the change in recommended CKD-EPI equation to CKD-EPI 2021 combined with race significantly decreased the time to hyperkalemia diagnosis within one year of CKD diagnosis. After the change to CKD-EPI 2021, there was a significant drop in the likelihood of patients being prescribed potassium-lowering drugs or having a doubled serum creatinine within a year of CKD diagnosis. While we expected the implementation of the race-free CKD-EPI 2021 to have a greater effect for Black patients than non-Black patients, we did not see this for most clinical endpoints. We did see decreased probabilities of all four outcomes in the post-update cohort which may indicate that implementation of the race-free CKD-EPI equation may result in the diagnosis of patients earlier in their disease course. Future work should elucidate the mechanisms driving these differences, whether there have been significant changes in treatment practices since the change in the GFR estimation equation, and long-term effects of each outcome.

## Data Availability

The data used in this study are available to all Truveta subscribers and may be accessed at studio.truveta.com.

## Declarations

## Ethics approval and consent to participate

Normalized electronic health record data are de-identified by expert determination under the HIPAA Privacy Rule before being made available to researchers. In accordance with 45 C.F.R. § 46.101 Protection of Human Subjects, our study did not require Institutional Review Board approval because it used only deidentified medical records. All data used in this study are publicly available to Truveta subscribers and may be accessed at studio.truveta.com.

## Availability of Data and Materials

The data and code used in this study is available to all Truveta subscribers and may be accessed at http://studio.truveta.com.

## Author contributions

Conceptualization: **CB, NLS, PJR, MS.** Methodology**: CB, PJR, RB, SG.** Data acquisition: **CB, SG, BMGC, RB**. Data analysis: **CB, SG, BMGC, PJR.** Interpretation: **CB, MS, NLS, PJR**. Initial draft: **CB.** Revisions: **CB, SG, BMGC, NLS, MS, PJR.** Final document review and approval for publication: All authors.

