## Supplement for "Time to Hyperkalemia and Other Outcomes Post-Implementation of the 2021 CKD-EPI Creatinine Equation: An Observational Cohort Study"

May 22, 2024

#### Data element definitions

Below are the codes used for defining chronic kidney disease, hyperkalemia, potassium lowering drugs, arteriovenous graft fistula, and serum creatinine. We also provide codes for exclusion criteria, all comorbidities included individually, and conditions included in the Elixhauser Comorbidity Index. All child codes are included even if not specified.

### 1 Study Outcomes

#### 1.1 Hyperkalemia

LOINC:
32713-0, 39789-3, 2823-3, 6298-4

#### 1.2 Potassium Lowering Drugs (Potassium Binders)

RxNorm:
1171402, 1170012, 1716211, 1716223, 730956, 1716213, 2047641, 2265525, 56512, 1716206, 1716217, 1799654, 2047628, 2101897, 2101898, 284757, 374633, 198221, 646148, 1716203, 1716212, 1716215, 1716219, 2101899, 1716207, 1716221, 2101896, 2375145, 2047629, 2047636, 2047638, 446440, 375288, 51670, 1181019, 1716208, 313072, 47630, 730957, 1086608, 1086609, 1159944, 1716214, 1716222, 1799649, 1799651, 2053927, 375287, 730958, 2047630, 2047639, 2375147, 422742, 446438, 546627, 730959, 546625, 646149, 1184185, 1716209, 1716216, 2047631, 2047634, 2375144, 1008140, 1171401, 1799650, 2047632, 2047633, 2047637, 2375146, 546626, 1086607, 1716210, 1716218, 313073, 2204723, 793741, 1170011, 1155083, 1184184, 1716220, 2053926, 446441, 1086606, 1159945, 1716205, 1799652, 1799653, 2047635, 2265524, 2265526, 316714, 422741, 793739, 793740, 1181018, 2047642

#### 1.3 Serum Creatinine

LOINC:
2160-0, 14682-9, 44784-7, 77140-2, 59826-8, 35203-9, 38483-4

#### 1.4 Arteriovenous Fistula Graft for Dialysis

CPT:
36836, 36837, 36818, 36819

SNOMED CT:
426340003

### 2 Exclusion

#### 2.1 Acute Kidney Injury

ICD10CM:
N17, N17.0, N17.1, N17.8, N17.2, N17.9

SNOMED CT:
22846003, 870589006, 609455009, 609482001, 88380005, 236432001, 12245621000119102, 423533009, 17380002, 71909003, 129721000119106, 36225005, 1177174007, 722278006, 14669001, 20483002, 298015003, 422593004, 733139004, 140031000119103, 307309005, 213231008, 269257004, 31005002, 430535006, 609472002, 722096006, 733137002, 438783006, 301814009, 722095005, 236429004, 368951000119105, 33561005, 236433006, 1263955004, 733839001, 52342006, 236428007, 145681000119101, 429489008, 429224003, 1263956003, 733138007, 200118004, 4451004, 236431008, 23697004, 51292008, 62216007

##### Polycystic Kidney Disease

ICD10CM: Q61.3, Q61.11, Q61.1, Q61.19, Q61.2

ICD9CM: 753.12, 753.13, 753.14

SNOMED CT: 253880009, 253884000, 771149000, 763891005, 717748005, 723409007, 737562008, 28770003, 236443009, 724094005, 717187000, 1216942009, 82525005, 765330003, 57088004, 5941000119101, 86463003, 253878003, 268234004, 253885004, 369071000119105, 717746009, 717749002, 1187460003, 765331004, 253879006, 1155732005, 773737004

#### 2.2 Renal Failure

ICD10CM:
I13.1, N18.1, Z49.32, N18.9, Z49, Z49.3, Z49.02, Z99.2, N25.0, N19, N18.5, N18, I12.0, I13.10, Z49.31, N18.32, Z49.01, Z49.0, N18.3, N18.6, I13.11, N18.4, Z94.0, N18.2, N18.31, N18.30

ICD9CM:
403.91, 585.6, 586, V45.11, V56.2, 404.92, V56, 403.01, V45.1, V42.0, 585.9, 404.12, 585.4, V56.31, 585.2, 404.13, 403.11, 404.02, V56.0, 588.0, 585.1, 404.93, 585.3, V56.3, 585.5, V45.12, 404.03, V56.1, 585, V56.32, V56.8

SNOMED CT:
731000119105, 430951000124108, 129161000119100, 22846003, 406168002, 153851000119106, 236436003, 718308002, 870589006, 897311008, 708975004, 609455009, 771000119108, 285851000119102, 431856006, 285001000119105, 722467000, 140101000119109, 285841000119104, 609482001, 88380005, 90751000119109, 96711000119105, 704667004, 285101000119109, 236432001, 430901000124109, 430941000124106, 12245621000119102, 26235008, 310647000, 363287001, 71421000119105, 700378005, 423533009, 90741000119107, 90791000119104, 71701000119105, 54155004, 275408006, 432601000124100, 43629001, 751000119104, 17380002, 723190009, 128001000119105, 368421000119108, 433144002, 96751000119106, 284961000119106, 71909003, 129721000119106, 285861000119100, 36225005, 1177174007, 29908007, 722278006, 700379002, 153891000119101, 140121000119100, 713696000, 111411000119103, 897310009, 45646000, 90761000119106, 432611000124102, 897312001, 368451000119100, 1217070004, 1263954000, 14669001, 20483002, 722150000, 129181000119109, 298015003, 129151000119102, 422593004, 733139004, 198708004, 449631000124102, 140031000119103, 285911000119109, 285881000119109, 307309005, 285081000119102, 285011000119108, 90731000119103, 49220004, 117681000119102, 1220586000, 368471000119109, 90688005, 46177005, 140131000119102, 285871000119106, 431855005, 127991000119101, 285061000119106, 43258006, 213231008, 430911000124107, 269257004, 737327002, 776416004, 96441000119101, 31005002, 10752771000119100, 430535006, 431857002, 90721000119101, 712487000, 609472002, 42399005, 722096006, 733137002, 236552002, 425369003, 268854008, 691421000119108, 438783006, 741000119101, 301814009, 368441000119102, 28196006, 718330001, 8468007, 433146000, 714153000, 1263953006, 609452007, 10757401000119104, 711000119100, 118781000119108, 722095005, 285831000119108, 236429004, 385971003, 96731000119100, 368951000119105, 284991000119104, 57557005, 33561005, 236435004, 284971000119100, 284981000119102, 236433006, 8996006, 1220648003, 198747002, 707324008, 16726004, 120261000119101, 285041000119107, 1263955004, 713313000, 430891000124105, 897308007, 721000119107, 1801000119106, 733839001, 52342006, 236428007, 145681000119101, 109477002, 429489008, 140111000119107, 429224003, 1263956003, 714152005, 368461000119103, 286371000119107,

194780003, 733138007, 49708008, 96741000119109, 200118004, 722098007, 4451004, 709044004, 269301005, 90771000119100, 430921000124104, 8501000119104, 724093004, 434431000124103, 430931000124101, 104931000119100, 236434000, 698591006, 368431000119106, 236431008, 96701000119107, 722149000, 1208934006, 285921000119102, 23697004, 10757481000119107, 129171000119106, 96721000119103, 51292008, 194781004, 4576001, 62216007

### 3 Comorbidities

#### 3.1 Alcohol Abuse

ICD10CM:
F10.98, T51.2X1A, T51.8X3, T51.94XA, F10.24, T51.1X3D, T51.3X3S, T51.8X1, F10.921, T51.3X1, T51.3X1D, F10.288, T51.8X4S, K29.20, T51.0X1D, T51.91, F10.988, T51.92XS, F10.25, T51.3X3D, F10.93, F10.229, T51.1X4, T51.8X3A, T51.2X2, F10.139, T51.0X3S, F10.929, T51.1X1A, T51.8, F10.121, T51.1X1S, F10.151, T51.2X3, T51.2X4S, T51.94, T51.0X4S, T51.2, T51.0X1A, T51.0X4, T51.1X2, T51.8X1A, T51.91XS, F10.950, F10.21, T51.8X2D, F10.130, F10.221, T51.91XA, T51.3X, F10.132, F10.930, F10.27, F10.281, T51.93XA, F10.231, K70.31, F10.230, T51.1X2D, T51.3X2A, T51.0X4A, T51.3X3A, F10.239, F10.95, F10.182, T51.8X4A, F10.23, K70.9, F10.181, T51.0X2D, F10.232, F10.22, T51.93XD, T51.2X4D, T51.92, F10.90, F10.11, Z71.4, T51.1X2S, T51.8X4, T51.2X2D, F10.180, F10.282, T51.2X3A, T51.8X, F10.28, T51.2X, T51.8X1S, F10.259, F10.29, F10.980, T51.1X3S, T51.1X4A, Z71.42, F10.129, T51.0X1S, F10.188, T51.8X1D, F10.18, F10.981, F10.20, F10.91, G62.1, K70.0, F10.13, T51.1X3, T51.8X2A, T51.1X2A, T51.8X3D, F10.1, T51.1X, T51.3X1A, T51.3X2S, F10.150, T51.2X2S, T51.3X3, T51.3X4S, K70.3, T51.92XD, F10.250, T51.1X3A, T51.94XD, T51.8X2S, T51.8X3S, T51.93, T51.0X2S, T51.91XD, F10.220, K70.30, T51.2X1, T51.2X3S, F10.15, T51.92XA, T51.1X4D, T51.2X1D, T51.3X4D, T51.0X2A, F10.131, F10.10, T51.3, T51.0X2, T51.0X, T51, F10.251, F10.26, T51.9, F10.120, F10, F10.12, I42.6, T51.1, T51.3X4, F10.9, T51.0X3A, Z71.41, F10.97, F10.19, F10.2, F10.99, T51.2X3D, T51.93XS, F10.932, F10.939, T51.1X1, T51.1X4S, F10.92, T51.8X4D, F10.931, F10.982, F10.920, T51.0X3, T51.2X2A, T51.0X3D, F10.14, F10.280, F10.96, T51.1X1D, F10.951, F10.959, T51.3X2, T51.3X2D, T51.3X4A, K29.21, K29.2, T51.2X4A, T51.3X1S, T51.2X4, T51.8X2, F10.94, T51.0X1, T51.94XS, F10.159, T51.0, T51.2X1S, T51.0X4D

ICD9CM:
303.90, 980.3, 303.01, 291.8, 303.02, 305.02, 291.89, 303.0, 303.00, 303.91, 425.5, 291.1, 291.9, 303.93, 303.92, 980, 535.3, 291.81, 305.01, 357.5, 980.9, 535.31, 305.03, 980.8, 980.2, 980.0, 291.2, 291.3, 303.9, 535.30, 303.03, V11.3, 980.1, 291.5, 305.00, 291.82, 305.0

SNOMED CT:
230800004, 291218002, 42423000, 216633005, 788542004, 41309000, 42344001, 1264172003, 207195004, 53936005, 235881000, 461231000124109, 185320006, 287166006, 191475009, 726007, 50687007, 284591009, 1149333003, 1153338005, 191883007, 1234783007, 398228004, 314849005, 386472008, 296270002, 698313007, 440068009, 425604002, 713776001, 866149003, 24882007, 48550003, 209099002, 439740005, 1255409006, 230353003, 133301000119102, 1234767007, 296259009, 308335008, 135331000119105, 68341005, 713370005, 212807002, 406547006, 31108002, 291226005, 44340006, 50357006, 82782008, 288836004, 724574008, 1234782002, 866204005, 314539001, 438515009, 698704008, 713281006, 1234774002, 315205008, 698307003, 698310005, 788339009, 18653004, 291274005, 19303008, 428931000124102, 281004, 420454001, 441244001, 765482002, 44047000, 185462000, 216651006, 291273004, 235880004,

281036007, 313183009, 440085006, 89507002, 400979004, 77406008, 108219001, 1234778004, 721710005, 85561006, 185387006, 1479971000168109, 82047000, 205791004, 296264008, 388975008, 296341009, 52703008, 1145073006, 296262007, 18170008, 191813001, 7200002, 77965002, 87810006, 185468001, 423215004, 6051000124107, 291211008, 270430005, 43695003, 308692003, 35755007, 61144001, 210098006, 216648004, 270425006, 1144767008, 216640006, 235942001, 408487007, 388977000, 361272001, 191802004, 440611006, 8635005, 59686008, 185348006, 191882002, 296950004, 291220004, 291221000, 308720009, 47237001, 1231846001, 86181006, 90526000, 34938008, 83362003, 296271003, 296951000, 711532000, 719410009, 440568002, 680007, 291923008, 723927004, 291217007, 1234779007, 30078008, 268645007, 1237136005, 73575006, 87790002, 191478006, 191804003, 185463005, 699208000, 237641009, 698308008, 291268005, 9953008, 69482004, 235875008, 401267002, 445450000, 73097000, 714829008, 788543009, 291216003, 401271004, 212813006, 698311009, 428921000124100, 1234750000, 5947002, 67426006, 185318008, 291214000, 1269515004, 29212009, 296260004, 83521008, 723882007, 408482001, 70495004, 296337005, 288031000119105, 291225009, 713583005, 4141000175103, 7052005, 58400001, 71318009, 421946003, 35437002, 698312002, 704126008, 15167005, 2530001, 12843005, 234366009, 291223002, 361273006, 439058005, 291219005, 269765000, 840449007, 25028003, 291270001, 291922003, 870194003, 21007002, 420650002, 439887005, 185346005, 185321005, 191805002, 234341005, 86013001, 6749002, 420054005, 87460008, 440146002, 185470005, 270424005, 440168007, 216645001, 713181003, 66590003, 191806001, 453131000124105, 309783001, 275693003, 65263004, 84251009, 278363000, 20881008, 723928009, 868184008, 310587004, 1197724009, 73007006, 191476005, 216635003, 135321000119107, 456201000124103, 205788004, 292881006, 34043003, 89291005, 1258986006, 390906007, 452081000124104, 59000001, 185464004, 440696002, 14736009, 49569001, 185345009, 185465003, 291215004, 296272005, 838386008, 1255408003, 71387007, 191884001, 698314001, 866203004, 773298008, 6061000124109, 408483006, 2043009, 296258001, 307757001, 216636002, 291224008, 310588009, 438516005, 135311000119100, 53923005, 191812006, 296949004, 426692001, 870191006, 719407002, 723103002, 108221006, 291275006, 212809004, 296338000, 423861007, 361268000, 609437000, 609438005, 7916009, 19681004, 453701000124103, 1234781009, 191480000, 36558000, 192811002, 270427003, 57346004, 65981005, 4525004, 4953006, 711420002, 1234772003, 12566000, 296340005, 445507008, 448337001, 97571000119109, 439708006, 28191001, 185469009, 291924002, 185389009, 291213006, 291276007, 41083005, 10755041000119100, 231467000, 461171000124100, 50325005, 291272009, 185349003, 185467006, 225929007, 386473003, 698321001, 3391000175108, 185347001, 838376007, 10741871000119101, 1234780005, 723930006, 185316007, 300992002, 711458003, 185466002, 1197766005, 719848005, 1234776000, 763184009, 185317003, 291222007, 296336001, 713862009, 288021000119107, 237738005, 291277003, 388970003, 698309000, 191471000, 25702006, 723929001, 11429006, 191811004, 291212001, 135301000119103, 15301000, 30274002, 8502007, 444971000124105, 235952002, 108220007, 296263002, 416520008, 723926008, 291269002, 291271002, 49463003, 69399002, 268796000, 296342002

#### 3.2 Blood Loss Anemia

ICD10CM:
D50.0

ICD9CM:
280.0

SNOMED CT:
234352004, 724556004, 413533008

##### 3.3 Cardiac Arrhythmias

ICD10CM:
T82.121S, R00.8, T82.110S, I47.2, T82.198D, I44.39, T82.198A, T82.199A, T82.119D, T82.118S, Z45.018, I47.11, I49.1, I47.1, I44.2, I47.19, T82.121A, I48.92, T82.119S, I47.20, I49.49, T82.110, T82.190, Z45.01, I49.3, I47.0, I48.4, T82.190A, I47.21, T82.128A, I48.1, I45.6, I48.91, T82.129, I44.30, T82.111, I49.01, I48.20, I48.11, I48.21, T82.199, I44.1, I48, R00.1, T82.198S, T82.119A, T82.120, T82.19, Z45.02, T82.121D, T82.191A, T82.129S, Z45.0, I48.9, T82.12, T82.199S, T82.191S, T82.119, T82.118A, R00.0, T82.111S, T82.118, T82.118D, I49.2, I49.5, T82.111A, T82.121, Z95.0, Z45.010, I49.8, I45.9, T82.199D, I49.9, I47, T82.111D, T82.110D, T82.190D, T82.129D, T82.110A, T82.198, I48.19, T82.190S, I49.02, I49.40, I47.10, T82.128, I44.3, T82.120A, I48.3, I47.29, T82.120D, I49.4, I49, I49.0, T82.128S, T82.120S, I47.9, T82.1, T82.129A, I48.0, Z45.09, I48.2, T82.11, T82.128D, T82.191, T82.191D

ICD9CM:
427.42, 427.32, V53.3, 426.10, 426.0, 426.13, V45.09, V53.31, 427.81, 996.04, 426.12, V45.02, 427.61, V53.39, 427.6, 427.9, V53.32, 427.69, 427.31, V45.00, 427.1, V45.0, 427.41, 427.4, 426.7, 427.3, 427.2, 427.89, 427.0, 427.8, 996.01, V45.01, 785.0, 426.9, 427.60

SNOMED CT:
49982000, 248652003, 251165007, 1186711002, 1217429005, 59118001, 82838007, 1204129001, 315027009, 1142067003, 1142123003, 1208864004, 6624005, 251167004, 40593004, 234214002, 418493005, 251158004, 698249005, 762247006, 708124001, 422348008, 251125008, 449846007, 31722008, 287057009, 397841007, 405806007, 703398004, 300997008, 1142086006, 280797561000119107, 721013001, 5561000124105, 314208002, 30667004, 1142069000, 1208909006, 234223004, 251180001, 733454004, 443325000, 715535009, 46319007, 233896004, 251173003, 251179004, 195070000, 444658006, 870697006, 32425009, 233900001, 462169004, 63467002, 195072008, 253528005, 870700005, 6285003, 251120003, 261195002, 10750641000119101, 13395001, 73459006, 251187003, 251188008, 472809000, 771179007, 33413000, 251152003, 410429000, 1142121001, 410430005, 1149362002, 63593006, 234181008, 69031000119105, 72654001, 181869007, 233901002, 251156000, 10745231000119102, 1237127006, 276513001, 698252002, 703162001, 715560009, 1142105006, 44602002, 426749004, 698271000, 1208866002, 473157001, 1208899000, 871832005, 2374000, 1208861007, 251163000, 129575004, 870695003, 120041000119109, 16574001, 36083008, 79325000, 716050002, 16415081000119104, 248653008, 251162005, 870694004, 432355004, 1142041008, 251160002, 1142103004, 27885002, 49436004, 1142093005, 111739000, 251124007, 233911009, 418341009, 609506003, 4006006, 234216000, 735685003, 770784003, 1204205002, 1230096008, 88412007, 162710004, 233916004, 251176006, 766883006, 789039008, 44273001, 233915000, 234211005, 251174009, 443478002, 10626002, 61277005, 5761000119100, 66657009, 74021003, 276796006, 419671004, 698251009, 162988008, 450919004, 441769002, 713419002, 15964501000119104, 762534000, 179924009, 440028005, 764732004, 1208824003, 240298005, 1142040009, 65778007, 71908006, 233909000, 240299002, 251092003, 233905006, 773587008, 180906006, 234225006, 68971000119104, 1142119006, 62026008, 698247007, 406461004, 251178007, 725060001, 1204167003, 49260003, 233898003, 397829000, 74615001, 723860000, 1163438008, 17869006, 285961000119107, 1142064005, 467643831000119105, 25569003, 78340007, 233919006, 429243003, 1208870005, 29320008, 233923003, 251157009, 703396000, 248651005, 251172008, 29894000, 699256006, 442559009, 472810005, 1142098001, 49044005, 234226007, 1208837000, 1264229001, 278085001, 39357005, 251094002, 735684004, 64545009, 13640000, 17366009, 213035005, 233899006, 251123001, 233892002, 449843004, 715865008, 233914001, 721010003, 1204124006, 251186007, 762681007, 1142068008, 60423000, 19092004, 1142090008, 1010446009, 4554005, 195069001, 1142057008, 46935006,

735683005, 62657007, 9651007, 1204171000, 20852007, 718104007, 46220003, 86014007, 111288001, 300996004, 251170000, 284470004, 47830009, 204384007, 251164006, 361137007, 195060002, 285951000119105, 233906007, 233897008, 234218004, 373905003, 283645003, 4973001, 6374002, 278086000, 1142204001, 72291000119104, 251168009, 1208868001, 41863008, 723866006, 1142120000, 111740003, 195080001, 233927002, 15964581000119107, 1142082008, 1142117008, 1163437003, 251171001, 423191000, 313377641000119105, 282825002, 1197418004, 1142066007, 195042002, 251161003, 706923002, 871644005, 86041000119107, 421656001, 735682000, 43906007, 870692000, 1142124009, 462170003, 1208911002, 710878005, 71792006, 720448006, 1230097004, 251177002, 1142111009, 309809007, 715395008, 59272004, 251155001, 764457005, 81681009, 234172002, 38566003, 195039008, 442917000, 77221000, 38274001, 1156821008, 63232000, 233904005, 10743741000119103, 5609005, 49710005, 251175005, 10760181000119109, 1172698005, 1220643007, 251182009, 289444003, 434392000, 737277001, 870696002, 1142110005, 1259226009, 195083004, 1230406008, 44103008, 233907003, 233918003, 285131000119102, 75532003, 32758004, 81898007, 1142122008, 12727006, 233910005, 413341007, 233891009, 15964901000119107, 46619002, 78240009, 251093008, 1208832006, 37760005, 10701000087104, 284941000119107, 284951000119109, 39260000, 28189009, 1208881008, 421348006, 16797001, 361136003, 54016002, 195105007, 233895000, 233903004, 871686004, 1010405004, 50799005, 251159007, 1142114001, 29717002, 425615007, 442946007, 489609371000119104, 26950008, 76887001, 162992001, 233894001, 27337007, 233902009, 270492004, 418818005, 21421000119109, 5370000, 233913007, 251181002, 234221002, 424571008, 78717006, 444605001, 184004, 67198005, 720507006, 10811961000119109, 55475008, 6180003, 195071001, 421869004, 74390002, 44808001, 83722004, 442515000, 119551000119102, 50006004, 1142118003, 1208863005, 12245071000119108, 3424008, 473006003, 6456007, 204383001, 733125004, 234220001, 11092001, 698504006, 719907006, 1179295004, 591071000124109, 426525004, 425582007, 698272007, 213213007, 441509002, 10164001, 20681005, 419752005, 719823007, 698270004, 11157007, 11849007, 725145002, 762682000, 789693005, 102451000119107, 12026006, 66568003, 251114004, 69730002, 233908008, 419400008, 1010443001, 1149361009, 1237128001, 86152005, 276512006, 422970001, 17338001, 195046004, 1222539006, 48867003, 420002000, 14718009, 251183004, 1208884000, 20143001, 413342000, 427665004, 233893007, 234219007, 423168004, 699748007, 715971003, 82226007, 233917008, 233922008, 418872003, 698250005, 251166008, 278482008, 440059007

#### 3.4 Chronic Pulmonary Disease

ICD10CM:
J67, J65, J45.991, J70.3, J45.22, J62, J63.5, J45.21, J45.902, J44, J45.51, J44.9, J44.1, J67.8, J67.0, J45.990, J61, I27.81, J44.0, J42, J43.0, I27.89, J45, J68.4, J43.8, J44.8, J67.1, J63, J66.0, J40, J63.0, J63.4, J70.1, J41.1, J66.2, J63.2, I27.82, J66.1, J67.3, I27.9, J67.7, J43.2, J43, J45.99, J63.3, J45.2, J45.909, J45.90, J43.1, J45.40, J45.32, I27.83, J60, I27.8, J67.4, J45.4, J45.50, J62.0, J45.20, J47.1, J63.6, J45.42, J63.1, J67.6, J45.5, J67.5, J45.9, J44.81, J66.8, J66, J41.0, J41.8, J47.0, J45.31, J47, J45.901, J45.30, J41, J64, J43.9, J67.9, J44.89, J45.998, J47.9, J62.8, J45.3, J45.41, J67.2, J45.52

ICD9CM:
493.11, 495.7, 493.12, 495.3, 493.82, 491.8, 495.9, 493.1, 496, 493.01, 501, 491.0, 500, 494.0, 494.1, 493.90, 495, 495.0, 416.8, 492.0, 495.5, 492.8, 504, 493.92, 493.22, 491.2, 502, 491.20, 490, 493.8, 491, 491.22, 505, 493.2, 493.20, 493.21, 492, 493.00, 493, 495.4, 416.9, 495.2, 493.10, 493.91, 493.0, 495.6, 491.21, 491.1, 495.8, 494, 493.02, 493.81, 491.9, 493.9, 495.1, 503

SNOMED CT:

19849005, 405720007, 707979007, 785728005, 76157009, 124991000119109, 135836000, 233687002, 233691007, 7343008, 16623004, 13617004, 40600002, 275495004, 313299006, 57607007, 87909002, 10692761000119107, 86638007, 195984007, 785744001, 83457000, 785737005, 52571006, 53926002, 10625671000119106, 313297008, 78723001, 195985008, 80010008, 846639005, 13394002, 15708009, 58691003, 64703005, 233760007, 195722003, 68328006, 233675009, 233751004, 31561003, 72270005, 870276004, 58890000, 707444001, 10692721000119102, 10785007, 123588006, 233757000, 13151001, 90623003, 233699009, 426853005, 79958002, 77690003, 734905008, 2360001000004109, 782513000, 50589003, 708093000, 72301000119103, 423889005, 16846004, 32139003, 51277007, 73144008, 805002, 49584005, 196001008, 879963005, 16003001, 11944003, 725415009, 10675991000119100, 88687001, 195725001, 103781000119103, 398640008, 424199006, 15964661000119102, 62371005, 67242002, 44547005, 1741000119102, 1751000119100, 10624991000119103, 408682005, 840351007, 80257001, 233761006, 10625151000119107, 86680006, 12295008, 370221004, 10625751000119106, 73448002, 195714005, 233672007, 10625551000119103, 312400008, 10676511000119109, 138389411000119105, 10625431000119105, 442025000, 12428000, 10674791000119101, 41269000, 233762004, 707511009, 233700005, 451981000124108, 10692681000119108, 33548005, 37981002, 404807005, 707513007, 106001000119101, 426979002, 445058002, 424643009, 79955004, 407674008, 735588005, 836477007, 10625791000119101, 10625391000119100, 70756004, 195728004, 10625031000119102, 40218008, 196049002, 196020005, 425748003, 29591002, 59786004, 10625231000119106, 12310001, 52333004, 74417001, 233599001, 266361008, 735465007, 233774000, 38729007, 785745000, 285381006, 370219009, 10625351000119105, 233683003, 5875001, 10625191000119102, 427295004, 404808000, 22951000119104, 10509002, 233679003, 10674711000119105, 707512002, 23022004, 65878001, 762521001, 25897000, 233702002, 47938003, 49691004, 233601004, 396286008, 840350008, 901000119100, 85438006, 233749003, 195727009, 703953004, 47515009, 195949008, 195989002, 782520007, 10674991000119104, 41553006, 64631008, 233677001, 404806001, 708038006, 35377009, 708095007, 27836007, 135181000119109, 72204002, 13217005, 10625631000119108, 40640008, 707981009, 10625071000119104, 10625511000119104, 195953005, 233759002, 396285007, 426656000, 733858005, 735589002, 785345002, 32398004, 1010333003, 68333005, 233598009, 10625871000119105, 71255007, 93432008, 85915003, 233758005, 846635004, 135171000119106, 63088003, 196026004, 16584951000119101, 86263001, 703954005, 63480004, 87433001, 707445000, 11641008, 36426008, 195720006, 10625471000119108, 195719000, 425969006, 714203003, 195967001, 16146001, 281239006, 48347002, 10625111000119106, 60805002, 10675871000119106, 233631005, 125001000119103, 19274004, 35301006, 1010334009, 708094006, 1010619008, 10675911000119109, 233754007, 10675751000119107, 196053000, 233695003, 707447008, 708096008, 10676271000119104, 293241000119100, 37471005, 51068008, 232430006, 233633008, 370220003, 10742121000119104, 78862003, 181007, 195729007, 233701009, 15199004, 707446004, 40122008, 36696005, 713175006, 735464006, 23958009, 34004002, 196017002, 233600003, 233756009, 233799004, 846638002, 266403005, 233674008, 733497009, 29422001, 71186008, 195721005, 233629001, 404804003, 10625271000119109, 89549007, 445378003, 55570000, 83291003, 115666004, 734904007, 1163489008, 111849006, 13645005, 233678006, 37711000, 42402006, 22607003, 707980005, 125011000119100, 95886009, 233630006, 18041002, 195959009, 1177120001, 12088005, 23315001, 49840000, 785736001, 61937009, 17996008, 389145006, 427603009, 27475006, 57686001, 92807009, 418395004, 233697006, 708090002, 50076003, 123587001, 19076009, 56968009, 85761009, 186178000, 708030004, 47895001, 195957006, 195958001, 233694004, 19325002, 34015007, 99031000119107, 33325001, 195726000, 10675471000119109, 405944004, 10625311000119109, 312371005, 786836003, 73414003, 233748006, 427679007, 52500008, 84409004, 762618008, 123589003, 195717003, 233627004, 233763009, 59940009, 409663006, 846637007, 10675431000119106, 18690003, 233688007, 125021000119107, 313296004, 370218001, 10676391000119108, 16055311000119107, 66987001, 77593006, 233755008, 266355005, 10676431000119103, 1259003, 185086009, 10625711000119105,

79479005, 225057002, 401000119107, 10675391000119101, 195977004, 69339004, 829976001, 233634002, 233698001, 445427006, 4981000, 31898008, 304527002, 15964701000119109, 233696002, 266356006, 707509000, 60485005, 89099002, 233632003, 233628009, 31387002, 195951007, 5281000124103, 10675551000119104, 10625591000119108, 64880000, 196019004

Truveta:
1215925

##### 3.5 Coagulopathy

ICD10CM:
D68.021, D68.61, D68.69, D68.03, D69.42, D69.5, D68.32, D68.59, D69.59, D68.8, D68.62, D69.51, D68.023, D68.52, D68.4, D68.311, D68.00, D69.1, D68.51, D68.02, D69.41, D68.022, D68.0, D69.3, D68.31, D67, D68.9, D69.6, D68.2, D68.029, D69.49, D68.01, D68.020, D68.04, D68.318, D68, D68.1, D68.5, D68.09, D68.6, D65, D69.4, D66, D68.3, D68.312

ICD9CM:
286, 286.4, 286.0, 286.2, 286.1, 286.59, 286.3, 287.33, 287.31, 286.9, 286.7, 287.1, 287.32, 287.3, 286.5, 287.39, 286.53, 286.52, 286.6, 287.41, 287.5, 287.30, 287.4, 287.49

SNOMED CT:
34478009, 234463000, 359536009, 328301000119102, 41461004, 237337003, 296932006, 425949001, 41106001, 441190003, 782909004, 71723006, 426199009, 200173001, 12546009, 43858000, 69500007, 128103006, 1264141002, 128099001, 721882001, 79674009, 50220002, 440924009, 5876000, 267199000, 438792009, 35554008, 439007008, 421102007, 441322009, 128108002, 38879000, 154818001, 359717002, 67406007, 191248000, 234370001, 6647006, 28505005, 234484005, 718553004, 30479005, 359711001, 733028000, 267272006, 275523003, 441191004, 789291002, 64509006, 128090002, 234464006, 6935003, 128102001, 16623961000119100, 234451005, 1285655009, 10934005, 276575001, 10749581000119100, 1285662000, 87397002, 191256002, 440820004, 1286003, 234444001, 860725002, 41788008, 328371000119107, 61744005, 95623001, 359730001, 60628003, 90935002, 4320005, 43346008, 123789000, 13172003, 441134009, 234448003, 1197595004, 81783000, 323079008, 128094006, 12501008, 30575002, 234486007, 267535004, 307517001, 724637001, 765748009, 715559004, 723489005, 23205009, 127034005, 296930003, 438373005, 86075001, 278366008, 402654005, 439455002, 783256001, 206510008, 439702007, 767713001, 50770000, 88540000, 307514008, 439274008, 1156838007, 307515009, 866152006, 12153008, 1162807005, 54569005, 58327003, 1148857003, 282707003, 1285657001, 33183004, 441188004, 49762007, 76771005, 128101008, 191244003, 191246001, 240486003, 720521008, 724356003, 191297009, 46760003, 19267009, 234466008, 302215000, 307342006, 720520009, 782934004, 1172685001, 37350004, 2897005, 51624005, 234445000, 402851000, 28975000, 80988005, 200030007, 722475006, 1172699002, 373420004, 439699000, 180481005, 33820001, 111396008, 439000005, 722401001, 123786007, 237271005, 302873008, 1563006, 328381000119105, 45366001, 10749641000119106, 128106003, 296931004, 440867000, 267524009, 717937006, 1148930004, 783251006, 440988005, 27068000, 41816006, 111589005, 359709005, 789660001, 76407009, 234485006, 403837005, 783255002, 64779008, 609462000, 1187615007, 307518006, 40855001, 128114009, 191255003, 416902009, 421766003, 27312002, 48788004, 36070007, 699208000, 61810006, 181456001, 1285661007, 234442002, 234483004, 64315007, 198912003, 118791000119106, 234447008, 47307007, 438827002, 65768009, 725157006, 46981006, 55907008, 128098009, 75331009, 123788008, 359721009, 1285663005, 713910008, 713508003, 234446004, 33297000, 33344008, 123790009, 296935008,

371106008, 31925001, 1217410004, 111452009, 128105004, 359727008, 724854007, 1264140001, 7933002, 49886003, 77084001, 95839005, 95840007, 359732009, 14230004, 84048006, 1285665003, 767712006, 128093000, 191323001, 296928000, 1285658006, 66909001, 439157002, 17592003, 18604004, 441101007, 413566007, 10854051000119101, 36351005, 1177171004, 359723007, 24663001, 234376007, 717407006, 1264232003, 47546008, 206511007, 234458004, 234482009, 62751000119108, 237270006, 191296000, 200031006, 234474009, 725034002, 237336007, 438360006, 28293008, 1908008, 21360006, 198917009, 2036003, 359531004, 440993008, 35066007, 191298004, 234462005, 296934007, 35913006, 89729000, 95841006, 306058006, 52137009, 39191000119103, 16773005, 237272003, 270117002, 1172604004, 3230006, 234456000, 95843009, 296936009, 19520006, 49177006, 1285656005, 128100009, 438492008, 713388002, 1259242002, 278365007, 234455001, 737221003, 768556005, 61551003, 95842004, 234460002, 439145006, 1285660008, 273986001, 32942005, 183005, 198918004, 234465007, 719021005, 95605009, 24149006, 34395002, 61802005, 73397007, 736024007, 9417000, 13993001, 62410004, 717936002, 719019000, 1187252002, 359714009, 26029002, 128096008, 33169001, 51720005, 267532001, 440866009, 723512008, 1148849001, 1156746003, 82835005, 85796009, 43302000, 10752381000119101, 95845002, 128091003, 240305000, 1162804003, 371074009, 1217409009, 16872008, 128107007, 1148859000, 82190001, 267527002, 721304007, 1285021005, 73975000, 234487003, 359704000, 86635005, 9489006, 37193007, 413565006, 45963004, 73162004, 78345002, 14024008, 105604006, 123787003, 19307009, 30182008, 402653004, 88776002, 154826009, 296927005, 439458000, 89962000, 234440005, 234459007, 725105006, 9311003, 13507004, 43217004, 51448009, 85589009, 716746003, 48976006, 711407000, 296926001, 50189006, 78129009, 25904003, 278504009, 439459008, 90414007, 37492005, 41690001, 34417008, 413567003, 6364000, 74576004, 128088003, 725291001, 783142006, 97571000119109, 10153004, 128113003, 234461003, 783194008, 16922007, 95844003, 267534000, 76642003, 111588002, 1196942001, 438372000, 439460003, 234450006, 1172901009, 128092005, 296929008, 773422002, 1208725005, 38970002, 440868005, 733096007, 234367000, 234457009, 87902006, 438476003, 234452003, 417626001, 441006000, 609456005, 717935003, 771075004, 3760002, 712922002, 128115005, 441192006, 774071007, 439156006, 1148862002, 15132005, 421132004, 1187614006, 1285666002, 32273002, 191322006, 1148929009, 206512000, 1264233008, 785308008, 62698000, 63444004, 191247005, 234454002, 717769007, 718554005, 1177197005, 1285664004, 438599002, 359700009, 296933001, 359725000, 441189007, 709465004, 716336002, 32605001, 128095007, 234453008, 1208617001, 1285659003

##### 3.6 Congestive Heart Failure

ICD10CM:
I25.5, I50.4, I42.9, I50.3, I50.30, I11.0, I50.22, I50.42, P29.0, I50.2, I50.33, I50.20, I42.0, I42.7, I13.2, I50.23, I50.32, I09.9, I50.41, I50.40, I50.21, I50.43, I42.6, I43, I42.5, I42.8, I50.31, I13.0

ICD9CM:
404.11, 404.01, 425.4, 428.2, 428.21, 428.9, 428.23, 428, 425.5, 428.32, 428.40, 428.30, 425.8, 428.41, 428.33, 425.9, 404.13, 428.42, 428.43, 428.20, 428.3, 428.31, 428.0, 404.91, 404.93, 402.91, 402.01, 404.03, 428.22, 428.1, 402.11, 398.91, 428.4, 425.7

SNOMED CT:
10091002, 15544002, 28284007, 415295002, 56941009, 312591002, 49699002, 71799002, 79754008, 90453003, 370141003, 721014007, 871639003, 56675007, 471875005, 82523003, 715655000, 763280005, 860839005, 446221000, 880040003, 46696008, 63183009, 426263006, 43736008, 472326004, 1149066008, 61012002, 195025008, 472325000, 6022005, 398716006, 101281000119107, 195023001, 83978005,

72481000119103, 39693003, 771513008, 48447003, 84114007, 111284004, 1172839002, 447935001, 472323007, 1954004, 702424003, 20005002, 44313006, 72011007, 91604001, 441541008, 715535009, 771509001, 55565007, 56501009, 54830008, 771478008, 1204468000, 34126003, 85898001, 96711000119105, 46165000, 285101000119109, 15629541000119106, 1264003007, 27097002, 84733009, 78069008, 756003, 52048004, 195112003, 253528005, 1197430005, 111285003, 472100003, 48872007, 87648004, 89819002, 233924009, 426300009, 58911001, 12023003, 64072006, 445336009, 49584005, 8209004, 424404003, 699668009, 775908005, 85232009, 120871000119108, 16063004, 23685000, 69609002, 195020003, 443253003, 443254009, 472320005, 79152002, 195021004, 15964661000119102, 7381000175100, 7391000175102, 96751000119106, 286011000119101, 1155719000, 717840005, 58629009, 84725008, 718713000, 68602002, 413995009, 609507007, 78031003, 472317002, 11104006, 67696008, 74960003, 720609003, 359617009, 766883006, 285221000119109, 1204462004, 30884007, 195027000, 195031006, 233878008, 67431000119105, 120851000119104, 1010447000, 890120009, 1208848007, 12563008, 53043001, 472316006, 1186866009, 5375005, 16855001, 53703001, 233875006, 441530006, 1137377003, 20529002, 30496006, 52029003, 277639002, 413994008, 364006, 24841007, 206586007, 719380003, 1153452008, 64077000, 79955004, 81552002, 719839000, 83014000, 471885006, 713523008, 314206003, 66989003, 83105008, 67441000119101, 788950000, 50270004, 21430002, 285911000119109, 276514007, 472319004, 715626008, 898208007, 74249003, 722919003, 285081000119102, 890121008, 195111005, 443344007, 15781000119107, 1010444007, 471880001, 1208846006, 64615000, 76931005, 7401000175100, 426856002, 286950001, 285061000119106, 120901000119108, 31085000, 441481004, 24584003, 25544003, 890122001, 60514000, 84871007, 871638006, 1234750000, 410431009, 471870000, 703276005, 774148007, 285211000119102, 233871002, 462172006, 471846004, 1204200007, 367363000, 85971001, 419097006, 717812000, 9909003, 34342000, 45227007, 83521008, 426611007, 715865008, 96311000119109, 19036004, 33644002, 1148765006, 40532006, 10633002, 88805009, 880014009, 194750008, 719835006, 35728003, 57803009, 93558006, 15629641000119107, 108701000119102, 16228931000119102, 2816000, 315608004, 7371000175103, 88318005, 472327008, 1086051000119107, 33006007, 46939000, 399020009, 18687009, 725416005, 10757401000119104, 53322007, 871617000, 417996009, 2024009, 471890009, 722095005, 788878000, 153931000119109, 45650007, 871649000, 194767001, 277638005, 96731000119100, 5148006, 44630002, 86995005, 50275009, 86466006, 194849004, 426012001, 204981000119101, 5053004, 472318007, 120881000119106, 60856006, 698594003, 860837007, 78862003, 153951000119103, 43121002, 7421000175106, 723993005, 64041007, 80479009, 789037005, 15629741000119102, 36471000, 16838951000119100, 1264404009, 398754006, 472315005, 719824001, 17759006, 471841009, 472324001, 89461002, 471858004, 429347004, 120891000119109, 35719004, 7411000175102, 1186718008, 120261000119101, 285041000119107, 16440002, 89456004, 703274008, 890119003, 460500001, 1230303001, 83291003, 724173009, 703272007, 59464004, 39041004, 41964001, 250999000, 195029002, 787001, 15629591000119103, 91529004, 194779001, 194709000, 719272007, 111000119104, 10335000, 120861000119102, 286031000119106, 62377009, 83898004, 111283005, 194741006, 119731000146105, 418304008, 462175008, 698296002, 700065003, 1204203009, 18192007, 92506005, 415509000, 44088000, 233873004, 1173036000, 1204206001, 40023000, 233877003, 1204194004, 447934002, 880038008, 1149109001, 1208843003, 72972005, 443343001, 13839000, 1208850004, 71892000, 462174007, 96741000119109, 42343007, 719451006, 195032004, 871646007, 28869005, 33258008, 46113002, 1155721005, 64749001, 86194008, 724550005, 704241002, 153941000119100, 1177176009, 195114002, 8501000119104, 15676002, 91442002, 406428006, 1204204003, 16253001, 240077004, 445236007, 720610008, 1187642008, 90828009, 286947004, 96701000119107, 880042006, 703273002, 15964701000119109, 285921000119102, 725027004, 10757481000119107, 23341000119109, 96721000119103, 90202001, 44993000, 194781004, 703275009, 195030007, 42099002, 1179293006, 28381002, 704242009, 1204460007

##### 3.7 Deficiency Anemia

ICD10CM:
D53.8, D52.1, D52.0, D51.9, D51.8, D53.0, D51.2, D53.2, D53.9, D53, D51.0, D52, D51.3, D51, D52.9, D53.1, D50.8, D51.1, D52.8

ICD9CM:
281.0, 281.1, 281.4, 281.2, 281.8, 280.9, 281, 280.8, 280.1, 281.3, 281.9

SNOMED CT:
85570009, 722005000, 717948004, 191142007, 1171357005, 84027009, 111574001, 18637002, 720401009, 48580008, 721295000, 44910003, 85746008, 49472006, 1153409008, 1148853004, 91217009, 721184003, 80126007, 191128004, 6659005, 234363001, 234361004, 82895008, 27798002, 267517008, 721186001, 52565000, 234360003, 1153403009, 1148856007, 191150003, 51667002, 87522002, 1142030006, 191161006, 24962009, 10812081000119108, 717946000, 191148006, 191149003, 290246007, 1153353002, 11491000, 199248002, 735452003, 735453008, 721185002, 191146005, 724557008, 86448001, 191154007, 724556004, 66612000, 1142087002, 34629009, 371315009, 1153333001, 1148855006, 87806008, 234362006, 1148861009, 85649008, 717947009, 267518003, 78677008

##### 3.8 Depression

ICD10CM:
F33.2, F34.1, F33.1, F33.42, F01.51, F33.9, F01.511, F33.41, F33.0, F32.1, F32.0, F33.3, F53.1, F32.2, F32.5, F32.9, F32.89, F32.3, F01.518, F32.4, F33.40, F34.81, F53.0, F34.89, F43.21, F33.8, F43.23

ICD9CM:
296.35, 309.1, 311, 296.36, 296.33, 296.34, 300.4, 296.24, 309.28, 290.43, 301.12, 296.31, 296.21, 296.22, 296.20, 298.0, 296.26, 296.23, 296.32, 296.30, 296.82, 309.0, 296.25, 290.21, 290.13

SNOMED CT:
274948002, 10835871000119104, 87512008, 765176007, 15193003, 237350002, 42810003, 81319007, 231542000, 15639000, 84788008, 231499006, 40568001, 63412003, 237349002, 724678004, 36923009, 788120007, 29929003, 33135002, 39809009, 69392006, 1234782002, 16266831000119100, 191616006, 413296003, 16265061000119105, 370143000, 25922000, 191630001, 71336009, 75084000, 67711008, 442057004, 60099002, 10811161000119107, 2618002, 724690002, 16238181000119101, 596004, 762345001, 83176005, 718636001, 288751000119101, 58703003, 192080009, 698957003, 63778009, 20250007, 84760002, 430852001, 726772006, 75837004, 38694004, 82218004, 1260298008, 30605009, 191659001, 77911002, 3109008, 83458005, 247803002, 16264901000119109, 724676000, 2506003, 77486005, 762329003, 18818009, 279225001, 16266991000119108, 762512002, 85080004, 319768000, 33736005, 16265301000119106, 1153570009, 191495003, 231504006, 720454007, 78667006, 268621008, 40379007, 48589009, 73867007, 191627008, 16265951000119109, 762321000, 79842004, 46244001, 724677009, 191604000, 320751009, 426578000, 10811121000119102, 762336002, 719593009, 723928009, 19527009, 321717001, 719592004, 191611001, 87842000, 33078009, 87414006, 832007, 76441001, 79298009, 720451004, 16264821000119108, 450714000, 10211000132109, 720452006, 94631000119100, 70747007, 36474008, 1153575004, 192079006, 231500002, 838530009, 35489007, 38451003, 191613003, 14183003, 104851000119103, 162722001, 16264621000119109, 251000119105, 720453001, 16238221000119109, 42925002, 762339009, 720455008,

19694002, 191634005, 36170009, 723930006, 28475009, 310495003, 191610000, 68019004, 231485007, 281000119103, 300706003, 14070001, 191629006, 712823008, 133121000119109, 191676002, 66344007, 310497006, 310496002

#### 3.9 Type 2 Diabetes

ICD10CM:
E11.49, E11.3539, E11.59, E11.52, E11.11, E11.3299, E11.21, E11.341, E11.40, E11.01, E11.3493, E11.3553, E11.3419, E11.51, E11.622, E11.44, E11.3213, O24.111, E11.3543, E11.3559, E11.3551, E11.3513, E11.3399, E11.3549, E11.319, E11.3311, E11.3533, E11.3413, E11.39, E11.311, E11.3592, E11.641, O24.119, E11.649, E11.3599, E11.3593, E11.37X1, E11.3511, E11.3292, E11.8, E11.9, E11.3591, E11.3492, E11.3212, E11.3491, E11.3531, E11.3541, E11.10, E11.43, E11.3529, E11.3412, E11.349, E11.359, E11.41, E11.3313, E11.29, E11.3319, E11.3523, E11.42, E11.3211, E11.331, E11.3519, E11.3552, E11.3542, E11.3499, O24.112, E11.3521, E11.65, E11.3291, E11.3532, E11.321, E11.3393, E11.3411, E11.3293, E11.329, E11.621, E11.3312, E11.3392, E11.3512, E11.610, E11.37X9, E11.3219, E11.628, E11.339, O24.12, E11.630, E11.36, E11.620, E11.3522, E11.3391, E11.22, E11.37X3, E11.618, E11.638, E11.69, E11.37X2, E11.00, O24.113, O24.13, E11.351

ICD9CM:
250.42, 250.70, 250.92, 250.40, 250.30, 250.50, 250.22, 250.90, 250.82, 250.60, 250.52, 250.20, 250.10, 250.32, 250.00, 250.12, 250.80, 250.62, 250.72, 250.02

SNOMED CT:
731000119105, 1501000119109, 16745131000119100, 420279001, 9859006, 190389009, 771000119108, 719216001, 16747021000119109, 28331000119107, 140101000119109, 421326000, 16747901000119104, 71421000119105, 104961000119108, 90791000119104, 87921000119104, 16747141000119104, 751000119104, 16745931000119102, 90781000119102, 16749661000119102, 16745531000119108, 1491000119102, 422034002, 140121000119100, 82551000119103, 237604008, 138941000119105, 190331003, 420436000, 314903002, 420756003, 769220000, 16746261000119108, 110181000119105, 140131000119102, 138921000119104, 127991000119101, 713706002, 421986006, 16746341000119103, 237599002, 1511000119107, 44054006, 60951000119105, 71441000119104, 1217044000, 82541000119100, 741000119101, 712883005, 97331000119101, 711000119100, 157141000119108, 16748141000119100, 16746901000119101, 703138006, 138911000119106, 609567009, 313436004, 41911000119107, 368581000119106, 16746131000119105, 1196922005, 421847006, 16745051000119107, 427027005, 314902007, 721000119107, 16747661000119109, 359642000, 422166005, 713703005, 420715001, 1481000119100, 140111000119107, 428007007, 368101000119109, 199230006, 237627000, 1551000119108, 16745651000119101, 781000119106, 421779007, 368591000119109, 1003605004, 97341000119105, 422099009, 81531005

#### 3.10 Diabetes with Complications

ICD10CM: E10.2, E11.33, E13.3593, E10.3419, E11.49, E13.359, E13.37, E10.44, E10.3523, E13.3319, E11.3539, E13.3291, E10.341, E11.59, E13.3392, E13.21, E10.37, E11.52, E13.33, E11.11, E13.4, E10.3393, E10.6, E11.62, E13.349, E13.3591, E13.6, E10.43, E11.355, E13.3393, E13.618, E10.3312, E10.3493, E10.35, E11.2, E11.5, E10.622, E13.3542, E10.319, E10.29, E13.3519, E13.3592, E10.3543, E11.3299, E10.351, E10.3553, E11.21, E10.3219, E10.3413, E11.341, E11.40, E13.37X2, E11.3493, E13.3599, E10.3319, E10.3522, E11.3553, E10.355, E11.3419, E11.51, E11.622, E13.3212, E13.59,

E11.44, E13.351, E13.37X9, E10.3552, E11.3213, E10.3499, E10.331, E10.3599, E11.354, E10.610, E11.37, E13.3553, E10.618, E13.610, E10.11, E10.354, E13.3521, E13.621, E13.3412, E10.64, E13.3413, E11.3543, E11.3559, E11.3551, E13.61, E10.3541, E11.34, E13.3532, E10.3542, E13.3539, E10.321, E10.3492, E11.3513, E13.2, E10.3532, E10.3539, E13.3551, E11.3399, E10.8, E11.3549, E13.3, E13.8, E10.3211, E11.319, E11.3311, E13.352, E11.3533, E11.64, E11.3413, E11.3, E11.39, E13.3399, E13.3531, E13.42, E10.329, E10.3399, E10.3292, E11.311, E13.3419, E13.43, E13.3549, E13.52, E11.3592, E13.3559, E13.37X3, E10.352, E10.359, E10.51, E10.3412, E10.3491, E11.641, E13.641, E11.352, E11.649, E13.29, E11.4, E13.3513, E10.3559, E10.37X1, E11.3599, E13.3391, E13.3411, E11.353, E13.3299, E13.65, E13.3529, E10.41, E11.3593, E11.37X1, E11.3511, E13.63, E11.3292, E13.39, E11.8, E13.62, E11.3591, E10.59, E11.3492, E11.3212, E13.339, E10.3411, E11.3491, E10.349, E11.3531, E11.63, E13.3313, E13.3311, E11.3541, E11.10, E13.3312, E11.43, E11.61, E10.37X3, E13.32, E13.3533, E13.354, E11.3529, E10.21, E11.640, E13.64, E13.3491, E10.3519, E11.3412, E11.349, E11.359, E11.41, E10.3392, E10.62, E10.339, E10.39, E13.353, E11.3313, E11.29, E11.3319, E11.3523, E13.3211, E11.42, E13.3552, E10.3391, E11.3211, E11.331, E10.31, E11.3519, E10.1, E11.3552, E10.620, E11.3542, E10.34, E11.3499, E10.4, E11.3521, E13.341, E13.3512, E11.65, E10.3293, E10.3521, E10.630, E10.649, E10.3529, E11.3291, E11.3532, E13.3492, E13.5, E10.61, E11.321, E13.3493, E10.3213, E11.3393, E13.37X1, E13.649, E11.3411, E13.44, E13.51, E11.3293, E10.5, E11.329, E11.6, E11.621, E11.3312, E11.35, E13.329, E10.3299, E10.37X9, E11.3392, E13.3543, E11.3512, E10.3313, E13.321, E13.3523, E13.355, E11.610, E10.10, E10.22, E10.42, E10.32, E11.37X9, E11.3219, E11.628, E13.331, E11.339, E13.3499, E10.3533, E11.1, E13.628, E10.3513, E13.69, E10.3212, E10.353, E13.319, E10.52, E10.40, E10.63, E11.630, E13.3292, E11.36, E13.31, E13.640, E10.37X2, E10.638, E10.65, E11.620, E10.3, E10.3512, E11.3522, E11.3391, E10.3593, E10.640, E10.69, E11.22, E11.31, E13.3219, E10.3592, E13.36, E10.36, E11.37X3, E13.40, E10.621, E13.311, E13.3511, E10.3511, E11.618, E10.311, E11.638, E10.49, E10.3549, E10.3291, E10.3591, E13.49, E13.34, E11.69, E13.35, E13.620, E13.638, E10.3311, E13.41, E10.33, E13.3522, E11.37X2, E13.22, E13.3541, E10.3531, E10.628, E10.641, E11.32, E10.3551, E11.351, E13.630, E13.3293, E13.622, E13.3213

ICD9CM: 250.93, 250.42, 250.83, 250.70, 250.92, 250.40, 250.43, 250.50, 250.4, 250.51, 250.90, 250.61, 250.7, 250.82, 250.91, 250.60, 250.71, 250.52, 250.73, 250.53, 250.63, 250.41, 250.6, 250.80, 250.62, 250.81, 250.72, 250.9, 250.8, 250.5

SNOMED CT: 426875007, 421437000, 731000119105, 1501000119109, 137941000119106, 127013003, 427134009, 81830002, 314010006, 138881000119106, 16745131000119100, 23045005, 238982009, 734022008, 190330002, 609575003, 769218003, 860798008, 73211009, 420279001, 609579009, 201252003, 368561000119102, 10656271000119102, 2751001, 232022001, 721088003, 368741000119105, 49817004, 237652003, 31321000119102, 9859006, 49455004, 16697521000119104, 190389009, 771000119108, 237651005, 199228009, 816067005, 71721000119101, 138591000119106, 16891191000119108, 190410002, 237620003, 719216001, 720519003, 16747021000119109, 59276001, 408411003, 609577006, 368721000119104, 421966007, 87461000119100, 609581006, 127012008, 420789003, 424736006, 735539005, 770582001, 28331000119107, 87471000119106, 721201000124104, 190411003, 713702000, 408410002, 39127005, 75682002, 199226008, 72021000119109, 870420005, 140101000119109, 1196923000, 421326000, 18521000119106, 138621000119108, 140381000119104, 368141000119106, 709147009, 90751000119109, 860980004, 399865004, 230574001, 237613005, 609580007, 722454003, 200687002, 789585000, 120711000119108, 420514000, 16747901000119104, 399863006, 140421000119108, 16747101000119101, 71421000119105, 104961000119108, 609572000, 703136005, 87441000119104, 1285518004, 199223000, 232020009,

367241000119104, 90741000119107, 90791000119104, 87921000119104, 71701000119105, 311366001, 769182000, 16747141000119104, 770324004, 126535008, 751000119104, 16745931000119102, 770097006, 722206009, 90781000119102, 105401000119101, 1259048007, 126551000119103, 16749661000119102, 21584002, 111556005, 128001000119105, 140391000119101, 16697471000119102, 368421000119108, 816962002, 860883001, 768792007, 769219006, 723074006, 137931000119102, 367261000119100, 16745531000119108, 368051000119109, 75524006, 237611007, 314893005, 770600002, 312908007, 724876003, 860979002, 237617006, 50620007, 399876000, 408539000, 421725003, 197605007, 1571000119104, 40801000119106, 769181007, 16746661000119106, 199227004, 361216007, 232021008, 312909004, 770094004, 1491000119102, 110141000119100, 71791000119104, 422034002, 140121000119100, 10754881000119104, 312903003, 789571005, 120731000119103, 90761000119106, 16749781000119101, 26298008, 420825003, 82551000119103, 368181000119101, 79554005, 368451000119100, 237603002, 769184004, 104951000119106, 109171000119104, 87491000119107, 237604008, 408414006, 445170001, 713704004, 126534007, 789568002, 72041000119103, 724067006, 10661671000119102, 368201000119100, 129599004, 769244003, 16746741000119102, 138941000119105, 39058009, 735538002, 16747741000119100, 721284006, 733072002, 71771000119100, 230577008, 193183000, 441628001, 40791000119105, 243421000119104, 190331003, 371087003, 408413000, 16748741000119101, 312904009, 140531000119105, 420436000, 425455002, 737212004, 10753491000119101, 11687002, 313435000, 421075007, 762489000, 110171000119107, 10660471000119109, 267604001, 314903002, 420422005, 126591000119108, 314537004, 420756003, 769220000, 90731000119103, 399877009, 420918009, 422228004, 38046004, 46635009, 16746261000119108, 420683009, 368471000119109, 170766006, 238984005, 860721006, 110181000119105, 769185003, 38205001, 237619009, 312905005, 140131000119102, 1217674007, 138921000119104, 16745611000119102, 890171006, 57886004, 691000119103, 60971000119101, 127991000119101, 16745891000119105, 769217008, 31211000119101, 72031000119107, 82571000119107, 103981000119101, 609566000, 367991000119101, 123763000, 10656231000119100, 713706002, 368171000119104, 310505005, 421986006, 16746341000119103, 399873008, 102621000119101, 230575000, 311782002, 96441000119101, 237599002, 1511000119107, 768794008, 90721000119101, 426705001, 44054006, 16891151000119103, 314014002, 84361000119102, 193350004, 60951000119105, 60961000119107, 8801005, 782755007, 721121000124104, 71441000119104, 368711000119106, 769186002, 770323005, 25412000, 48951005, 193489006, 783722008, 724136006, 420996007, 735200002, 138611000119101, 42954008, 310387003, 390834004, 860978005, 421895002, 1217044000, 399866003, 414894003, 421305000, 82541000119100, 70694009, 399862001, 735537007, 82980005, 237635002, 762291006, 741000119101, 770599000, 368441000119102, 111307005, 190368000, 712883005, 769245002, 201724008, 870529009, 609576002, 445260006, 97331000119101, 140521000119107, 164881000119109, 1144894004, 190407009, 721283000, 421920002, 16746941000119104, 1255271005, 193184006, 428896009, 770362001, 822995009, 368521000119107, 399868002, 609573005, 711000119100, 157141000119108, 16748141000119100, 782825008, 16745211000119100, 16746901000119101, 703137001, 788878000, 75022004, 419100001, 421750000, 761000119102, 60991000119100, 190372001, 276560009, 408409007, 789572003, 126571000119107, 703138006, 706894000, 138911000119106, 421365002, 104941000119109, 201251005, 769191001, 871778008, 701000119103, 770095003, 770765001, 39181008, 230579006, 609567009, 368541000119101, 768793002, 25093002, 313436004, 421256007, 789569005, 41911000119107, 199229001, 238981002, 106281000119103, 768799003, 770098001, 368551000119104, 16747341000119101, 237632004, 609563008, 368581000119106, 74627003, 193185007, 770581008, 816178004, 5368009, 402864004, 789562001, 16746131000119105, 237601000, 816177009, 138811000119100, 59079001, 609562003, 609565001, 420270002, 111231000119109, 769188001, 190412005, 51002006, 422126006, 1521000119100, 1196922005, 792926007, 119831000119106, 312907002, 420486006, 427571000, 368251000119101, 193349004, 399875001, 421847006, 199225007, 201723002, 43959009, 309426007, 816961009, 16745051000119107, 368151000119108, 1172389004, 76751001, 238983004, 427027005, 201250006, 609568004, 237621004,

280137006, 421631007, 609578001, 97621000119107, 127014009, 314902007, 721000119107, 16747661000119109, 408412005, 408540003, 427943001, 716362006, 138901000119108, 359642000, 4855003, 769190000, 769222008, 19378003, 422166005, 713703005, 201000119106, 16697421000119103, 420715001, 713457002, 84371000119108, 768798006, 769183005, 4783006, 609570008, 1481000119100, 193141005, 721111000124107, 312906006, 314011005, 399872003, 140111000119107, 140411000119101, 1251386008, 33559001, 368461000119103, 1217068008, 232023006, 417677008, 421893009, 428007007, 609569007, 368101000119109, 46894009, 62260007, 422183001, 530558861000132104, 190447002, 739681000, 24203005, 28032008, 199230006, 421468001, 399870006, 237600004, 420662003, 72051000119101, 367391000119102, 126531000119109, 138891000119109, 140401000119104, 770766000, 237612000, 420868002, 422275004, 1285519007, 63510008, 190406000, 237627000, 713705003, 72061000119104, 82581000119105, 399864000, 609574004, 35777006, 422088007, 426907004, 1551000119108, 769221001, 5969009, 127011001, 230576004, 704241002, 72141000119104, 90771000119100, 16745651000119101, 190416008, 860977000, 367211000119103, 190388001, 395204000, 413183008, 427089005, 609571007, 414910007, 707221002, 789567007, 769187006, 91352004, 770096002, 102781000119107, 791000119109, 609561005, 609564002, 781000119106, 368431000119106, 230572002, 421779007, 770361008, 236500003, 237633009, 399869005, 312910009, 398140007, 60981000119103, 368591000119109, 1003605004, 312912001, 789570006, 1531000119102, 16745411000119104, 359611005, 716020005, 97341000119105, 422099009, 314015001, 1142044000, 199231005, 422014003, 789542009, 870421009, 111552007, 399874002, 712882000, 768797001, 1197592001, 81531005, 236499007, 441656006, 399871005, 368601000119102, 871781003, 15936501000119100

#### 3.11 Diabetes without Complications

ICD10CM:
E11.11, E11.01, E11.0, E10.11, E13.00, E13.01, E11.10, E13.10, E13.1, E13.0, E13.11, E10.1, E10.10, E11.1, E10.9, E11.00, E13.9

ICD9CM:
250.93, 251.1, 253.6, 250.42, 250.11, 250.3, 250.83, 250.70, 252.1, 251.4, 250.92, 250.40, 250.43, 251.8, 253.9, 250.30, 250.50, 253.7, 250.21, 250.31, 250.4, 250.51, 252.00, 250.2, 251, 250.03, 250, 251.9, 250.22, 253.2, 250.0, 250.90, 250.61, 250.7, 250.82, 252.0, 252, 253.5, 250.91, 250.60, 250.1, 250.71, 250.52, 250.73, 253.8, 250.20, 253, 253.4, 250.53, 251.5, 252.8, 250.10, 250.32, 253.0, 251.0, 250.63, 253.1, 250.00, 250.41, 252.02, 250.6, 250.12, 250.80, 250.62, 250.81, 250.01, 252.9, 250.13, 250.72, 250.9, 251.2, 250.02, 250.8, 251.3, 253.3, 250.23, 250.5, 250.33, 252.08, 252.01

SNOMED CT:
426875007, 23045005, 734022008, 609575003, 73211009, 609579009, 2751001, 721088003, 49817004, 237652003, 31321000119102, 190389009, 237651005, 199228009, 816067005, 190410002, 720519003, 609577006, 609581006, 127012008, 721201000124104, 75682002, 199226008, 709147009, 237613005, 609580007, 722454003, 609572000, 703136005, 1285518004, 199223000, 722206009, 105401000119101, 111556005, 75524006, 237611007, 237617006, 408539000, 40801000119106, 199227004, 10754881000119104, 26298008, 237603002, 237604008, 724067006, 733072002, 40791000119105, 737212004, 10753491000119101, 11687002, 313435000, 421075007, 420422005, 46635009, 237619009, 890171006, 57886004, 609566000, 123763000, 237599002, 426705001, 44054006, 8801005, 782755007, 721121000124104, 783722008, 138611000119101, 42954008, 1217044000, 70694009, 111307005, 190368000, 609576002, 445260006, 190407009, 1255271005, 368521000119107, 609573005, 782825008, 703137001, 75022004, 421750000, 190372001, 276560009, 703138006, 609567009, 313436004, 199229001, 106281000119103, 609563008,

5368009, 237601000, 59079001, 609562003, 609565001, 420270002, 190412005, 51002006, 421847006, 199225007, 76751001, 609568004, 609578001, 408540003, 716362006, 359642000, 4783006, 609570008, 1481000119100, 14735008, 721111000124107, 33559001, 1217068008, 609569007, 368101000119109, 46894009, 530558861000132104, 190447002, 24203005, 28032008, 199230006, 237600004, 367391000119102, 237612000, 1285519007, 190406000, 237627000, 609574004, 5969009, 190416008, 413183008, 427089005, 609571007, 91352004, 609561005, 609564002, 1142044000, 199231005, 111552007, 1197592001, 81531005

Truveta:
1215938

#### 3.12 Drug Abuse

ICD10CM:
F12.222, F13.230, F15.121, F13.91, F12.980, F14.9, F15.94, F18.24, F12.950, F13.232, F16.180, F12.22, F14.95, F19.99, F11.921, F13.239, F15.251, F11.259, F16.24, F12.95, F15.181, F15.988, F19.27, F19.930, F19.98, F13.180, F14.180, F14.23, F18.15, F16.29, F15.222, F16.951, F11.92, F13.2, F13.22, F13.90, F15.20, F11.222, F11.981, F13.20, F15.22, F18.151, F12.229, F13.281, F16.1, F18.188, F19.159, F16.25, F13.14, F18.259, F11.28, F11.920, F13.182, F14.221, F19.92, F12.1, F15.182, F11.221, F11.988, F15.129, F19.239, F13.950, F15.950, F19.921, F19.151, F11.951, F12.951, F14, F16.10, F16.920, F18.129, F18.988, F19.1, F11.95, F13.150, F12.2, F11.229, F14.22, F15.29, F12.19, F14.288, F15.929, F15.229, F16.19, F18.25, F19.90, F19.951, F19.929, F19.939, F13.96, F18.11, F13.931, F14.18, F14.181, F14.922, F19.129, F19.22, F13.282, F15.920, F11.122, F13.188, F19.950, F18.220, F19.222, F14.222, F13.982, F15.18, F18.27, F11.182, F15.24, F18.20, F19.21, F13.97, F14.982, F15.2, F16.120, F12.988, F13.21, F13.981, F18.920, F13.951, F13.139, F11.98, F14.921, F15.15, F18.959, F19.130, F16.251, F11.10, F14.250, F19, F14.28, F12.98, F12.121, F13.19, F19.131, F19.139, F19.23, F13.28, F14.981, F15.982, F19.280, F11.121, F13.129, F13.27, F18.951, F16.92, F11.11, F11.94, F12.93, F15.93, F16.121, F12.188, F12.251, F12.280, F15.14, F19.922, F12.11, F12.25, F13.939, F15.280, F16.98, F13.250, F14.90, F16.14, F11.220, F16.980, F19.97, F12.15, F19.231, F12.13, F11.23, F12.29, F13.131, F11.24, F18.159, F19.988, F14.12, F18.2, F19.230, F12.92, F13.121, F14.150, F13.95, F15.159, F15.92, F19.16, F15.951, F16.28, F19.18, F19.182, F14.25, F14.99, F16.12, F12.91, F15.19, F19.9, F11.14, F15.12, F15.188, F12.250, F18.17, F19.981, F12.21, F13.288, F16.283, F13.930, F13.988, F18.280, F11.93, F18.95, F19.180, F12.129, F16.9, F12.90, F13.26, F15.98, Z71.5, F15.959, F16.90, F19.132, F18.94, F11.12, F19.121, F13.13, F16.129, F18.98, F19.19, F12.122, F13.280, F13.980, F11.922, F18.921, F19.120, F13.99, F14.10, F11.282, F18.12, F11.250, F12.180, F15.180, F14.951, F13.1, F15.95, F16.950, F19.150, F14.282, F15.10, F16.183, F19.188, F14.120, F14.14, F14.15, F15.21, F18.250, F12.18, F12.20, F14.92, F18.97, Z71.52, F11.129, F13.18, F18.10, F18.229, F11.120, F13.29, F18.150, F19.13, F13.98, F14.121, F16.122, F19.20, F19.11, F11.929, F14.2, F18.251, F19.2, F19.281, F13.130, F12.922, F13, F19.25, F12.921, F13.959, F12.151, F15.120, F12.120, F14.1, F18.9, F13.10, F19.181, F19.15, F19.259, F11.29, F14.280, F19.229, F19.95, F11, F14.182, F13.11, F15.221, F16.188, F13.94, F16.2, F11.21, F12.220, F11.2, F14.251, F12.221, F12.23, F16.220, F12.150, F13.93, F14.988, F15.220, F15.23, F16.22, F16.988, F19.91, F11.251, F11.150, F15.921, F19.94, F11.13, F15.9, F16.250, F19.28, F12.99, F15.150, F15.980, F18.29, F19.24, F15.981, F18.1, F13.132, F15.90, F18.99, Z71.51, F12.920, F14.11, F15.259, F11.91, F11.25, F18.221, F15.250, F15.281, F16.11, F18.288, F14.24, F11.288, F13.259, F13.929, F14.98, F15.288, F19.282, F16.91, F15.91, F13.12, F13.181, F13.92, F14.281, F14.29, F11.281, F19.17, F19.250, F14.159, F19.10, F19.251, F13.25,

F13.9, F16.21, F18.21, F19.932, F14.21, F15.922, F16.259, F18.120, F11.959, F14.950, F16, F12.12, F13.24, F14.220, F16.94, F16.959, F19.220, F16.280, F16.921, F12.288, F15.25, F11.20, F16.18, F14.959, F18.980, F19.12, F13.921, F14.19, F14.920, F15.282, F16.288, F19.920, F14.91, F19.29, F11.159, F13.15, F14.20, F14.929, F16.151, F16.983, F19.93, F19.982, F11.181, F18.91, F14.980, F11.18, F11.982, F13.159, F18.28, F15.13, F18.180, F13.231, F19.122, F19.931, F13.23, F16.150, F14.94, F19.232, F11.9, F12.9, F13.251, F15.28, F18.19, F19.96, F14.13, F12.259, F19.14, F19.288, F11.15, F11.90, F12.10, F13.221, F13.229, F19.980, F13.120, F13.932, F15.122, F13.151, F11.19, F11.188, F16.221, F18.22, F15.11, F18.92, F19.959, F15.99, F19.221, F14.188, F14.259, F18.929, F11.1, F12.28, F12.929, F14.229, F16.15, F16.929, F14.151, F16.95, F18.950, F11.950, F13.220, F13.920, F19.26, F14.129, F16.99, F18, F18.14, F14.122, F15, F11.151, F12, F18.18, F14.93, F15.151, F16.20, F15.1, F16.159, F16.229, F18.90, F11.22, F11.99, F12.159, F12.959, F18.121

ICD9CM:
304.00, 305.21, 304.6, 304.83, 304.52, 304.63, 304.82, 304.3, 305.20, 305.9, 292.84, 305.30, 305.23, 305.70, 305.82, 292.1, V65.42, 304.2, 304.12, 305.43, 304.93, 305.41, 305.92, 304.41, 304.51, 304.03, 305.63, 305.71, 292.11, 292.0, 304.92, 304.31, 304.01, 305.73, 304.91, 304.13, 292.2, 304.62, 305.22, 305.93, 292.12, 304.71, 304.80, 305.80, 305.51, 305.50, 305.83, 304.4, 292.8, 292.82, 305.62, 292.81, 292.83, 304.21, 292.9, 305.31, 304.81, 305.72, 304.33, 304.0, 305.60, 304.60, 304.50, 304.42, 304.5, 305.81, 304.22, 304.73, 305.3, 304.10, 304.72, 292, 305.40, 304.43, 304, 305.5, 304.7, 304.70, 305.8, 304.02, 305.91, 304.20, 304.30, 304.61, 304.40, 292.89, 305.6, 305.2, 304.9, 305.32, 305.61, 304.53, 304.32, 304.11, 304.23, 304.1, 305.4, 305.53, 305.90, 305.52, 304.90, 305.33, 292.85, 305.42, 304.8, 305.7

SNOMED CT:
47774001, 290169000, 290247003, 290606005, 291034008, 291053003, 291139000, 291162003, 291266009, 291590009, 291610004, 291997002, 292154009, 292382006, 292454003, 292887005, 293385003, 293443003, 295334008, 295534007, 295615001, 295832004, 296129007, 296162004, 296248000, 296472007, 296533005, 296570001, 296649002, 296651003, 296786005, 418475009, 427640001, 717889000, 737334000, 871572007, 328301000119102, 1157257000, 20871009, 41461004, 46604003, 64600001, 83367009, 84584008, 89137004, 95911005, 218488009, 230800004, 290231000, 290255005, 290426000, 290651005, 290813007, 290995002, 291094006, 291196005, 291218002, 291425006, 291688004, 291839002, 291881008, 292350008, 292374006, 292718001, 293195004, 293555007, 295547002, 295699008, 295831006, 296115006, 296281004, 296358004, 296384005, 296385006, 296636007, 296932006, 297023004, 297036004, 403655004, 419347003, 427046006, 871569000, 288851000119106, 1156753007, 15277004, 42423000, 61544007, 89451009, 191493005, 191837001, 290228001, 290616002, 291379003, 291611000, 291841001, 292960005, 293568004, 295219000, 295345006, 295364000, 295490003, 295654000, 295819009, 296042005, 296186002, 296250008, 296433008, 296550007, 296591009, 296747006, 296764009, 296795002, 403658002, 724662001, 762330008, 870409005, 153501000119105, 18311006, 35776002, 41106001, 47923004, 74851005, 194791005, 216519004, 216633005, 218969006, 290134002, 290289006, 290372000, 290751000, 290781008, 290786003, 290798008, 291185002, 291254003, 291280002, 291785008, 292179007, 292210007, 292312002, 292675000, 293049008, 293502001, 293553000, 295296006, 295303009, 295409009, 295431001, 295743001, 295841009, 296026003, 296583000, 296757007, 296849000, 419055000, 704055002, 713524002, 713775002, 722321001, 762531008, 788542004, 306181000119104, 28926001, 293544007, 41309000, 42344001, 55967005, 79044002, 212695003, 233687002, 237640005, 290137009, 290459002, 290549001, 290560005, 290960000, 291042009, 291065004, 291166000, 291321003, 292252002, 292522009, 292805004, 292955006, 293223006, 293383005, 295332007, 295407006, 295500006, 295620001, 295717002, 295725000, 295730001, 295980002, 296089007, 296130002, 296513006,

296623007, 296682004, 296705007, 402746001, 403646002, 416073003, 716005004, 871679007, 298601000119107, 62473009, 78358001, 191891003, 207195004, 290304000, 290515009, 290822008, 290837007, 291207002, 291355000, 291408009, 291538000, 291700001, 291847002, 291908003, 292153003, 292318003, 292474005, 292487004, 292867009, 293038002, 293380008, 293436009, 293538008, 295505001, 295661001, 295918002, 296112009, 296847003, 296924003, 296943003, 296970005, 297011005, 308374001, 314422000, 418328005, 724722007, 292992006, 1144817005, 1144836000, 21511000, 53936005, 78695006, 109804004, 212771008, 218697003, 218794000, 235881000, 290173002, 290384003, 290493003, 290648003, 290803004, 290850008, 291208007, 291323000, 291375009, 291545000, 291588008, 292351007, 292380003, 292504006, 292547006, 292840003, 293036003, 293064008, 293310001, 295184007, 295412007, 295415009, 295484002, 295535008, 295537000, 295789008, 296013002, 296435001, 296481001, 296488007, 296789003, 296793009, 296833002, 296880005, 724666003, 762328006, 461231000124109, 448178009, 1148671008, 7184000, 51814004, 110284009, 185320006, 191888003, 212596008, 237509009, 238894004, 290293000, 290705001, 290871003, 291054009, 291170008, 291682003, 291880009, 291948006, 292554000, 292855007, 295464001, 295794008, 296068003, 296187006, 296584006, 296979006, 403629000, 711545001, 724657002, 735235000, 871521003, 15921411000119106, 1148543009, 1323006, 11659006, 12898000, 72057009, 191475009, 218635008, 290487006, 290506002, 290526003, 290623001, 290762000, 291261004, 291607006, 291850004, 291899000, 292130004, 292565005, 295123003, 295517004, 295634001, 295662008, 295964007, 295994001, 296116007, 416093006, 425885002, 21841000119100, 1163356007, 726007, 5189009, 18689007, 50687007, 57305009, 69956002, 91014009, 206018005, 218951002, 231462006, 235650007, 284591009, 290260009, 290412009, 290936004, 291050000, 291308005, 291428008, 291527000, 291601007, 291614008, 292017003, 292399007, 292442003, 293499000, 293518007, 293572000, 295362001, 295405003, 295576004, 295775003, 295787005, 295931007, 295966009, 296133000, 296191001, 296450005, 296631002, 296642006, 296738003, 296801006, 712742003, 724727001, 789560009, 298531000119104, 678421000119106, 295325003, 878878007, 1149333003, 1153338005, 4771009, 22583005, 30365001, 42373003, 191360002, 199255000, 216471009, 230682007, 290300009, 290759003, 291510008, 291543007, 291615009, 292192005, 292217005, 292711007, 293146004, 293454008, 295200005, 295352008, 295501005, 295577008, 295578003, 295690007, 295807001, 295814004, 296027007, 296046008, 296224009, 296492000, 296633004, 296790007, 296978003, 297101001, 870749003, 871565006, 306171000119102, 680531000119103, 15999391000119104, 84483000, 111760006, 191883007, 212674008, 212694004, 216560006, 235619005, 269269007, 291566008, 291635005, 291687009, 291690003, 292401001, 292725008, 292727000, 292967008, 295361008, 295783009, 296405008, 296448002, 296585007, 296713008, 297017009, 403650009, 720683004, 722200003, 724523000, 1149407007, 18108002, 34917002, 62052002, 191483003, 216551003, 280983004, 290135001, 290845002, 291158009, 291337009, 291442000, 292293005, 292495000, 292875003, 293172008, 293217001, 295193008, 295268000, 295530003, 295567006, 295623004, 296053004, 296377002, 296875001, 296882002, 296901005, 418949004, 713694002, 762419009, 306571000119106, 56043005, 1179727001, 34085004, 45701006, 58104004, 95919007, 111350000, 290235009, 290530000, 290684005, 291361002, 291364005, 291620009, 292132007, 292352000, 292462006, 292691002, 292742002, 295328001, 295680004, 295700009, 295901001, 296005003, 296180008, 296749009, 296872003, 419219000, 700425005, 788863007, 331981000119104, 1153364004, 1153541000, 70932007, 72182007, 84811001, 95247003, 102448004, 206129006, 216585002, 237809000, 242831003, 290680001, 290890006, 290915001, 291052008, 291182004, 291286008, 291393005, 291467002, 291604004, 291627007, 291664001, 291986002, 292304005, 293004005, 295267005, 296458003, 296788006, 398228004, 724715004, 870410000, 16238741000119105, 10070005, 292739008, 448090008, 1686006, 40382002, 57319009, 85005007, 216503006, 216615004, 241960008, 290492008, 291887007, 292069008, 292137001, 292149000, 292309000, 292735002, 293471009, 295160004, 295300007, 295724001, 295737003, 295932000, 296031001, 296277004, 296279001, 296334003, 296511008, 296655007, 296729000, 297081000, 314849005, 386472008, 402428008, 715059007, 7665002, 129660004, 1144820002, 471771000124106, 7201003,

36784008, 54594006, 61104008, 191844005, 191850000, 191853003, 191920007, 218533007, 290209008, 290513002, 290969004, 291309002, 291330006, 291435000, 291930002, 292879009, 292925004, 293279008, 295539002, 295561007, 295686005, 295718007, 296660006, 296666000, 296969009, 297083002, 309742004, 312958000, 397841007, 762507003, 306561000119100, 15920041000119105, 191873008, 235944000, 290386001, 290690009, 290821001, 290911005, 291295000, 292029007, 292605001, 293560006, 295335009, 296173006, 296270002, 296329002, 296707004, 296863006, 297006008, 418579000, 698313007, 46193001, 11387009, 38879000, 40550004, 67933004, 86624000, 206023005, 212758005, 216530001, 216596005, 218471002, 290151009, 291267000, 291347007, 291569001, 292166003, 292629008, 293008008, 293012002, 293337004, 295240004, 295440002, 295555009, 296001007, 296037002, 296553009, 296587004, 296806001, 296873008, 296914006, 297022009, 371341003, 403639006, 416714005, 440068009, 27051000119102, 91561000119109, 17231009, 28348003, 109861009, 216627001, 218813003, 230315008, 290238006, 290288003, 290364002, 290701005, 290949009, 293470005, 293535006, 295247001, 295568001, 295655004, 296054005, 296062002, 296167005, 296413009, 296805002, 296844005, 297060000, 297941009, 361148000, 425604002, 713776001, 723653009, 724678004, 724704009, 724708007, 762510005, 866149003, 306551000119102, 15919781000119104, 1142050005, 24882007, 30897009, 48550003, 102449007, 209099002, 216494005, 290166007, 290174008, 290532008, 290536006, 290628005, 291599005, 292085003, 293074006, 293289007, 293473007, 295712008, 295776002, 296066004, 296466007, 296581003, 296800007, 296940000, 296959003, 439740005, 720681002, 35279005, 111491000119107, 200907003, 212533002, 230353003, 234645009, 290391000, 290433000, 290982007, 291179009, 291247009, 291263001, 291389004, 291817004, 292027009, 292223000, 292637000, 293075007, 295156002, 295283005, 295306001, 295356006, 295389006, 295613008, 295659005, 296038007, 296188001, 296904002, 297053006, 297073001, 403672006, 713496008, 724711008, 871513009, 133301000119102, 403622009, 9291000, 55680006, 57005003, 87816000, 212643006, 237811009, 239849004, 281510005, 290278004, 290398006, 290463009, 290665007, 290681002, 290742008, 290843009, 290967002, 291229003, 291414002, 291491006, 292240009, 292261002, 292340007, 292651000, 292974003, 293516006, 295455006, 295678005, 295874009, 296259009, 296699003, 297086005, 308335008, 312098001, 313260003, 135331000119105, 34743008, 45421006, 52382004, 67438002, 68341005, 212641008, 238989000, 290745005, 290966006, 291049000, 291165001, 291282005, 291291009, 291596003, 291891002, 291953001, 291969008, 292282008, 292414000, 293135002, 293164006, 293349003, 295242007, 295682007, 296225005, 296307001, 296432003, 296625000, 296986003, 713370005, 724664000, 762517008, 10838971000119103, 470701000124108, 10984003, 25980005, 26532007, 66698002, 81711008, 191937000, 212763009, 237228001, 290199009, 290354006, 290401009, 290418008, 290448006, 290561009, 290610008, 290818003, 290830009, 290930005, 290988006, 291285007, 291591008, 291825002, 291958005, 292127006, 292170006, 292438001, 292748003, 293165007, 295145002, 296125001, 296160007, 296367004, 296460001, 296726007, 312959008, 402662002, 403654000, 724671005, 724723002, 762388009, 1148696006, 1149353004, 1162818005, 35224002, 212807002, 290294006, 290841006, 291107007, 291234004, 291492004, 291802003, 291898008, 291954007, 292280000, 292334000, 292453009, 292494001, 292497008, 292533007, 292853000, 293058001, 293206002, 293280006, 295167001, 295243002, 295275004, 295597003, 296122003, 296157000, 296190000, 296201002, 296343007, 296573004, 406547006, 724705005, 762576008, 838526006, 125851000119106, 897046009, 1144816001, 1149469007, 1230075001, 1973000, 32371003, 65941003, 71328000, 201436003, 216591000, 230665005, 235751001, 275400004, 290198001, 290363008, 290396005, 290507006, 290572004, 290765003, 290829004, 290990007, 291306009, 292139003, 292327002, 292333006, 292346001, 292923006, 293239004, 295759001, 295847008, 295887003, 296294002, 296389000, 296753006, 442406005, 698493007, 870621003, 871676000, 306151000119106, 16623961000119100, 14002008, 31108002, 53126001, 89183005, 169465000, 235130007, 238635004, 290466001, 290670000, 290731004, 290766002, 290876008, 291226005, 291697000, 291857001, 291862000, 291979005, 292047001, 292202006, 292913002, 293483006, 295746009, 296158005, 296312000, 296447007, 296675003, 300993007, 402659000, 838336002, 1155737004, 22574000,

111480006, 212509008, 290215008, 290689000, 290901003, 292090000, 292193000, 292204007, 292391005, 292544004, 292865001, 293302002, 293453002, 295136003, 295196000, 295747000, 295897007, 296567000, 296892005, 297005007, 737341006, 18597007, 21647008, 44340006, 50357006, 51339003, 212761006, 233673002, 280985006, 290193005, 290194004, 290210003, 290441000, 290711003, 290810005, 290918004, 290999008, 291878003, 291987006, 292112006, 292390006, 292405005, 292409004, 292799005, 292802001, 295166005, 295441003, 295672006, 295683002, 295840005, 296006002, 296748001, 762346000, 871573002, 1162824004, 10916006, 78267003, 190283008, 212658003, 212737004, 218808002, 218972004, 231474005, 233725001, 288836004, 290203009, 290264000, 290608006, 290734007, 290758006, 290820000, 290838002, 291490007, 291679008, 291783001, 291855009, 292398004, 293446006, 295480006, 295481005, 295486000, 295858004, 295877002, 295951006, 296159002, 296874002, 370511006, 698402008, 724574008, 724729003, 785277001, 23601000119102, 298541000119108, 1145067005, 1148724006, 328371000119107, 13151001, 21782001, 31715000, 43125006, 59274003, 85871000, 191932006, 216487007, 216526004, 216527008, 290636001, 291384009, 291433007, 291450009, 291515003, 291624000, 291649001, 291853002, 291983005, 292126002, 292254001, 292457005, 292483000, 292508009, 292682001, 292904004, 293450004, 293552005, 295800004, 295825008, 295987004, 296698006, 296868002, 297008009, 428676003, 698492002, 866204005, 10625831000119107, 15735361000119109, 1179732000, 93435005, 129662007, 238999005, 290145004, 290232007, 290270006, 290287008, 290494009, 290509009, 290676006, 291018005, 291117002, 291791005, 291819001, 292013004, 292174002, 292233008, 292292000, 292535000, 292863008, 293010005, 293496007, 295621002, 295667002, 295803002, 296394000, 296670008, 296858007, 314539001, 438515009, 698704008, 713281006, 860826006, 306581000119109, 41234002, 71320007, 1149239002, 25599007, 72700005, 80258006, 83864008, 212715006, 290462004, 290795006, 290802009, 291362009, 291378006, 291593006, 291831004, 291932005, 292256004, 292372005, 293043009, 293056002, 295805009, 295853008, 295946000, 296123008, 296517007, 296864000, 297098000, 315205008, 402754004, 698307003, 698310005, 735456000, 788339009, 298611000119105, 1149229008, 18653004, 48445006, 57310008, 73340005, 74984006, 216500009, 216550002, 218356002, 235874007, 237908009, 254247000, 269723006, 290775000, 290801002, 290861000, 290885008, 290925006, 291274005, 291377001, 291400002, 291456003, 291567004, 291907008, 292006004, 292142009, 292224006, 292449007, 292777002, 292938007, 293248009, 293351004, 295170002, 295466004, 295684008, 295739000, 295896003, 296140004, 296302007, 296519005, 296600009, 296877009, 417360004, 16000231000119105, 1148539005, 1148673006, 19303008, 45088005, 50589003, 86733006, 191833002, 212706004, 241756003, 242828004, 290248008, 290283007, 290310000, 290423008, 290477007, 290887000, 290905007, 291066003, 291562005, 292032005, 292368006, 292540008, 293472002, 293576002, 295205000, 295551000, 295821004, 295915004, 295979000, 296045007, 296051002, 296182000, 296423000, 296590005, 296657004, 297942002, 426450003, 428931000124102, 281004, 50452004, 290149005, 290196002, 290404001, 290583008, 292034006, 292052006, 292284009, 292811001, 292977005, 293057006, 293390000, 295225001, 295260007, 295278002, 295317000, 295369005, 296093001, 296520004, 296656008, 296857002, 297032002, 420454001, 441244001, 441681009, 699064007, 699066009, 703850002, 720672008, 724682002, 765482002, 766219005, 838524009, 91461000119108, 15233006, 14700006, 28800007, 46673008, 50601005, 74338006, 199253007, 199549009, 290443002, 290624007, 290846001, 291144007, 291233005, 291602000, 291786009, 292375007, 292890004, 293160002, 295171003, 295371005, 295507009, 295579006, 295696001, 295722002, 295995000, 296189009, 296697001, 296769004, 296839003, 402750008, 724558003, 762302008, 306141000119109, 306611000119102, 44047000, 237694000, 1162817000, 46878002, 65223000, 185462000, 218643003, 290183003, 290695004, 290839005, 290886009, 290944004, 291273004, 291973006, 292033000, 292176000, 292288007, 292385008, 292465008, 292685004, 293293001, 295284004, 295573007, 296477001, 296836005, 297085009, 414996002, 699012001, 722139003, 724526008, 762518003, 762672001, 453441000124105, 10553007, 1145152008, 1149148001, 28213007, 28224009, 39994006, 63484008, 212778002, 235880004, 281036007, 290350002, 290739002, 290794005, 291411005,

291598002, 292084004, 292266007, 292478008, 292870008, 292922001, 293215009, 293242005, 293244006, 293464004, 295137007, 295234006, 295384001, 295720005, 295912001, 295942003, 296111002, 296156009, 296172001, 296216004, 296438004, 296480000, 296616006, 313183009, 417143004, 427975003, 440085006, 717127002, 22681004, 1145065002, 1163013005, 10327003, 46759008, 61351004, 73891003, 90755006, 212748006, 290732006, 291137003, 291409001, 291650001, 291812005, 292030002, 292118005, 292131000, 292151001, 292496004, 292752003, 292760002, 295531004, 295810008, 295950007, 296141000, 296664002, 296710006, 296783002, 296903008, 296908004, 296966002, 297063003, 400979004, 403673001, 702693007, 833317005, 870411001, 9982009, 51228004, 75122001, 77406008, 84393008, 108219001, 216514009, 230354009, 231461004, 234171009, 237545007, 237853005, 242259007, 277805004, 290656000, 291011004, 291429000, 291644006, 291965002, 292482005, 292988004, 292998005, 293141009, 293152003, 293226003, 293232008, 293317003, 293565001, 295423006, 295424000, 296292003, 296310008, 296420002, 296848008, 296957001, 297057007, 402751007, 838523003, 840300001, 12398201000119102, 7690000, 7895008, 448091007, 26617005, 38941006, 65425009, 238813009, 290156004, 290214007, 290217000, 290249000, 290708004, 290725002, 290939006, 290980004, 291112008, 291404006, 291846006, 292823007, 293073000, 295174006, 295468003, 295827000, 295905005, 296019003, 296069006, 296321004, 296374009, 296930003, 297084008, 297200009, 302895007, 417895009, 424338005, 711538001, 721710005, 830280003, 290592006, 292856008, 1017197007, 1144827004, 1145046005, 1157240004, 4688008, 12876009, 18323000, 85561006, 190284002, 277131000, 290155000, 290943005, 291021007, 291067007, 291184003, 291190004, 291239009, 292338002, 295141006, 295635000, 295693009, 295856000, 295872008, 296193003, 296306005, 296313005, 296339008, 296459006, 296773001, 403625006, 403632002, 723937009, 724655005, 724668002, 12398571000119105, 16270221000119108, 471751000124101, 4836003, 185387006, 216612001, 290365001, 290968007, 290996001, 291113003, 291345004, 291357008, 291554002, 291672004, 291796000, 291900005, 291909006, 292978000, 293021001, 295252006, 295742006, 296719007, 296770003, 302507002, 403614002, 403620001, 408667000, 420545001, 712950002, 713528004, 724656006, 316061000119106, 1148653001, 471761000124104, 1479971000168109, 20179009, 82047000, 191934007, 205791004, 206126004, 230334008, 290188007, 290309005, 290924005, 291063006, 291078004, 291209004, 291344000, 291513005, 291579004, 291608001, 292307003, 292311009, 292424008, 292952009, 295815003, 295833009, 296208008, 296264008, 296997003, 388975008, 403642000, 448177004, 11047881000119101, 49968000, 212523005, 212545006, 218727000, 234986004, 237798008, 290451004, 290933007, 291086004, 291528005, 291876004, 291955008, 292045009, 292066001, 292207000, 292249005, 292383001, 292428006, 292525006, 292668001, 292980006, 293194000, 293307008, 293421006, 295497000, 296149003, 296341009, 296386007, 296485005, 296606003, 296775008, 296861008, 414874007, 419914000, 724696008, 104521000119108, 290591004, 8837000, 23142005, 47318007, 52703008, 64595005, 212619008, 236520002, 291382008, 291858006, 292012009, 292328007, 292377004, 292420004, 292564009, 292729002, 293153008, 293346005, 295460005, 296228007, 296720001, 296732002, 297031009, 429299000, 431307001, 735458004, 788955005, 1145073006, 1149236009, 67725009, 191494004, 233706004, 290240001, 291010003, 291118007, 291524007, 291950003, 292500002, 292541007, 292704008, 292759007, 292851003, 293281005, 295194002, 295207008, 295540000, 295545005, 295676009, 296170009, 296262007, 296662003, 296668004, 297035000, 427488005, 724690002, 870715006, 433231000124100, 455441000124106, 16238181000119101, 1148533006, 1230074002, 18170008, 191813001, 238990009, 290280005, 290662005, 290797003, 291172000, 291390008, 291432002, 291453006, 291523001, 291546004, 291552003, 291585006, 291896007, 292995008, 293150006, 293173003, 293495006, 295318005, 295630005, 295731002, 295826009, 296499009, 296579000, 296686001, 296741007, 296782007, 296913000, 296947002, 297028008, 427590008, 715431002, 435491000124103, 1230081009, 7200002, 58327003, 59369008, 77277003, 77965002, 87810006, 185468001, 216472002, 238814003, 242825001, 290500008, 290538007, 290694000, 291055005, 291138008, 291508006, 291821006, 292102004, 292212004, 292512003, 292623009, 292669009, 293139008, 293251002, 293392008, 293406000, 293438005, 295478000, 296240007,

296474008, 296498001, 296607007, 296772006, 296890002, 297050009, 423215004, 762337006, 6051000124107, 453431000124100, 1162819002, 1383008, 55720000, 68563002, 95915001, 212516009, 242256000, 290562002, 290650006, 291211008, 291299006, 292425009, 292446000, 292534001, 292546002, 292798002, 293140005, 295226000, 295663003, 295708002, 295744007, 295770008, 295921000, 296088004, 296104001, 296244003, 296560003, 296615005, 296648005, 297020001, 297038003, 724701001, 762345001, 772133000, 86401000119104, 429211000124107, 10385009, 191857002, 191938005, 216479006, 216623002, 233672007, 239000007, 270430005, 290769009, 290840007, 291230008, 291305008, 291360001, 291363004, 291407004, 291879006, 291982000, 292692009, 292848005, 293221008, 295124009, 295392005, 295617009, 295645003, 295664009, 296064001, 296094007, 296357009, 296398002, 296679009, 296982001, 402747005, 21831000119109, 1003534004, 1145044008, 21413004, 21673003, 37344009, 38247002, 43695003, 191875001, 290989003, 291461001, 291638007, 291980008, 292036008, 292089009, 292186004, 292221003, 292243006, 292325005, 293497003, 293521009, 295286002, 295360009, 295491004, 295688006, 296197002, 296238002, 296582005, 296866003, 403668007, 425581000, 713542007, 715301007, 721898008, 860915001, 871674002, 54531000, 13504006, 109254000, 235051007, 240165007, 290139007, 290267007, 290352005, 290377006, 290574003, 290998000, 291024004, 291163008, 291346003, 291646008, 292145006, 292290008, 292510006, 292556003, 293054004, 293082006, 295183001, 295465000, 295485001, 295801000, 296192008, 296344001, 296419008, 296825003, 698622004, 698707001, 724688003, 369091000119106, 81119003, 1148544003, 46721000, 62942003, 191889006, 206128003, 216488002, 216584003, 233640009, 241939002, 290416007, 290452006, 290514008, 290525004, 290587009, 290855003, 290904006, 291479002, 291597007, 291698005, 291970009, 292264005, 292355003, 292956007, 292990003, 293037007, 295168006, 295502003, 295587007, 296545001, 296879007, 426364002, 708655006, 720676006, 762384006, 770709005, 788905002, 6348008, 14130006, 59651006, 61458006, 129576003, 236074001, 237665007, 290635002, 291037001, 291133004, 291324006, 292262009, 292460003, 293046001, 293579009, 295212009, 295246005, 295602005, 295924008, 296379004, 296509004, 296721002, 296727003, 297048001, 308692003, 762331007, 1149406003, 13857004, 35755007, 41136005, 61144001, 66351003, 210098006, 216532009, 218916005, 218953004, 231451006, 270425006, 290147007, 290236005, 290541003, 290927003, 291505009, 291804002, 292167007, 292234002, 292361000, 292388005, 293177002, 295125005, 295400008, 295476001, 296231008, 296462009, 297095002, 403611005, 403657007, 766054009, 871574008, 91471000119102, 1144767008, 1148569003, 1148734002, 11306002, 25277000, 41047001, 61558009, 242255001, 290346009, 290557003, 290595008, 290970003, 290991006, 291072003, 291436004, 291488006, 291894005, 292846009, 292854006, 293017008, 293240002, 295413002, 295510002, 295556005, 295748005, 295785002, 296378007, 296674004, 296850000, 297045003, 707524009, 715300008, 838522008, 429061000124102, 455411000124107, 455431000124101, 27795004, 1162805002, 34958009, 49540005, 235942001, 290417003, 290597000, 291089006, 291105004, 291332003, 291334002, 291600008, 291621008, 292115008, 292172003, 292232003, 292283003, 293138000, 293201007, 295289009, 295297002, 295339003, 295344005, 295383007, 296036006, 296444000, 296592002, 296843004, 408487007, 417906002, 442197003, 442351006, 443478002, 708122002, 720677002, 724665004, 724721000, 80868005, 111764002, 290387005, 290555006, 290678007, 290707009, 290737000, 290832001, 291039003, 291136007, 291181006, 291520003, 292432000, 292710008, 295873003, 296059000, 296131003, 296282006, 296694008, 297078005, 406448004, 10692681000119108, 52335006, 58700000, 73692007, 80092001, 89322006, 218704008, 218933003, 290233002, 290849008, 290964009, 291292002, 291468007, 291530007, 291631001, 291648009, 291928004, 292040004, 292803006, 293549002, 295178009, 295195001, 295589005, 295608009, 296316002, 296463004, 296544002, 296724005, 296921006, 297051008, 388977000, 403656003, 713728002, 1162825003, 1230078004, 13054007, 31956009, 58960006, 66409006, 68099003, 231469002, 290503005, 290520009, 290634003, 290945003, 291782006, 292406006, 292545003, 292549009, 292812008, 292900008, 293343002, 295213004, 295277007, 295357002, 295394006, 295462002, 296392001, 296559008, 296605004, 296953002, 296994005, 361272001, 419330004, 430973002, 699449003, 762562007, 838325009, 16076691000119100,

1148730006, 1149470008, 1156769000, 20876004, 37714008, 191802004, 191825003, 212725001, 242258004, 290421005, 291852007, 292054007, 292140001, 292177009, 292786007, 292843001, 293015000, 295173000, 295692004, 295929003, 296003005, 296390009, 296733007, 296826002, 302897004, 402258000, 403664009, 418572009, 427205009, 440611006, 720680001, 762516004, 770708002, 830281004, 298581000119103, 1145076003, 328381000119105, 8635005, 74264003, 89715001, 185348006, 191882002, 191906000, 212603001, 242829007, 290251001, 290269005, 290937008, 291148005, 291193002, 291232000, 291583004, 292190002, 292426005, 293068006, 293220009, 293373006, 293519004, 295302004, 295381009, 295549004, 296138009, 296206007, 296230009, 296746002, 296950004, 296990001, 724702008, 1144831005, 1163008006, 1179731007, 7511009, 29290000, 75683007, 79953006, 216620004, 275388009, 290791002, 291220004, 291221000, 291437008, 291474007, 292203001, 292301002, 292419005, 292477003, 293013007, 293055003, 293455009, 295463007, 295603000, 295798006, 296234000, 296285008, 296322006, 296546000, 296575006, 296608002, 296630001, 296735000, 296891003, 296942008, 308720009, 403665005, 428383000, 703848005, 724658007, 724661008, 306131000119100, 47237001, 1149412008, 30887000, 61406000, 64808005, 79982007, 86181006, 90526000, 231459008, 242835007, 290313003, 290430002, 290663000, 290753002, 290773007, 292156006, 292185000, 292816006, 293228002, 293308003, 293371008, 295288001, 295322000, 296222008, 296330007, 296345000, 296421003, 296429001, 296906000, 296931004, 403609001, 713698004, 15993751000119108, 1144835001, 34938008, 83362003, 191855005, 193609000, 200893007, 216539000, 216569007, 219015004, 230443000, 290162009, 291145008, 291256001, 291371000, 291448001, 291452001, 291625004, 292078002, 292208005, 292315000, 292506008, 292667006, 292828003, 292944006, 292947004, 293050008, 293550002, 293561005, 295158001, 295238009, 295305002, 295308000, 295854002, 296118008, 296271003, 296369001, 296370000, 296437009, 296596004, 296737008, 296846007, 296951000, 296981008, 297102008, 403653006, 789255009, 699687003, 191849000, 216616003, 239900005, 290244005, 290306003, 290393002, 290556007, 290760008, 290889002, 290908009, 291440008, 291519009, 291684002, 292503000, 292687007, 292906002, 293237002, 293348006, 293442008, 295762003, 295938001, 295959008, 296039004, 296359007, 403613008, 403652001, 407674008, 703845008, 711532000, 719410009, 720580006, 724675001, 773000004, 315951000119104, 293386002, 295427007, 52305004, 73383004, 83168008, 212701009, 231468005, 231963007, 238995004, 254250002, 290298009, 290378001, 290609003, 290621004, 290727005, 291017000, 291074002, 291110000, 291341008, 291667008, 291832006, 292205008, 292619001, 293161003, 293200008, 293408004, 295290000, 295642000, 295806005, 295852003, 295963001, 296530008, 296689008, 296938005, 297030005, 403648001, 427327003, 428720002, 440568002, 724679007, 455421000124104, 88031005, 422593004, 1145050003, 1149468004, 680007, 191918009, 212615002, 216541004, 216589008, 216733002, 218641001, 237449003, 290290002, 290809000, 291176002, 291586007, 291923008, 292272007, 292281001, 295223008, 295454005, 295952004, 296273000, 296400003, 296416001, 402260003, 723927004, 724654009, 403633007, 890232004, 24669002, 41390006, 64386003, 218326006, 218395001, 218606000, 290243004, 290408003, 290872005, 291130001, 291438003, 291469004, 291553008, 291807009, 292000005, 292158007, 292302009, 292521002, 292717006, 292767004, 293444009, 293536007, 293574004, 295375001, 295482003, 295955002, 296000008, 296213007, 296523002, 296703000, 296792004, 296794003, 297018004, 297026007, 297072006, 414689001, 306731000119104, 1149145003, 41602003, 75496000, 95918004, 127060006, 206570000, 216469009, 218929005, 230346007, 232032008, 290449003, 290622006, 290640005, 290860004, 290873000, 290906008, 290909001, 291217007, 291395003, 291403000, 291472006, 291509003, 291518001, 291984004, 292209002, 292360004, 292463001, 292592004, 293007003, 293229005, 293309006, 293332005, 295802007, 296081005, 296286009, 296449005, 296676002, 296831000, 402752000, 419253001, 698302009, 771073006, 1149140008, 1654001, 30078008, 33217001, 53556002, 95909001, 216498008, 268645007, 290847005, 291048008, 291555001, 291605003, 291623006, 291659000, 291962004, 292067005, 292144005, 292349008, 292381004, 292815005, 293275002, 293487007, 293545008, 295144003, 295155003, 295254007, 295473009, 295786001, 295820003, 295940006, 296226006, 296696005, 296766006, 296779002,

296915007, 296976004, 297052001, 297202001, 11196001, 48606007, 111782004, 218534001, 290783006, 290856002, 290958002, 290961001, 291006001, 291080005, 291155007, 291405007, 291413008, 292107005, 292182002, 292308008, 293044003, 293059009, 295472004, 295765001, 295866008, 295882009, 296381002, 296884001, 297089003, 396344000, 762508008, 870691007, 306161000119108, 1172690003, 1237136005, 22915003, 110281001, 206021007, 212616001, 290305004, 290483005, 291070006, 291083007, 291104000, 291534003, 291629005, 291674003, 291828000, 292320000, 292714004, 293211000, 293420007, 295671004, 295767009, 295781006, 295829002, 295914000, 295984006, 295997008, 296090003, 403623004, 441858005, 702809001, 723406000, 770702001, 153491000119103, 3053001, 129664008, 191856006, 216580007, 290716008, 290724003, 291169007, 291195009, 291301004, 291652009, 291680006, 292141002, 292492002, 292818007, 292877006, 293149006, 293167004, 295189002, 295636004, 295928006, 296396003, 296403001, 296704006, 296752001, 445902007, 724684001, 762390005, 1144828009, 47743003, 73575006, 87790002, 95239003, 191478006, 191492000, 191804003, 191826002, 290858001, 291015008, 291051001, 291809007, 292610002, 292779004, 293028007, 293066005, 295188005, 295426003, 295772000, 295878007, 296070007, 296925002, 296958006, 297259001, 439011002, 439990003, 724663006, 724676000, 725060001, 737338002, 24151000119106, 306651000119101, 28368009, 16077051000119102, 26679003, 42524008, 79815000, 111772000, 290518006, 290771009, 290894002, 290965005, 291161005, 291356004, 292171005, 292196008, 292730007, 292785006, 292793006, 292849002, 292883009, 293159007, 293243000, 293333000, 295291001, 295319002, 295619007, 295679002, 295860002, 295991009, 296426008, 296431005, 296536002, 296634005, 296784008, 698088001, 306541000119104, 461061000124108, 23833000, 1003473002, 1144838004, 59400006, 74524003, 185463005, 219063001, 290213001, 290353000, 290359001, 290804005, 291031000, 292050003, 292459008, 293398007, 295143009, 295217003, 295280008, 295643005, 295727008, 295846004, 296117003, 296176003, 296909007, 403621002, 699020004, 715430001, 15921731000119106, 1145546002, 39502009, 42894006, 212633008, 218614006, 290167003, 290461006, 291099001, 291204009, 292178004, 292244000, 292410009, 292433005, 292616008, 295201009, 295241000, 295422001, 295694003, 295715005, 295726004, 295756008, 295859007, 295974005, 296761001, 403637008, 428370001, 702702004, 762391009, 770711001, 133361000119101, 43124005, 1144778000, 12442008, 42377002, 44838007, 200892002, 216468001, 233741000, 290164005, 290470009, 290524000, 290913008, 291493009, 291574009, 292023008, 292049003, 292133002, 292468005, 292724007, 293027002, 293030009, 293069003, 293169001, 293230000, 295129004, 295165009, 295522004, 295871001, 295884005, 295961004, 296061009, 296150003, 296691000, 297001003, 699036002, 713244007, 722077007, 724519008, 725939009, 1148751009, 1156517008, 1231160001, 34764007, 69047008, 191871005, 218665003, 237641009, 290332009, 290484004, 290693006, 290854004, 291128003, 291668003, 291824003, 291870005, 291872002, 291977007, 292213009, 292269000, 292795004, 292821009, 292936006, 292937002, 293249001, 293276001, 293566000, 295376000, 295382002, 295812000, 296049001, 296108003, 296137004, 296168000, 296174000, 296200001, 296325008, 296401004, 296443006, 296504009, 296622002, 296624001, 296690004, 297075008, 361129004, 403301006, 403641007, 403666006, 698308008, 1148522008, 1156013003, 1837006, 70330008, 111756008, 212505002, 216533004, 216547000, 233945002, 238998002, 290190008, 290410001, 290414005, 290476003, 290522001, 292630003, 292958008, 292985001, 293142002, 293175005, 293505004, 293523007, 295799003, 295927001, 296035005, 296214001, 296290006, 296478006, 296483003, 296503003, 296639000, 296964004, 402394004, 715302000, 766045006, 105624005, 1186867000, 5444000, 67794001, 218608004, 218845008, 290431003, 290719001, 290746006, 291268005, 291458002, 291584005, 292363002, 292387000, 292606000, 292695006, 292860006, 293234009, 293542006, 295261006, 295301006, 295420009, 295732009, 295949007, 296566009, 296700002, 296885000, 296995006, 288461000119105, 337601000119109, 15911111000119105, 295429005, 1148534000, 9953008, 34738001, 47664006, 69482004, 82339009, 290284001, 290776004, 290938003, 290962008, 291121009, 291205005, 291402005, 291639004, 291643000, 293102007, 293193006, 295229007, 295581008, 295596007, 295651008, 296022001, 296098005, 296184004, 296445004, 296680007, 296898009, 297055004,

402756002, 430398007, 699072009, 708079007, 762329003, 298621000119103, 111769007, 191867007, 218568007, 218581000, 235875008, 241754000, 290311001, 290748007, 290827002, 291470003, 291486005, 291581002, 291830003, 292358001, 292421000, 292488009, 292515001, 292676004, 292891000, 293512008, 295186009, 295349000, 295363006, 295492006, 295986008, 296004004, 296165002, 296406009, 296613003, 296862001, 296912005, 724692005, 762335003, 16077091000119107, 5602001, 111755007, 231962002, 269268004, 290161002, 290165006, 290229009, 290237001, 291103006, 291455004, 291603005, 291869009, 291944008, 291994009, 292091001, 292251009, 292560000, 292939004, 293269006, 293335007, 293511001, 295520007, 295599000, 295937006, 296017001, 296074003, 296436000, 296991002, 297016000, 297054000, 401267002, 418944009, 445450000, 449845006, 118791000119106, 306601000119100, 890086003, 47307007, 71277006, 73097000, 190282003, 290710002, 291253009, 291342001, 291632008, 291681005, 291818009, 291976003, 292014005, 292810000, 293402003, 293500009, 293539000, 295483008, 295702001, 296275007, 296617002, 296671007, 699067000, 714829008, 724833000, 788543009, 2638001, 13216001, 50722006, 61803000, 216593002, 218814009, 290138004, 290202004, 290276000, 290361005, 290733001, 290848000, 290946002, 291216003, 291445003, 291576006, 291592001, 291645007, 291685001, 292063009, 292111004, 292314001, 293238007, 293334006, 293432006, 295202002, 295314007, 295913006, 295969002, 295999006, 296007006, 296087009, 296139001, 296171008, 296422005, 296580002, 296755004, 296962000, 361128007, 401271004, 724660009, 737335004, 762512002, 10853021000119107, 51725000, 421527008, 8481000, 18337009, 23877007, 26768000, 65435003, 87858002, 111779009, 275387004, 290406004, 290547004, 290601000, 290615003, 290667004, 290789005, 290893008, 290977000, 291120005, 291485009, 291544001, 291616005, 291895006, 291992008, 292227004, 292455002, 292537008, 292731006, 292814009, 295203007, 295626007, 295948004, 296024000, 296987007, 296996007, 297007004, 297199006, 419639006, 427180006, 698311009, 713529007, 22621000119103, 129141000119104, 428921000124100, 12398361000119105, 30564001, 292366005, 1144832003, 1148752002, 1162822000, 1162823005, 36261000, 42395004, 43893007, 81270000, 109776005, 111766000, 190448007, 203447001, 290171000, 290563007, 291525008, 291683008, 291861007, 292519007, 292808002, 292943000, 295152000, 295498005, 295625006, 296079008, 296672000, 296787001, 296948007, 403303009, 442760001, 709062008, 713693008, 725940006, 5947002, 60454001, 212898003, 216507007, 218475006, 231479000, 290375003, 290469008, 290529005, 291111001, 291159001, 291540005, 291696009, 291836009, 291851000, 291934006, 292114007, 293079001, 293350003, 293491002, 295135004, 295250003, 295525002, 295526001, 295843007, 296030000, 296195005, 296203004, 296347008, 297039006, 297068007, 297087001, 713910008, 447961000124105, 37370005, 44961008, 75544000, 111763008, 185318008, 212599001, 231470001, 237646004, 238815002, 290148002, 290351003, 290407008, 290696003, 290796007, 290892003, 291085000, 291093000, 291214000, 291985003, 292086002, 292259006, 292516000, 293459003, 295399001, 295513000, 296097000, 296124002, 296507002, 312936002, 361149008, 762320004, 770707007, 145101000119102, 292134008, 1144823000, 1269515004, 7072000, 29212009, 77650001, 129663002, 191495003, 212738009, 230317000, 231477003, 242975003, 290176005, 290191007, 290218005, 290239003, 290370008, 290411002, 290671001, 290874006, 290956003, 291153000, 292253007, 292819004, 292839000, 295180003, 295294009, 295353003, 295448009, 295698000, 295989001, 296044006, 296103007, 296260004, 296742000, 296935008, 403647006, 429672007, 762325009, 836439001, 57125001, 1144777005, 1148536003, 1156001008, 18869009, 22706002, 38968006, 191868002, 191936009, 212713004, 290158003, 290508001, 290763005, 290777008, 290815000, 291164002, 291651002, 291833001, 291849004, 292009006, 292051004, 292473004, 292994007, 293071003, 293347001, 295650009, 295681000, 295842002, 296091004, 296132005, 296366008, 296701003, 296832007, 296854009, 296881009, 297070003, 699018002, 827074008, 838525005, 15993671000119108, 403674007, 1148655008, 4526003, 55043004, 212785003, 216577006, 218961009, 237539004, 242060008, 290367009, 290618001, 290619009, 291069005, 291199003, 291236002, 291521004, 291609009, 291628002, 291789002, 291995005, 292129009, 292148008, 292189006, 292228009, 292348000, 292373000, 292769001, 293344008, 295154004, 295425004, 295461009, 295590001,

295649009, 296040002, 296067008, 296327000, 296382009, 296677006, 296894006, 296972002, 297056003, 419056004, 723882007, 788983007, 871576005, 306191000119101, 44969005, 1187103009, 70655008, 212503009, 216466002, 218620007, 218636009, 230333002, 290362003, 290642002, 290657009, 290699005, 290729008, 290736009, 291124001, 291354001, 291974000, 291981007, 292064003, 292226008, 292436002, 292528008, 292679006, 293528003, 295285003, 295516008, 295839008, 296554003, 296684003, 296754000, 296916008, 403631009, 408482001, 426790001, 724716003, 788904003, 288861000119108, 69434005, 70495004, 81459009, 87396006, 216525000, 241763003, 290285000, 290392007, 290686007, 290828007, 291481000, 291568009, 291960007, 292048006, 292157002, 292600006, 292832009, 293270007, 295185008, 295221005, 295594005, 296337005, 296440009, 296595000, 296712003, 296714002, 296911003, 296980009, 296984000, 297077000, 395215005, 420059000, 426095000, 441527004, 724689006, 737337007, 288031000119105, 876800006, 1179730008, 6900000, 11974006, 30984008, 34729003, 216486003, 218799005, 290168008, 290216009, 290409006, 290578000, 290586000, 290755009, 291140003, 291225009, 291326008, 291844009, 291886003, 291996006, 292527003, 292591006, 293006007, 293214008, 293529006, 293543001, 295377009, 295893006, 296075002, 296678001, 296785009, 296928000, 442108004, 713583005, 766221000, 4141000175103, 1156002001, 5319009, 7052005, 58400001, 71318009, 191827006, 191919001, 290253003, 290652003, 290660002, 290698002, 291367003, 291563000, 291617001, 291814006, 292899003, 295230002, 295237004, 295784003, 295898002, 295917007, 296095008, 296107008, 296196006, 296391008, 296471000, 415344002, 421946003, 427195008, 306211000119100, 1149240000, 276008, 9233004, 17592003, 32393008, 35437002, 63649001, 206022000, 216573005, 240102007, 280982009, 290371007, 290703008, 291087008, 291191000, 291238001, 291480004, 291501000, 291968000, 292180005, 292362007, 292499006, 292671009, 292771001, 292838008, 292912007, 293292006, 293437000, 293445005, 293562003, 295198004, 295560008, 296050001, 296291005, 296414003, 296876000, 296889006, 296920007, 698312002, 704126008, 306381000119102, 1144839007, 1145058005, 1155740004, 471791000124107, 1977004, 15167005, 36193003, 86321003, 191928000, 241757007, 290382004, 290464003, 290645000, 290679004, 290864008, 291114009, 291372007, 291580001, 291622001, 292197004, 292764006, 292852005, 293078009, 293372001, 293403008, 293517002, 295159009, 295199007, 295385000, 295697005, 295864006, 295981003, 296346004, 296351005, 296457008, 296725006, 724697004, 10854051000119101, 16077171000119107, 1149139006, 15919991000119104, 2530001, 7743006, 12843005, 32046005, 33871004, 38149009, 61356009, 190524001, 200852004, 277804000, 290302001, 291036005, 291200000, 291660005, 291784007, 291941000, 292125003, 292303004, 292367001, 292696007, 295474003, 295629000, 295669004, 295706003, 295714009, 295795009, 295988009, 296744004, 296975000, 403607004, 724720004, 838520000, 290425001, 54561008, 95912003, 111768004, 129666005, 216582004, 219050007, 234366009, 237884007, 290175009, 290564001, 290599002, 290654002, 290659007, 290778003, 290979002, 291223002, 291457007, 291499008, 291556000, 291578007, 292082000, 292378009, 292437006, 292723001, 292885002, 292986000, 293210004, 293423009, 293533004, 295153005, 295499002, 295533001, 295763008, 296008001, 296500000, 296654006, 296739006, 296781000, 296963005, 361273006, 439058005, 86062001, 288293001, 2819007, 11061003, 23527004, 241750009, 241947002, 290257002, 290568003, 290741001, 290917009, 290983002, 291142006, 291219005, 291634009, 291945009, 292046005, 292194006, 292220002, 292296002, 292444002, 292493007, 292689005, 295310003, 295729006, 295793002, 296183005, 296223003, 296293008, 296645008, 296758002, 296797005, 297003000, 419737003, 440385001, 724653003, 870674008, 1145075004, 47546008, 62276003, 212704001, 216605005, 290380007, 290784000, 291057002, 291064000, 291463003, 291626003, 291963009, 292056009, 292285005, 292364008, 292505007, 292951002, 293035004, 293257003, 293261009, 293354007, 293397002, 295265002, 295354009, 295574001, 295757004, 295956001, 296018006, 296328005, 296611001, 296954008, 296968001, 296971009, 297047006, 400072006, 403606008, 403675008, 403872008, 406593009, 737557007, 840449007, 871522005, 16228931000119102, 87904007, 292061006, 1155741000, 1230079007, 7071007, 25028003, 30857006, 32722006, 40857009, 77857001, 218952009, 235955000, 238901000, 290133008, 290234008, 290589007,

291270001, 291478005, 291922003, 292072001, 292911000, 293039005, 295122008, 295287006, 295329009, 295542008, 295571009, 295583006, 295701008, 295837005, 296010004, 296289002, 296851001, 296983006, 296989005, 724680005, 762323002, 770710000, 870194003, 21007002, 1149405004, 31430005, 74265002, 93432008, 95916000, 158058005, 191486006, 196052005, 238997007, 276581009, 280955001, 290565000, 290787007, 290922009, 290959005, 291116006, 291396002, 291517006, 291647004, 292169005, 292255000, 292291007, 292456001, 292894008, 292973009, 292996009, 293005006, 293052000, 293433001, 295593004, 295703006, 296218003, 296304008, 296353008, 296464005, 296827006, 296917004, 420650002, 439887005, 713450000, 736825009, 737226008, 762324008, 306531000119108, 403644004, 1148570002, 1148880006, 11268001, 49929007, 53523003, 73108003, 185346005, 290295007, 290614004, 291098009, 292105002, 292247007, 292617004, 292826004, 295272001, 295469006, 295494007, 295528000, 295741004, 295971002, 296057003, 296164003, 296324007, 296399005, 296564007, 296610000, 296799008, 296955009, 404647000, 699068005, 713691005, 788956006, 789046004, 817962007, 13889008, 1145077007, 1148567001, 2296009, 10028000, 61411003, 185321005, 191805002, 216464004, 235876009, 290275001, 290544006, 290932002, 291231007, 291426007, 291951004, 291952006, 292071008, 292306007, 292395001, 292400000, 292581000, 292965000, 293219003, 293530001, 295271008, 295330004, 295753000, 295857009, 295892001, 295968005, 296048009, 296411006, 296542003, 296663008, 297009001, 312960003, 414061000, 426950009, 724674002, 762321000, 762503004, 70885001, 216528003, 216590004, 230242007, 234341005, 235873001, 290447001, 290460007, 290641009, 290720007, 290891005, 291023005, 291092005, 291154006, 291298003, 291368008, 291589000, 291943002, 292317008, 292443008, 292532002, 292914008, 293024009, 293070002, 293080003, 293158004, 293174009, 293247004, 293253004, 293306004, 293457001, 293551003, 295518009, 295939009, 296233006, 296371001, 296510009, 296895007, 296965003, 297029000, 724687008, 762509000, 840592004, 212757000, 234426009, 290799000, 291020008, 291189008, 291227001, 291676001, 291873007, 292018008, 292963007, 292969006, 295146001, 295258005, 295418006, 296065000, 296470004, 296540006, 296973007, 300907002, 403636004, 403669004, 413851004, 722120008, 724719005, 772808000, 15993711000119107, 1153538009, 1231161002, 8732001, 86013001, 103297000, 191893000, 218408007, 218566006, 218792001, 290341004, 290496006, 290527007, 290558008, 290584002, 290844003, 291047003, 291061008, 291206006, 291424005, 291536001, 291946005, 292031003, 292257008, 292469002, 292979008, 295262004, 295506000, 295766000, 296144008, 296552004, 296934007, 296956005, 297002005, 297033007, 406434004, 419007000, 723933008, 723975009, 762515000, 789273007, 838519006, 1003842009, 1148654007, 1156734002, 1162821007, 89729000, 237450003, 241937000, 290271005, 290291003, 290366000, 290422003, 290638000, 290792009, 291056006, 291076000, 291313009, 291822004, 291837000, 292165004, 292277001, 292357006, 292593009, 292628000, 293300005, 293389009, 296086000, 296154007, 296202009, 296241006, 296278009, 297013008, 720679004, 767013007, 770890009, 306831000119106, 27992006, 30871003, 51443000, 53001008, 118951003, 206127008, 218795004, 290376002, 290523006, 290612000, 290824009, 291957000, 292490005, 292991004, 293255006, 295622009, 295752005, 295834003, 295895004, 295978008, 296181007, 296205006, 296229004, 296242004, 296456004, 296479003, 296688000, 296702005, 296765005, 296902003, 296961007, 420054005, 762322007, 838527002, 212718008, 16773005, 45775001, 52020004, 216556008, 216614000, 218613000, 290348005, 290478002, 290480008, 290723009, 291447006, 292025001, 292517009, 292901007, 293001002, 293285001, 293295008, 293331003, 293422004, 295235007, 295881002, 295908007, 296178002, 296387003, 296576007, 296650002, 296743005, 297043005, 403661001, 403667002, 440146002, 724529001, 724677009, 762513007, 1145056009, 1145057000, 1153542007, 16800004, 22888007, 39951001, 44003006, 47191007, 72527006, 185470005, 191148006, 216521009, 270424005, 290347000, 290403007, 290626009, 290997005, 291192007, 291318000, 291431009, 291931003, 292138006, 292396000, 292945007, 292964001, 293547000, 295299004, 295564004, 295673001, 295687001, 295695002, 296102002, 296529003, 296547009, 296723004, 296842009, 297071004, 418801006, 421530001, 724667007, 762671008, 866258001, 98811000119103, 1089991000119109, 1145547006, 1155772001, 29733004, 39807006, 50320000, 58025002,

82674000, 216586001, 290488001, 290495005, 290605009, 290842004, 291194008, 291284006, 291587003, 292163006, 292324009, 292536004, 293031008, 293520005, 293558009, 295367007, 295544009, 296211009, 296487002, 296569002, 296936009, 440168007, 10625951000119109, 16236661000119100, 1230080005, 43624006, 64921004, 218903005, 241746002, 241767002, 290552009, 291106003, 291658008, 291848007, 292037004, 292057000, 292121007, 292164000, 292279003, 292603008, 292738000, 292982003, 293034000, 293170000, 293171001, 293305000, 293394009, 293557004, 295209006, 295244008, 295337001, 295401007, 295627003, 296012007, 296506006, 296944009, 403630005, 713181003, 871577001, 306631000119107, 1162803009, 28504009, 48534006, 66590003, 67893003, 191806001, 212712009, 235022008, 254246009, 287165005, 290140009, 290582003, 290637005, 290974007, 291033002, 291160006, 291365006, 292080008, 292229001, 292250005, 292486008, 293341000, 293427005, 293554006, 293575003, 295404004, 295580009, 295644004, 295830007, 296085001, 296175004, 296771004, 296776009, 297062008, 297066006, 723936000, 762578009, 86391000119101, 290424002, 402748000, 453131000124105, 897590006, 1231159006, 191831000, 191869005, 216610009, 241959003, 254249002, 290274002, 290315005, 290692001, 290884007, 290929000, 291129006, 291228006, 291352002, 291867006, 291940004, 292076003, 292103009, 292113001, 292188003, 292485007, 292518004, 293166008, 293456005, 293515005, 295449001, 295488004, 295902008, 296199004, 296245002, 296522007, 296558000, 296638008, 296830004, 296967006, 309783001, 419476001, 699065008, 762340006, 871774005, 306621000119109, 293258008, 1148725007, 1153508003, 93433003, 218615007, 275693003, 290157008, 291004003, 291571001, 291575005, 291865003, 291866002, 292216001, 292484006, 292941003, 293452007, 293531002, 295312006, 295338006, 295457003, 295609001, 295616000, 296101009, 296317006, 296681006, 296734001, 296774007, 762336002, 37121000, 105625006, 25555003, 50933003, 55082002, 70273001, 109483004, 196757008, 290491001, 290668009, 290779006, 290819006, 290920001, 291399009, 291441007, 291956009, 291993003, 291998007, 292345002, 292817002, 292929005, 293029004, 293216005, 295632002, 295754006, 295869001, 295976007, 296364006, 296494004, 296514000, 296637003, 296835009, 297046002, 402744003, 439014005, 713726003, 725121001, 762505006, 838382005, 24000001, 40844001, 65263004, 74882009, 77721001, 84251009, 218619001, 231466009, 290222000, 290559000, 290852000, 291084001, 291471004, 291793008, 291840000, 291843003, 291975004, 292035007, 292359009, 292665003, 293265000, 293312009, 295487009, 295584000, 295585004, 295953009, 295965008, 296060005, 296128004, 296288005, 296453007, 297082007, 297097005, 425841004, 770705004, 432391000124104, 45472002, 4690009, 216502001, 216536007, 216558009, 218790009, 254252005, 278363000, 290250000, 290328003, 290919007, 291177006, 291636006, 291967005, 292007008, 292147003, 292430008, 295211002, 295231003, 295388003, 295395007, 295566002, 295575000, 295646002, 295813005, 295907002, 296412004, 296673005, 737336003, 298551000119105, 18768006, 66595008, 200794009, 216522002, 216661004, 239845005, 242823008, 290607001, 290811009, 291095007, 291537005, 291559007, 291573003, 291829008, 292618009, 292699000, 292908001, 292933003, 295181004, 295239001, 295379007, 295403005, 295414008, 295618004, 295954003, 296032008, 296052009, 296056007, 296368009, 296468008, 296594001, 296621009, 296803009, 297079002, 698609007, 1149408002, 20202008, 20881008, 33845003, 68629007, 69819005, 76400006, 93434009, 196053000, 216604009, 218379001, 268646008, 290312008, 290437004, 290537002, 290639008, 290658004, 290770005, 290899007, 291203003, 291317005, 291338004, 291373002, 291803008, 292004001, 292222005, 292305006, 292408007, 292609007, 292636009, 293203005, 295359004, 295557001, 295637008, 295855001, 295870000, 296548004, 296568005, 296571002, 296777000, 296798000, 312961004, 403635000, 720665007, 723928009, 838529004, 868184008, 1156515000, 2121004, 36706008, 82276009, 212727009, 216523007, 242833000, 290252008, 290344007, 290373005, 290455008, 290674009, 290682009, 291060009, 291307000, 291925001, 292918006, 292970007, 293225004, 293417004, 293424003, 295249003, 295336005, 295347003, 295433003, 295477005, 295993007, 296246001, 296355001, 296409002, 296718004, 296731009, 296905001, 310587004, 473188002, 762511009, 16000271000119108, 1145548001, 60634005, 216535006, 218586005, 218726009, 241758002, 241762008, 290445009, 290456009, 290926007, 291029009,

291147000, 291197001, 291359006, 291503002, 291901009, 291914005, 291929007, 292008003, 292175001, 292332001, 292910004, 293040007, 293245007, 295204001, 295707007, 295738008, 296265009, 296283001, 296907009, 427399008, 724530006, 840591006, 871629009, 367421000119109, 23842007, 30310000, 41839000, 47580006, 67888001, 68709002, 73007006, 73397007, 84758004, 90687000, 191476005, 191905001, 216480009, 216594008, 218680009, 290241002, 290342006, 290594007, 290673003, 290812002, 290914002, 291201001, 291398001, 291430005, 291506005, 291884000, 292624003, 293132004, 293163000, 293207006, 295161000, 295233000, 295257000, 295298007, 295447004, 295595006, 295709005, 295773005, 295985007, 296362005, 296451009, 296602001, 296603006, 296922004, 403612003, 724144006, 736024007, 789119007, 111767009, 290427009, 1142059006, 10650000, 13993001, 57628003, 61438005, 218950001, 276582002, 290208000, 290620003, 290706000, 291100009, 291689007, 292435003, 293212007, 293460008, 295508004, 296467003, 296619004, 296919001, 297024005, 297025006, 297064009, 408673004, 430136001, 698481003, 702811005, 1153374001, 1162806001, 70328006, 76346005, 216570008, 218705009, 290180000, 290201006, 290429007, 290675005, 290767006, 290863002, 291108002, 291168004, 291358003, 292466009, 292498003, 292864002, 293157009, 293507007, 295340001, 295553002, 295641007, 295736007, 295934004, 295936002, 296077005, 296620005, 296709001, 297092004, 135321000119107, 456201000124103, 47362003, 1144834002, 16959008, 63983005, 76547000, 83627000, 191832007, 290265004, 290434006, 290669001, 290738005, 290973001, 291264007, 291657003, 291662002, 292001009, 292231005, 292287002, 292411008, 292886001, 292898006, 295311004, 295316009, 295390002, 295817006, 295919005, 296628003, 406447009, 409564005, 870750003, 1148571003, 1231158003, 27331008, 39521001, 88320008, 109957002, 205788004, 216579009, 231942005, 232145009, 290467005, 290539004, 290617006, 290957007, 291038006, 291333008, 291594000, 291808004, 291926000, 292184001, 292440006, 292507004, 292542000, 292712000, 292961009, 293155001, 293311002, 295163002, 295192003, 295380005, 295598008, 295764002, 295848003, 296221001, 296395004, 296715001, 427229002, 724700000, 871525007, 871685000, 367631000119105, 12398651000119100, 1179217006, 4223005, 28711005, 48310002, 65679006, 80471007, 191819002, 212553003, 216622007, 218796003, 267206008, 290185005, 290226002, 290301008, 290438009, 290579008, 290664006, 291119004, 291126004, 291806000, 291890001, 291902002, 292044008, 292239007, 292245004, 292920009, 293183004, 293395005, 293573005, 295130009, 295563005, 295570005, 295849006, 296083008, 296236003, 296331006, 296455000, 296497006, 296577003, 296614009, 296706008, 296750009, 297012003, 403750002, 762334004, 306201000119103, 367411000119102, 1186983001, 5368009, 26714005, 34043003, 49141004, 57695009, 89291005, 89535003, 191838006, 191874002, 216709008, 218696007, 241753006, 290571006, 291030004, 291257005, 291294001, 291366007, 291640002, 291675002, 291897003, 292235001, 292415004, 292713005, 292953004, 293176006, 293192001, 293563008, 295451002, 295459000, 295975006, 296055006, 296109006, 296469000, 296618007, 296693002, 296708009, 711539009, 846605006, 298561000119107, 306221000119107, 196607008, 1258986006, 62014003, 218382006, 218698008, 237849008, 238503007, 290517001, 290631006, 291001006, 291127008, 291150002, 291314003, 292323003, 292386009, 293103002, 293156000, 293370009, 293537003, 295164008, 295169003, 295179001, 295182006, 295236008, 295391003, 295408001, 295750002, 295867004, 295926005, 296025004, 296252000, 296372008, 296424006, 296883007, 296888003, 390906007, 403671004, 410501000, 444271000, 708653004, 838528007, 453421000124103, 1148849001, 1155990006, 3902000, 24984009, 37331004, 95910006, 196051003, 216736005, 219031007, 230441003, 238992001, 290299001, 290750004, 290975008, 291016009, 291090002, 291380000, 291443005, 291477000, 291813000, 291882001, 292150000, 292237009, 292286006, 292427001, 292822002, 292824001, 293400006, 293407009, 293466002, 295504002, 295816002, 295865007, 296210005, 296756003, 296796001, 404694008, 722536004, 737340007, 762344002, 871681009, 306591000119107, 452081000124104, 1148651004, 1156746003, 59000001, 65242001, 66413004, 185464004, 218927007, 241948007, 290630007, 290713000, 291237006, 291401003, 291498000, 291988001, 292709003, 295410004, 295749002, 296354002, 296442001, 296952007, 313915006, 403670003, 724728006, 15911041000119105, 11862006, 32835006,

43302000, 191484009, 191845006, 200844002, 216600000, 216618002, 290261008, 290757001, 290805006, 290870002, 291059004, 291434001, 291460000, 291507001, 291686000, 292106001, 292289004, 292434004, 292452004, 292551008, 293451000, 293546009, 295227009, 295436006, 295546006, 295733004, 296146005, 296309003, 296532000, 296589001, 296643001, 296658009, 713186008, 1142110005, 32552001, 52367009, 81914009, 87363009, 216499000, 230304003, 290206001, 290242009, 290419000, 290718009, 290831008, 291514004, 291868001, 292329004, 292756000, 292907006, 292927007, 293009000, 293218006, 293236006, 293246008, 293504000, 293524001, 295295005, 295434009, 295658002, 295734005, 295925009, 295960003, 296072004, 296495003, 296588009, 402757006, 413837004, 428219007, 440696002, 306821000119108, 1149146002, 21836003, 26715006, 43497001, 216588000, 240101000, 242253008, 290603002, 290712005, 290752007, 290825005, 290940008, 291669006, 291805001, 291972001, 292200003, 292726009, 292794000, 293101000, 293278000, 293435008, 293513003, 295398009, 295402000, 295475002, 295704000, 296284007, 296301000, 296635006, 296640003, 296780004, 296869005, 403662008, 724698009, 724706006, 871683007, 890095006, 1153419002, 7538002, 14736009, 35228004, 40748005, 47753002, 49569001, 185345009, 185465003, 238988008, 290687003, 290740000, 290875007, 290928008, 291081009, 291215004, 291484008, 291495002, 291823009, 291856005, 292042007, 292136005, 292294004, 292523004, 292888000, 293490001, 295512005, 295552007, 295562000, 295607004, 295728003, 295823001, 295900000, 296143002, 296152006, 296272005, 296473002, 296531007, 296865004, 297080004, 720666008, 838386008, 846646001, 5851000119107, 292800009, 1144818000, 1149235008, 71387007, 191884001, 290647008, 290972006, 291082002, 291289001, 291810002, 291888002, 291964003, 292555004, 292587001, 292831002, 292835006, 293384004, 293401005, 293468001, 293498008, 295253001, 295647006, 295943008, 296063007, 296151004, 296215000, 296349006, 296361003, 296489004, 296711005, 296939002, 296946006, 297099008, 870367008, 871675001, 7761000119106, 1153540004, 15919911000119108, 18052008, 28864000, 53796002, 95917009, 110990003, 111558006, 212728004, 218943000, 290546008, 290709007, 291131002, 291516002, 291885004, 292263004, 292404009, 292417007, 292842006, 292844007, 293301009, 293396006, 295495008, 295631009, 295910009, 295911008, 403619007, 704206001, 725938001, 870677001, 1148859000, 1155738009, 236452000, 267527002, 290258007, 290577005, 290627000, 290862007, 291122002, 291132009, 291423004, 291548003, 291692006, 292055008, 292143004, 292501003, 292502005, 292806003, 293144001, 293154002, 293252009, 293303007, 295172005, 295653006, 295691006, 296080006, 296326009, 296410007, 296428009, 296515004, 296574005, 296762008, 296910002, 698314001, 704203009, 720678007, 866203004, 16031471000119107, 4863002, 46571000, 53702006, 68822001, 83150005, 191820008, 200075005, 218924000, 218928002, 236015007, 290200007, 290230004, 290308002, 290611007, 290793004, 290814001, 290823003, 290888005, 291040001, 291320002, 291427003, 291577002, 292335004, 293224000, 293488002, 293577006, 295150008, 295342009, 295355005, 295755007, 295809003, 295824007, 296105000, 296365007, 296439007, 297014002, 297090007, 441690002, 724672003, 737342004, 773298008, 6061000124109, 236522005, 1003535003, 1156000009, 1179725009, 14607001, 20117002, 109813002, 191931004, 212736008, 241849000, 290450003, 290596009, 290604008, 291152005, 291630000, 292273002, 293085008, 293129002, 293208001, 293508002, 293567009, 295396008, 295554008, 295600002, 295685009, 295689003, 296311007, 296335002, 296415002, 296565008, 296653000, 297000002, 408483006, 698480002, 724686004, 1148674000, 1187219008, 9489006, 13632006, 15649002, 20215004, 199547006, 206125000, 231480002, 231482005, 242830002, 290356008, 290420006, 290479005, 290490000, 290986005, 290992004, 291096008, 291115005, 291123007, 291171007, 291522006, 291531006, 291560002, 292003007, 292917001, 293014001, 295393000, 295439004, 295610006, 295863000, 295880001, 295990005, 296258001, 296556001, 296804003, 296896008, 307757001, 363101005, 403663003, 789272002, 871570004, 12815001, 39003006, 77355000, 191390009, 191865004, 212644000, 216463005, 218394002, 290432005, 290489009, 290519003, 291224008, 291504008, 291677005, 291797009, 291874001, 291990000, 292041000, 292461004, 292563003, 292719009, 292940002, 295151007, 295222003, 295386004, 295416005, 295550004, 295633007, 296308006, 724714000, 1144776001, 49450009, 269270008, 290179003,

290625008, 291619003, 291641003, 291694007, 291892009, 291942007, 291978002, 291989009, 292124004, 292298001, 292450007, 292543005, 292858009, 295232005, 295251004, 295279005, 295601003, 296177007, 296212002, 296427004, 310588009, 405544006, 438516005, 699071002, 762315004, 762387004, 838530009, 135311000119100, 1104801100011910З, 53923005, 66780009, 87132004, 191812006, 212617005, 216534005, 290297004, 290369007, 290728000, 290900002, 290902005, 291383003, 291482007, 291653004, 291655006, 291670007, 291678000, 291889005, 292274008, 292297006, 292641001, 293375004, 295190006, 295435005, 295586003, 295614002, 295639006, 295674007, 295875005, 295922007, 296020009, 296096009, 296136008, 296145009, 296287000, 296486006, 296665001, 296834008, 296949004, 408540003, 838521001, 840589003, 870191006, 1149147006, 1149228000, 1647007, 50026000, 56698004, 212636000, 212691007, 290717004, 290878009, 291178001, 291327004, 291331005, 291496001, 292128001, 292238004, 292242001, 292924000, 292946008, 292999002, 293509005, 295248006, 295348008, 295397004, 295471006, 295493001, 295638003, 295778001, 295909004, 296073009, 296148006, 296418000, 296562006, 296856006, 297041007, 299969005, 719407002, 723103002, 434761000124100, 4418008, 19307009, 110285005, 216501008, 218369003, 218370002, 218941003, 230316009, 290415006, 290588004, 290633009, 290702003, 291541009, 291699002, 292331008, 292841004, 293162005, 293288004, 293440000, 293506003, 295264003, 295346007, 295421008, 295515007, 295835002, 296084002, 296425007, 296461002, 296541005, 296555002, 297004006, 403616000, 403618004, 720682009, 12398281000119105, 1149410000, 1155997009, 108221006, 109774008, 191851001, 241949004, 290504004, 290785004, 291186001, 291279000, 291512000, 291539008, 291875000, 292919003, 295309008, 295453004, 295958000, 296227002, 296446003, 296927005, 419901001, 735616007, 31438003, 56294008, 216554006, 231472009, 290754008, 290978005, 291275006, 291903007, 292075004, 292371003, 292471002, 292845008, 292957003, 293076008, 295523009, 295628008, 295930008, 296058008, 296194009, 296356000, 296860009, 296941001, 403645003, 699011008, 702810006, 91411000119105, 453451000124107, 678411000119104, 295324004, 32093003, 75831003, 82768008, 89765005, 103079001, 191816009, 242832005, 290303006, 290349002, 290440004, 290836003, 290987001, 291820007, 291971008, 292159004, 292615007, 292896005, 293011009, 293418009, 293463005, 293484000, 295351001, 295411000, 295417001, 295868009, 296099002, 296142007, 296338000, 296695009, 403608009, 405542005, 423861007, 429692000, 713638002, 724683007, 724695007, 724724008, 762332000, 871673008, 70539005, 72820004, 1179385009, 6024006, 60199004, 77607006, 200068009, 236505008, 290394008, 290465002, 290869003, 290923004, 291151003, 291188000, 291526009, 291572008, 291860008, 292313007, 292319006, 292431007, 292700004, 292950001, 293148003, 295126006, 295139005, 295479008, 296393006, 296407000, 296999000, 361268000, 609437000, 609438005, 724694006, 737339005, 1148516004, 7916009, 19681004, 47836003, 76636002, 88674007, 231920009, 290374004, 290569006, 290575002, 292024002, 292509001, 293002009, 295138002, 295370006, 295406002, 295967000, 296134006, 296380001, 296505005, 296599006, 296845006, 297061001, 297094003, 297940005, 311173003, 403626007, 441636005, 735750005, 762420003, 315801000119108, 453701000124103, 20310007, 45110008, 89739006, 191887008, 212709006, 216621000, 231478008, 241748001, 290154001, 290343001, 290436008, 290666008, 290683004, 291079007, 291241005, 291340009, 291606002, 291799007, 291949003, 292059002, 292120008, 292268008, 292384007, 292403003, 292694005, 293198002, 295270009, 295519001, 296237007, 296352003, 361151007, 371627004, 403615001, 446468007, 830282006, 288295008, 1003530008, 1156735001, 1162820008, 54912002, 193548006, 242261003, 290197006, 290355007, 290444008, 290851007, 290954000, 291102001, 291248004, 291376005, 291494003, 291666004, 292199001, 292356002, 292439009, 292573001, 292642008, 292684000, 293256007, 295536009, 296015009, 296243009, 296402006, 296667009, 296852008, 296926001, 402753005, 403265002, 724717007, 870412008, 7248001, 18631001, 70340006, 111762003, 191480000, 191843004, 233637009, 290475004, 290722004, 290826006, 291045006, 291863005, 291959002, 292909009, 293283008, 293449004, 295176008, 295350000, 295656003, 295883004, 296135007, 296261000, 296476005, 296855005, 296899001, 296923009, 297037008, 426001001, 295430000, 403638003, 1149411001, 1179728006, 1231163004, 36265009,

36558000, 65986000, 72610007, 74902008, 191907009, 192811002, 212723008, 269688005, 270427003, 276628009, 290146003, 290390004, 290402002, 290550001, 290580006, 290790001, 291143001, 291927009, 291991001, 292866000, 292903005, 295438007, 295808006, 295844001, 296161006, 296454001, 296516003, 296551006, 296627008, 296945005, 297040008, 724673008, 306641000119103, 461071000124101, 1230077009, 184004, 11949008, 30138004, 66884007, 85761009, 191895007, 216601001, 218663005, 218869007, 290395009, 290548009, 290598005, 291475008, 291535002, 292379001, 292393008, 292451006, 292475006, 292479000, 293060004, 293241003, 293352006, 295157006, 295208003, 296247005, 296408005, 296482008, 296496002, 296629006, 403651008, 426873000, 295428002, 13503000, 65981005, 216545008, 218724007, 230760006, 236519008, 290207005, 290256006, 290458005, 290481007, 290853005, 290934001, 290941007, 290953006, 291046007, 291167009, 291449009, 291476009, 292598000, 292734003, 292966004, 293151005, 293263007, 293315006, 295140007, 295304003, 295521006, 295543003, 295565003, 295606008, 296217008, 296763003, 698403003, 871551009, 216518007, 1163367008, 4525004, 16248006, 41690001, 111345006, 200069001, 290182008, 290279007, 290435007, 290474000, 290896000, 291489003, 291532004, 291905000, 292109008, 292336003, 292342004, 292470001, 292650004, 293100004, 293282003, 293469009, 295256009, 295374002, 295529008, 295716006, 295780007, 296249008, 296300004, 296315003, 296363000, 403610006, 403659005, 724685000, 85337000, 890202007, 1148753007, 8995005, 12777009, 16332007, 50393008, 53275008, 190447002, 216562003, 234020005, 238519000, 290400005, 290442007, 290446005, 290935000, 290952001, 291487001, 291558004, 291561003, 292123005, 292276005, 292376008, 292402008, 292531009, 292897001, 293025005, 293271006, 293448007, 295293003, 295886007, 295947009, 296563001, 417229001, 703846009, 711420002, 720675005, 724707002, 762327001, 455461000124105, 16238221000119109, 12566000, 16260007, 85302003, 199252002, 201146008, 290399003, 290485003, 290498007, 290950009, 290981000, 291156008, 291374008, 291549006, 291695008, 292260001, 292627005, 292737005, 292997000, 293571007, 295378004, 295503008, 295511003, 295538005, 295796005, 295879004, 295906006, 296521000, 296537006, 296683009, 297058002, 428867008, 724670006, 725287006, 860699005, 368281000119108, 295327006, 520841000124109, 1848004, 28389000, 29602005, 32518004, 46975003, 52866005, 83465002, 203483005, 212607000, 216609004, 269265001, 290379009, 290756005, 291529002, 291663007, 292010001, 292101006, 292201004, 292215002, 292825000, 292847000, 292850002, 293404002, 295197009, 295206004, 295255008, 295273006, 295365004, 295572002, 295791000, 295983000, 295996004, 296033003, 296047004, 296120006, 296333009, 296340005, 296535003, 296878004, 296993004, 297042000, 297067002, 445507008, 448337001, 724659004, 724712001, 724843002, 762339009, 762462005, 762506007, 838551007, 97571000119109, 456311000124109, 666000, 1179724008, 4390004, 72972005, 191496002, 216467006, 216520005, 216599003, 290227006, 290685006, 291035009, 291134005, 291246000, 291381001, 292326006, 292834005, 293196003, 293222001, 293447002, 295175007, 295437002, 295945001, 296441008, 296669007, 296841002, 296897004, 296998008, 425339005, 439708006, 704205002, 724725009, 678401000119102, 1144824006, 1148731005, 10486009, 28191001, 32875003, 51510002, 55410006, 185469009, 190815001, 191916008, 216576002, 218764006, 290688008, 291026002, 291062001, 291412003, 291550006, 291691004, 291827005, 291924002, 292135009, 292230006, 292520001, 292553006, 292561001, 292761003, 293266004, 295307005, 295652001, 295657007, 295666006, 295862005, 295891008, 295962006, 296011000, 296235004, 296501001, 296518002, 296974001, 418370000, 698838008, 870414009, 367391000119102, 1149242008, 18051001, 20260003, 50529000, 57588009, 62711001, 111758009, 185389009, 216546009, 241761001, 290212006, 290646004, 290931009, 291183009, 291202008, 291213006, 291276007, 291565007, 291613002, 292005000, 292011002, 292070009, 292108000, 292489001, 292514002, 293097009, 293277005, 293485004, 295368002, 295611005, 295612003, 295890009, 296185003, 296539009, 296960008, 403643005, 439407002, 442654007, 715269009, 770704000, 782579006, 871776007, 38751004, 890087007, 15026003, 35528000, 40571009, 41083005, 51862005, 216583009, 216603003, 242824002, 254253000, 290160001, 290602007, 290879001, 290947006, 291028001, 291135006, 291288009, 291353007, 291551005, 291656007, 291842008, 291854008, 292058005,

292181009, 292191003, 292316004, 292458000, 292530005, 292837003, 295224002, 295343004, 295446008, 295836001, 295889000, 295916003, 296491007, 296593007, 296751008, 296859004, 297010006, 439591006, 10755041000119100, 25508008, 32267003, 51493001, 56277006, 88993007, 91267002, 191839003, 216555007, 218718001, 231467000, 242260002, 278023006, 290211004, 290314009, 290545007, 290697007, 290764004, 290772002, 290994003, 291003009, 291533009, 291798004, 292295003, 293000001, 293184005, 295269008, 295419003, 295489007, 295675008, 295745008, 296314004, 296397007, 296685002, 296838006, 403640008, 715058004, 724732000, 762504005, 1461000119109, 50325005, 111588002, 212700005, 218630003, 269272000, 290219002, 290383009, 290551002, 290590003, 290768001, 291175003, 291272009, 291800006, 291815007, 292248002, 292347005, 292392003, 292447009, 292981005, 293197007, 293274003, 293294007, 293336008, 293467006, 295341002, 295541001, 296155008, 296169008, 296274006, 296641004, 296900006, 407590002, 762333005, 1144822005, 8325007, 19445006, 61704003, 76941008, 185349003, 185467006, 199254001, 216485004, 216703009, 225929007, 241749009, 242834006, 268640002, 290159006, 290389008, 290453001, 290655001, 290749004, 290774001, 290808008, 290951008, 290976009, 291459005, 291794002, 291871009, 292162001, 292218000, 292344003, 292745000, 292801008, 293147008, 293434007, 293559001, 295276003, 295387008, 295444006, 295452009, 296082003, 296661005, 371089000, 386473003, 403649009, 419406002, 698321001, 715267006, 724681009, 724713006, 772999000, 871529001, 3391000175108, 1145047001, 5002000, 27064003, 111460005, 185347001, 191894006, 212602006, 216483006, 218791008, 231473004, 238996003, 291302006, 291339007, 291351009, 291557009, 291838005, 291845005, 292275009, 292720003, 293254005, 293461007, 295292008, 295445007, 295582001, 295941005, 295970001, 295982005, 296549007, 296557005, 296604000, 296659001, 296745003, 296929008, 309279000, 402745002, 762514001, 838376007, 10741871000119101, 199550009, 218868004, 218925004, 235750000, 235889003, 237844003, 290152002, 290533003, 290921002, 291454000, 292088001, 292322008, 292353005, 292412001, 293081004, 293205003, 295333002, 295442005, 295524003, 295758009, 295771007, 296204005, 296430006, 296534004, 296722009, 296829009, 296886004, 418603006, 723930006, 724710009, 12398321000119100, 1187205008, 77131002, 80300008, 110283003, 185316007, 212563006, 216624008, 290385002, 290585001, 290661003, 290835004, 290868006, 290898004, 290910006, 291329001, 292195007, 292407002, 292868004, 293489005, 293503006, 295148000, 295149008, 295779009, 296002000, 296512001, 296647000, 296853003, 297096001, 297100000, 300992002, 310202009, 711458003, 713692003, 866260004, 1081000119105, 455451000124108, 1148519006, 15244003, 19920003, 45536007, 52671001, 81648008, 91388009, 185466002, 216619005, 218610002, 218626001, 235648004, 269722001, 290254009, 290273008, 290277009, 290345008, 290542005, 290677002, 290700006, 290816004, 290857006, 290877004, 291007005, 291044005, 291297008, 291388007, 291582009, 292155005, 292270004, 292538003, 292666002, 292688002, 292796003, 292959000, 293062007, 293227007, 293250001, 295624005, 295648001, 295777006, 295933005, 296028002, 296110001, 296239005, 296434002, 296538001, 296887008, 418108003, 724699001, 871678004, 315791000119107, 1163009003, 1197766005, 20385005, 59826007, 79156004, 218416003, 218572006, 219019005, 235044003, 290360006, 290486002, 290613005, 290672008, 290963003, 291283000, 291406008, 291834007, 292441005, 292732004, 292763000, 292859001, 293023003, 293136001, 293262002, 295266001, 295456007, 295458008, 295532006, 295760006, 295920004, 296100005, 296113004, 296153001, 296323001, 296376006, 297088006, 698577000, 719848005, 724730008, 145841000119107, 367761000119105, 1187220002, 1231162009, 14784000, 15276008, 23146008, 38323006, 51160004, 95635002, 190416008, 191899001, 216475000, 216506003, 218481003, 219009004, 231481003, 254251003, 290221007, 290501007, 290735008, 292015006, 292026000, 292152008, 292198009, 292827008, 295331000, 295735006, 296041003, 312962006, 397696004, 427131001, 430400006, 724693000, 763184009, 788958007, 9408000, 1144826008, 90210000, 185317003, 216559001, 216572000, 268647004, 290454007, 290497002, 290534009, 290643007, 291041002, 291141004, 291222007, 291595004, 291795001, 291835008, 291906004, 292073006, 292971006, 293042004, 295187000, 295470007, 295527005, 295660000, 296336001, 296828001, 297074007, 403340001, 735493006, 870413003, 22719003,

53050002, 56918001, 58727001, 74934004, 196318002, 219036002, 241958006, 290268002, 290357004, 290368004, 290714006, 290948001, 292065002, 292110003, 292146007, 292339005, 292762005, 293020000, 293340004, 293465003, 293540003, 295210001, 295313001, 295604006, 295923002, 295972009, 296126000, 296232001, 296597008, 296652005, 296767002, 296768007, 297201008, 413183008, 713862009, 724718002, 766220004, 288021000119107, 237738005, 890088002, 1153539001, 136281000119109, 31752000, 64415008, 85975005, 290272003, 290912003, 291075001, 291277003, 291564006, 291633003, 292267003, 292968003, 293084007, 295191005, 295605007, 295677000, 295788000, 295894000, 295899005, 296198007, 296209000, 296253005, 296644007, 296716000, 296840001, 297021002, 297076009, 304605000, 388970003, 418541005, 446337009, 698309000, 717947009, 720673003, 762326005, 863953007, 871684001, 191471000, 1144830006, 1148542004, 25702006, 26543004, 212600003, 212708003, 218406006, 268797009, 275471001, 290163004, 290528002, 290553004, 290644001, 290743003, 290867001, 291310007, 291497005, 292241008, 292915009, 292972004, 293374000, 293462000, 295134000, 295640008, 295668007, 295705004, 295713003, 295957005, 296016005, 296977008, 297044004, 297049009, 425413006, 720674009, 723929001, 724669005, 789271009, 290593001, 1003532000, 1657008, 11429006, 14154001, 30491001, 56194001, 85618003, 290136000, 290566004, 291125000, 291300003, 291462008, 291500004, 291542002, 291642005, 291877008, 291883006, 292062004, 292397009, 292448004, 292480002, 292659003, 292962002, 292975002, 293041006, 295121001, 295281007, 295792007, 295904009, 296029005, 296092006, 296502008, 296561004, 296646009, 427897002, 724726005, 78731006, 7339009, 15382009, 32009006, 88165009, 111765001, 212664005, 235837008, 236521003, 290153007, 290177001, 290184009, 290245006, 290516005, 290629002, 291000007, 291792003, 291859003, 291893004, 292820005, 293393003, 295259002, 295432008, 295665005, 295797001, 296071006, 296147001, 296220000, 296609005, 296728008, 715268001, 871526008, 306371000119100, 306811000119101, 1078301000112109, 10755161000119102, 129665009, 880042006, 1149328002, 1155739001, 216564002, 235681003, 242262005, 290895001, 291235003, 291637002, 292278006, 293003004, 293016004, 293458006, 295218008, 295450001, 295782004, 296043000, 296475009, 296490008, 296572009, 296601008, 418505005, 427670006, 871922002, 292168002, 1156518003, 27956007, 49020008, 62388003, 95661003, 191322006, 191811004, 238146000, 269271007, 290186006, 290381006, 290521008, 290581005, 290833006, 291027006, 291077009, 291212001, 291315002, 291570000, 291904001, 292206009, 292389002, 292476007, 292693004, 292716002, 292809005, 295147005, 295373008, 295790004, 295822006, 295851005, 295973004, 296119000, 296121005, 296303002, 296305009, 296348003, 296360002, 296404007, 420094007, 703849002, 715303005, 762338001, 782593004, 15735401000119100, 470681000124105, 5578008, 17570002, 110282008, 191821007, 216491002, 236814005, 290189004, 290263006, 291654005, 292236000, 292690001, 292833004, 292942005, 295127002, 295245009, 295274000, 295588002, 295811007, 295828005, 295935003, 296127009, 296452002, 297015001, 297065005, 297091006, 403627003, 425974003, 703847000, 1145066001, 1531000, 45747003, 56916002, 63628002, 206124001, 212562001, 275482009, 290282002, 290405000, 291068002, 291343006, 291439006, 291473001, 291811003, 291966001, 292122000, 292369003, 292370002, 292413006, 293339001, 293548005, 293578001, 295177004, 295558006, 295711001, 295861003, 295944002, 296465006, 726543008, 788957002, 135301000119103, 1149241001, 13187008, 15301000, 25753007, 30274002, 39679005, 73935008, 77718003, 200070000, 218968003, 241764009, 290511000, 290576001, 290730003, 290866005, 291071005, 291174004, 291673009, 291787000, 291788005, 291826001, 292934009, 293131006, 293213002, 293494005, 295818001, 296373003, 296632009, 296717009, 296988002, 297019007, 301643003, 722329004, 846758008, 1162505005, 1230076000, 8502007, 14651005, 86225009, 216531002, 269689002, 290531001, 290721006, 290744009, 290761007, 290817008, 290865009, 291502007, 291801005, 292539006, 292677008, 292701000, 292740005, 293019006, 293209009, 293441001, 295838000, 295845000, 296023006, 296106004, 296350006, 296612008, 296760000, 699021000, 715270005, 104831000119109, 444971000124105, 235952002, 699022007, 1155999007, 2403008, 28563001, 40398009, 108220007, 190965006, 191900006, 191901005, 191930003, 212518005, 216481008, 216497003, 290220008, 290428004, 290535005, 290859009,

290942000, 291287004, 291304007, 292028004, 292394002, 292445001, 293501008, 295128007, 295282000, 295372003, 295443000, 295514006, 295740003, 296021008, 296166001, 296207003, 296263002, 296493005, 296578008, 296598003, 296730005, 296837001, 416520008, 426020004, 772020009, 72566003, 1144368001, 35189002, 36533005, 212742007, 212773006, 290286004, 290307007, 290916000, 291444004, 291671006, 291693001, 291933000, 292702007, 292902000, 292976001, 293268003, 293482001, 295548007, 296009009, 296076001, 296163009, 296870006, 296933001, 723926008, 724691003, 367201000119101, 1155736008, 32287002, 44206008, 66491001, 91074004, 216595009, 290358009, 290468000, 290502000, 290632004, 290715007, 290726001, 290800001, 290897009, 290985009, 291269002, 291271002, 291483002, 291910001, 291947001, 292173008, 292472009, 292491009, 292829006, 295228004, 295768004, 295804008, 295888008, 295998003, 296034009, 296543008, 296626004, 296759005, 296985004, 297034001, 713965007, 715971003, 870515008, 1144771006, 21445009, 32358001, 49463003, 206130001, 212739001, 216548005, 218973009, 235705002, 235830005, 241747006, 290172007, 290204003, 290457000, 290903000, 291303001, 291350005, 291446002, 291547008, 292225007, 292310005, 292744001, 292949001, 295559003, 295721009, 295751003, 295885006, 296251007, 296802004, 296871005, 297059005, 43599001, 63645007, 69399002, 72120001, 91433009, 191485005, 218621006, 218634007, 268796000, 290388000, 290413004, 291157004, 291242003, 291618006, 291961006, 293367005, 295142004, 295220006, 295710000, 295769007, 295903003, 295992002, 296114005, 296219006, 296276008, 296342002, 296417005, 296692007, 297027003, 390833005, 403628008, 699025009

##### 3.13 Fluid and Electrolyte Disorders

ICD10CM:
E87.22, E87.4, E87, E87.1, E87.71, E87.3, E86.1, E87.70, E87.7, E87.20, E87.5, E87.29, E87.79, E87.0, E87.6, E87.21, E87.8, E86.0, E87.2, E22.2, E86.9, E86

ICD9CM:
253.6, 276.50, 276.1, 276.61, 276.9, 276.5, 276.2, 276.6, 276.69, 276.0, 276.7, 276, 276.52, 276.51, 276.3, 276.4, 276.8

SNOMED CT:
47774001, 124201006, 124286004, 124435007, 236477004, 254256008, 276584001, 299465007, 426875007, 707755000, 712641002, 724351008, 124364002, 58618005, 119247004, 124361005, 124553004, 237682009, 237883001, 238104009, 303096003, 399144008, 402727002, 421437000, 722207000, 733457006, 763778003, 783157004, 783768006, 137941000119106, 435091000124105, 1085091000119108, 5667009, 722037004, 3642008, 21420006, 58136006, 237905007, 237993009, 239852007, 271847005, 361091007, 1611000119108, 1187542000, 2725003, 22910008, 40951006, 45369008, 47526003, 111559003, 124239003, 124444008, 124608006, 237840007, 276555005, 403828005, 722468005, 725393000, 733636005, 763312008, 763366000, 771469002, 783717008, 860858001, 133791000119107, 306181000119104, 81987005, 398664009, 1186825009, 5354002, 62151007, 63246000, 124168008, 124232007, 124451004, 236464008, 237640005, 238057007, 238066006, 360866000, 410059004, 722292000, 124698002, 1003862001, 1156858008, 1187534001, 23045005, 66931009, 83832001, 111397004, 124462004, 129456006, 236460004, 237959005, 267431006, 272938000, 734022008, 763280005, 771445001, 6479008, 8849004, 190330002, 428014009, 459063003, 609575003, 724279004, 726021008, 733086003, 143101000119101, 238000002, 1003938005, 41572006, 73211009, 111385000, 190242005, 237509009, 360653002, 609579009, 860860004, 124348005, 124367009, 124534009, 1155841005, 3427001, 11659006, 22062008, 58459009, 116020001, 124418008, 124663000, 206493009, 237913008, 399959003, 713890008, 723552005, 303891000119104, 368561000119102,

16006551000119104, 30652003, 1197747005, 2751001, 6669004, 50143004, 59399004, 111382002, 124280005, 124567008, 162685008, 237925006, 238020001, 423350005, 721088003, 722343009, 763721006, 773492007, 304331000119108, 72523005, 874900008, 1148934008, 1156853004, 1196872006, 13003007, 48839007, 49817004, 61451000, 237652003, 267454002, 302736004, 718211004, 770567006, 31321000119102, 306171000119102, 403834003, 1142071000, 1187113001, 28212002, 124131005, 124215008, 124460007, 190755005, 190794006, 237727005, 240168009, 402474007, 725031005, 367621000119107, 1186865008, 21529005, 35885006, 87074006, 124423008, 124559000, 124682009, 190389009, 190913009, 190920002, 192788009, 195025008, 237636001, 237806007, 238006008, 238068007, 238100000, 403397004, 733630004, 773666007, 306571000119106, 16005951000119104, 124660002, 237651005, 237955004, 735421004, 1208937004, 23272008, 39239006, 69718008, 124388000, 199228009, 237863002, 238115004, 314467007, 702377007, 717773005, 733599009, 816067005, 137421000119106, 124190008, 1153364004, 1162916008, 1264009006, 63275006, 78373000, 84121007, 124142001, 124312008, 190410002, 230352008, 237620003, 237911005, 237978005, 719216001, 720519003, 721840000, 310101000119107, 237643007, 1149073003, 27718001, 47757001, 61663001, 75934005, 77274005, 124324001, 124668009, 124707009, 186628001, 190694001, 190846002, 238022009, 238821003, 361090008, 609577006, 22933009, 1003387003, 25739007, 49973006, 62278002, 70199000, 75853001, 124476003, 124657009, 206492004, 237728000, 360579002, 421966007, 768553002, 125921000119106, 306561000119100, 25606004, 84727000, 1220589007, 1220596009, 90054000, 124517001, 124596009, 124607001, 190856003, 267433009, 609581006, 717263009, 20269002, 1187548001, 15771004, 18684002, 34818008, 53378008, 63656007, 127012008, 190884008, 237847005, 237965005, 402471004, 715633008, 722924000, 724096007, 735539005, 771513008, 55341000119107, 1155913007, 721201000124104, 12901004, 16966009, 34528009, 55296004, 56692003, 66789005, 70731005, 80908008, 111378004, 124283007, 237862007, 238874008, 239848007, 360771001, 699681002, 736652002, 306551000119102, 10743351000119102, 28140001000004102, 303094000, 1172839002, 1256073006, 1264112006, 21639008, 32612005, 44574006, 55178001, 64624009, 71785001, 124141008, 190643008, 237921002, 237988006, 396338004, 721172000, 722947004, 724784006, 725289009, 764962002, 783734000, 119181000119104, 124718009, 1592005, 6885006, 12602008, 35912001, 36891003, 70140000, 75682002, 81475007, 124148002, 124676006, 199226008, 237726001, 238019007, 238076009, 276557002, 403672006, 778065005, 1197052008, 1258977001, 1269423000, 359007, 20165001, 21764004, 30985009, 37548006, 72442006, 75598001, 124646004, 237983002, 238051008, 281611002, 717276003, 827115000, 1073961000119106, 14052004, 25998009, 124155000, 124274002, 1156856007, 12326000, 35759001, 124458005, 124489009, 124499004, 124535005, 190829000, 190866006, 267446004, 277365009, 138621000119108, 11179002, 57119000, 124622006, 1148936005, 1254946006, 1255121003, 26015003, 30967002, 37200009, 72262000, 76314005, 88380005, 111383007, 124146003, 124182008, 124310000, 124541003, 403806005, 405733001, 424925008, 709147009, 717046003, 723307008, 763320005, 771509001, 783176002, 870590002, 1264157002, 9345005, 22169002, 44176004, 49547008, 75387001, 91632005, 123967008, 124336007, 230426003, 238102008, 403738008, 403831006, 717044000, 310201000119103, 331481000119108, 16075931000119103, 124192000, 237914002, 1230003009, 4997005, 26254008, 59957008, 65791008, 84260001, 124654002, 276569005, 281362006, 360589003, 361010007, 361087002, 402476009, 717182006, 733623005, 771478008, 306151000119106, 10676831000119101, 1217065006, 76220009, 124152002, 124591004, 124661003, 124703008, 194909006, 237559000, 238157005, 276549000, 303087003, 303098002, 718603002, 733601006, 124621004, 1156857003, 1251488008, 5388008, 15978003, 35145002, 46885003, 61688009, 73475009, 84937002, 124180000, 124374004, 124455008, 237613005, 281610001, 609580007, 722454003, 724575009, 16040071000119106, 18104000, 75238000, 83470009, 124183003, 124350002, 124693006, 124696003, 1187550009, 4519003, 68581004, 80119007, 124687003, 404166001, 698729002, 719377004, 722937002, 763211004, 778067002, 120711000119108, 303971000119105, 11718301000119106, 440009, 7405009, 68724006, 124467005, 124616002, 128264007, 370992007, 399947002, 402788005, 413356003, 720864008,

763110007, 124701005, 1142075009, 1251499005, 29596007, 30529005, 85880000, 124299001, 124443002, 124626009, 237624007, 238161004, 303852004, 444173008, 733084000, 733115009, 733637001, 1149103000, 1187528008, 1197430005, 1256071008, 1682008, 13795004, 20756002, 24013007, 123965000, 124505003, 124598005, 360781002, 713401006, 68061000119109, 306581000119109, 1187040004, 41326006, 85020001, 124539004, 398958000, 609572000, 702327009, 703136005, 719454003, 724638006, 733600007, 765100000, 782772000, 11718401000119103, 16015911000119102, 26745009, 35633007, 1186847009, 1208936008, 1285518004, 11380006, 55236002, 124112004, 124128009, 124203009, 124589007, 180485001, 191172001, 199223000, 237697007, 237908009, 297226004, 733466005, 147211000119101, 16716281000119108, 51294009, 367241000119104, 10180006, 61071003, 124126008, 124496006, 124502000, 124640005, 237508001, 237807003, 237824009, 297288000, 445612002, 83201000119108, 367481000119108, 6483008, 123963007, 124277009, 124409006, 124642002, 190247004, 238027003, 238098000, 254221009, 267374005, 276645004, 428530009, 726081005, 765326001, 778068007, 307130006, 444944006, 1279844009, 10469003, 25646005, 29291001, 38063000, 54155004, 69478001, 80006005, 83858001, 124234008, 124441000, 124481007, 715338007, 718558008, 764733009, 783558004, 788758003, 306141000119109, 306611000119102, 124692001, 1217212009, 1237514002, 1264212004, 16851005, 24790002, 39929009, 66266003, 83563007, 86667008, 124463009, 124569006, 162742005, 206490007, 238045003, 297257004, 361066007, 83586000, 237994003, 1220625008, 16784003, 81685000, 85444005, 86353007, 124244005, 124407008, 190948002, 237638000, 237981000, 276568002, 360348000, 718880003, 1269235004, 8764008, 27943000, 70737009, 124223005, 124304000, 124362003, 124492008, 124650006, 360631004, 360812008, 716863007, 722302009, 775908005, 1153332006, 21952001, 23860007, 29512005, 45188008, 81896006, 124153007, 124396005, 124543000, 237853005, 361058003, 702440000, 722206009, 722488009, 105401000119101, 4887000, 26336006, 81986001, 83469008, 230677000, 267432004, 403813005, 403832004, 717962006, 21584002, 23569000, 24326000, 33316007, 89579000, 111556005, 124147007, 297235006, 773329005, 112421000119102, 1216941002, 9723006, 33116002, 39112005, 56661000, 61120003, 68985000, 71858003, 123970007, 124211004, 236446001, 237984008, 367406009, 472320005, 725078006, 773418007, 782886007, 830052009, 111377009, 317006, 25067009, 37934003, 45744005, 55822004, 75066002, 124177001, 237622006, 403807001, 718210003, 720940008, 721183009, 137931000119102, 367261000119100, 124691008, 1256075004, 1269231008, 50122000, 71946004, 124198001, 124345008, 124347000, 129590000, 237055002, 237957007, 286923006, 709110009, 720858001, 783057002, 368051000119109, 58112007, 7752002, 13644009, 50855007, 70610001, 75524006, 111387008, 124507006, 237611007, 237928008, 237989003, 360834005, 609463005, 783696009, 124110007, 124695004, 10399008, 66978005, 124165006, 235667000, 237919007, 238112001, 699260009, 721100009, 765146000, 1230272009, 9128006, 61172008, 62009002, 74185008, 95569006, 124305004, 124321009, 124475004, 192816007, 237617006, 237923004, 238048001, 254222002, 773576000, 784370005, 366961000119106, 34225008, 43929004, 45812003, 71638002, 71938000, 84193000, 124307007, 190737001, 238871000, 408539000, 421725003, 426898004, 443894002, 718713000, 721163008, 12246008, 1255323007, 50029007, 75610003, 79169000, 91273001, 124326004, 124669001, 124710002, 238078005, 238101001, 419503008, 709490002, 763204003, 65764006, 1137328006, 1172586007, 25404008, 59178007, 65524005, 111567006, 124672008, 237941007, 239000007, 773649005, 1571000119104, 40801000119106, 124166007, 1172841001, 15616006, 190680002, 199227004, 236467001, 237499004, 237966006, 238083002, 718212006, 801000124108, 298961000119102, 1073971000119100, 1075021000119104, 16040031000119108, 1187461004, 1237181009, 15877009, 42930003, 86444004, 124159006, 124290002, 190687004, 230358007, 237891005, 240165007, 277605001, 715374003, 724356003, 725033008, 733450008, 303881000119102, 310191000119101, 16076171000119109, 360721004, 1003367004, 4702003, 9386003, 51387008, 124313003, 124504004, 129591001, 201666003, 286909009, 360664004, 402478005, 426387005, 716721003, 303951000119101, 435101000124104, 10754881000119104, 15933901000119104, 124680001, 1173035001, 1279890001, 37295009, 42021008, 62144003, 62369005, 79488001, 123966004,

164208001, 237769009, 237842004, 297254006, 428839004, 720520009, 724769002, 725296006, 120731000119103, 360892007, 400128006, 4409006, 11225002, 45646000, 55783001, 81677009, 124113009, 124261008, 124392007, 124599002, 442009005, 782918002, 303911000119102, 1186844002, 3140001, 4477007, 24052000, 32891000, 33513003, 40751003, 67049004, 124511000, 190732007, 192782005, 341536001, 410050000, 720978005, 7265005, 53254000, 733452000, 1010397000, 1187616008, 26298008, 47632004, 124220008, 190682005, 708122002, 16076131000119106, 82319005, 1187529000, 1208620009, 14637005, 26399002, 26436007, 44673006, 111388003, 123810000, 124384003, 124573009, 124636001, 277894008, 297233004, 399087009, 403736007, 445120004, 146371000119104, 77365006, 1187511005, 1237272007, 28987007, 59761008, 124309005, 124429007, 234407001, 237603002, 237953006, 267424007, 276265003, 281612009, 360619001, 402725005, 450849003, 702366001, 304131000119106, 111396008, 1153361007, 1155910005, 15346004, 31368008, 78921008, 124376002, 190774002, 234176004, 371019009, 399126000, 716716001, 722457005, 789762004, 86531000119105, 18756002, 238069004, 12957008, 50967008, 124565000, 124606005, 237604008, 238033007, 765137006, 237642002, 1237418002, 3944006, 41300001, 59686008, 84618009, 124133008, 124295007, 297231002, 361086006, 703254001, 304301000119101, 1186878008, 1269236003, 7810004, 23800005, 43916004, 124470009, 124634003, 190894003, 230359004, 238037008, 405288003, 709556009, 715923003, 717774004, 724067006, 870313002, 306131000119100, 68067009, 880066000, 1251447008, 14725002, 44730006, 64081000, 84264005, 124308002, 190932003, 213281004, 236548002, 237902005, 238085009, 286920009, 302250008, 399149003, 405566004, 732246009, 304321000119105, 11718351000119105, 88393000, 1187525006, 124285000, 124398006, 237903000, 254223007, 360339005, 426896000, 82236004, 6183001, 46556004, 59531002, 124445009, 127339009, 238094003, 716747007, 721187005, 735538002, 5963005, 25211000119101, 15991002, 34530006, 53124003, 65389002, 66185005, 67528009, 124175009, 124658004, 190701006, 237852000, 237979002, 238023004, 239847002, 715632003, 718750004, 723675006, 310301000119105, 1269271003, 9537004, 55056006, 63329001, 65419005, 71751002, 82994006, 88512005, 95565000, 124684005, 237597000, 238038003, 361093005, 424099008, 717934004, 721284006, 723557004, 733072002, 337091000119107, 82003006, 238065005, 1003851001, 1153363005, 72945002, 124262001, 124330001, 124355007, 124430002, 236463002, 238158000, 782167001, 306731000119104, 39390005, 87827003, 190764000, 1003847003, 7085002, 24754009, 37703005, 237230004, 238007004, 276550000, 399053004, 764456001, 781386002, 827117008, 84969006, 1153334007, 1196873001, 1821000146108, 1388004, 6160004, 42484009, 49379003, 60118008, 68528007, 73068003, 124120002, 124323007, 124448006, 124516005, 124609003, 124667004, 198748007, 237598005, 237856002, 699331002, 778029000, 40791000119105, 124713000, 1237339005, 1260391004, 37666005, 42883007, 48655003, 124287008, 124450003, 124675005, 190331003, 190645001, 276426004, 721234004, 306161000119108, 44553005, 67202007, 1156791007, 1156792000, 1187523004, 22011005, 54736008, 124194004, 237920001, 278991002, 361001004, 734434007, 782745008, 16076011000119100, 238004006, 238026007, 54898003, 64654004, 124132003, 124520009, 124579008, 424846003, 722057000, 737212004, 783065004, 10753491000119101, 1196870003, 1216939003, 4107000, 11160000, 11687002, 81891001, 124243004, 124338008, 124456009, 124656000, 128289001, 234363001, 238009001, 313435000, 421075007, 445646001, 707756004, 735746003, 782757004, 306651000119101, 63844009, 1196948002, 1256066001, 16195001, 16279005, 23536000, 24308003, 33985005, 35208003, 46727001, 53836006, 73869005, 124144000, 124164005, 124181001, 124241002, 124296008, 236455003, 237696003, 363090004, 420422005, 709282004, 734173003, 773423007, 306541000119104, 24661004, 70348004, 28575006, 29692004, 47709007, 60268006, 124236005, 124281009, 124306003, 124416007, 235908005, 237934001, 297278001, 361075009, 718712005, 722851003, 880065001, 1264156006, 23156007, 42669007, 52139007, 58275005, 124139007, 124298009, 124359001, 190640006, 236461000, 237658004, 238028008, 360910004, 447292006, 709412006, 725026008, 836343001, 43253002, 89597008, 124714006, 9326001, 124595008, 236466005, 276629001, 297256008, 360586005, 724344004, 46635009, 88518009, 124366000, 124694000,

1187174002, 1187532002, 2243000, 21630007, 22886006, 62578003, 80710001, 91180009, 124149005, 124222000, 124248008, 124344007, 237641009, 237937008, 238021002, 238084008, 360943000, 361129004, 721236002, 308821000119109, 1142078006, 1187535000, 1822007, 80887004, 170766006, 238099008, 402472006, 429735007, 444707001, 709561006, 723579009, 298941000119101, 360361000, 1186849007, 22350004, 32284009, 37497004, 60414003, 78311009, 124335006, 126877002, 237619009, 276563006, 698854006, 699328003, 723624008, 724766009, 768554008, 18927009, 25443007, 124127004, 791000124107, 1187517009, 1256077007, 20052008, 40278002, 61336008, 236447005, 237729008, 405287008, 410058007, 719680009, 732264002, 774151000, 367041000119108, 239087008, 890171006, 1172627007, 1187514002, 44940001, 57886004, 124339000, 190745006, 190919008, 201670006, 267436001, 371117009, 389087006, 402787000, 403400008, 722928002, 732252005, 783615009, 3591006, 237985009, 1003850000, 1153375000, 1208738002, 15770003, 20155007, 29914000, 43258006, 46683007, 50056009, 62403005, 73281004, 123808002, 124154001, 124469008, 190786004, 238875009, 287266002, 303092001, 312857009, 717181004, 722054007, 725464001, 735386008, 735497007, 773575001, 782771007, 816156006, 306601000119100, 5335002, 1137357005, 1142077001, 1156317007, 1222644009, 27642008, 57507002, 124255005, 124575002, 238089003, 238952003, 402473001, 609566000, 367991000119101, 123763000, 124185005, 124478002, 307128009, 1074021000119103, 1172966001, 2638001, 14763005, 20987006, 36799008, 65327002, 124137009, 124157008, 192787004, 236532003, 237990007, 267426009, 274864009, 297253000, 397915002, 719974003, 722058005, 80963002, 124187002, 1156826003, 10033001, 33982008, 63103006, 124509009, 235915002, 237793004, 238063003, 239837000, 310505005, 732959007, 735422006, 784346006, 308901000119105, 17025000, 124612000, 1163454006, 1237417007, 14210003, 79886009, 90739004, 124284001, 124341004, 124715007, 190448007, 237501007, 237625008, 238087001, 276687002, 303097007, 307201006, 373607009, 21412009, 1237512003, 80394007, 124206001, 124302001, 206481000, 236468006, 239843003, 703526007, 724099000, 770924008, 774148007, 816155005, 124346009, 1220595008, 2186007, 15285008, 27373000, 44018007, 59655002, 124122005, 124269005, 237646004, 360697003, 733448000, 776416004, 124690009, 237644001, 238049009, 1208485009, 10741005, 15892005, 28710006, 54781007, 60743005, 77098009, 124191007, 124259004, 124557003, 124679004, 237599002, 238011005, 302661005, 733451007, 304261000119109, 1740001000004102, 16068491000119107, 1187524005, 15707004, 62999006, 124136000, 124219002, 124390004, 190429008, 419097006, 717812000, 720977000, 725028009, 763209008, 782739000, 309701000119101, 9105005, 37800003, 38032004, 40946000, 41797007, 67434000, 73856006, 124363008, 124473006, 190681003, 238103003, 238160003, 426705001, 860859009, 97361000119109, 306191000119101, 1092691000119109, 1186734006, 1187527003, 25425008, 36985004, 44054006, 123977005, 124438009, 124723009, 237556007, 240042004, 361203007, 416676005, 717788000, 770655004, 304291000119102, 124689000, 7573000, 52186006, 124613005, 201663006, 240307008, 387817006, 402786009, 724770001, 763325000, 763720007, 13901000119100, 84361000119102, 308791000119101, 1187526007, 1231309005, 1260128008, 5291005, 58263000, 95622006, 124597000, 124706000, 190304001, 237929000, 238163001, 398114001, 784347002, 55912009, 6400008, 8801005, 17885001, 56811006, 60952007, 124170004, 124497002, 237498007, 276627004, 360878001, 427195008, 704166007, 709111008, 723994004, 782755007, 830053004, 306211000119100, 1074001000119107, 1776003, 1220598005, 721121000124104, 17901006, 49562005, 49748000, 67148009, 70644008, 71690006, 95613005, 124254009, 124360006, 124461006, 124503005, 124665007, 190512008, 237771009, 237940008, 277893002, 360969006, 361043006, 370491005, 427627006, 719975002, 725079003, 306381000119102, 27503000, 31220004, 238002005, 1208747005, 1860003, 28259009, 30287008, 72744008, 89627008, 90500005, 124356008, 124471008, 124644001, 237623001, 360994007, 361088007, 410052008, 783174004, 15933941000119102, 897592003, 4598005, 7425008, 123811001, 124209008, 124529005, 124625008, 126533001, 237938003, 238025006, 238162006, 240167004, 271327008, 276262000, 360733009, 363140000, 719449007, 783722008, 309901000119103, 4920001, 124426000, 1003375005, 1196950005, 12045002, 40777006, 124600004, 233720006, 237647008, 237884007, 237890006, 237939006, 238003000,

360953004, 719816006, 303931000119107, 7259005, 20766005, 895227002, 1236807002, 21367009, 37814003, 62440002, 67312003, 254116003, 765329008, 1601000119105, 310121000119103, 238001003, 1251362006, 1255116001, 54470008, 57863006, 80473005, 82260000, 123813003, 124264000, 124314009, 124491001, 124581005, 238050009, 276634002, 403814004, 699869003, 708672004, 720860004, 724142005, 788756004, 138611000119101, 16010751000119107, 1230310007, 2241003, 42954008, 59455009, 123815005, 124436008, 124673003, 124699005, 237926007, 238035000, 238054000, 281365008, 361082008, 715830008, 719257008, 773726000, 124316006, 445275003, 1153362000, 1186809004, 1217044000, 35019008, 40873003, 77624000, 124256006, 217341008, 237637005, 363203000, 721838005, 771113001, 306531000119108, 15933861000119105, 70694009, 275598004, 1156181008, 987840791000119102, 43465001, 43498006, 45414006, 52600004, 67132008, 123812008, 124130006, 124266003, 124268002, 277895009, 735537007, 771339005, 789657008, 310061000119109, 58588007, 1187619001, 1222658006, 1231141008, 58256000, 58976002, 69614003, 83817008, 87049008, 124213001, 124315005, 190952002, 235906009, 237635002, 238030005, 360686003, 450316000, 718214007, 762291006, 310271000119108, 415764005, 1222662000, 1279891002, 9414007, 19604005, 63127008, 87694001, 124378001, 238061001, 276567007, 718193005, 771115008, 403830007, 1177118004, 28196006, 29633007, 52677002, 111307005, 124439001, 190368000, 479003, 1222655009, 1228849007, 8468007, 20313009, 39011001, 74479008, 124184009, 124394008, 124533003, 124686007, 238059005, 238953008, 275850001, 1237515001, 22774003, 28882002, 29028009, 39784009, 67854007, 124325000, 124592006, 237725002, 237922009, 277481003, 425079005, 609576002, 816082000, 306831000119106, 52165006, 124265004, 1153376004, 1187462006, 1172703004, 37702000, 118951003, 124279007, 190785000, 238041007, 410056006, 445260006, 309721000119105, 78642008, 237998000, 707742001, 46939000, 124118000, 124372000, 124380007, 124464003, 124633009, 276562001, 387814004, 1173034002, 10170007, 28193003, 30102006, 40488004, 61367005, 82342003, 106000008, 124605009, 186624004, 190407009, 276646003, 410055005, 721283000, 742876007, 310241000119101, 10743311000119103, 237602007, 1148694009, 1255271005, 88351001, 124340003, 124382004, 237886009, 297232009, 307127004, 360763000, 428896009, 700338003, 709109004, 717791000, 720830009, 766876004, 822995009, 304341000119104, 1208486005, 40527005, 78548001, 124472001, 124493003, 124521008, 124531001, 124648003, 203471008, 206491006, 237749004, 240096000, 771305006, 773276004, 788492001, 306631000119107, 308801000119100, 368521000119107, 124156004, 124525004, 124697007, 1187522009, 1217052002, 1254896008, 10877007, 16652001, 56051008, 124253003, 279081001, 303093006, 402726006, 429428003, 609573005, 718219002, 773415005, 773648002, 308811000119102, 86859003, 353295004, 56090007, 57414003, 60805002, 124288003, 124530000, 190915002, 237964009, 270369000, 360675000, 402785008, 444755001, 703527003, 724039002, 724227000, 782825008, 306621000119109, 124121003, 767133009, 1264565005, 42496002, 124343001, 237972006, 238040008, 277547006, 283839008, 315057000, 402475008, 700112007, 703137001, 303941000119103, 310181000119104, 37121000, 1186810009, 1284855000, 8900005, 75022004, 190902006, 201667007, 276644000, 421750000, 717942003, 720831008, 761000119102, 16076051000119104, 55711009, 890436003, 52767006, 63450009, 72925005, 190372001, 237843009, 238010006, 238081000, 239844009, 276560009, 431335002, 609490001, 764860006, 766251006, 310261000119102, 58833000, 1156855006, 1187640000, 1172843003, 1196869004, 4690009, 38495009, 55571001, 60216004, 124260009, 124674009, 124721006, 206489003, 237631006, 238079002, 286924000, 344587002, 408670001, 703138006, 720827002, 265569002, 360887005, 1153420008, 1172683008, 1197360001, 49013001, 54119007, 57557005, 76938004, 124332009, 124337003, 237855003, 238060000, 238093009, 239845005, 720941007, 122811000119101, 1075001000119108, 1260133007, 14783006, 30101004, 42012007, 124200007, 124510004, 234422006, 237854004, 709604005, 722059002, 701000119103, 237945003, 237954000, 879937000, 1230273004, 15689008, 26389007, 37183000, 46166004, 56233004, 74116004, 124135001, 124204003, 124351003, 124486002, 124619009, 206506005, 237907004, 268846006, 609567009, 768846004, 783064000, 784373007, 60634005, 78544004, 90093009, 281363001, 788757008,

870383008, 368851000119102, 82500001, 124270006, 28183005, 52724003, 123964001, 124301008, 124400005, 313436004, 360642008, 426655001, 699189004, 700111000, 720513002, 732951005, 734990008, 783767001, 310321000119101, 23447005, 57514000, 5181007, 82800008, 124677002, 237909001, 237950009, 238951005, 363204006, 424340000, 716200002, 303921000119109, 39925003, 73146005, 124349002, 237960000, 1222649004, 1217380005, 1310004, 22450000, 34349009, 35400008, 56041007, 56574000, 59229005, 80544005, 124297004, 124318007, 124547004, 190823004, 199229001, 235903001, 238014002, 238043005, 342553006, 439006004, 718721006, 106281000119103, 68295002, 1187536004, 1208621008, 1237349008, 7046009, 29736007, 111303009, 124143006, 124294006, 192812009, 398123003, 303961000119104, 368551000119104, 16006591000119109, 16015951000119101, 71322004, 124317002, 10518000, 43123004, 52452006, 78812008, 124601000, 237632004, 237975008, 403398009, 609563008, 726079008, 733112007, 771444002, 304121000119108, 310081000119100, 310221000119107, 237946002, 403829002, 722762005, 1179217006, 19726003, 27277001, 43339004, 60853003, 87235005, 116021002, 124250000, 124333004, 237841006, 398680004, 723367005, 306201000119103, 82245003, 5368009, 58558003, 70079009, 93466004, 124109002, 124303006, 124319004, 124334005, 124410001, 124442007, 360822002, 700107006, 711151004, 717185008, 306221000119107, 1222671009, 13920009, 39452003, 47417003, 62332007, 95612000, 237601000, 237628005, 237849008, 276261007, 399187006, 719430008, 776417008, 1197757006, 18829008, 59079001, 88740003, 124161002, 237860004, 238031009, 238090007, 238954002, 392559009, 402724009, 403399001, 445596006, 609562003, 609565001, 735732003, 306591000119107, 366951000119109, 238064009, 1172698005, 1197758001, 4575002, 11455007, 14140009, 66751000, 70528007, 190722000, 237506002, 420270002, 721085000, 764943000, 821000124103, 111231000119109, 5280003, 21535005, 64852002, 72452005, 72682008, 84816006, 190412005, 190882007, 237885008, 778025006, 41527003, 51002006, 31798004, 37695001, 50490005, 50869007, 51022005, 61165007, 61961002, 77817004, 124151009, 124158003, 124174008, 124214007, 124465002, 124638000, 236496000, 371067004, 424224001, 717821004, 306821000119108, 62201009, 422126006, 1236805005, 15059000, 23501004, 47641009, 73873008, 82141001, 124282002, 124422003, 124561009, 124670000, 237894002, 238047006, 238067002, 718106009, 718124006, 721235003, 726702005, 770755007, 783616005, 119831000119106, 309681000119104, 1153419002, 1177177000, 4170004, 16517004, 49513005, 74991009, 124257002, 124386001, 129589009, 360902000, 10672271000119100, 59289007, 124354006, 124480008, 360757009, 1186718008, 22830006, 26206000, 124115002, 124119008, 124169000, 124263006, 124289006, 124406004, 124498007, 124659007, 124708004, 206553008, 237910006, 238036004, 420200001, 423148005, 428173007, 15771000119109, 103931000119102, 58067006, 237952001, 1197753005, 57835009, 67855008, 81873006, 124629002, 237555006, 254220005, 402479002, 403805009, 421847006, 724146008, 732245008, 771516000, 783741006, 336941000119100, 25362006, 32442003, 57777000, 67845009, 111408001, 124212006, 124402002, 199225007, 212971003, 782744007, 303095004, 1153378003, 1187541007, 5655007, 8868001, 25010000, 26513001, 30390004, 60334003, 124171000, 124370008, 124453001, 124652003, 237895001, 240306004, 343570008, 367368009, 441690002, 609451000, 709414007, 724138007, 735909008, 763348005, 124179003, 1230303001, 1196949005, 25966003, 76751001, 123809005, 124162009, 124230004, 124610008, 206458002, 709075008, 721173005, 723583009, 368361000119108, 237996001, 4307007, 67455003, 124176005, 237750004, 238072006, 276263005, 276633008, 360410009, 387816002, 399170009, 609568004, 717792007, 724173009, 773331001, 28093001, 79385002, 360709003, 1264155005, 40607004, 47683004, 56595005, 190724004, 190855004, 214290000, 237621004, 238080004, 340519003, 360358001, 360932008, 399045007, 403732009, 609578001, 698953004, 723454008, 16006631000119109, 23490001000004109, 24529006, 56112001, 124202004, 124527007, 190644002, 237986005, 240095001, 297252005, 302870006, 360804000, 404163009, 445252005, 777998000, 819953000, 368961000119107, 124594007, 13866000, 15470004, 18789002, 51626007, 66095000, 69880002, 124549001, 124664006, 124671001, 237630007, 276558007, 408540003, 716362006, 717054001, 722285005, 724559006, 765403009, 773644000, 830054005, 309661000119108,

34852006, 23849003, 41013004, 47719001, 52760008, 124614004, 237977000, 297225000, 302866003, 359642000, 778022009, 339651000119100, 25422006, 63055005, 124528002, 7409003, 74450001, 76175005, 124615003, 190244006, 398796005, 763212006, 303901000119100, 86664001, 238062008, 1254947002, 43193009, 52311001, 61764000, 71702000, 111394006, 124365001, 281364007, 725587007, 774205007, 787412002, 80231000119105, 237995002, 47906008, 61031008, 72825009, 74162007, 83761007, 109477002, 123816006, 124454007, 124618001, 190953007, 237861000, 238159008, 414380008, 722927007, 84371000119108, 9311003, 48983004, 124611007, 237997005, 50658006, 51097006, 57218003, 90073001, 124291003, 124452006, 124587009, 128596003, 212970002, 237560005, 276586004, 699299001, 700109009, 737203001, 771442003, 62151000119109, 28428009, 69080001, 72831007, 124563007, 237858001, 238053006, 239026002, 717041008, 717961004, 101301000119106, 4783006, 48286001, 124700006, 238044004, 4996001, 11282001, 22684007, 124276000, 124358009, 124437004, 201669005, 238018004, 238872007, 446923008, 609570008, 716712004, 767134003, 1481000119100, 17827007, 444838008, 1222680009, 16242007, 59252009, 61164006, 61772003, 87898000, 124522001, 190773008, 190847006, 190860000, 236798003, 273700000, 715318006, 717787005, 725903003, 773643006, 256571000119105, 16914911000119107, 1179455003, 721111000124107, 124711003, 236541008, 238095002, 360854006, 402469004, 426255005, 719096006, 721973006, 304351000119102, 73123008, 124352005, 237512007, 6509007, 33559001, 237770005, 237999008, 238073001, 403827000, 445571008, 449902003, 712640001, 774154008, 124368004, 1156591005, 1279845005, 53783003, 124178006, 238017009, 238088006, 399249008, 721099001, 723452007, 306641000119103, 74653006, 88469006, 1217068008, 1237619001, 1255269005, 30174008, 124193005, 124226002, 254122007, 276564000, 297251003, 297255007, 361092000, 609569007, 710733002, 711152006, 723360007, 771448004, 773398005, 784349004, 368101000119109, 13144005, 238070003, 2538008, 32394002, 46894009, 59848001, 124221007, 124275001, 124519003, 190646000, 190842000, 234589002, 235909002, 267427000, 360373000, 402470003, 763688008, 782828005, 530558861000132104, 62311004, 129645002, 360416003, 361017005, 1153373007, 1251446004, 9360008, 29094004, 68451005, 124186006, 124501007, 129644003, 198709007, 236475007, 237507006, 238013008, 427784006, 721285007, 725286002, 763531001, 773668008, 783559007, 1220651005, 24338009, 51080000, 78590007, 124251001, 124293000, 124459002, 190447002, 237880003, 238092004, 360597005, 410051001, 721834007, 724768005, 1082511000119102, 1173036000, 1187171005, 9635004, 12770006, 24203005, 66521008, 68390005, 69525003, 75652008, 77402005, 124240001, 124617006, 237904006, 237933007, 238032002, 238097005, 66576001, 124189004, 1003860009, 1172605003, 1196871004, 7772007, 28032008, 45235005, 50375007, 124320005, 124604008, 199230006, 234405009, 276232006, 370493008, 371316005, 421784001, 711155008, 788491008, 298951000119104, 1172844009, 13196008, 43941006, 51984006, 64814003, 123807007, 237888005, 237980004, 238091006, 276559004, 276561008, 371628009, 717888008, 726080006, 783740007, 109041000119107, 114831000119107, 378007, 32123005, 1156813002, 1187512003, 12403008, 22935002, 23585005, 124246007, 237600004, 238074007, 238096001, 297238008, 360979008, 709413001, 717045004, 367391000119102, 360745000, 25765006, 33669002, 124208000, 197483008, 237659007, 238056003, 275404008, 276702002, 360921008, 361051009, 778048001, 124258007, 3744001, 35528000, 45116002, 54627004, 54905006, 61598006, 69329005, 124252008, 124432005, 124434006, 237931009, 271077003, 272064009, 449901005, 707551007, 310281000119106, 1038000, 5315003, 12002009, 22558005, 40802007, 89719007, 124421005, 124512007, 237554005, 237612000, 237645000, 387815003, 720976009, 25792000, 90505000, 1153331004, 1155842003, 1217050005, 1285519007, 16813005, 30913008, 34095006, 46785007, 59702004, 83850008, 124124006, 124583008, 235905008, 459062008, 698870008, 725044000, 445274004, 64235006, 86095007, 124237001, 124272003, 170765005, 236495001, 237639008, 237851007, 418470004, 304281000119100, 304311000119103, 71253000, 12313004, 20957000, 44759006, 59586001, 74003001, 111395007, 124224004, 190406000, 190859005, 236444003, 237627000, 237889002, 780820008, 783620009, 691471000119109, 1074031000119100, 124140009, 124357004, 1003849000, 10866001, 50327002, 54064006, 60737008, 124404001, 124536006,

190241003, 237844003, 405581005, 609574004, 65520001, 74320008, 124163004, 124427009, 1153351000, 124134002, 124245006, 124433000, 124440004, 190889003, 239846006, 297237003, 441963008, 722008003, 724576005, 725046003, 735423001, 7361000175106, 68393007, 403717002, 35691006, 43449002, 44231009, 73480000, 82793005, 105593004, 216325001, 710734008, 718751000, 782696001, 784381008, 1264154009, 5969009, 42393006, 52070001, 52270006, 52832001, 53210006, 82701004, 124242009, 124419000, 124538007, 237503005, 238822005, 269387007, 707441009, 13490001000004105, 16068331000119102, 1220574003, 13306007, 21263006, 26151008, 60045007, 61860000, 65277001, 124125007, 124532008, 124620003, 124709007, 128190004, 190416008, 206494003, 267434003, 445009001, 720859009, 765401006, 367681000119106, 48440001, 1197489003, 34566007, 83076007, 124238006, 124300009, 190869004, 360800009, 767139008, 783175003, 783178001, 79935000, 1187515001, 1187043002, 1222672002, 15890002, 38795005, 41345002, 58808003, 111569009, 124172007, 395204000, 413183008, 427089005, 609571007, 773426004, 36444000, 48638002, 64716005, 124311001, 124331002, 124524000, 190888006, 360792001, 448834003, 785303004, 55731008, 73843004, 20825002, 24595009, 34420000, 41142009, 50909009, 63702009, 72488000, 91352004, 124322002, 124342006, 124702003, 237505003, 237982007, 238024005, 288581000119102, 334051000119107, 9936001, 5579000, 8808004, 59451000, 124145004, 124411002, 124514008, 267447008, 360378009, 609561005, 609564002, 722212004, 725394006, 237961001, 1234819007, 30188007, 90616004, 95582003, 124412009, 124483005, 124508001, 237963003, 445010006, 733085004, 735475005, 763203009, 306371000119100, 306811000119101, 59990008, 124123000, 1148820009, 1187530005, 9979004, 73877009, 124267007, 124329006, 237633009, 238077000, 400170001, 700342000, 700463002, 717048002, 721225009, 722923006, 8933000, 124526003, 1145530005, 124138004, 124216009, 124585001, 236445002, 237755009, 237859009, 238955001, 278932006, 361089004, 398140007, 711409002, 720861000, 1148928001, 16060001, 41305006, 237951008, 361094004, 724040000, 107421000119101, 1217061002, 1237225007, 10899004, 58258004, 60193003, 78586005, 124278004, 124327008, 360959000, 363041004, 444756000, 725027004, 1003858007, 1156803005, 1187635008, 26132002, 33763006, 66002008, 124116001, 124273008, 267435002, 702365002, 153091000119109, 66937008, 124188007, 1179376009, 1187623009, 1186863001, 2359002, 6075009, 49227001, 54879000, 70134007, 72836002, 85487008, 124129001, 124678007, 237846001, 360546002, 426161002, 124688008, 890435004, 1142044000, 42183005, 69281008, 95837007, 124150005, 124228001, 124712005, 192817003, 199231005, 237944004, 237987001, 405256006, 426020004, 445261005, 711412004, 1082621000119108, 58610003, 1003368009, 1187250005, 3634007, 111552007, 124197006, 124628005, 234177008, 237848000, 360353005, 390951007, 398036000, 410053003, 423580001, 698290008, 709465004, 717047007, 734019006, 735496003, 69463008, 1197592001, 37982009, 44359008, 47922009, 81531005, 90560007, 111548007, 124111006, 124408003, 124466001, 238082007, 254069004, 360376008, 441656006, 733111000, 309741000119104, 10406007, 1148926002, 42927005, 44779003, 50279003, 62216007, 80902009, 124428004, 124593001, 190749000, 190787008, 238071004, 360369003, 699315005, 720951008, 733083006, 331331000119101, 368601000119102, 16005991000119109, 6974005, 12066005, 16754006, 43599001, 54954004, 55004003, 78960005, 111386004, 123814009, 124205002, 124577005, 124602007, 124704002, 190760009, 237879001, 238870004, 360987009, 403737003, 700449008, 762292004, 773406003

Truveta:
1215938

#### 3.14 HIV/AIDS

ICD10CM:
Z21, O98.712, O98.711, O98.73, B97.35, O98.719, O98.72, B20, O98.71, O98.713, O98.7

SNOMED CT:
414604009, 442537007, 713483007, 421529006, 230202002, 421020000, 420244003, 713275003, 733835007, 420549007, 421929001, 422282000, 421102007, 719522009, 420938005, 236406007, 713316008, 713967004, 422012004, 420384005, 421047005, 697904001, 421283008, 40780007, 240103002, 420281004, 713297001, 713446007, 713527009, 91947003, 420718004, 398329009, 713497004, 72631000119101, 397763006, 713487008, 1187001009, 713505000, 421660003, 235726002, 733834006, 713734009, 840498003, 1260096003, 713897006, 735527002, 713325002, 713341001, 713349004, 713490002, 713299003, 713340000, 713844000, 15928141000119107, 315019000, 713318009, 735528007, 445945000, 713511002, 10755671000119100, 713339002, 80191000119101, 420945005, 722557007, 735525005, 87117006, 713696000, 713845004, 735524009, 421508002, 76991000119109, 186707002, 713507008, 713570009, 713544008, 735523003, 1187000005, 420877009, 713488003, 713523008, 713881001, 421571007, 420614009, 186706006, 421415007, 713572001, 713445006, 420543008, 421077004, 421766003, 420687005, 713731001, 713260006, 422003001, 713729005, 713695001, 421454008, 442134007, 713533000, 713543002, 420452002, 771119002, 421708000, 422189002, 713546005, 771127006, 422136003, 713504001, 713887002, 422337001, 281388009, 713508003, 421272004, 48794007, 421312009, 840442003, 420308006, 81000119104, 86406008, 713733003, 713491003, 420544002, 713880000, 420818005, 421230000, 402901009, 421827003, 713489006, 421874007, 1153348007, 421597001, 421706001, 416729007, 420658009, 186708007, 713506004, 713722001, 90691000119105, 420321004, 111880001, 1142045004, 422177004, 421394009, 721166000, 765434008, 235009000, 713298006, 714464009, 422089004, 422194002, 91948008, 62479008, 90681000119107, 735521001, 421023003, 735526006, 421998001, 421851008, 713342008, 421403008, 713444005, 713545009, 714083007, 230180003, 735522008, 421710003, 421883002, 713964006, 420302007, 420721002, 420774007, 420787001, 52079000, 79019005, 713718006, 713732008, 420524008, 414376003, 421983003, 700053002, 421460008, 1142055000, 713510001, 95892003, 713571008, 76981000119106, 420900006, 697965002, 838377003, 713526000, 276665006, 420691000, 420554003, 713730000, 10746341000119109, 713320007, 422127002, 713531003, 713278001, 421671002, 405631006, 420801006, 421315006, 420395004, 713530002, 713532005, 420764009, 713503007, 230598008, 420403001, 421431004, 72621000119104, 713484001, 276666007, 713300006, 771126002

#### 3.15 Hypertension

ICD10CM:
I13.1, I12, I15.2, I11.0, I12.9, I12.0, I13.10, I13, I13.2, I13.11, I11, I10, I15, I15.9, I15.1, I15.0, I15.8, I11.9, I13.0

ICD9CM:
402.1, 404.11, 403.91, 402.9, 404.01, 403.90, 404.0, 404.92, 642.03, 403.01, 405.19, 642.0, 401.0, 403.9, 404.90, 405, 403.00, 405.99, 642.01, 404.12, 402.90, 404.1, 403.0, 404.13, 405.1, 403.11, 404.9, 401.1, 404.02, 403.10, 404.91, 404.00, 405.11, 642.00, 404.93, 642.04, 405.01, 402.10, 401.9, 402, 402.0, 402.91, 402.01, 404.03, 405.0, 403, 404, 405.09, 403.1, 402.11, 404.10, 642.02, 402.00, 405.9, 405.91

SNOMED CT:
22966008, 429457004, 54225002, 194791005, 129161000119100, 153851000119106, 111438007, 194788005, 285851000119102, 428163005, 1474004, 35303009, 367821000119106, 285001000119105, 9901000, 73410007, 198949009, 78808002, 285841000119104, 14973001, 56218007, 169465000, 96711000119105,

74451002, 704667004, 285101000119109, 26078007, 1208839002, 31881008, 766937004, 128001000119105, 123799005, 96751000119106, 284961000119106, 871642009, 285861000119100, 59720008, 66052004, 461301000124109, 153891000119101, 199006004, 39727004, 48146000, 111411000119103, 19769006, 36315003, 20753005, 129181000119109, 129151000119102, 66610008, 83105008, 449631000124102, 285911000119109, 285881000119109, 285081000119102, 1208845005, 285011000119108, 1201005, 15781000119107, 117681000119102, 285871000119106, 127991000119101, 285061000119106, 49102001, 762463000, 275516004, 712487000, 77970009, 123800009, 31992008, 108701000119102, 86234004, 57684003, 65518004, 427889009, 48552006, 10757401000119104, 23717007, 194783001, 10725009, 46481004, 118781000119108, 285831000119108, 18416000, 194767001, 96731000119100, 5148006, 284991000119104, 39018007, 284971000119100, 284981000119102, 63287004, 34273003, 120261000119101, 285041000119107, 40511000119107, 72022006, 194779001, 36221001, 64715009, 194785008, 199007008, 79355007, 286371000119107, 194780003, 96741000119109, 10759031000119106, 46113002, 199008003, 89242004, 60899001, 59733002, 8501000119104, 434431000124103, 104931000119100, 371125006, 96701000119107, 1078301000112109, 59621000, 285921000119102, 10757481000119107, 129171000119106, 78975002, 428575007, 96721000119103, 10757441000119102, 194781004, 28119000, 71874008

#### 3.16 Hypertension, Complicated

ICD10CM:
I13.1, I12, I15.2, I11.0, I12.9, I12.0, I13.10, I13, I13.2, I13.11, I11, I15, I15.9, I15.1, I15.0, I15.8, I11.9, I13.0

ICD9CM:
402.1, 404.11, 403.91, 402.9, 404.01, 403.90, 404.0, 404.92, 403.01, 405.19, 403.9, 404.90, 405, 403.00, 405.99, 404.12, 402.90, 404.1, 403.0, 404.13, 405.1, 403.11, 404.9, 404.02, 403.10, 404.91, 404.00, 405.11, 404.93, 405.01, 402.10, 402, 402.0, 402.91, 402.01, 404.03, 405.0, 403, 404, 405.09, 403.1, 402.11, 404.10, 402.00, 405.9, 405.91

SNOMED CT:
22966008, 54225002, 194791005, 129161000119100, 153851000119106, 111438007, 194788005, 285851000119102, 428163005, 1474004, 367821000119106, 285001000119105, 73410007, 198949009, 285841000119104, 14973001, 169465000, 96711000119105, 74451002, 704667004, 285101000119109, 26078007, 1208839002, 31881008, 766937004, 128001000119105, 123799005, 96751000119106, 284961000119106, 871642009, 285861000119100, 66052004, 153891000119101, 199006004, 39727004, 111411000119103, 36315003, 20753005, 129181000119109, 129151000119102, 66610008, 83105008, 449631000124102, 285911000119109, 285881000119109, 285081000119102, 1208845005, 285011000119108, 15781000119107, 117681000119102, 285871000119106, 127991000119101, 285061000119106, 49102001, 275516004, 712487000, 77970009, 123800009, 31992008, 108701000119102, 86234004, 57684003, 427889009, 48552006, 10757401000119104, 194783001, 118781000119108, 285831000119108, 194767001, 96731000119100, 5148006, 284991000119104, 39018007, 284971000119100, 284981000119102, 34273003, 120261000119101, 285041000119107, 194779001, 36221001, 64715009, 194785008, 199007008, 79355007, 286371000119107, 194780003, 96741000119109, 10759031000119106, 46113002, 199008003, 89242004, 60899001, 59733002, 8501000119104, 434431000124103, 104931000119100, 96701000119107, 285921000119102, 10757481000119107, 129171000119106, 428575007, 96721000119103, 10757441000119102, 194781004, 28119000

#### 3.17 Hypertension, Uncomplicated

ICD10CM:
I10

ICD9CM:
642.03, 642.0, 401.0, 642.01, 401.1, 642.00, 642.04, 401.9, 642.02

SNOMED CT:
429457004, 35303009, 9901000, 73410007, 78808002, 56218007, 74451002, 59720008, 461301000124109, 48146000, 19769006, 1201005, 762463000, 65518004, 23717007, 10725009, 46481004, 18416000, 63287004, 40511000119107, 72022006, 194785008, 371125006, 1078301000112109, 59621000, 78975002, 71874008

#### 3.18 Hyperthyroidism

ICD10CM:
K74.02, K76.5, K70.40, K74.00, K71.4, K73.8, K74.01, K70.41, K73.9, K74.0, K70.31, K71.11, K74.1, K70.9, K76.7, K74.4, K71.51, K72.10, K71.7, K71.3, K76.3, K70.0, K74.69, K70.2, K76.6, K74.3, K70.30, K74.5, K71.50, K74.60, K73.1, K71.10, K72.11, K71.0, K73.0, K76.2, K74.2, K76.1, K73.2

ICD9CM:
070.32, 571.9, 070.9, 456.21, 070.6, 571.4, 571, 573.1, 573.4, 571.5, 572.2, 571.42, 571.41, 571.6, 571.49, 573.3, 070.23, 570, 070.44, 571.0, 572.8, 571.2, 571.1, 571.8, 572.4, 573.0, 573.8, 573.5, 070.33, 573.9, 573.2, 070.54, 571.3, 572.1, 572.0, 070.22, 456.2, 456.0, 456.1, 456.20, 571.40, 572.3, V42.7

SNOMED CT:
1761006, 79720007, 235917005, 186639003, 1092801000119102, 1155841005, 235898000, 37688005, 703866000, 1186865008, 1220580006, 771149000, 235896001, 1155913007, 713370005, 43904005, 123717006, 38662009, 73475009, 768289009, 16859701000119109, 235900003, 197310003, 235886005, 197301006, 235880004, 19943007, 235869004, 721710005, 15999000, 723829000, 34736002, 197299004, 419728003, 197291001, 266468003, 435101000124104, 16070004, 12368000, 724278007, 31712002, 10295004, 76301009, 271440004, 721847002, 197303009, 1186854003, 123605001, 197305002, 128302006, 1234822009, 6183001, 715864007, 197284004, 715401008, 1197150002, 768006009, 328383001, 235899008, 725939009, 88518009, 76783007, 717187000, 716203000, 735451005, 863957008, 724766009, 123607009, 1197739005, 61977001, 725940006, 1230342001, 281388009, 41889008, 870517000, 235901004, 1197736003, 50167007, 737202006, 89789003, 420054005, 725416005, 774204006, 713181003, 768125005, 79607001, 197442005, 57339008, 123606000, 72925005, 123716002, 197293003, 60037002, 699189004, 424340000, 190823004, 871619002, 10690671000119109, 197279005, 197441003, 314963000, 74669004, 197300007, 266471006, 725938001, 831000119103, 235902006, 16098491000119109, 702969000, 9843006, 1208726006, 307757001, 27156006, 768288001, 838380002, 235895002, 713966008, 838305005, 58282009, 109819003, 735733008, 21861000, 33144001, 74162007, 78208005, 197294009, 768127002, 1010616001, 708198006, 1237346001, 240792005, 432908002, 371139006, 235897005, 266470007, 62484002, 197286002, 767809001, 838377003, 1197704005, 235889003, 31155007, 767810006, 86454000, 266469006, 425413006, 123604002, 30188007, 45256007, 536002, 66870002, 722870008, 768126006, 186698009, 153091000119109, 43634002,

197296006, 1116000, 450880008, 713965007, 103611000119102, 89580002, 773737004

##### 3.19 Lymphoma

ICD10CM:
C82.5, C84.1, C84.53, C82.33, C84.13, C81.01, C84.11, C82.94, C84.96, C85.18, C85.71, C84.33, C82.41, C84.23, C81.96, C82.62, C83.1, C84.0, C84.44, C83.43, C84.79, C84.20, C83.88, C85.96, C84.49, C84.Z7, C85.88, C81.75, C84.Z, C85.22, C83.55, C83.60, C84.95, C81.38, C83.01, C83.36, C85.8, C81.3, C83.64, C84.A3, C83.12, C85.21, C84.4, C84.A1, C84.A6, C83.91, C86, C81.2, C82.21, C82.6, C84.05, C82.59, C84.39, C81, C82.84, C85.00, C83.96, C84.47, C84.63, C82.60, C84.94, C81.70, C82.87, C86.5, C83.82, C83.97, C84.69, C84.73, C85.72, C84.68, C85.20, C83.71, C86.3, C82.53, C82.75, C82.42, C81.37, C81.49, C82.52, C82.30, C82.36, C84.67, C84.34, C84.28, C85.04, C85.08, C86.4, C81.10, C83.37, C84.Z1, C81.11, C81.34, C82.4, C84.A4, C81.12, C84.41, C85.27, C84.9, C85.06, C81.32, C82.72, C82.74, C82.93, C84.21, C85.90, C82, C83.03, C83.72, C84.Z9, C81.05, C82.55, C83.53, C84.A0, C85.92, C85.83, C81.36, C83.73, C82.32, C84.65, C85.85, C81.31, C83.92, C81.47, C83.30, C83.58, C85.99, C82.11, C82.04, C81.23, C84.Z0, C83.05, C83.3, C83.98, C82.28, C84.66, C85.76, C83.0, C83.59, C83.21, C83.61, C82.64, C83.50, C82.24, C83.31, C83.49, C84.32, C82.19, C82.26, C82.57, C84.7, C82.09, C85.81, C82.45, C81.15, C83.13, C84.60, C84.76, C83.68, C84.75, C82.43, C84.30, C81.19, C81.39, C81.45, C81.97, C83.07, C83.83, C85.1, C81.71, C83.19, C83.44, C84.93, C85.82, C82.1, C82.66, C82.92, C83.28, C84.27, C85.02, C85.16, C81.77, C84.77, C81.00, C82.03, C84.51, C84.56, C85.15, C81.46, C84.35, C85.2, C83.00, C81.22, C83.20, C84.03, C84.Z2, C84.18, C82.67, C82.48, C83.22, C85.93, C84.54, C84.57, C81.95, C84.06, C84.16, C84.A, C82.80, C82.31, C83.45, C81.94, C82.17, C82.29, C83.42, C83.46, C84.99, C81.16, C82.05, C86.2, C88.4, C81.44, C84.40, C85.12, C83.34, C82.3, C82.37, C82.83, C83.27, C83.86, C84.02, C81.02, C81.25, C84.31, C86.1, C82.65, C83.7, C84.15, C81.0, C81.4, C81.48, C82.91, C84.37, C84.09, C82.14, C81.08, C82.50, C83.16, C83.52, C83.85, C81.30, C84.Z6, C83.57, C84.04, C84.26, C82.56, C82.78, C83.87, C84.45, C84.91, C81.74, C84.64, C85.14, C83.41, C83.89, C83.70, C81.29, C82.13, C82.96, C84.A8, Z85.71, C84.7A, C81.91, C84.6, C84.98, C84.Z8, C81.06, C82.02, C84.24, C85.10, C85.78, C81.17, C83.02, C83.32, C85.29, C85.79, C84.70, C84.A5, C81.42, C82.51, C84.46, C85.11, C85.74, C81.03, C82.49, C82.54, C82.71, C83.94, C84.Z4, C82.47, C82.61, C82.77, C84.Z3, C81.40, C85.97, C86.6, C81.27, C82.06, C84.12, C82.23, C83.24, C85.13, C81.93, C86.0, C84.22, C82.39, C85, C85.89, C81.92, C84.72, C84.97, C83.51, C84.50, C84.62, C84.42, C85.05, C83.38, C81.35, C82.12, C82.76, C84.Z5, C85.07, C81.20, C83.18, C83.48, C85.94, C82.79, C83.67, C82.10, C83.23, C81.33, C82.15, C82.8, C82.98, C83.39, C83.69, C84.55, C85.19, C85.25, C82.08, C83.84, C84.29, C82.58, C85.80, C82.16, C82.18, C82.9, C83.54, C85.95, C84.52, C84.A2, C82.97, C82.40, C84.A7, C83.79, C84.07, C84.14, C85.01, C82.89, C83.80, C85.17, C81.79, C83.62, C83.63, C84.25, C82.27, C83.81, C81.21, C82.35, C82.44, C81.41, C83.99, C85.70, C85.86, C82.20, C82.86, C83.26, C82.22, C81.14, C81.76, C83.11, C83.78, C83.5, C83.77, C84.90, C85.84, C82.70, C85.73, C83.14, C82.0, C83.10, C83.9, C85.09, C83.15, C83.25, C81.13, C81.43, C83.90, C84.74, C84.78, C85.87, C84.48, C85.77, C82.95, C83, C83.76, C82.46, C85.23, C85.75, C82.00, C85.24, C85.26, C81.26, C82.2, C83.09, C83.40, C81.24, C81.28, C82.90, C83.8, C81.9, C83.95, C84.36, C84.71, C81.78, C82.99, C83.17, C83.75, C84.38, C82.88, C84.58, C83.33, C81.90, C81.18, C85.03, C81.98, C82.38, C83.56, C83.93, C84.00, C83.04, C85.28, C81.73, C83.29, C84, C84.10, C84.59, C81.09, C82.25, C85.91, C82.68, C84.08, C84.19, C82.01, C84.92, C85.98, C82.63, C82.73, C83.06, C83.08, C83.74, C81.99, C84.17, C84.43, C83.66, C85.9, C81.04, C81.72, C82.34, C82.81, C83.65, C82.69, C84.A9, C81.1, C81.7, C82.07, C82.85, C81.07, C82.82, C83.35, C83.47, C84.01, C84.61

ICD9CM:
201.40, 201.54, 202.63, 201.95, 200.22, 200.46, 200.25, 200.75, 202.48, 200.43, 200.57, 202.50, 200.14, 200.33, 201.12, 200.82, 200.62, 202, 203.01, 201.22, 200.77, 201.60, 202.71, 200.4, 200.84, 202.64, 200.12, 201.67, 202.12, 200.2, 201.55, 202.46, 200.7, 202.17, 202.10, 200.70, 201.64, 202.97, 202.85, 200.73, 200.80, 202.74, 200.21, 238.6, 200.56, 202.5, 200, 201.18, 200.68, 202.3, 201.62, 200.61, 202.23, 202.41, 200.24, 201.4, 201.94, 202.06, 201.76, 202.82, 201.0, 200.67, 200.76, 202.1, 201.90, 202.11, 202.07, 201.92, 200.54, 202.38, 200.00, 200.17, 202.42, 201.28, 201.42, 200.83, 202.03, 201.2, 202.18, 200.52, 201.41, 202.53, 201.96, 201.10, 202.56, 201, 200.55, 201.75, 202.84, 203.00, 202.33, 202.28, 202.45, 201.01, 201.08, 202.54, 200.85, 200.13, 202.31, 202.80, 200.32, 202.20, 202.34, 202.4, 202.98, 201.66, 202.32, 201.26, 200.02, 200.05, 200.18, 201.73, 201.78, 202.01, 202.35, 202.65, 200.26, 201.51, 201.47, 202.7, 201.70, 202.47, 203.0, 202.44, 202.75, 201.65, 201.72, 201.97, 200.20, 200.38, 201.00, 202.67, 202.73, 201.53, 202.04, 200.86, 201.1, 201.57, 202.78, 200.44, 202.62, 201.15, 202.91, 200.36, 201.13, 200.42, 202.36, 201.5, 200.88, 201.48, 201.58, 201.25, 201.45, 200.34, 202.24, 200.04, 202.15, 202.93, 202.88, 201.24, 201.68, 202.43, 202.9, 202.94, 200.10, 201.50, 200.6, 202.00, 201.56, 202.83, 200.47, 202.55, 202.87, 202.58, 200.45, 201.93, 202.95, 201.77, 202.92, 200.35, 201.04, 200.15, 202.57, 202.30, 202.68, 202.8, 202.90, 200.51, 202.02, 201.43, 201.74, 200.30, 200.06, 201.02, 200.78, 200.0, 202.26, 200.64, 202.22, 201.11, 201.61, 200.01, 200.66, 201.20, 201.06, 201.52, 201.9, 200.40, 200.8, 202.13, 203.02, 201.44, 201.46, 201.7, 202.6, 202.60, 200.1, 202.21, 200.23, 202.81, 202.2, 200.08, 201.63, 201.98, 200.03, 200.74, 201.23, 201.27, 201.21, 202.37, 200.72, 202.0, 202.16, 200.37, 202.05, 202.14, 202.27, 201.17, 202.51, 200.5, 200.63, 202.61, 200.65, 202.25, 200.07, 200.11, 200.48, 200.58, 201.16, 202.40, 200.60, 201.07, 201.03, 201.05, 201.6, 202.08, 200.41, 202.70, 202.66, 200.71, 201.91, 202.96, 202.72, 200.28, 200.3, 200.53, 202.52, 202.76, 200.81, 200.16, 200.31, 200.50, 201.71, 202.77, 201.14, 200.27, 202.86, 200.87

SNOMED CT:
448372003, 93516008, 414780005, 1255898004, 93542008, 426336007, 449219006, 277609007, 1255808008, 93195001, 188502002, 277617004, 404141000, 448317000, 448865007, 93543003, 442537007, 448666006, 830057003, 109962001, 1255791002, 188674006, 302848008, 713483007, 307633009, 1153347002, 188507008, 133751000119102, 93534007, 39086001, 189984004, 400122007, 739301006, 1255789005, 127220001, 763477007, 277626001, 188541000, 1255798008, 188570009, 404135007, 276811008, 371134001, 93541001, 109970006, 303055001, 314926009, 128808004, 188506004, 404117003, 765136002, 1255796007, 188635004, 188568000, 188591001, 109966003, 95226002, 188631008, 840424008, 188575004, 188634000, 441313008, 447100004, 448269008, 1255799000, 19340000, 404113004, 448867004, 188503007, 448354009, 118607005, 116711000119103, 421283008, 449065000, 68261000119102, 277610002, 95186006, 93496009, 1172592001, 128921005, 734141009, 188529007, 1255743003, 93191005, 448371005, 12351000132102, 404157000, 1163404000, 404137004, 117211000119105, 188512009, 188672005, 420028002, 117091000119108, 277611003, 95187002, 254792006, 277627005, 118605002, 373168002, 448774004, 115245001, 188492005, 404118008, 188513004, 1196900003, 188589009, 762690000, 74654000, 898197001, 94690004, 404127009, 733860007, 77381001, 303017006, 419386004, 188562004, 307635002, 713897006, 93494007, 93547002, 445269007, 314923001, 404121005, 420788006, 40411000, 302841002, 449217008, 449318001, 713325002, 93522004, 1255750004, 404144008, 448220006, 93514006, 93523009, 110007008, 190818004, 404149003, 447656001, 1153354008, 55150002, 94688000, 128806000, 449292003, 93555009, 1237578005, 95193005, 95264000, 128802003, 188718006, 1172695008, 763884007, 109967007, 414166008, 1255687003, 76481008, 94686001, 94711005, 188637007, 188590000, 143291000119104, 445406001, 448465000, 448555009, 352791000119108, 448319002, 419392005, 450958009, 1157162007, 1255890006, 415110002, 109979007,

1255892003, 313427003, 404128004, 1255812002, 724645006, 840423002, 188587006, 763666008, 118617000, 1187136005, 1255726001, 722954005, 93489007, 1187135009, 448738008, 188572001, 307625008, 92511007, 46744002, 95209008, 314924007, 450911001, 702785000, 68291000119109, 93196000, 703626001, 188537004, 1156403002, 1255666008, 189985003, 188516007, 112687003, 188609000, 116811000119106, 116871000119103, 188500005, 93527005, 94687005, 118611004, 307624007, 414553000, 448217003, 93193008, 103685007, 448376000, 449418000, 783541009, 188565002, 1163043007, 93192003, 447989004, 713516007, 1255727005, 1255745005, 303056000, 419879004, 276836002, 419586003, 93495008, 93509007, 188579005, 188585003, 128801005, 397352006, 1255802009, 307647008, 93518009, 103686008, 277637000, 448447004, 93493001, 93533001, 189982000, 314929002, 449053004, 189962004, 277643003, 404138009, 420519005, 726721002, 188576003, 425657001, 448387008, 441559006, 117061000119101, 277545003, 448254007, 733895005, 93505001, 93536009, 836277009, 109965004, 95260009, 447806008, 448386004, 116741000119104, 118599009, 128803008, 445105005, 449220000, 188517003, 1677531000119107, 1255811009, 448384001, 721555001, 786960000, 93510002, 1255786003, 94704006, 448709005, 1255801002, 94708009, 128807009, 277632006, 836276000, 93497000, 93520007, 188515006, 109989006, 404108003, 443487006, 1255785004, 188577007, 1255806007, 404120006, 714463003, 71109004, 118601006, 1148851002, 1255753002, 93194002, 55020008, 1187129009, 115246000, 188498009, 277625002, 426071002, 783220004, 93552007, 109972003, 45572000, 277618009, 418789003, 456911000124108, 118600007, 402881008, 64575004, 188501009, 399648005, 351211000119104, 109964000, 95194004, 302842009, 404111002, 449173006, 116841000119105, 1163005009, 277613000, 445030005, 188569008, 188593003, 733627006, 404115006, 440422002, 722953004, 724647003, 1222550009, 93492006, 188531003, 1148845007, 1255754008, 188547001, 1187107005, 397467006, 735332000, 1255888005, 188613007, 188627002, 716788007, 93521006, 1217301006, 95230004, 449075002, 93525002, 404143002, 302845006, 314922006, 404119000, 68271000119108, 188511002, 1255788002, 402882001, 404147001, 422853008, 449063007, 461351000124108, 50102004, 188487008, 277579002, 449177007, 449221001, 449419008, 450910000, 109968002, 1153348007, 1255792009, 314925008, 118608000, 236513009, 188679001, 1255887000, 52248008, 449074003, 404140004, 448468003, 277615007, 1255804005, 94709001, 307636001, 404136008, 448560008, 4950009, 188566001, 95188007, 397469009, 109969005, 188538009, 109978004, 93531004, 188632001, 404131003, 734067001, 117111000119100, 93500006, 188554007, 419770008, 93515007, 1255784000, 277614006, 1255895001, 277616008, 686981000119108, 786909001, 93487009, 1255905007, 836486002, 188505000, 441962003, 449307001, 188586002, 43985008, 314927000, 765328000, 93532006, 93550004, 1255790001, 122981000119101, 404114005, 724648008, 118602004, 277622004, 762691001, 1255800001, 128800006, 1153357001, 1255803004, 13048006, 397350003, 418628003, 277623009, 404109006, 444910004, 95263006, 307637005, 414785000, 448553002, 448609001, 721762007, 277473004, 773995001, 307646004, 449072004, 93498005, 1255907004, 94715001, 188493000, 445925008, 1148847004, 452291000124109, 93506000, 93507009, 1155957006, 1187190002, 702786004, 128805001, 110459008, 93519001, 449216004, 93530003, 420063007, 1153336009, 1255906008, 95224004, 188633006, 277580004, 314930007, 1155956002, 103682005, 404133000, 404145009, 415112005, 448561007, 1255751000, 404150003, 109971005, 1162264008, 109975001, 188675007, 276815004, 116691000119101, 314931006, 421246008, 784551004, 1162973007, 448231003, 449176003, 715950008, 93524003, 1153346006, 404130002, 699657009, 277612005, 413537009, 1255749004, 400001003, 188580008, 420302007, 404129007, 448607004, 93548007, 118610003, 1255793004, 447658000, 447766003, 128875000, 1197344000, 713718006, 77430005, 128798004, 188551004, 67824002, 95231000, 240531002, 188612002, 94710006, 402880009, 404110001, 449222008, 188524002, 308121000, 277629008, 58961005, 448672006, 866098005, 116821000119104, 1153383006, 1255728000, 188504001, 277651000, 109977009, 255102004, 397349003, 787565006, 352411000119109, 461311000124107, 70600005, 93197009, 93198004, 109976000, 426885008, 16893006, 188558005, 118618005, 404116007, 93199007, 188489006, 448212009, 736322001, 95225003, 277567002, 450908002, 188514005, 419094004,

1255270006, 93546006, 93554008, 188536008, 307634003, 95261008, 447805007, 93526001, 188582000, 188548006, 353211000119103, 1255883001, 413587002, 448213004, 448663003, 94707004, 397468001, 718200007, 1255886009, 269476000, 307622006, 118606001, 41529000, 716789004, 92512000, 118609008, 1153356005, 277664004, 277474005, 307623001, 95210003, 277624003, 277628000, 419662008, 449058008, 90120004, 1255737000, 307649006, 427141003, 449218003, 450959001, 734066005, 1153382001, 1255752007, 278052009, 399456004, 699818003, 787198005, 277654008, 1255739002, 285776004, 354851000119101, 93488004, 1153355009, 1255807003, 95192000, 404148006, 444597005, 93501005, 1162576007, 835009, 94714002, 350951000119101, 92513005, 93551000, 285420006, 404142007, 766935007, 188592008, 404106004, 128799007, 274905008, 278051002, 450521000124109, 1172704005, 94712003, 404126000, 787594004, 12341000132100, 188567005, 285422003, 277642008, 448995000, 352251000119109, 787036009, 188578002, 1255797003, 449059000, 734076008, 1255894002, 128922003, 404134006, 702977001, 404107008, 93549004, 1172702009, 1255814001, 733917002, 93528000, 277653002, 314934003, 93537000, 1255902005, 303057009, 188559002, 255101006, 404112009, 686991000119106, 1255738005, 1255744009, 1255897009, 188676008, 232075002, 285421005, 450909005, 92516002, 1255805006

#### 3.20 Metastatic Cancer

ICD10CM:
C78.4, C77.3, C78.1, C80.2, C77.0, C80.1, C78.39, C79.32, C79.89, C79.40, C79.82, C79.60, C79.63, C79.71, C78.3, C77.5, C78.30, C77.4, C79.72, C79.70, C77.2, C79.19, C78.00, C79.81, C79.5, C79.0, C78.01, C79.49, C79.4, C77.1, C79.11, C80.0, C78.80, C78.2, C79.61, C79, C78.02, C78.7, C79.9, C79.00, C80, C77.9, C79.31, C78.5, C79.7, C78.8, C79.2, C77, C79.6, C79.1, C78.89, C79.10, C79.3, C78, C79.01, C77.8, C79.52, C78.0, C78.6, C79.51, C79.02, C79.8, C79.62

ICD9CM:
197.5, 197.7, 199, 197.0, 198.1, 198.82, 199.2, 198.0, 198.81, 198.4, 198, 196.2, 196, 198.3, 196.6, 197.4, 196.9, 197, 197.6, 196.3, 196.5, 197.3, 196.0, 197.1, 198.2, 196.1, 198.6, 199.0, 196.8, 198.89, 197.8, 198.8, 199.1, 198.7, 197.2, 198.5

SNOMED CT:
92254003, 92447006, 94002000, 94069006, 94077005, 94462009, 109882003, 188073001, 188159008, 254987003, 369567009, 371969003, 448372003, 448674007, 700423003, 707383000, 707429002, 716855006, 722687007, 833285007, 87151000119105, 681601000119101, 69408002, 93516008, 109385007, 414780005, 352751000119103, 689441000119102, 1196876009, 1255190008, 1255898004, 1259406007, 1259446001, 1259502006, 1259512004, 1259590008, 1259637001, 1260094000, 1268393000, 1268485001, 187947006, 255031003, 255037004, 371965009, 373888000, 427038005, 446939001, 449386007, 722444002, 764855007, 93542008, 16527721000119108, 16906561000119108, 1259367002, 1259653003, 1236948000, 1254727003, 1268532006, 1268638003, 1268955002, 93911008, 94275007, 94404007, 94450005, 94571002, 94572009, 94600009, 109370001, 188282004, 236512004, 302837001, 363497007, 371982006, 403899005, 408644002, 426336007, 447886005, 449219006, 699357004, 716653001, 721579004, 735385007, 763131005, 763408003, 771080008, 785879009, 816205008, 838346000, 1080341000119105, 15635721000119108, 15959061000119101, 94719007, 277609007, 402911002, 1255808008, 1259477004, 1259711008, 1251490009, 93195001, 94129007, 94627008, 188502002, 188666005, 254877001, 275396004, 277617004, 371968006, 448668007, 721644006, 722445001, 735718000, 767444009, 773775004, 782915004, 99111000119103, 10708511000119108, 682041000119100, 1259449008, 1260045000, 1263654002, 1264500002, 93141006, 93689003, 93865007, 93930008, 94000008, 94294002, 94312000, 94518002, 94639000, 187708004, 187821001, 188010006,

188462001, 254733003, 255081007, 363420003, 363517008, 369464004, 369478003, 372005006, 393563007, 403978008, 404141000, 448317000, 448865007, 698040004, 707392002, 733346002, 108101000119101, 130371000119101, 184861000119102, 687981000119106, 93543003, 1079581000119107, 1081361000119102, 681691000119107, 685981000119100, 1255652003, 1259314002, 1254749009, 1264340007, 1268387008, 93872008, 94087009, 94230009, 94266005, 109264009, 109876001, 127246002, 242862004, 254840009, 277549009, 363419009, 369479006, 442537007, 448666006, 707627009, 766046007, 830057003, 870318006, 109962001, 685571000119105, 690141000119103, 1162282006, 1255791002, 1255809000, 1259390007, 1259721000, 1260077002, 1263641009, 92324006, 93638003, 93803002, 94041003, 94515004, 94539009, 94602001, 109887009, 188060000, 188321006, 188674006, 254713002, 255096006, 285641009, 302848008, 314976006, 371964008, 372111007, 403918002, 405546008, 449211009, 713483007, 735916009, 307633009, 1153347002, 689251000119101, 1197276007, 1231732006, 1259695007, 1236943009, 1237467000, 1268539002, 94001007, 94360002, 127236004, 188051008, 188289008, 188319001, 188507008, 255029007, 363402007, 363458004, 369496007, 402499008, 404080006, 404094007, 423158009, 448435005, 449803009, 703387000, 716651004, 133751000119102, 93534007, 187734007, 16905141000119107, 685951000119107, 1255112004, 1255656000, 1259372006, 1259420000, 1259530006, 1259583003, 1237037004, 1268502003, 93133006, 93645003, 93798006, 93993007, 94082003, 94122003, 94191001, 94443006, 94453007, 307601000, 314988008, 315006004, 400122007, 698043002, 721568009, 722677008, 739301006, 1254844005, 1255789005, 1259370003, 1259410005, 1259472005, 1260057009, 1268459000, 93744007, 93991009, 94016005, 94465006, 109821008, 109832008, 127220001, 187738005, 187989003, 188308004, 188648000, 189815007, 372112000, 449637003, 707673006, 721637002, 722666002, 763477007, 363357005, 404810003, 1079071000119101, 16636101000119105, 687841000119105, 1259594004, 1260071001, 1268701008, 93146001, 93667002, 94264008, 94342002, 94345000, 94454001, 94567000, 94716000, 128462008, 187792009, 187972006, 188091006, 188133005, 188252005, 255017003, 274902006, 277626001, 310498001, 402529002, 403946000, 404037002, 413441006, 423424005, 425231005, 428905002, 707388009, 707674000, 785807007, 788972003, 87121000119102, 15956181000119102, 16091171000119106, 16260631000119101, 110005000, 188541000, 684931000119100, 685941000119105, 1255798008, 1259744002, 1259785007, 1237047001, 1237456001, 91860005, 93814004, 93927001, 93944002, 94022001, 94170009, 94218003, 187613005, 187622006, 187926007, 188256008, 254764001, 254938000, 314969001, 363408006, 363410008, 371992003, 372108006, 403979000, 422833009, 449632009, 721605003, 762458004, 770402000, 108211000119108, 130891000119106, 93890009, 188570009, 404135007, 1156416002, 1156460002, 683041000119105, 1179377000, 1197268005, 1259819000, 1260070000, 1260074009, 92135006, 94250008, 127070008, 254632001, 274087000, 276811008, 285769009, 363515000, 369568004, 371134001, 371978009, 402520003, 403714009, 423673009, 449578008, 698045009, 707391009, 733470002, 765741003, 422282000, 1080551000119100, 1162354002, 1254805002, 1260063000, 91855006, 94011000, 95214007, 187942000, 188122007, 188135003, 269460009, 269473008, 369557004, 722515001, 734522002, 1082901000112103, 93541001, 109970006, 359631009, 1078981000119108, 1080581000119107, 15955141000119104, 1197313009, 1254803009, 91854005, 93652001, 93925009, 93957005, 94015009, 94107008, 94343007, 94415002, 187601000, 188209005, 254969001, 255109008, 303055001, 363495004, 423463003, 707581004, 721302006, 721576006, 783704005, 789721005, 682031000119109, 1186869002, 1259376009, 1260039008, 1237050003, 1263657009, 93787005, 94043000, 94115006, 94217008, 94283001, 127232002, 188506004, 254650008, 254828009, 255070009, 255072001, 404117003, 703228009, 765136002, 133531000119104, 1156795003, 16527601000119105, 352391000119109, 689791000119102, 1196911006, 1255655001, 1255795006, 1255796007, 1236954004, 92144007, 94323001, 94395004, 94396003, 94674004, 187633009, 188635004, 369499000, 369510000, 402873007, 403889000, 424052001, 721716004, 723845003, 10737861000119101, 188568000, 188591001, 685701000119105, 939595491000119108, 1163357003, 1186631005, 352551000119106, 1255742008, 1260040005, 1237487004, 1237488009, 1268526001, 93788000, 230156002, 254404003, 286894008,

304545002, 359987004, 371966005, 402509003, 404056007, 413843002, 447707005, 448931007, 699356008, 90831000119105, 109966003, 123731000119106, 1254852008, 1255085007, 1251487003, 1269121005, 93775003, 94417005, 94677006, 95226002, 109837002, 188281006, 188631008, 276419004, 402503002, 402522006, 423295000, 448509007, 449034009, 723182009, 724642009, 765095002, 822969007, 840424008, 107601000119107, 1078881000119102, 1082281000119105, 1081291000119104, 1082241000119100, 1259794001, 1260080001, 1237495000, 92414002, 93816002, 94014008, 94097000, 94247005, 94324007, 188017009, 254874008, 363403002, 404008001, 424078005, 448710000, 449248000, 707397008, 707590006, 713609000, 721577002, 1681000119102, 188575004, 1080561000119103, 1186615002, 1196856003, 1260018001, 1260091008, 1237520001, 1251466008, 1263643007, 92198006, 92200000, 93757004, 94581003, 188634000, 254408000, 254797000, 314408000, 363412000, 369448007, 369507007, 423195009, 441313008, 447100004, 448269008, 448675008, 715904005, 718220008, 721561003, 721672004, 735678002, 782697005, 866072004, 1098981000119101, 15635761000119103, 16091411000119109, 1197332003, 1255799000, 1259401002, 1259662001, 1259697004, 10737961000119108, 1264357006, 93687001, 93701008, 93891008, 94179005, 94640003, 94647000, 254886006, 277569004, 315009006, 363477002, 369511001, 399590005, 402497005, 404113004, 423468007, 448867004, 715412008, 724805000, 13351431000119102, 895358004, 15958021000119103, 687331000119109, 1255358009, 1259330005, 1259532003, 1259563004, 1259693000, 1260115001, 1251495004, 94154007, 94595000, 187903000, 188234005, 188245008, 188503007, 254474002, 309245001, 363475005, 369572000, 373082000, 402500004, 403929003, 448354009, 449630001, 707407001, 715664005, 764735002, 1651000119109, 67821000119109, 681621000119105, 107831000119101, 1196854000, 1236957006, 1269044006, 93190006, 93809003, 93996004, 94120006, 94244003, 94442001, 94473002, 94486006, 94658009, 118607005, 187791002, 187927003, 187945003, 188065005, 188193007, 188269007, 315003007, 363431006, 369533004, 369551003, 447712006, 721580001, 792907004, 827162007, 116711000119103, 421283008, 890528009, 1081351000119104, 685641000119102, 1197331005, 1255385002, 1255725002, 1259491005, 1264116009, 1268392005, 93714009, 94112009, 94540006, 187681002, 187726001, 253001006, 255039001, 276751004, 277574007, 363365008, 369476004, 372014001, 373081007, 424454009, 448711001, 449065000, 698646006, 707470000, 713574000, 722513008, 68261000119102, 188029000, 277610002, 1010624006, 1079011000119109, 1079051000119105, 1156469001, 1254786005, 1254825003, 1259672003, 1263911005, 93145002, 93684008, 93842002, 94035005, 94330007, 94463004, 95186006, 109831001, 187864008, 188121000, 188180002, 254631008, 363349007, 363452003, 363496003, 369505004, 707408006, 823017009, 61301000119102, 96901000119105, 107791000119107, 93496009, 1082181000119103, 689851000119102, 1186614003, 1260059007, 1251484005, 93760006, 94195005, 94478006, 94506006, 94607007, 94641004, 187798008, 188307009, 254601002, 254637007, 278433008, 314975005, 363453008, 369518007, 372105009, 402526009, 12311000132101, 107771000119106, 188529007, 1010681001, 1078971000119105, 1156410008, 16774591000119105, 688061000119104, 1196998001, 1197285007, 1255743003, 1259435004, 1259813004, 1260032004, 93191005, 93650009, 93776002, 94489004, 94493005, 94671007, 187666008, 188033007, 188313000, 254611009, 269515006, 277543005, 363450006, 404154007, 404156009, 448371005, 449628003, 713189001, 764952009, 787091002, 816203001, 122961000119105, 353441000119103, 1080981000119104, 404157000, 1081721000119106, 1153345005, 15955941000119102, 15958101000119106, 16782241000119104, 1259486009, 1259830002, 1254735000, 1268486000, 1268538005, 91856007, 93958000, 94494004, 94528006, 109991003, 110006004, 127255004, 187862007, 188103003, 254732008, 254824006, 255074000, 363405009, 363449006, 363498002, 369495006, 369536007, 369544007, 403915004, 404137004, 445738007, 707359008, 721700001, 733162002, 818967003, 117211000119105, 188512009, 188672005, 1010656004, 687421000119101, 688081000119108, 1197322005, 1259340008, 1259580000, 1259643004, 1259700000, 1237041000, 1268534007, 1269123008, 92268005, 93200005, 93655004, 93862005, 93912001, 94113004, 94255003, 94500000, 109992005, 127251008, 187700006, 276804009, 369602008, 371984007, 372017008, 107581000119103, 117091000119108, 277611003,

1156404008, 1156806002, 686461000119108, 1259829007, 1268716006, 93770008, 94098005, 94215000, 94298004, 94327000, 94344001, 95187002, 187969004, 254625005, 254792006, 254974009, 255078002, 277627005, 312104005, 315000005, 363383000, 363421004, 369483006, 369595009, 370511006, 403924008, 423793008, 429033009, 724467001, 458321000124102, 1081021000119109, 1081561000119108, 118605002, 363510005, 16905581000119107, 687971000119108, 689831000119108, 1255661003, 1259528009, 1260072008, 1237461004, 1237524005, 1254729000, 1268388003, 1268540000, 93692004, 93772000, 93945001, 94231008, 94662003, 109348004, 188000002, 254876005, 301756000, 313250007, 315002002, 363429002, 363441009, 369512008, 373168002, 404087009, 443144000, 448774004, 713327005, 734015000, 762457009, 822970008, 107591000119100, 1079101000119105, 351251000119103, 1208744003, 1259660009, 1236965009, 1254731009, 1268545005, 93801000, 93837005, 93878007, 93917007, 94084002, 94287000, 94625000, 94631002, 188492005, 254878006, 269533000, 369576002, 403940006, 404081005, 404091004, 404118008, 707349007, 707421004, 707452003, 716652006, 763309005, 45221000119105, 188513004, 1010603001, 1144252005, 1156409003, 683761000119107, 689371000119102, 1196900003, 1208745002, 1259391006, 1259745001, 1268352008, 1268905005, 93228004, 93998003, 94110001, 94182000, 94560003, 94635006, 94657004, 109367000, 187608006, 188062008, 188191009, 269581007, 276809004, 276962007, 403913006, 449309003, 707377003, 707472008, 789689004, 107571000119101, 188589009, 1080531000119106, 1217020002, 1259423003, 1251492001, 93229007, 93665005, 94024000, 94194009, 94273000, 94367004, 94548004, 94580002, 94594001, 94675003, 109870007, 109931000, 363461003, 369563008, 371962007, 423284006, 733834006, 737312002, 762690000, 783183009, 91181000119105, 116381000119105, 127961000119108, 1080111000119108, 254727007, 898197001, 350681000119101, 352481000119103, 1255813007, 1259332002, 1259374007, 1259645006, 1259716003, 1268639006, 1268720005, 94003005, 94131003, 94204005, 94285008, 94370000, 94590005, 94628003, 94690004, 188034001, 254551004, 286887005, 302816009, 363351006, 371972005, 372096000, 404071006, 404127009, 404169008, 417570003, 423812005, 707457009, 721698005, 722678003, 766758001, 766979005, 71111000119109, 681651000119102, 1081591000119101, 1156452003, 1156810004, 1197343006, 1259455003, 1259742003, 1260019009, 1237428006, 94067008, 94140004, 94551006, 127237008, 187865009, 187900002, 187906008, 188195000, 188325002, 254708001, 254986007, 255086002, 280959007, 303017006, 371975007, 423480004, 707390005, 781382000, 1078931000119107, 10749871000119100, 188562004, 307635002, 1080701000119109, 15956701000119109, 16637101000119107, 354081000119101, 1229871006, 1259357007, 1259523000, 1259543006, 1236968006, 1237498003, 1237529000, 1251489000, 1254732002, 1263659007, 1268386004, 1268705004, 1268710000, 1268904009, 94265009, 94436000, 94618007, 123313007, 127245003, 187844003, 254424004, 277572006, 278046008, 363369002, 363500001, 369485004, 369488002, 448274000, 707342003, 713897006, 733362008, 93494007, 93547002, 445269007, 878808006, 1080051000119105, 1080721000119100, 15977551000119100, 352331000119105, 352701000119102, 688011000119102, 1259733005, 1259753009, 1260083004, 1263656000, 1269413009, 92416000, 93653006, 93796005, 93947009, 93974005, 94046008, 94142007, 94460001, 94637003, 94718004, 276799004, 281702006, 369472002, 369516006, 369603003, 371983001, 403906006, 404121005, 420788006, 422572002, 428100006, 449268006, 721607006, 724554001, 1010679003, 895345000, 10976631000119101, 352381000119106, 1259798003, 1237528008, 93771007, 93892001, 94051002, 94296000, 94545001, 94553009, 276860003, 302841002, 307592006, 363355002, 369522002, 369528003, 369596005, 372025005, 403916003, 404055006, 448221005, 449217008, 449318001, 707426009, 713325002, 721311006, 722685004, 788874003, 79281000119106, 91061000119100, 103511000119103, 93522004, 1010607000, 1082271000119107, 684901000119107, 1162351005, 1255750004, 1259381000, 1259684001, 1237465008, 1263998000, 93222003, 93846004, 93916003, 93971002, 94347008, 94364006, 188030005, 188064009, 188110009, 254654004, 255075004, 373090000, 403941005, 404069006, 404144008, 423535002, 427685000, 448220006, 707596000, 721314003, 721718003, 770602005, 354671000119106, 93514006, 93523009, 110007008, 1010674008, 1148904003, 681751000119106, 352581000119104, 1259382007, 1260075005, 1268715005,

92199003, 93710000, 93812000, 94052009, 94297009, 109879008, 127267002, 187658004, 190818004, 254437001, 278491007, 314993006, 363366009, 363508008, 369502001, 702476004, 733356003, 764940002, 836274002, 866070007, 1701000119104, 10769100011910, 457721000124104, 1080211000119103, 1082311000119107, 110004001, 1080071000119101, 1157062009, 682231000119100, 145298771000119104, 1259434000, 1263450003, 45061000087102, 93871001, 94156009, 94192008, 109996008, 187604008, 187631006, 187853005, 187948001, 188020001, 188290004, 188478004, 276954004, 281565009, 363386008, 397011009, 398670003, 403922007, 404149003, 423325007, 444712000, 447656001, 447710003, 707423001, 724058006, 724060008, 133881000119100, 351961000119109, 423829008, 1153354008, 1156417006, 682541000119106, 1268962006, 93216007, 93899005, 94245002, 94362005, 94441008, 94458003, 9459008, 94591009, 94616006, 94648005, 94688000, 187698002, 187833006, 187841006, 187992004, 276975007, 363385007, 369542006, 371996000, 372121004, 413842007, 418372008, 449292003, 723301009, 725390002, 733345003, 461371000124103, 93555009, 187752007, 255114007, 15930821000119105, 1237578005, 1259705005, 1259759008, 1259799006, 1260110006, 93763008, 93956001, 94058008, 94102002, 94267001, 94619004, 94668004, 95193005, 95264000, 109848009, 109925003, 187697007, 187976009, 188718006, 189814006, 254863004, 415287001, 447883002, 448554008, 449073009, 721627007, 773624006, 10745291000119103, 1144945000, 1157061002, 684941000119109, 1259350009, 1259514003, 10738001000119105, 1237491009, 1240346000, 1251486007, 93186009, 93977003, 94145009, 94162004, 94259009, 94366008, 94513006, 94535003, 94582005, 255056009, 276821000, 363406005, 371971003, 372013007, 404017001, 698287002, 707487003, 721623006, 721626003, 763884007, 109967007, 414166008, 1082261000119101, 1156414004, 350631000119102, 689491000119105, 1251564008, 1255687003, 1259589004, 1236955003, 1263652003, 94426008, 94544002, 94686001, 94711005, 109945006, 187916000, 188637007, 314964006, 363378008, 363469001, 369604009, 413445002, 449101009, 707582006, 707705008, 733349009, 866094007, 91171000119107, 123631000119103, 123781000119107, 188590000, 143291000119104, 15931021000119108, 1196881000, 1268396008, 1268537000, 1268712008, 94139001, 94422005, 94476005, 94485005, 94561004, 109985000, 187694000, 188001003, 188301005, 188662007, 254434008, 254652000, 278453007, 285598005, 363501002, 403925009, 423005002, 445406001, 447266004, 448465000, 448555009, 448559003, 448954003, 449096009, 707339009, 724468006, 778066006, 838355002, 352791000119108, 1259366006, 1259498004, 1259810001, 1260042002, 1237518004, 1251497007, 93225001, 93745008, 93994001, 94093001, 127259005, 188090007, 188283009, 310526005, 372128005, 413446001, 448233000, 448319002, 449206007, 708921005, 721557009, 733136006, 788982002, 870327007, 352201000119105, 1081751000119103, 1141626005, 1156783003, 1157141006, 681991000119104, 685611000119101, 1255885008, 1259176001, 1259327003, 1259484007, 1259521003, 1259577001, 1263467005, 1268897007, 61493004, 92073008, 93651008, 94279001, 94474008, 188235006, 254462001, 278050001, 314961003, 314973003, 369470005, 369565001, 400058002, 448215006, 712750007, 786710002, 1082291000119108, 1082121000119102, 1157162007, 130941000119107, 682311000119102, 685851000119104, 1255890006, 1259520002, 1254750009, 1264498003, 1268901001, 93952004, 94006002, 94168000, 94306005, 94400003, 94411006, 94562006, 94573004, 94621009, 127274007, 188733003, 254626006, 254856004, 363417006, 363516004, 369538008, 404014008, 415110002, 415286005, 425048006, 425688002, 448248006, 707464006, 725437002, 109979007, 1079201000119101, 16823941000119108, 1255892003, 1259633002, 1259719005, 1254755004, 1268702001, 1268898002, 93784003, 93902008, 93940006, 94175004, 94449005, 127269004, 188071004, 188089003, 188324003, 313427003, 369500009, 402819001, 404128004, 447596005, 449497006, 721556000, 722527003, 766978002, 100721000119109, 118613001, 1082171000119101, 15950141000119105, 353811000119104, 1197299006, 1197279000, 1254847003, 1255812002, 1236931008, 1240355002, 1263475004, 1268460005, 1268488004, 1268518009, 94121005, 94529003, 94584006, 188741003, 254412006, 276953005, 285615003, 307576001, 363464006, 371967001, 447705002, 447730004, 723854000, 724645006, 735917000, 840423002, 105111000119109, 188587006, 15958381000119101, 1254787001, 1259427002, 1259437007, 1259803007, 1263466001,

20224008, 93220006, 93712008, 93755007, 93854002, 93894000, 94068003, 94169008, 94171008, 94186002, 94378007, 94495003, 94520004, 94565008, 109843000, 187994003, 188323009, 254973003, 314962005, 369469009, 369503006, 372116002, 424486004, 440501006, 710195004, 716649003, 716657000, 723843005, 733355004, 733598001, 735919002, 763666008, 99101000119101, 118617000, 1255726001, 1259251005, 1259447005, 1251471001, 93729006, 94274006, 94371001, 188454009, 254425003, 254887002, 363396004, 369575003, 372010005, 403895004, 417417007, 421249001, 609515005, 707353009, 722954005, 763479005, 866176003, 91141000119100, 236811000119101, 1080101000119105, 93489007, 895354002, 1079141000119107, 1255746006, 1259727001, 1259731007, 1260021004, 1268536009, 92251006, 93690007, 93972009, 94029005, 94155008, 94512001, 127271004, 254555008, 281564008, 314191009, 363423001, 369468001, 402563000, 448165009, 448298007, 448738008, 188572001, 1082231000119109, 1156786006, 1259439005, 1259457006, 1259535001, 1259626002, 1259726005, 1237455002, 1251474009, 36222008, 93715005, 93777006, 94185003, 94336001, 94527001, 187693006, 188154003, 240163000, 254638002, 255004001, 307625008, 312115000, 314989000, 363411007, 369490001, 371986009, 443520009, 448558006, 133871000119103, 92511007, 682241000119109, 1259561002, 1259720004, 1268641007, 92282005, 94118008, 94271003, 94328005, 94346004, 94556001, 95209008, 188649008, 302851001, 363502009, 369562003, 424849005, 430556008, 448450001, 702785000, 707346000, 717922007, 721673009, 68291000119109, 105121000119102, 354351000119105, 1082051000119106, 1144928005, 1259399008, 1259623005, 93196000, 93900000, 93961004, 93985007, 94369001, 94604000, 109844006, 109927006, 187952001, 188004006, 188072006, 188729005, 254730000, 277619001, 285607001, 285631006, 363416002, 369521009, 400074007, 424276002, 424487008, 443488001, 703626001, 707350007, 717736007, 721696009, 770685009, 773774000, 188537004, 1080541000119102, 1153422000, 1156403002, 15977631000119105, 353721000119100, 145194251000119106, 1255644001, 1255647008, 1255666008, 1259680005, 1259805000, 1237039001, 1263660002, 92441007, 93693009, 94103007, 94152006, 94208008, 94280003, 94393006, 94656008, 109836006, 254629004, 707424007, 1080201000119101, 188516007, 404122003, 10737781000119101, 1081611000119106, 1144891007, 1157064005, 16527481000119100, 243971001000119103, 688891000119103, 1259334001, 1259408008, 1259429004, 1259715004, 1237414000, 1240351006, 1263653008, 1269458003, 93169003, 94049001, 94089007, 94423000, 127266006, 187641009, 188609000, 254955001, 281563002, 307502000, 369550002, 372007003, 402541009, 404038007, 433067002, 116811000119106, 147101000119108, 1081571000119102, 890534002, 1156461003, 145511000119106, 15951061000119102, 15958301000119109, 1217490003, 1259537009, 1236962007, 1263655001, 1269122003, 93677000, 93901001, 94257006, 187861000, 363377003, 363447008, 448377009, 448669004, 721641003, 721669006, 723077004, 773537001, 866079008, 866087009, 116871000119103, 1010669000, 1080021000119102, 1162286009, 1197324006, 1197260003, 1259387001, 1259400001, 1259412002, 1259724008, 1259748004, 1260116000, 1237415004, 1251494000, 1264068000, 93857009, 93888008, 188003000, 188500005, 254731001, 254748009, 399326009, 402544001, 402562005, 403917007, 408643008, 422782004, 449636007, 698048006, 707491008, 707592003, 766048008, 87131000119104, 184801000119103, 93527005, 254898001, 12241031000119108, 145501000119108, 689451000119100, 1259368007, 1259585005, 1259776007, 1260033009, 1264458000, 1269045007, 93695002, 94687005, 109378008, 118611004, 188005007, 254547001, 254851009, 302823005, 307624007, 369467006, 414553000, 448217003, 709517003, 709830006, 716648006, 721301004, 721547008, 128041000119107, 130351000119105, 71101000119106, 1081321000119107, 1081701000119102, 400375351000119102, 1197266009, 1259474006, 1260067004, 1240352004, 92306009, 93193008, 93793002, 93815003, 93849006, 93966009, 94005003, 94196006, 94557005, 109267002, 109939003, 187941007, 188016000, 254445006, 285603002, 314997007, 372133009, 402518001, 402530007, 408646000, 448376000, 448882009, 448990005, 448993007, 449418000, 698011002, 707471001, 735680008, 783541009, 184891000119109, 1079111000119108, 424779008, 1078951000119101, 1079641000119101, 1080711000119107, 16527441000119105, 681611000119103, 1197328009, 1259325006, 1259433006, 1259822003, 1260031006, 1268355005, 93943008, 94073009,

94117003, 94310008, 94419008, 94480000, 94612008, 109994006, 127249009, 188302003, 188641006, 231829006, 254714008, 255191003, 363354003, 369524001, 369554006, 369584003, 402515003, 449054005, 707361004, 707662004, 722672002, 783155007, 1079191000119104, 188565002, 1079121000119101, 686431000119100, 1162531007, 1197294001, 1259403004, 1259567003, 1259823008, 1237422007, 93192003, 93826009, 94189009, 94213007, 94236003, 94242004, 94246001, 94386007, 94503003, 188002005, 254635004, 254726003, 363373004, 363398003, 371991005, 414676007, 447989004, 707703001, 713516007, 733134009, 766247009, 1153349004, 351041000119109, 682481000119100, 688981000119107, 689281000119108, 1254781000, 1255727005, 1255745005, 1259394003, 1259763001, 1259780002, 1237483000, 1268640008, 92213006, 93679002, 93806005, 94133000, 94206007, 94249008, 94468008, 94537006, 127234001, 188737002, 188746008, 255112006, 269469005, 303056000, 363352004, 363505006, 363518003, 369460008, 372244006, 403893006, 413442004, 424190005, 708504008, 715903004, 721563000, 721643000, 764938007, 122951000119108, 1080941000119109, 1144304002, 87141000119108, 1197283000, 1259402009, 1268966009, 1269052009, 92215004, 93915004, 94111002, 94307001, 94359007, 109842005, 187652003, 188168005, 254402004, 254465004, 254771006, 276836002, 369471009, 369497003, 372003004, 402561003, 448952004, 721549006, 723847006, 723852001, 93495008, 93509007, 94407000, 188579005, 188585003, 277550009, 353801000119102, 1187622004, 1259395002, 1259489002, 1259790005, 1260092001, 1268533001, 94446003, 94456004, 187640005, 187742008, 187918004, 187932002, 424413001, 447783007, 449260004, 722676004, 723853006, 838344002, 431911000124102, 459411000124109, 10737561000119103, 1081891000119103, 1156785005, 1254783002, 1255802009, 1259337008, 1259760003, 1269047004, 1285321001, 93201009, 94290006, 94521000, 94632009, 187682009, 188318009, 255129009, 307647008, 369514009, 369555007, 372124007, 402540005, 403890009, 425230006, 448436006, 448992002, 1082071000119102, 93518009, 15956661000119102, 1162346004, 1237473004, 1240353009, 1251485006, 92814006, 93868009, 94161006, 94225005, 254609000, 277573001, 277637000, 369582004, 371973000, 415283002, 425869007, 448447004, 93493001, 93533001, 123661000119106, 1144303008, 1197334002, 1259175002, 1259747009, 1237457005, 1268961004, 1269042005, 92202008, 93898002, 94200001, 94420002, 94598003, 94645008, 109933002, 187831008, 237832001, 369597001, 372136001, 400174005, 440173001, 448216007, 448378004, 449053004, 449259009, 707492001, 707528007, 721636006, 722686003, 781076008, 352311000119100, 1080261000119100, 353511000119101, 10737521000119108, 107811000119106, 1081341000119101, 15958181000119103, 682361000119104, 688811000119107, 1179390007, 1259801009, 1259802002, 1260037005, 1268634001, 1268909004, 40312006, 93213004, 93705004, 94042005, 187688008, 187804005, 277643003, 313428008, 372027002, 372107001, 397015000, 402532004, 403902008, 404138009, 405945003, 420519005, 449077005, 723076008, 726721002, 733144006, 353561000119103, 188576003, 402910001, 16091371000119108, 16905461000119104, 353571000119109, 681701000119107, 688731000119109, 1259450008, 1259481004, 1259569000, 1259723002, 1260076006, 1251460002, 1254728008, 94012007, 94209000, 94425007, 94655007, 188013008, 188726003, 254457002, 254872007, 255030002, 255066001, 363443007, 369452007, 369454008, 425657001, 448387008, 702392008, 703429003, 704152002, 707343008, 707587000, 722683006, 724536000, 724552002, 733064004, 29431000119108, 1156833003, 682381000119108, 688931000119106, 1254957001, 1259341007, 1259428007, 1259615008, 1259783000, 1251491008, 1268965008, 92143001, 93202002, 93906006, 94039004, 94356000, 109853004, 187866005, 277589003, 302855005, 313354001, 363404008, 369540003, 369606006, 403892001, 441559006, 443675005, 448388003, 707484005, 713572001, 721621008, 786038001, 117061000119101, 133841000119105, 1080241000119104, 1081741000119100, 1156456000, 12246481000119106, 15956741000119106, 352961000119103, 688751000119103, 550991000124107, 1259392004, 1259692005, 1259736002, 1259786008, 1237477003, 1251468009, 1263997005, 1268351001, 92104000, 92156008, 94081005, 94301000, 94649002, 127264009, 127272006, 187740000, 188156001, 189924002, 254837009, 277545003, 303012000, 369573005, 369590004, 372138000, 399969009, 402506005, 448254007, 449416001, 707385007, 96281000119107, 93505001, 93536009,

16906481000119103, 353081000119103, 1197263001, 1255649006, 1259566007, 1259770001, 1259782005, 112691000112107, 1259237005, 92028001, 93209006, 93686005, 93876006, 93941005, 94132005, 94248000, 94416001, 94620005, 94679009, 109368005, 187937008, 188102008, 254734009, 254940005, 255071008, 307617006, 363384006, 363482009, 402504008, 402521004, 707451005, 722827008, 123701000119104, 353521000119108, 1082191000119100, 15959101000119103, 109965004, 1079611000119100, 15958741000119107, 351051000119106, 688801000119109, 689781000119100, 1179389003, 1255393002, 1255632004, 1254734001, 1268958000, 94028002, 94412004, 95260009, 109374005, 127252001, 254895003, 255090000, 255128001, 307591004, 363745004, 369492009, 372139008, 402539008, 402820007, 447806008, 448386004, 717735006, 723887001, 116741000119104, 19090001000004101, 118599009, 898198006, 1156406005, 681971000119100, 1259396001, 1260058004, 1264367001, 1268391003, 39795003, 93986008, 94030000, 94268006, 94337005, 123842006, 188177003, 363466008, 402871009, 408647009, 445105005, 449220000, 733348001, 770686005, 780817000, 866069006, 98981000119103, 188517003, 1081761000119101, 10976711000119104, 1144307009, 1153425003, 16775311000119107, 16905541000119102, 688031000119107, 689401000119104, 1255811009, 1259321002, 1259551009, 1259758000, 1237476007, 1254747006, 93658002, 93834003, 94017001, 94623007, 94921006, 109359008, 187784000, 189847002, 254897006, 255021005, 277571004, 278054005, 307341004, 312111009, 315008003, 371990006, 402495002, 403927001, 404172001, 417554000, 424887002, 446189008, 448930008, 449377002, 699317002, 773283006, 870355006, 1080471000119105, 894171571000119107, 1259409000, 1259582008, 1259735003, 1259777003, 1260061003, 1263465002, 1263912003, 94079008, 94239005, 94331006, 94457008, 94501001, 94570001, 187661003, 254975005, 286897001, 363391009, 369501008, 371993008, 372127000, 402498000, 447706001, 448384001, 707360003, 721555001, 721629005, 723879002, 735679005, 93510002, 1010675009, 16527561000119105, 352921000119108, 353061000119107, 1197329001, 1196874007, 1255786003, 1259451007, 1259497009, 1237044008, 1237480002, 1251459007, 1268908007, 93913006, 94219006, 94220000, 94278009, 94304008, 94379004, 94704006, 254701007, 254917006, 255032005, 285635002, 363353009, 363487003, 369465003, 371989002, 402507001, 448709005, 681721000119103, 687891000119102, 689821000119105, 985355341000119101, 1237341006, 1255801002, 1259734004, 1237454003, 91987008, 92149002, 93221005, 93820003, 93880001, 93981003, 93989001, 94124002, 94135007, 94201002, 94241006, 94467003, 94609005, 94708009, 109998009, 187829004, 277632006, 363489000, 363493006, 363503004, 372140005, 404046008, 707467004, 711414003, 716593008, 721602000, 722517009, 766045006, 838343008, 87101000119106, 1081581000119104, 93497000, 93520007, 188515006, 15951021000119107, 16527521000119100, 1177173001, 1197327004, 1259326007, 1259353006, 1259438002, 1259702008, 1264416003, 68979007, 93681000, 93928006, 94116007, 94221001, 109847004, 109919002, 109989006, 127262008, 187944004, 188050009, 188668006, 254843006, 255040004, 255068000, 307593001, 307603002, 313356004, 314990009, 314996003, 363381003, 363514001, 372101000, 400175006, 404108003, 422541001, 422679002, 443487006, 699354006, 707628004, 707660007, 722680009, 104981000119104, 1082331000119102, 12246561000119101, 895356000, 1081901000119104, 15955901000119104, 1255192000, 1255785004, 1259722007, 1236941006, 1237527003, 1269043000, 92811003, 93778001, 94263002, 94295001, 94491007, 254563009, 369574004, 417181009, 447738006, 448775003, 722665003, 838342003, 11471000224106, 24111000119105, 188577007, 1082141000119108, 1255806007, 1260060002, 1237057000, 1237521002, 93694003, 94187006, 94233006, 94498001, 94525009, 127233007, 127270003, 254844000, 254852002, 281560004, 285642002, 372020000, 403729006, 403920004, 404120006, 448451002, 714463003, 783615009, 866052009, 118601006, 1148851002, 1153341001, 16845341000119105, 1255753002, 1259356003, 1259419006, 1259448000, 1259821005, 1251456000, 93194002, 93699008, 93978008, 94123008, 110013004, 187940008, 254918001, 277587001, 363462005, 402876004, 403712008, 423610004, 443679004, 707593008, 733352001, 733359005, 91151000119103, 1082111000119109, 1187464007, 1259773004, 93187000, 94092006, 94243009, 94353008, 187950009, 187978005, 188074007, 231831002, 254559002, 277530005, 314992001, 403891008, 404015009, 447390000, 702391001, 707337006, 707466008, 721620009, 353661000119102,

423294001, 143401000119106, 1079531000119106, 1080691000119109, 1099571000119100, 1155991005, 1186612004, 1259749007, 1259768005, 1268350000, 93781006, 94270002, 94340005, 94522007, 109369002, 187801002, 188327005, 254431000, 254567005, 254820002, 271943005, 274085008, 277601005, 285640005, 302849000, 312112002, 369461007, 369564002, 372099007, 698044008, 700488005, 721635005, 788979007, 7391000119103, 21851000119103, 1079061000119107, 1144924007, 16774671000119108, 685671000119109, 1259343005, 1259375008, 1259678004, 1259738001, 1237034006, 1237051004, 1237426005, 1260217004, 1268484002, 1268503008, 1268718007, 93217003, 93636004, 93713003, 93740003, 93908007, 93920004, 94410007, 94511008, 94654006, 187824009, 188061001, 188292007, 188498009, 255008003, 276829003, 277625002, 313429000, 314968009, 404039004, 443136000, 449269003, 722103009, 1081841000119106, 1156472008, 689501000119103, 1208460000, 1259422008, 1259817003, 1260082009, 1260085006, 1263658004, 92280002, 94019003, 94032008, 94108003, 94211009, 94466007, 188022009, 269580008, 275397008, 363488008, 371979001, 372087000, 404093001, 426071002, 352321000119107, 93552007, 109972003, 1081281000119102, 685621000119108, 682461000119109, 1254806001, 1255629002, 1259313008, 1237032005, 1237519007, 1240363001, 93711001, 93969002, 94341009, 94564007, 94613003, 188063003, 254703005, 277618009, 363375006, 424600001, 426217000, 448401007, 717921000, 830150003, 456911000124108, 459391000124109, 1078901000119100, 118600007, 402881008, 685831000119105, 687821000119104, 1255660002, 1260064006, 1237043002, 1240357005, 1251445000, 1268389006, 1268637008, 93671004, 93782004, 94083008, 94319009, 94444000, 94464005, 188272000, 188501009, 241861008, 369559001, 369560006, 371995001, 373083005, 403905005, 404072004, 721558004, 723849009, 766248004, 830151004, 61471000119100, 351211000119104, 353421000119109, 109964000, 1156418001, 681781000119104, 685721000119101, 1208483002, 1259509002, 1259602004, 1259704009, 1259812009, 1260268001, 93182006, 93707007, 93726004, 93830007, 94050001, 94094007, 94508007, 94588009, 95194004, 109840002, 109947003, 187635002, 188242006, 302842009, 363350007, 363367000, 363413005, 398903003, 404111002, 425941003, 447800002, 449173006, 699537002, 707531008, 721640002, 116841000119105, 208041000119100, 354701000119107, 681631000119108, 123761000119103, 683131000119100, 1197311006, 1197330006, 1208349005, 1259378005, 1259462007, 1259820006, 1237048006, 1263468000, 1268462002, 1268698000, 93893006, 94672000, 109839004, 255028004, 277613000, 285610008, 314951005, 363382005, 369463005, 369527008, 369583009, 372019006, 372097009, 423121009, 699659007, 188569008, 188593003, 733627006, 78461000119105, 1157063004, 688681000119100, 1237340007, 1259453005, 1259466005, 1259493008, 1268956001, 92419007, 93767009, 94253005, 94403001, 94452002, 94681006, 369486003, 369588000, 404115006, 430338009, 440422002, 449153001, 722953004, 723173003, 724647003, 764846009, 359640008, 682451000119107, 684921000119103, 1259442004, 1259518000, 1268888008, 94569002, 187973001, 254441002, 254645002, 314965007, 314994000, 371988005, 402505009, 402511007, 402513005, 402558004, 404067008, 424666005, 443648003, 447781009, 449254004, 705061009, 713038003, 722512003, 735735001, 143411000119109, 93492006, 188531003, 1148845007, 15950221000119108, 687861000119109, 688411000119109, 681921000119101, 1254801006, 1255663000, 1255754008, 1259732000, 1259816007, 1268461009, 93139005, 93751003, 94352003, 94483003, 94526005, 94678001, 187793004, 189509003, 231834005, 255069008, 255087006, 363418001, 369558009, 397009000, 707583001, 707595001, 717731002, 721541009, 764990003, 9541000119105, 67841000119103, 109921007, 188547001, 1156807006, 15949941000119101, 352461000119107, 685901000119108, 1197342001, 1254691000, 1254733007, 1263484004, 1268633007, 1268699008, 93654000, 94033003, 94148006, 94388008, 94633004, 187811009, 187869003, 187997005, 188261005, 255107005, 314987003, 363481002, 369556008, 403894000, 448863000, 449108003, 721540005, 722707001, 735332000, 135091000119106, 109392002, 15958221000119106, 353791000119103, 689471000119109, 1255645000, 1255888005, 1259384008, 1259775006, 1237033000, 1268544009, 1268563005, 1279841001, 93135004, 93747000, 94020009, 94163009, 94334003, 94391008, 94394000, 188613007, 188627002, 254561006, 254710004, 269467007, 277602003, 363506007, 403939009, 441535001, 449308006, 699704002,

707485006, 707493006, 716788007, 61311000119104, 133861000119109, 93521006, 1156812007, 682291000119101, 1217301006, 1259452000, 1259668002, 1259755002, 1259796004, 1269126000, 93963001, 94205006, 94496002, 94499009, 94586008, 187653008, 254398008, 269459004, 707395000, 735921007, 15956341000119105, 359648001, 878858004, 689771000119103, 1229988003, 1255658004, 1259756001, 1237481003, 1254730005, 92197001, 93742006, 93828005, 94153001, 94305009, 94470004, 95230004, 109828002, 187809000, 188076009, 231833004, 254909003, 255012009, 314998002, 369498008, 372103002, 403954003, 422479008, 424993006, 445448008, 449075002, 707460002, 716859000, 722543005, 734099007, 459401000124106, 1080121000119101, 12240951000119107, 93525002, 404143002, 123721000119108, 1079021000119102, 1080571000119109, 354391000119100, 1187123005, 1255630007, 1259344004, 1259825001, 1268546006, 92274005, 93983000, 94013002, 94176003, 94414003, 94610000, 187870002, 188297001, 285605009, 285616002, 302845006, 369525000, 369545008, 404119000, 440525003, 707355002, 713290004, 733608000, 68271000119108, 145831000119103, 684911000119105, 15956381000119100, 188511002, 1082131000119104, 1153426002, 1156495004, 1156808001, 1157071000, 16899941000119102, 687361000119101, 1186616001, 1162311007, 1254790007, 1255654002, 1255788002, 1259369004, 1259479001, 1259769002, 1236942004, 1251457009, 1251472008, 93214005, 94099002, 94235004, 94429001, 123845008, 254655003, 402525008, 402882001, 404147001, 413389003, 422853008, 425178004, 449063007, 707453008, 707489000, 721699002, 108201000119105, 461351000124108, 15957181000119104, 353111000119108, 1196879002, 1264118005, 1281812003, 93700009, 93817006, 93845000, 94075002, 94587004, 188487008, 188754005, 253003009, 255042007, 255077007, 276822007, 277551008, 277568007, 277579002, 285639008, 313353007, 372008008, 427056005, 447885009, 448315008, 449177007, 449221001, 449419008, 724649000, 724806004, 733603009, 109968002, 895355001, 1080041000119108, 1153348007, 1197280002, 1255657009, 1255792009, 1259336004, 1259424009, 1259500003, 10737681000119106, 1237522009, 1254723004, 1264258003, 1268349000, 1268704000, 92271002, 93663003, 93825008, 94193003, 94326009, 94597008, 187977000, 188280007, 254520006, 254794007, 277156006, 286899003, 363454002, 363509000, 372126009, 372135002, 424229006, 424260006, 448299004, 449055006, 710196003, 733357007, 118608000, 895111002, 1157060001, 1259475007, 1264115008, 134421000, 187660002, 187884007, 187920001, 188042000, 188055004, 236513009, 254435009, 254995004, 269617008, 307651005, 363388009, 402510008, 403903003, 408642003, 448314007, 449487002, 697993003, 707575007, 721306009, 721567004, 772992009, 1079171000119100, 109386008, 188679001, 352731000119109, 689231000119107, 1186630006, 1201780008, 1255887000, 1259414001, 1259458001, 1259789001, 1259814005, 1260086007, 1236964008, 1268957005, 1268964007, 93749002, 94479003, 94626004, 109366009, 109371002, 271468000, 315007008, 449074003, 707486007, 722514002, 770684008, 866050001, 352001000119100, 11010461000119101, 1082091000119101, 687351000119103, 1255096004, 1259750007, 1259797008, 1237045009, 1268499002, 1269273000, 93709005, 94322006, 94351005, 94603006, 94629006, 187727005, 187854004, 188009001, 188188009, 254539001, 363374005, 363399006, 373088001, 402538000, 403901001, 404140004, 448468003, 448864006, 721670007, 722511005, 764737005, 866073009, 1081651000119107, 1259388006, 1259779000, 1237471002, 1237526007, 93640008, 93789008, 93995000, 94034009, 94408005, 94549007, 94579000, 94663008, 187929000, 188220005, 254712007, 254948003, 254993006, 277615007, 285611007, 313248004, 363446004, 363486007, 423816008, 707468009, 707579001, 724553007, 683031000119101, 91211000119109, 1255628005, 1255804005, 1259454004, 1259761004, 1260084005, 1237492002, 1264523004, 93675008, 93850006, 93889000, 94314004, 94381002, 94709001, 187857006, 300988009, 307636001, 314960002, 359785002, 403911008, 448229007, 449156009, 723850009, 96981000119102, 97051000119105, 12240991000119102, 404136008, 1153377008, 1157067003, 682251000119106, 1231267007, 1237474005, 93648001, 93657007, 93970001, 94172001, 188100000, 254888007, 269516007, 303194003, 363451005, 372023003, 372106005, 404052009, 404068003, 448560008, 715414009, 721548003, 783056006, 188566001, 878857009, 1081331000119105, 16229011000119100, 1255648003, 1259322009, 93185008, 94027007, 94281004, 94311007, 94574005, 94666000, 95188007,

109830000, 109835005, 127256003, 187956003, 188038003, 188192002, 188458007, 188734009, 254707006, 275265005, 277570003, 281562007, 307216009, 307340003, 314970000, 359782004, 363407001, 402516002, 422599000, 722688002, 10813100011910, 109969005, 1197359006, 1217617009, 1259312003, 1260024007, 1260046004, 1254725006, 1254745003, 1263469008, 92141004, 94025004, 94197002, 94291005, 94399005, 94409002, 94614009, 94665001, 187683004, 187881004, 188152004, 254423005, 255123005, 314966008, 363471001, 402508006, 402537005, 422758009, 424952003, 440527006, 444596001, 448218008, 448712008, 449224009, 449627008, 707389001, 733360000, 764961009, 783771003, 91081000119109, 353671000119108, 354591000119108, 188538009, 10737601000119103, 1141621000, 685731000119103, 687911000119100, 1186735007, 123691000119104, 1259698009, 1268911008, 1269050001, 1269412004, 93144003, 93909004, 94292003, 94424006, 94530008, 94534004, 94555002, 109978004, 188138001, 254436005, 269544008, 286895009, 363456000, 372119009, 372142002, 402818009, 408648004, 422968005, 423494003, 439478008, 449631002, 707539005, 93531004, 188632001, 10737401000119100, 1080451000119101, 249389151000119103, 353841000119100, 1259426006, 1259714000, 1259717007, 93218008, 93733004, 93768004, 94157000, 94210005, 94418000, 127268007, 187606005, 187736009, 187991006, 188075008, 188095002, 255052006, 312114001, 402494003, 404131003, 721310007, 721569001, 789690008, 860923004, 871843002, 18121000119104, 94291000119103, 117111000119100, 93500006, 188554007, 878805009, 1081631000119101, 683121000119103, 689361000119108, 1254785009, 1259318004, 1269049001, 1279882001, 93637008, 93660000, 93821004, 254980001, 255119002, 326072005, 363435002, 369458006, 369487007, 369547000, 402527000, 423697000, 424151006, 425127006, 707379000, 721642005, 765740002, 770559003, 87111000119109, 99121000119105, 93515007, 686441000119109, 1162855005, 1255784000, 1259320001, 1259352001, 1259421001, 1259682002, 1259690002, 1237460003, 1268395007, 92417009, 93843007, 93959008, 94164003, 94384005, 94447007, 109823006, 109878000, 109885001, 187776004, 254908006, 274084007, 277614006, 278044006, 307226002, 307610008, 369508002, 372115003, 402817004, 707357005, 707394001, 712849003, 136641000119101, 452241000124100, 255115008, 688671000119103, 428487561000119102, 1254845006, 1255895001, 1268519001, 1268707007, 92155007, 93189002, 93697005, 93746009, 93905005, 94018006, 94368009, 94502008, 94601008, 110000005, 187733001, 277616008, 286896005, 363376007, 372000001, 399884001, 403898002, 403926005, 403977003, 427642009, 438946002, 443333004, 449066004, 699358009, 707399006, 721562005, 721632008, 721697000, 722673007, 782672006, 15957141000119109, 352421000119102, 681821000119109, 686981000119108, 1172793008, 1197011004, 1197275006, 1237479000, 60620005, 93934004, 94104001, 94284007, 187949009, 188274004, 254481009, 271467005, 285634003, 363437005, 369493004, 372137005, 396198006, 423032007, 424440001, 447785000, 707409003, 707591005, 722510006, 726652005, 350921000119109, 93487009, 188208002, 404124002, 1081671000119103, 1156804004, 1157157006, 1197259008, 1255905007, 1237484006, 1263914002, 1269051002, 93670003, 93723007, 93867004, 94008001, 94086000, 94214001, 94339008, 94638008, 94669007, 94676002, 187830009, 188770007, 302847003, 373091001, 716588005, 733163007, 836486002, 61291000119103, 354491000119109, 1079551000119100, 1081921000119108, 354461000119102, 354731000119100, 1162361003, 1208741006, 1255747002, 1259505008, 1237517009, 1268487009, 93646002, 93730001, 94105000, 94302007, 94514000, 187662005, 188243001, 188287005, 188505000, 277604002, 372015000, 372018003, 403914000, 403951006, 403955002, 404041003, 441962003, 449307001, 449635006, 1079151000119109, 188586002, 1080751000119108, 1148901006, 16905421000119109, 681711000119105, 1255395009, 1259310006, 1259349009, 1259358002, 1237035007, 92090002, 93948004, 94484009, 94554003, 94605004, 254904008, 254915003, 255015006, 275266006, 363428005, 369610009, 372016004, 707406005, 715901002, 765328000, 830196006, 860831008, 866068003, 93532006, 93550004, 1081851000119108, 1144922006, 1197293007, 1196839006, 1255790001, 1259345003, 93722002, 94026003, 94445004, 94461002, 94477001, 127257007, 187814001, 187996001, 285309005, 314952003, 369598006, 402502007, 449067008, 726019003, 12301000132103, 67831000119107, 122981000119101, 681591000119108, 688091000119106, 1259329000, 1259445002, 1259674002, 1237425009,

93134000, 93210001, 93783009, 93946000, 93964007, 94348003, 127238003, 187757001, 254586002, 254620000, 363359008, 363433009, 402542002, 404114005, 423691004, 424938000, 707580003, 716659002, 722828003, 735918005, 827186009, 1157068008, 1597759100119105, 683981000119101, 1197296004, 1255191007, 1259669005, 1259707002, 1260087003, 1236953005, 1260270005, 1268711001, 92201001, 93708002, 94143002, 94158005, 94160007, 94402006, 94517007, 94659001, 254651007, 254838004, 254850005, 254900004, 254983004, 363470000, 369515005, 369578001, 402517006, 402546004, 449633004, 698288007, 707356001, 723846002, 723851008, 724648008, 118602004, 1156455001, 1156797006, 1179375008, 1255651005, 1259444003, 1259767000, 1237423002, 1251469001, 1251493006, 93137007, 93754006, 93922007, 109874003, 109880006, 188147009, 188744006, 254622008, 277622004, 285312008, 369585002, 423114009, 717734005, 721625004, 78411000119107, 459371000124108, 1082601000112109, 12246641000119104, 681661000119100, 1197307000, 1255800001, 1259443009, 1259592000, 1259772009, 1259806004, 93833009, 94059000, 94224009, 94350006, 94670008, 94680007, 187767006, 188044004, 188077000, 254393004, 269579005, 363372009, 363440005, 363484005, 402523001, 402545000, 719052006, 109390005, 123751000119100, 1153357001, 354331000119104, 1187519007, 1230318000, 1255074005, 1255392007, 1255631006, 1255803004, 1259762006, 1236944003, 1254753006, 13048006, 93140007, 93151007, 93903003, 94338000, 94398002, 94510009, 94630001, 94646009, 254543002, 281566005, 314953008, 369561005, 372141009, 404089007, 430621000, 443261008, 707465007, 714251006, 766980008, 148911000119107, 1010677001, 15950061000119105, 1197355000, 1255740000, 1259440007, 1259740006, 1260048003, 1237052006, 1251498002, 1268721009, 93203007, 93741004, 93766000, 93804008, 187803004, 254466003, 254503007, 277623009, 314954002, 363387004, 369591000, 372009000, 404109006, 431396003, 444910004, 707347009, 707402007, 721628002, 722682001, 12281000132104, 94365007, 1156453008, 687961000119102, 1162348003, 1231381006, 1259627006, 1236945002, 1268697005, 93886007, 93979000, 94053004, 94277004, 94372008, 94401004, 95263006, 187845002, 276870001, 285644001, 307637005, 363370001, 369548005, 404010004, 404153001, 414785000, 423280002, 444374006, 447792005, 448553002, 448609001, 707396004, 707686002, 721762007, 733471003, 866093001, 1079601000119103, 1153423005, 1217692004, 1259260002, 1259613001, 1260050006, 1237046005, 1237489001, 93682007, 93932000, 94177007, 109833003, 188006008, 188366002, 254829001, 277473004, 314967004, 363494000, 371977004, 426964009, 448989001, 449634005, 709031009, 724650000, 773995001, 1082221000119106, 1080731000119102, 1259380004, 1251470000, 1254740008, 1268541001, 93885006, 94023006, 94286009, 94321004, 94373003, 94599006, 109349007, 109858008, 127244004, 127250009, 188183000, 188469005, 188748009, 255003007, 307646004, 369580007, 425749006, 449072004, 707405009, 707479004, 733353006, 783706007, 865954003, 96291000119105, 99131000119108, 1082301000119109, 93498005, 315058005, 372131006, 10737821000119106, 682261000119108, 689801000119101, 1255907004, 1259373001, 1259389003, 1259800005, 1260088008, 1263476003, 1268548007, 1268642000, 1268706003, 93797001, 93808006, 94212002, 94385006, 94431005, 94660006, 94667009, 109923005, 187724003, 187773007, 188045003, 188317004, 254841008, 363394001, 363490009, 369506003, 423987006, 424549003, 426642002, 427658007, 449223003, 707469001, 720587009, 721639004, 722377004, 15636991000119103, 1081861000119105, 1196841007, 1251465007, 93942003, 94289002, 94376006, 94390009, 94715001, 187836003, 188471005, 188493000, 255093003, 363463000, 372004005, 372065009, 404079008, 722542000, 61321000119106, 91041000119104, 895361003, 1148847004, 16527641000119107, 1196844004, 1217383007, 1254848008, 1259507000, 1251467004, 1263786003, 93716006, 93982005, 94109006, 94203004, 94382009, 109886000, 127225006, 187794005, 254535007, 254869000, 254893005, 254912000, 254934003, 276952000, 369513003, 372028007, 404139001, 446807009, 702446006, 184781000119102, 452291000124109, 93506000, 93507009, 307608006, 16218761000119102, 1259351008, 1259655005, 1268717002, 93142004, 93183001, 93753000, 93951006, 94488007, 94543008, 254870004, 277505007, 314995004, 363363001, 404054005, 702786004, 707458004, 708971008, 721604004, 721617001, 721671006, 67811000119102, 1079631000119105, 15958701000119105, 354301000119106,

91271000119101, 1222643003, 1259564005, 93223008, 94355001, 94433008, 94487002, 94505005, 94542003, 187659007, 188273005, 188725004, 254450000, 255038009, 285608006, 363414004, 363445000, 369480009, 404045007, 447951009, 707348004, 707422006, 713293002, 722509001, 830055006, 90811000119100, 130381000119103, 352071000119105, 1080151000119109, 93519001, 105041000119109, 683791000119100, 689311000119105, 1162353008, 1197323000, 1196846002, 1260065007, 1260100002, 1236947005, 1237036008, 1237427001, 1254752001, 1263999008, 93152000, 93704000, 93918002, 94184004, 94576007, 188132000, 188151006, 188298006, 188315007, 254570009, 307599002, 363444001, 363492001, 372011009, 403912001, 420890002, 422378004, 449216004, 698042007, 789004008, 93530003, 1153336009, 1156809009, 1255906008, 1259468006, 1259587002, 1259751006, 1259754003, 1259787004, 1260089000, 1237053001, 1268501005, 1269046008, 93227009, 94040002, 94252000, 94299007, 94617002, 94650002, 95224004, 127240008, 188015001, 188296005, 188633006, 277580004, 399934007, 443493003, 707538002, 723855004, 726654006, 763064007, 1078961000119104, 1255741001, 1259316000, 1259510007, 1259575009, 1237485007, 1237490005, 1268632002, 1268703006, 93659005, 93874009, 94482008, 187692001, 187999008, 275394001, 307219002, 314963000, 369609004, 372063002, 404133000, 404145009, 415112005, 419052002, 423447006, 448561007, 449294002, 707393007, 707576008, 722674001, 733350009, 766759009, 1148551007, 1156471001, 1208461001, 1196898000, 1197354001, 1255394008, 1255650006, 1255751000, 1259371004, 1259791009, 93230002, 93933005, 94038007, 94475009, 109356001, 128466006, 187993009, 188340000, 188669003, 238636003, 278024000, 314999005, 369450004, 369455009, 403909004, 404150003, 449417005, 703230006, 724644005, 1080161000119106, 109971005, 192681000119104, 1080681000119106, 1081911000119101, 1082211000119104, 1162264008, 1255641009, 1255642002, 1260090009, 1237472009, 93143009, 93215006, 93761005, 94057003, 94318001, 94380001, 94651003, 109875002, 109975001, 187722004, 188675007, 271568003, 276815004, 278042005, 285633009, 369549002, 372130007, 424302003, 707430007, 707473003, 860830009, 116691000119101, 351461000119100, 1078941000119103, 1079181000119102, 682411000119106, 1259648008, 1260068009, 1237424008, 1251473003, 35868009, 93683002, 93875005, 93939009, 94333009, 94389000, 94577003, 127218004, 187709007, 233940007, 369594008, 373089009, 419240004, 421246008, 423349005, 443936004, 447782002, 698200003, 707403002, 718604008, 722519007, 722670005, 737308008, 764845008, 187716008, 1153424004, 1197204009, 1237458000, 1237523004, 1237531009, 1251475005, 1264066001, 1268959008, 92817004, 94437009, 94472007, 94593007, 94615005, 109383000, 187777008, 187842004, 188163001, 254582000, 269616004, 286891000, 369535006, 369601001, 402524007, 403458008, 428281000, 448231003, 449176003, 703625002, 707378008, 715950008, 721624000, 722524005, 722795004, 93524003, 1153346006, 1197326008, 682441000119105, 1197349005, 1259436003, 1259456002, 1259548002, 1237530005, 92152005, 93923002, 94010004, 94207003, 94558000, 94575006, 94652005, 303201005, 313249007, 363426009, 369605005, 404130002, 447882007, 699318007, 699657009, 702368000, 707483004, 721574009, 721645007, 722530005, 763771009, 94682004, 277612005, 16636051000119105, 16774551000119100, 350851000119109, 687771000119106, 1196883002, 1255730003, 1259333007, 1259459009, 1259737006, 1259792002, 1260056000, 93737003, 93936002, 94036006, 94061009, 94062002, 94190000, 94421003, 94432003, 94592002, 255118005, 313357008, 372113005, 402514004, 402564006, 413537009, 423106003, 423615009, 425066001, 721304007, 721559007, 764847000, 780844005, 789051005, 838341005, 866096009, 82761000119108, 91161000119101, 1079591000119105, 689211000119102, 1196853006, 1255086008, 1255749004, 1259417008, 1259441006, 1259601006, 1268902008, 92325007, 93759001, 94397007, 94469000, 94507002, 109834009, 188099008, 188736006, 254513004, 255108000, 369453002, 369571007, 372143007, 399068003, 400001003, 403910009, 404016005, 404155008, 416842003, 423464009, 423506005, 425376008, 707400004, 721618006, 722712000, 135931000119106, 353431000119107, 1081811000119107, 188580008, 231835006, 420302007, 686451000119106, 1259383002, 1259596002, 1259681009, 1237525006, 1264499006, 93661001, 93725000, 93984006, 94387003, 187614004, 285614004, 363504005, 369546009, 372021001, 403897007, 404070007, 404129007, 422691006, 445238008, 448607004, 707588005, 722679006,

354361000119107, 459381000124106, 93548007, 118610003, 890529001, 1080501000119104, 1153379006, 1255793004, 1259616009, 1259624004, 1237497008, 1263913008, 1268900000, 93931007, 94509004, 109358000, 285307007, 285618001, 369531002, 369592007, 372001002, 403949007, 422676009, 428322007, 447658000, 447766003, 448214005, 707411007, 707454002, 733358002, 773284000, 1080231000119108, 128875000, 1148906001, 688861000119105, 1197344000, 1259418003, 1240359008, 93703006, 93727008, 93987004, 94125001, 94624001, 187995002, 188054000, 188219004, 188250002, 188642004, 255124004, 269475001, 274088005, 315005000, 363348004, 363491008, 369579009, 423746001, 449420002, 712525007, 713718006, 722718001, 762315004, 816204007, 94144008, 1156459007, 11717361000119108, 15957261000119102, 352771000119107, 91191000119108, 1163568002, 1251565009, 1259347006, 1259541008, 1259664000, 1251483004, 1254689008, 93219000, 93647006, 93929003, 93975006, 127242000, 127261001, 187843009, 188263008, 188361007, 369474001, 369482001, 369530001, 371987000, 372002009, 372120003, 402652009, 403742006, 449267001, 449293008, 705176003, 707354003, 707358000, 707481002, 724866001, 764951002, 770601003, 84811000119107, 101371000119101, 354641000119104, 402912009, 105051000119106, 1079621000119107, 1080741000119106, 1153421007, 16782321000119104, 16905501000119104, 350881000119102, 1197295000, 1197298003, 1259612006, 1260081002, 1236960004, 1268348008, 1268354009, 92150002, 93836001, 94228007, 109911004, 313355000, 369491002, 369529006, 369534005, 416901002, 419842002, 426248008, 722671009, 1080131000119103, 1080191000119104, 118614007, 188551004, 1153427006, 687881000119100, 1254788006, 1259774005, 1259826000, 1268390002, 1268631009, 1269414003, 93792007, 93858004, 93883004, 94329002, 94578008, 95231000, 187786003, 187988006, 188155002, 188745007, 253005002, 254634000, 285310000, 363368005, 363476006, 373080008, 404051002, 413656006, 733347006, 240531002, 1082061000119108, 681831000119107, 91231000119104, 1208451008, 681221000124106, 93184007, 93674007, 93818001, 109951001, 188612002, 314955001, 363438000, 372026006, 372100004, 402547008, 403942003, 420524008, 443250000, 448250003, 448296006, 723848001, 184741000119107, 780821007, 15956261000119100, 682321000119109, 1259342000, 1259525007, 1259688003, 1236961000, 1251458004, 1251482009, 1263601000, 1268561007, 94072004, 94090003, 94238002, 94261000, 94430006, 94710006, 188125009, 363434003, 369475000, 369589008, 369608007, 371012000, 372094002, 402880009, 404110001, 448273006, 448300007, 448868009, 449222008, 707380002, 707384006, 721633003, 733351008, 147131000119101, 352231000119103, 188524002, 308121000, 1079031000119104, 355141000119106, 1162349006, 1255193005, 1260073003, 1236969003, 91858008, 93642000, 93644004, 93769007, 93997008, 94159002, 254549003, 275395000, 277629008, 285308002, 363459007, 369519004, 369581006, 372006007, 372022008, 372098004, 403921000, 444604002, 448665005, 707584007, 707664003, 717968005, 764694005, 353741000119106, 1082701000112100, 15635801000119106, 455733521000119103, 1197310007, 1259416004, 1259516001, 1259578006, 1259593005, 1259795000, 1236940007, 58961005, 93138002, 93226000, 93863000, 94007006, 94165002, 109856007, 187834000, 285613005, 363390005, 369494005, 422736007, 424132000, 721578007, 352301000119103, 458581000124106, 1079161000119106, 1080091000119100, 1080181000119102, 109389001, 1080621000119107, 688691000119102, 1259315001, 1259650000, 1259788009, 1260079004, 1237478008, 1263474000, 1264497008, 93662008, 93968005, 94222008, 94531007, 94589001, 188040008, 188293002, 188445006, 253042009, 277575008, 311779007, 315001009, 363360003, 363483004, 369504000, 399954008, 423607006, 448259002, 448672006, 448994001, 722425009, 733926004, 766757006, 866098005, 116821000119104, 1141627001, 1148852009, 1153383006, 16906601000119108, 687801000119108, 688321000119107, 1255728000, 1259659004, 1259804001, 1259808003, 92818009, 94056007, 94180008, 187846001, 188265001, 188504001, 254557000, 277651000, 312949007, 315004001, 363393007, 369566000, 372123001, 403981003, 423708008, 426370008, 429114002, 448449001, 707495004, 707704007, 709471005, 734884008, 93451002, 109977009, 16699241000119100, 1259338003, 1259348001, 1259553007, 1259666003, 1264502005, 1268906006, 44721000087109, 93204001, 93973004, 94047004, 94060005, 94181007, 94258001, 94653000, 187675005, 187925006, 255102004, 275399006, 302856006, 363380002, 363499005,

707398003, 707589002, 721560002, 723844004, 724056005, 838349007, 352411000119109, 461311000124107, 1081781000119105, 1255110007, 1259533008, 1259676000, 1259686004, 1263651005, 93197009, 93198004, 93756008, 93882009, 94054005, 94198007, 94532000, 94550007, 109976000, 118612006, 127241007, 127248001, 187810005, 187974007, 255111004, 278043000, 285609003, 286902000, 363400004, 363436001, 363457009, 402531006, 408645001, 423973006, 426885008, 702467006, 722518004, 723889003, 733344004, 764856008, 15930861000119100, 188558005, 123741000119102, 1081271000119100, 682471000119103, 686591000119102, 1259620008, 1259827009, 1237469002, 1240362006, 1263642002, 1264364008, 93720005, 93831006, 93884005, 93960003, 93962006, 94044006, 94316002, 94497006, 94608002, 118618005, 187637005, 188153009, 188244007, 188286001, 188295009, 285638000, 285645000, 369481008, 370987005, 404116007, 423645007, 721622001, 762665002, 1671000119100, 91251000119105, 1197297008, 16899901000119104, 1255646004, 1259415000, 1259728006, 1264117000, 1268500006, 1268881002, 93199007, 93851005, 93914000, 93990005, 187760008, 187835004, 188247000, 188489006, 235966007, 254860001, 271323007, 363478007, 423050000, 447757002, 447804006, 448212009, 707404008, 723281005, 726653000, 736322001, 67771000119102, 878806005, 1081311000119100, 15958661000119103, 690121000119109, 93953009, 94173006, 94276008, 94519005, 94566009, 95225003, 187685006, 187968007, 254702000, 276810009, 277461004, 277567002, 369532009, 397008008, 403943008, 449208008, 699028006, 707490009, 707672001, 721709000, 723856003, 133891000119102, 109388009, 188514005, 1081691000119102, 16228971000119104, 1162352003, 1255270006, 1259425005, 1259431008, 1236956002, 1237466009, 92269002, 93724001, 93860002, 93861003, 94313005, 94317006, 94435001, 94492000, 109993000, 187741001, 237833006, 269464000, 363358000, 363395000, 369577006, 369600000, 403919005, 432328008, 443439001, 447109003, 448139005, 722529000, 737313007, 830060005, 93546006, 93554008, 1080591000119105, 352911000119101, 763101411000119100, 1259377000, 1259739009, 1259815006, 62497000, 93839008, 93955002, 94128004, 94661005, 109347009, 187917009, 187970003, 188049009, 188189001, 188239000, 188312005, 188665009, 254612002, 314974009, 363415003, 369509005, 372024009, 443937008, 707344002, 737058005, 866109006, 91421000119103, 354681000119109, 188536008, 307634003, 1078891000119104, 1081301000119103, 1153381008, 681941000119107, 685601000119104, 793197981000119106, 1259598001, 1259651001, 1260093006, 93879004, 94045007, 94055006, 94481001, 94596004, 94643001, 95261008, 127258002, 127273001, 188204000, 253018005, 285637005, 286890004, 363465007, 369466002, 369569007, 369593002, 399660006, 403950007, 423896007, 424334007, 426124006, 447805007, 708054009, 716655008, 33581000119109, 234941000119100, 242951000119108, 93526001, 188582000, 1156788007, 687941000119101, 688911000119101, 1259407003, 1259483001, 94066004, 94076001, 94606003, 188190005, 188268004, 188326001, 254619006, 286889008, 369489005, 371974006, 385478001, 403274000, 403711001, 405822008, 428061005, 706970001, 709285002, 721619003, 724059003, 1082101000119106, 188548006, 404123008, 1144764001, 353211000119103, 1222581000, 1255883001, 1259309001, 1259328008, 1259354000, 1259765008, 1264369003, 1268394006, 91861009, 93649009, 93698000, 93864006, 94240007, 94490008, 94583000, 109912006, 254972008, 363425008, 369456005, 369541004, 403713003, 413587002, 418529003, 443719001, 448213004, 448663003, 700057001, 866082003, 1691000119104, 432131000124100, 12241071000119106, 15950101000119108, 16260431000119104, 688901000119104, 1255662005, 1259559006, 1259709004, 1259781003, 1259828004, 1251496003, 1263639008, 92203003, 93224002, 93924008, 94037002, 94260004, 94707004, 187957007, 188107002, 188211001, 188241004, 188728002, 254417000, 369570008, 404092006, 707427000, 707475005, 707532001, 718200007, 833295000, 10737911000119105, 15931061000119103, 354721000119103, 689481000119107, 1255886009, 1259404005, 1259599009, 93848003, 94308006, 109854005, 109855006, 188052001, 255084004, 269476000, 278189009, 307622006, 369523007, 404040002, 404090003, 404152006, 422886007, 707455001, 707585008, 866097000, 12235601000119105, 118606001, 1079571000119109, 1137644008, 1144305001, 353761000119105, 687991000119109, 1197325007, 1259346002, 1259461000, 1268543003, 1284849005, 94080006, 94167005, 94262007, 94448002, 369553000, 371997009,

372064008, 372125008, 448257000, 449207003, 698285005, 721305008, 724537009, 763796007, 12291000132102, 354741000119109, 92512000, 118609008, 1010631005, 1153356005, 1156420003, 16260391000119109, 1255748007, 1259379002, 1259393009, 1259495001, 1259557008, 1260062005, 92299006, 93731002, 127235000, 188019007, 188021002, 254389005, 277664004, 363439008, 372092003, 403468003, 448408001, 698286006, 721630000, 866080006, 87091000119101, 91031000119108, 122901000119109, 1082321000119100, 403947009, 16905381000119100, 681681000119109, 808558991000119105, 1259504007, 1259832005, 1254748001, 93827000, 93887003, 94085001, 110002002, 127254000, 188070003, 254454009, 254950006, 277474005, 307623001, 363474009, 402559007, 449295001, 722714004, 107751000119102, 1079561000119103, 1099581000119102, 352341000119101, 1196860000, 1259470002, 1259710009, 1260038000, 1237031003, 1268353003, 93732009, 93748005, 93802007, 93832004, 94226006, 94232001, 94413009, 94585007, 94634005, 95210003, 109838007, 109948008, 127247006, 187769009, 187822008, 187939006, 187975008, 188068007, 277624003, 277628000, 363485006, 370967009, 402560002, 449058008, 353501000119104, 352471000119101, 1255659007, 1259324005, 1259386005, 1259712001, 1259743008, 1259778008, 1237493007, 94288005, 94459006, 128465005, 187725002, 187967002, 188691005, 254478004, 363507003, 399897004, 403896003, 403944002, 404009009, 405843009, 721575005, 721725005, 735450006, 763063001, 84831000119102, 253017000, 350621000119100, 1255737000, 1255815000, 1259487000, 1259544000, 1260036001, 1236963002, 1237038009, 1264495000, 1268635000, 92813000, 93835002, 93870000, 94374009, 118615008, 127265005, 187723009, 187938003, 254526000, 254709009, 275490009, 285432005, 307609003, 307649006, 363346000, 363389001, 372012002, 372129002, 402528005, 402815007, 416769008, 423189008, 423274005, 423384009, 423600008, 447416008, 707425008, 707671008, 717916003, 721308005, 722229001, 1156811000, 16414611000119100, 16845381000119100, 1162280003, 1259641002, 1259752004, 1260066008, 1260069001, 1236958001, 1237494001, 1237496004, 1254746002, 93717002, 93728003, 94004004, 94269003, 187701005, 188184006, 188322004, 310599006, 363409003, 371970002, 371976008, 400173004, 423700001, 427141003, 449218003, 721603005, 734066005, 783736003, 866071006, 866095008, 72891000119103, 91281000119103, 130911000119108, 1081551000119106, 1153382001, 681911000119108, 685971000119103, 1162284007, 1255752007, 1259335000, 1259413007, 1237042007, 1237372005, 94071006, 94078000, 94293008, 94303002, 94428009, 109915008, 187634003, 253096008, 254849005, 276738009, 278052009, 302815008, 314991008, 363379000, 369526004, 371980003, 402816008, 408649007, 423038006, 425148008, 447784001, 699818003, 707410008, 707456000, 707482009, 721546004, 721634009, 722681008, 350861000119106, 277654008, 685861000119102, 1187285005, 1217010007, 1237368006, 1255739002, 1259311005, 1259571000, 1264257008, 1264524005, 93672006, 93910009, 94138009, 94254004, 94309003, 109824000, 127066000, 254896002, 285776004, 372093008, 427492003, 448664009, 448776002, 449100005, 707586009, 721638007, 733135005, 122881000119107, 354851000119101, 93488004, 1153355009, 352431000119104, 682221000119103, 685921000119104, 687401000119105, 1255807003, 1259635009, 1259757005, 1254724005, 1268542008, 93734005, 93799003, 93800004, 94300004, 94315003, 94536002, 95192000, 127243005, 188230001, 231832009, 307650006, 402533009, 402536001, 404053004, 404148006, 422399001, 424632001, 444597005, 446022000, 707529004, 767448007, 82501000119102, 1079131000119103, 93501005, 895359007, 1145245004, 15955101000119101, 688711000119104, 1255194004, 1259323004, 1259385009, 1259464008, 1260406003, 1264501003, 93188005, 93643005, 93980002, 94644007, 94714002, 109937001, 109982002, 127239006, 188176007, 254427006, 254653005, 255088001, 285604008, 285839005, 369457001, 369520005, 369543001, 402496001, 402879006, 404088004, 723265000, 733361001, 737309000, 761958009, 766981007, 838340006, 350951000119101, 92513005, 93551000, 285420006, 1080631000119105, 1197350005, 1259331009, 10737731000119102, 1237055008, 1263640005, 1264358001, 93696001, 94048009, 94357009, 94375005, 94383004, 94546000, 94552004, 94568005, 109357005, 187828007, 253000007, 254633006, 255035007, 372030009, 393573009, 402636006, 403986008, 404142007, 424970000, 443961001, 707535004, 766935007, 782722002, 188592008, 404106004,

16091131000119108, 1255643007, 1259718002, 1236966005, 1240370001, 93676009, 93786001, 94021008, 94134006, 94320003, 94434002, 94439007, 109841003, 109988003, 187732006, 255073006, 285619009, 363392002, 372062007, 722516000, 782827000, 91131000119109, 226521000119108, 354651000119102, 16091251000119109, 551001000124108, 1208459005, 1259319007, 1268885006, 94096009, 94234000, 94547009, 187987001, 190030009, 254459004, 254988008, 255016007, 255110003, 274905008, 276826005, 276828006, 278051002, 285643007, 310504009, 423807009, 440397000, 449253005, 721606002, 735757008, 737310005, 766930002, 450521000124109, 1172704005, 1259463002, 1259618005, 1259746000, 1259831003, 1260035002, 1254726007, 1268636004, 1268899005, 25050002, 93211002, 93738008, 93807001, 93824007, 94272005, 94361003, 94504009, 94524008, 94712003, 188157005, 188253000, 254484001, 254517003, 276420005, 363427000, 363467004, 371999007, 372095001, 402519009, 404126000, 721303001, 184871000119108, 683991000119103, 1081711000119104, 188567005, 285422003, 10737361000119109, 1156413005, 1259432001, 1259555000, 1259764007, 1237516000, 1264496004, 93743001, 93764002, 93907002, 94101009, 94229004, 94358004, 94533005, 187946002, 254530002, 269463006, 277642008, 363430007, 415177008, 424954002, 448995000, 449472007, 707386008, 721695008, 722832009, 738527001, 133851000119107, 352251000119109, 1080171000119100, 10737641000119101, 1080611000119100, 688781000119105, 1255092002, 1259730008, 1259741005, 1259771002, 1259818008, 1264067005, 1268547002, 1268714009, 92333008, 94451009, 109941002, 188339002, 254916002, 276803003, 369587005, 399490008, 404151004, 448988009, 702405001, 707345001, 707494000, 716647001, 735920008, 1661000119106, 184881000119106, 188578002, 16906521000119103, 687811000119106, 1186629001, 1255797003, 1259573002, 1259629009, 1259833000, 1268458008, 82546001, 91857003, 93641007, 94427004, 94611001, 94683009, 254390001, 254509006, 276797002, 312113007, 314418005, 402543007, 403904009, 423425006, 449059000, 722112006, 860828007, 1144306000, 688941000119102, 1255894002, 1259657002, 1237056009, 1237416003, 1237482005, 1268700009, 93680004, 93721009, 93844001, 94166001, 94325008, 94392001, 109980005, 254553001, 254994000, 363422006, 369473007, 369586001, 404134006, 415284008, 423318000, 447708000, 448670003, 702977001, 192701000119101, 878807001, 1080001000119106, 1156415003, 15950021000119100, 15957221000119107, 15958261000119101, 16905341000119105, 1259539007, 1260034003, 1251454002, 1268528000, 1268719004, 1269347008, 93706003, 93739000, 93967000, 94335002, 94354002, 94455000, 94538001, 109851002, 134312002, 188046002, 188240003, 254839007, 363424007, 402501000, 404107008, 415285009, 445227008, 447949005, 721313009, 733343005, 783006001, 865951006, 100731000119107, 12246601000119101, 93549004, 109391009, 276876007, 1079541000119102, 1079991000119101, 1081731000119109, 1179762006, 1197286008, 1208457007, 1255814001, 1255899007, 1259411009, 1259546003, 1259639003, 1259766009, 1268960003, 93136003, 93904009, 93976007, 94237007, 109943004, 188266000, 188645002, 254800003, 269578002, 277782009, 285606005, 285612000, 285617006, 363401000, 369517002, 369552005, 371998004, 372110008, 425303004, 707362006, 707537007, 707670009, 707697002, 716274007, 354621000119105, 93528000, 255121007, 1187521002, 1197258000, 1254800007, 1259317009, 1259405006, 1264521002, 1269048009, 93150008, 93829002, 93926005, 94440009, 94673005, 109984001, 109995007, 188069004, 188264002, 255091001, 277653002, 404047004, 404664002, 425225007, 29421000119105, 93537000, 1080601000119103, 1254789003, 1255902005, 1259339006, 1259398000, 1259549005, 1259784006, 1263479005, 93773005, 94126000, 94183005, 94227002, 94332004, 94406009, 254996003, 276827001, 303057009, 359780007, 363460002, 369449004, 403923002, 404073009, 423595004, 447389009, 699355007, 707401000, 713573006, 722684000, 737311009, 763409006, 840573001, 459421000124101, 188559002, 1010633008, 1157070004, 1157072007, 1255084006, 1259355004, 92812005, 93669004, 94063007, 94202009, 94523002, 94622002, 109918005, 188032002, 188285002, 254711000, 255067005, 255101006, 276750003, 278060005, 363432004, 363468009, 363473003, 369459003, 402534003, 404112009, 423627007, 702369008, 722533007, 732201008, 860832001, 454721000124105, 15930901000119106, 15930941000119108, 354421000119107, 682271000119102, 686991000119106, 1197312004, 1254846007, 1255738005, 1255744009,

1259610003, 1259713006, 1259793007, 1259807008, 93205000, 93779009, 93950007, 93992002, 94405008, 94664002, 127253006, 127263003, 187644001, 187943005, 369451000, 369477008, 371963002, 371981004, 389216001, 398623004, 402512000, 444911000, 698041000, 713646001, 716586009, 722528008, 13487100011910O, 1079091000119100, 1081601000119108, 1082251000119103, 15956221000119105, 16845301000119108, 682421000119104, 685691000119105, 91221000119102, 1255897009, 1259526008, 1259631000, 1259646007, 1259824002, 94256002, 94349006, 94636007, 109384006, 127260000, 188676008, 188732008, 232075002, 254989000, 286893002, 369484000, 369599003, 369607002, 371994002, 372104008, 400057007, 403996004, 448258005, 733354000, 12235561000119105, 285421005, 686601000119109, 1255729008, 1259725009, 1236967001, 1237621006, 1269125001, 92032007, 93841009, 94282006, 94471000, 94516003, 94642006, 109822001, 188768003, 277597005, 278055006, 363364007, 372122006, 403900000, 404077005, 413847001, 721701002, 866048009, 92516002, 1148931000, 1156454002, 350801000119105, 688381000119106, 84821000119100, 1197292002, 1255805006, 1259430009, 1240372009, 1268531004

#### 3.21 Obesity

ICD10CM:
Z68.43, Z68.38, E66.8, E66.09, Z68.34, Z68.32, Z68.31, Z68.33, Z68.42, Z68.45, E66.3, E66.9, Z68.39, Z68.30, Z68.36, E66.2, Z68.41, Z68.37, E66.1, E66.01, Z68.35, Z68.44

SNOMED CT:
722037004, 1187531009, 294493008, 717761005, 15750121000119108, 360566006, 700150001, 72894001, 408512008, 190966007, 5036006, 295509007, 770680004, 414917005, 270486005, 785722006, 415530009, 414919008, 444862003, 719160009, 296526005, 238135003, 715628009, 238131007, 293481008, 238132000, 10750551000119100, 111036000, 443381000124105, 248312008, 248311001, 80660001, 275947003, 1229946007, 788996008, 162690006, 1208987006, 171000119107, 722595002, 62999006, 238133005, 783556000, 44772007, 1260134001, 238134004, 414918000, 297500005, 292464007, 1260139006, 162863004, 162864005, 238136002, 1229943004, 268915006, 414915002, 721231007, 1076711000119101, 83911000119104, 461341000124106, 268916007, 1255335006, 290439001, 57337005, 776204008, 722053001, 702949005, 414438005, 774102003, 783549006, 1003380001, 722596001, 819948005, 298464002, 53146006, 770750002, 450451007, 414916001, 722051004, 414920002, 724137002, 763350002, 773663004, 719834005, 1076701000119104, 783719006, 82793005, 63702009, 717269008, 443371000124107, 190965006

#### 3.22 Other Neurological Disorders

ICD10CM:
G12.0, G13.1, G20.B2, G11.0, G40.00, G40.B0, G13, G12, G40.4, G37.1, G31.2, G32, G25.5, G40.10, G40.31, G40.C, G40.C0, G40.821, G93.6, G31.9, G37.0, G40.824, R56.01, G36.1, G11.6, G12.24, G40.90, G40.21, G40.C19, G40.A09, G40.B09, G40.C01, G40.C11, G13.2, G21.9, G40.919, G40.802, G40.11, G11.8, G11.9, G93.1, G11.10, G13.8, G40.89, G20.B1, G12.23, G21.2, G40.50, G40.80, G40.401, G93.0, R47.02, R56.1, G12.9, G21.3, G40.A11, G21.8, G40.209, G40.311, G40.A0, G12.2, G93.5, G20.C, G93.82, G11.11, G40.109, G40.219, R56, G12.25, G40.011, G40.5, G40.101, G40.211, G93.49, G20.A2, R56.00, G13.0, G36.8, G40.409, G36.0, G93.41, G93.40, G21, G40.201, G40.A01, G32.0, G21.11, G40.0, G40.301, G93.7, G12.20, G40.82, G32.89, G40.41, G40.814, G40.01, G40.812, G93.3, G37.4, G40.901, G40.B, G36.9, G40.91, G37.5, G40.419, G40.8, G40.83, G40.511, G31.81, G93.2, G12.22, G20.A1, G40.519, G40.C1, G40.009, G40.909, G31.8, G40.822, G25.4, G40.B1, G93.8, G11.19, G40.B01, G93.89, G37, G93.9, G37.3, G93.32, G20, G40.A, G31.86, R47.0,

G12.1, G40.81, G40.111, G37.81, G40.30, G40.823, G11.2, G21.19, G31.83, G40.001, G40.A1, G37.2, G37.8, G40.119, G40.319, G93.39, G40.804, G40.809, G40.811, G31.80, G40.509, G37.89, G21.4, G93.81, G21.0, G40.801, G32.81, G40.20, G40.B11, G93.44, G40.309, G93.4, G12.21, G93.42, G93.43, G31.84, G40.42, G40.833, G40.501, G40.911, R56.0, G20.B, G40.813, G12.29, G31.89, R47.01, G40.1, G11, G40.3, G32.8, G40.019, G36, G11.4, G11.1, G40.9, G93, G40.C09, G40.40, G21.1, G31.85, G37.9, G40.B19, G12.8, G35, G40.411, G40, G40.A19, G20.A, G40.834, G10, G93.31, G11.3, G31.82, G40.803, G40.819, R56.9, G11.5, G40.2

ICD9CM:
345.61, 345.80, 335.8, 335.0, 340, 335.20, 345.3, 784.3, 335.22, 345.7, 334.3, 331.9, 345.91, 335.19, 335.21, 335.23, 345.10, 341.8, 333.4, 335.1, 345.8, 348.1, 335.24, 780.33, 333.5, 345, 334.1, 780.3, 336.2, 335.9, 345.01, 341.1, 334.9, 345.40, 345.81, 332.1, 345.71, 335.11, 345.70, 332.0, 341.20, 335.10, 334.8, 334.2, 345.51, 348.31, 334, 335.2, 341.22, 345.0, 345.00, 341.9, 345.50, 345.5, 345.1, 335.29, 345.60, 780.31, 345.90, 341, 345.2, 345.11, 348.30, 345.6, 345.9, 334.0, 780.32, 341.2, 341.0, 348.3, 780.39, 334.4, 345.41, 341.21, 348.39, 333.92, 335, 345.4

SNOMED CT:
3119002, 4113009, 4439001, 17409003, 31033002, 34427002, 35486000, 38583007, 40781006, 47774001, 72839009, 79801002, 81642009, 83351003, 94069006, 111499002, 163932009, 164094002, 192716003, 193028008, 194843003, 196474009, 200063000, 203943005, 203976009, 212316005, 213207003, 230294009, 230525002, 231450007, 237700006, 249440006, 254256008, 267977009, 276584001, 424669003, 424761009, 699261008, 711524008, 722614006, 724351008, 764812008, 840464007, 87151000119105, 292441000119109, 330041000119103, 341441000119102, 346911000119103, 350471000119102, 431741000124101, 460731000124105, 15639321000119106, 16206821000119104, 1073003, 230301006, 276221006, 425887005, 335471000119108, 16065251000119109, 352751000119103, 1163102005, 1179296003, 1169356004, 1222657001, 20930002, 23096007, 32931001, 36209000, 45894003, 51323005, 54287007, 66393002, 69757000, 83367009, 91377003, 94787001, 126951006, 126952004, 126956001, 128208007, 189179009, 192730008, 209095008, 230210001, 230515007, 230671004, 230800004, 231996009, 236692005, 237682009, 253128003, 253178001, 253746001, 254177003, 267389001, 271479005, 277507004, 421437000, 425936006, 441705005, 444545003, 609554006, 698743007, 699314009, 705130002, 716771000, 717043006, 721378006, 721975004, 725011006, 766246000, 782675008, 783157004, 288851000119106, 341691000119108, 435091000124105, 1085091000119108, 10943311000119108, 12236911000119107, 15697281000119100, 15738481000119102, 16039431000119105, 123535000, 890433006, 1176992006, 1231751000, 1237571004, 1255640005, 1268955002, 5582005, 7126001, 7819003, 15277004, 40130009, 53096005, 56267009, 76157009, 78693004, 89451009, 90860001, 94450005, 94600009, 188282004, 189016009, 191493005, 192704009, 193165008, 198993009, 201625003, 202674000, 202685003, 230401007, 230471006, 239852007, 239884000, 246783003, 267388009, 277189006, 363497007, 400018004, 404235004, 404654006, 406566006, 417484006, 420932006, 422513000, 717761005, 722110003, 722914008, 735462005, 735614005, 762330008, 816205008, 840576009, 137991000119103, 330791000119108, 343721000119103, 433891000124100, 444441000124109, 16058111000119103, 16207211000119105, 59032001, 445308004, 16903531000119107, 1156412000, 38595071000119104, 1173997008, 1217226000, 1230142006, 1231632004, 1260322001, 1260378000, 18311006, 29197009, 40099009, 45369008, 67305007, 70576008, 75983009, 81830002, 85821003, 91951001, 111487009, 123707001, 192708007, 201847001, 230193008, 230506000, 230643005, 230674007, 234199000, 236531005, 238701007, 238945000, 238947008, 262718005, 267611002, 307686007, 312923002, 314010006, 416633008, 431034009, 432354000, 442925003, 609382000, 698781002, 700331009, 711566009, 719819004, 721404006, 721513005, 721522006, 722620007, 723584003, 732933009, 733636005, 763312008, 763366000, 771469002, 788978004, 860852000, 5601000124105,

291711000119109, 293941000119105, 433941000124104, 1074091000119101, 1074451000119102, 1156764005, 1156798001, 1169360001, 1186825009, 1177042002, 1208339007, 1259196009, 1259609008, 1263597000, 1281842000, 5566009, 15758002, 19514005, 23374007, 27358001, 37356005, 42344001, 55967005, 63246000, 69134001, 71587006, 76236006, 84306007, 93930008, 94294002, 111568001, 123953004, 126971002, 188462001, 191306005, 201066002, 230158001, 230281007, 230679002, 230715005, 232439007, 253144007, 254970000, 288276001, 297946004, 396275006, 403646002, 414173003, 416073003, 445095002, 608874000, 709012003, 715751004, 715776003, 716664003, 718213001, 719316004, 721261004, 721477000, 722055008, 722292000, 724425005, 724560001, 733071009, 762707000, 764850002, 5321000124109, 140961000119107, 678691000119106, 15678801000119102, 1003447007, 890368007, 1156858008, 1186881003, 1162852008, 1220600004, 1260223009, 12204031000119101, 1260370007, 1263555006, 1263577005, 1269375005, 1284851009, 7343008, 7454002, 22419002, 24956004, 36540006, 40631009, 53633000, 56155002, 59770006, 70936005, 78358001, 83832001, 94266005, 95459002, 95484004, 192711008, 193071004, 204138006, 230206004, 230367007, 230557001, 230754006, 230786001, 231518004, 231907006, 238518008, 238699007, 238982009, 240468001, 241991002, 253523001, 254189001, 282806000, 314007004, 315048006, 359569009, 388979002, 399971009, 430399004, 442421004, 702463005, 703621006, 708673009, 709145001, 715655000, 721220004, 722929005, 723716009, 724722007, 734022008, 763280005, 765046002, 766767001, 770667002, 773555005, 140921000119102, 347271000119100, 368311000119105, 12239181000119108, 15705441000119107, 15738281000119101, 16024191000119100, 88125005, 1003432003, 1003584006, 720261501000119107, 1172688004, 1230020008, 1259721000, 1263546008, 2463005, 5587004, 15802004, 22067002, 40787005, 49736003, 53936005, 70673006, 79468000, 126962006, 188321006, 190330002, 191458003, 203944004, 208060001, 209077005, 210038008, 212267006, 212288002, 213056007, 230197009, 230386004, 230452009, 240058001, 240472002, 246535004, 247147005, 265377002, 267639001, 285641009, 304603007, 312947009, 427065003, 450363003, 703369003, 721385005, 721502007, 721766005, 723157009, 733086003, 762328006, 766872002, 771186004, 771420004, 788907005, 830015009, 107311000119104, 291851000119100, 292021000119105, 292421000119103, 313831000119100, 330311000119100, 367601000119103, 1084701000119100, 82361000119107, 1176984002, 1208841001, 1208629005, 1259122001, 1260224003, 1279978002, 11256005, 16171003, 23732000, 25859000, 64009001, 67930001, 95004009, 110284009, 111385000, 127303003, 163988000, 188289008, 188319001, 198992004, 203967002, 206234004, 230391003, 240211000, 253147000, 253180007, 254957009, 402468007, 403403005, 425500002, 472916000, 698848005, 707443007, 719678003, 720010009, 721729004, 721974000, 722631004, 722992006, 723582004, 735235000, 735752002, 766815007, 773692000, 866132005, 870265003, 111641000119102, 134771000119108, 335861000119104, 338691000119104, 348071000119106, 13015781000119100, 16026871000119101, 28978003, 1010666007, 897311008, 1187564009, 1163131009, 1163132002, 1259117001, 1260130005, 1260384002, 1263585001, 1281836004, 330651000119107, 39181000087106, 5112009, 11659006, 13274008, 20075001, 25816005, 28676002, 52421005, 53741008, 56097005, 70884002, 86695008, 87764000, 87909002, 88233000, 89476005, 89478006, 91175000, 95148002, 95453001, 191475009, 193102008, 203974007, 229747003, 230420009, 230568001, 230756008, 238852002, 254965007, 277373000, 284186002, 312957005, 371115001, 442693003, 449904002, 699814001, 700312003, 708975004, 718752007, 721408009, 721770002, 735648006, 762300000, 771334000, 782945001, 838332000, 860811009, 21841000119100, 68131000119100, 100941000119100, 331381000119100, 342911000119104, 347021000119109, 432161000124109, 434451000124105, 12236681000119108, 16006551000119104, 16899261000119106, 231930000, 1003373003, 1141628006, 1197747005, 1231283007, 1254941001, 1260334009, 1281837008, 16637211000119100, 5318001, 18689007, 22126005, 40912001, 50143004, 61974008, 64036004, 70843000, 76902006, 82725007, 87450004, 91187007, 93744007, 95126008, 111498005, 128188000, 188308004, 193120004, 201117001, 206457007, 212198000, 230467008, 230654000, 230655004, 230710000, 235039005, 238020001, 240565001, 251001002, 254982009, 271723009, 276587008, 278466007, 398100001, 410472006, 703542000, 709587006,

715748006, 721088003, 721222007, 721452001, 724727001, 725433003, 762294003, 763721006, 763794005, 766931003, 773492007, 778005007, 782881002, 783739005, 105451000119102, 149821000119103, 291081000119100, 293961000119109, 295501000119104, 15685081000119102, 15738401000119105, 15739881000119108, 15863451000119107, 786120041000132108, 29570005, 44145005, 52859009, 238844000, 406582007, 421529006, 1052319006, 1075761000119107, 1144936001, 1156853004, 806161651000119106, 1186877003, 1196872006, 1237625002, 105171000220101, 1259591007, 1268701008, 33209009, 39823006, 44395000, 44983007, 52072009, 83716008, 88281007, 111515007, 111872008, 186284005, 191463004, 192899004, 204067005, 208063004, 209100005, 230202002, 230682007, 387712008, 410074002, 441980007, 445355009, 699184009, 717977003, 721509003, 722854006, 723127000, 732954002, 733192006, 762535004, 782690007, 782737003, 788916009, 838345001, 13601000119109, 28721000119108, 87121000119102, 306171000119102, 680531000119103, 15731561000119100, 16058191000119107, 25410001000004103, 6475002, 417607009, 420244003, 438513002, 684931000119100, 1177010008, 1259744002, 1259471003, 9859006, 11045000, 48522003, 62158001, 75076004, 76628006, 91012008, 128204009, 171822009, 172276003, 202679005, 212229008, 229692002, 230567006, 230619007, 231925004, 232449005, 237727005, 239839002, 240053005, 240393003, 254151006, 254938000, 277527003, 282800006, 403650009, 406592004, 410568009, 416463008, 421506003, 441574008, 442300000, 442884002, 698999002, 721788005, 722618009, 722660008, 722962002, 724002003, 762363001, 764854006, 770560008, 773307006, 5771000119106, 291481000119105, 292851000119109, 294071000119100, 333231000119107, 333531000119105, 438411000124101, 15686121000119109, 15738441000119107, 15960781000119108, 15986271000119103, 49455004, 76880004, 15704241000119107, 10954781000119103, 1156416002, 1156460002, 1179377000, 1172594000, 10861091000119102, 334371000119100, 1259819000, 1263490000, 4523006, 7449006, 34977006, 37145001, 45781009, 61701006, 69131009, 191483003, 192788009, 193463006, 195025008, 209182000, 212169004, 212243003, 230395007, 230596007, 230599000, 231837003, 240073000, 247157006, 282748000, 285757001, 402172000, 403397004, 403790004, 439732004, 443002008, 702448007, 712644005, 716592003, 718519003, 721441007, 721493009, 721526009, 721765009, 722382006, 733630004, 763632004, 774212003, 782829002, 866121005, 17771000119103, 77461000119109, 124171000119105, 306571000119106, 311211000119105, 330421000119102, 338701000119104, 15630201000119101, 15633051000119102, 15989271000119107, 16005951000119104, 230380005, 735421004, 1141882007, 1145215002, 1187042007, 1217651008, 1208937004, 1234783007, 1259654009, 18822004, 18960007, 27206009, 31382008, 45283008, 50429003, 54184008, 56531003, 61337004, 66454007, 69718008, 71760005, 73453007, 78875003, 83157008, 86393005, 91502009, 95834000, 109940001, 111298007, 193074007, 193186008, 201070005, 201929006, 202830000, 206196005, 230298007, 230330004, 230545005, 230687001, 230709005, 230782004, 231949001, 253162003, 297943007, 311829000, 314467007, 417017003, 419990005, 428876001, 445331004, 608817003, 702377007, 711483003, 711567000, 717061002, 719156006, 720813007, 722984002, 723502001, 724836008, 737237009, 783012006, 785298001, 788863007, 816067005, 2961000119106, 34041000119108, 292351000119100, 294061000119106, 303161000119101, 331981000119104, 679231000119101, 15987391000119105, 14493003, 66264000, 16891191000119108, 1163064006, 10861131000119100, 1220650006, 1237130004, 8011004, 15671007, 37754005, 47000000, 51742006, 52723009, 59753003, 62195001, 65312002, 70917000, 70932007, 85102008, 102448004, 191464005, 192667007, 193191009, 200065007, 212210002, 230236008, 230253001, 230352008, 230622009, 237577005, 237809000, 241993004, 246530009, 254969001, 276650005, 442928001, 442964000, 609558009, 715465001, 718096004, 719216001, 720519003, 721499008, 722033000, 724384008, 763070001, 870563006, 135811000119107, 292601000119109, 432011000124107, 461441000124103, 12265501000119101, 16016551000119101, 16238741000119105, 238937005, 768663003, 1052243006, 1156840002, 769023031000119104, 1173024006, 1228881003, 15675921000119107, 1260143005, 1263983002, 1281834001, 1207009, 1686006, 5217008, 16110005, 16695002, 30877004, 38523005, 39150004, 46252003, 57917004, 61663001, 67320001, 77274005, 78211006, 90310002, 94969009, 94970005, 95695004, 109905002, 126975006, 126983000,

128207002, 128239009, 163933004, 186628001, 193152004, 208078006, 212247002, 212276004, 230755007, 235047005, 237877004, 238022009, 238821003, 238839007, 240552005, 254951005, 262699006, 271386001, 271971004, 276219001, 402856005, 425007008, 445014002, 451035002, 609395005, 699867001, 703280000, 713829001, 732923001, 782822006, 783091003, 860881004, 870290001, 870339004, 32651000119109, 290211000119105, 290531000119102, 290931000119108, 291721000119102, 313841000119109, 330061000119104, 340311000119104, 672571000119109, 679181000119107, 1621920100011910 1, 10532003, 56048001, 1119281000, 1196911006, 1281840008, 25331007, 61104008, 74225001, 75853001, 78723001, 85136002, 109561000, 129614001, 163593005, 195200006, 199096006, 203522001, 209086000, 212141007, 212301007, 230198004, 230799003, 231926003, 237728000, 246546001, 402340009, 403256009, 403687001, 403808006, 405756005, 416351002, 421966007, 428199000, 441806004, 609592007, 700150001, 717733004, 720954000, 733332002, 762507003, 768553002, 840459008, 860822008, 82371000119101, 125921000119106, 290111000119100, 290461000119109, 345671000119105, 347531000119100, 15737201000119108, 26852004, 41455002, 84727000, 430404002, 344271000119107, 1172603005, 1186874005, 1220589007, 1197182004, 1254939002, 1259476008, 1259542001, 8627007, 17689009, 29426003, 48528004, 49453006, 49481000, 51371005, 72880002, 95775000, 109926002, 109938006, 110998005, 126964007, 190856003, 192990004, 192999003, 193881001, 194118007, 202747007, 230156002, 230199007, 230759001, 238834002, 253223002, 254958004, 289273003, 312925009, 402717008, 406446000, 418856006, 442212003, 450379002, 460880006, 609614000, 698738000, 698763000, 721464009, 722574007, 723170000, 735556002, 870347004, 68861000119103, 90831000119105, 291371000119100, 291841000119102, 338851000119105, 348151000119103, 440211000124100, 679121000119108, 1666193100011910 2, 192656006, 425390006, 3549009, 8847002, 11387009, 14309005, 15708009, 15771004, 17111003, 58691003, 92122002, 95657009, 111338006, 125609005, 127012008, 129623003, 188281006, 202690000, 206192007, 208006009, 212281008, 212357003, 230711001, 232002000, 249944006, 253165001, 276817007, 360418002, 405757001, 416714005, 424736006, 427758000, 438511000, 711103007, 715564000, 715726000, 716787002, 718766002, 735755000, 771081007, 771143004, 77471000119103, 124001000119104, 292131000119109, 1086991000119103, 10972981000119108, 15637511000119100, 16052551000119108, 16077211000119109, 16477021000119103, 204739008, 230267005, 230388003, 1003881009, 1163106008, 1187566006, 10941451000119104, 1259540009, 1269374009, 442001, 5822000, 6208003, 18071005, 30243004, 43262000, 50366005, 87227007, 92474007, 94247005, 94789003, 95894002, 189162003, 192898007, 192991000, 208058003, 230142004, 230183001, 230315008, 230628008, 235040007, 237349002, 253010003, 276280009, 276591003, 277185000, 395505000, 397573005, 399091004, 399107008, 399305009, 402997002, 419395007, 422446008, 428212003, 713702000, 715826005, 721769003, 723308003, 724678004, 724708007, 762510005, 770792007, 783090002, 840421000, 4061000119104, 124081000119107, 290681000119102, 290921000119105, 294041000119107, 306551000119102, 318671000119108, 337401000119107, 430051000124107, 15639361000119101, 15729961000119108, 15989831000119105, 16026591000119103, 28140001000004102, 417065002, 422012004, 1003923009, 1142050005, 1144906005, 1156967007, 1157161000, 1172634009, 1187540008, 1187004001, 1172839002, 1167371007, 1208966001, 1259096007, 1259703003, 1260327007, 1263538007, 9638002, 12331003, 32595002, 60505005, 64624009, 65096006, 79556007, 81543006, 95457000, 102449007, 106169008, 187100003, 190643008, 230676009, 233760007, 237716008, 237988006, 240401005, 347079002, 396338004, 402463003, 415035006, 419648001, 421684006, 425892007, 427926004, 457767003, 715904005, 723159007, 724784006, 725289009, 726614009, 733110004, 763400005, 764962002, 773421009, 783734000, 789283006, 860865009, 870551001, 34191000119104, 83231000119101, 112971000119109, 113001000119106, 330111000119103, 333081000119102, 434521000124102, 10749811000119108, 15738521000119102, 16644681000119102, 4817008, 60318001, 15989951000119107, 1197332003, 1259038005, 1259049004, 1259701001, 1263551002, 1263558008, 1268463007, 8301004, 9753004, 12348006, 13753008, 29834003, 30292005, 39127005, 49747005, 81475007, 109904003, 128485009, 192719005, 200907003, 206419007, 209178002, 230353003, 230661001, 230766000, 231971006,

237676006, 237726001, 238019007, 238830006, 239086004, 262712006, 276706004, 311495006, 363237008, 363477002, 404224009, 405760008, 406563003, 445213003, 450377000, 714279000, 717266001, 718719001, 720493003, 722386009, 722582007, 722853000, 723666009, 724805000, 726709001, 765135003, 782432008, 72021000119109, 124071000119109, 133301000119102, 290471000119103, 1074181000119103, 1596061000119102, 16001951000119102, 16206981000119106, 16594881000119101, 1144911007, 1148918005, 687331000119109, 1163057009, 342031000119105, 1197052008, 1259693000, 1260382003, 1269423000, 359007, 12010005, 20377001, 23283004, 34680005, 41370002, 41918006, 43661002, 44433009, 47444008, 72442006, 85799002, 86103006, 86819001, 87900003, 92405007, 94154007, 109668000, 111508004, 193008009, 193137006, 209197002, 230204001, 230335009, 230536009, 233751004, 239849004, 267386008, 281510005, 360530005, 363475005, 386805003, 404687005, 445468002, 699059008, 715415005, 717276003, 722989007, 725461009, 732958004, 733028000, 733202007, 762299008, 762350007, 765756007, 773414009, 827115000, 860806007, 1074501000119100, 10624571000119106, 15730961000119100, 15737841000119109, 253184003, 890422008, 1145239007, 1156814008, 1156827007, 1156856007, 1156963006, 1179547007, 1259547007, 1268819009, 1281832002, 16206461000119106, 292281000119103, 23882000, 44771000, 68577004, 69017006, 74297002, 78152008, 94442001, 111306001, 128203003, 128206006, 201067006, 204013004, 209958006, 212179002, 237720007, 247056004, 252246005, 253098009, 301848005, 314783008, 315056009, 403512005, 408671002, 421326000, 426934001, 429765002, 444980006, 609605005, 700381000, 715369006, 719186004, 722385008, 725146001, 725420009, 783139000, 787932009, 866064001, 292681000119101, 1084741000119103, 15633131000119109, 62382002, 403716006, 421283008, 1010663004, 368141000119106, 1187126002, 1187304005, 1177044001, 1179388006, 1222646006, 1259074001, 1259118006, 12056002, 13752003, 15492000, 21323007, 26015003, 32106001, 44341005, 65605001, 76462000, 95833006, 111383007, 126948004, 193000002, 193021002, 193139009, 202659002, 212265003, 212328002, 230150008, 230240004, 230403005, 230694003, 232267001, 238154003, 268956009, 277479000, 315826004, 371632003, 397185006, 403654000, 405761007, 422474003, 423248004, 427939000, 430947007, 457672001, 473429002, 699521008, 703054007, 718770005, 719191003, 721297008, 722633001, 723307008, 724671005, 724723002, 725411000, 732929002, 783176002, 860864008, 5491000124101, 21361000119109, 87651000119100, 334711000119109, 343821000119109, 15733121000119102, 16206621000119103, 25784009, 1156469001, 1204172007, 544102301000119103, 675351000119100, 313781000119108, 1231212006, 1259113002, 1259558003, 1260450002, 1269358004, 488408691000119104, 5510009, 5747006, 9345005, 25185004, 29188005, 35154004, 37732008, 43829003, 51004007, 52276000, 72129000, 84116009, 84461004, 86787000, 90253000, 95414005, 126941005, 186956008, 193130008, 193190005, 206195009, 212214006, 212307006, 212329005, 230390002, 230426003, 230514006, 230526001, 247141006, 298123004, 371022006, 397734008, 403810008, 404660006, 419074008, 420805002, 428635007, 445388002, 702426001, 720494009, 721241005, 721820007, 722985001, 724705005, 733333007, 763715007, 766871009, 770722002, 860841006, 23941000119108, 61301000119102, 307391000119101, 313811000119105, 318701000119109, 331481000119108, 334911000119107, 344451000119102, 15737721000119109, 16052431000119101, 16075931000119103, 16419701000119108, 230268000, 230280008, 230300007, 1217367007, 1231654002, 1259077008, 10941261000119103, 1251484005, 1269377002, 1279839002, 1973000, 17769000, 32232003, 36276008, 71328000, 72960004, 86489003, 95628005, 123710008, 188307009, 192689006, 194063006, 200072008, 208012004, 212194003, 230379007, 230665005, 238828009, 253156008, 254984005, 278027007, 281362006, 398136003, 400947008, 427919004, 442676003, 448045004, 700328008, 710360007, 713527009, 715419004, 715562001, 716023007, 717225001, 718610008, 722201004, 724815006, 733623005, 766818009, 773610007, 784372002, 290411000119106, 291311000119108, 291571000119106, 306151000119106, 430071000124102, 12236991000119103, 16320791000119106, 40230002, 111502003, 1145220002, 1156410008, 1187520001, 1163534008, 1208895006, 1217688006, 1254858007, 1259813004, 1269370000, 21201006, 22135003, 38606009, 45853006, 60338000, 72270005, 73064001, 77224008, 78784005, 94489004, 164109003, 188313000, 191461002, 202687006, 205749001,

209190000, 230258005, 230574001, 232071006, 238635004, 239134005, 247140007, 254171002, 276549000, 282804002, 300993007, 371079004, 386033004, 387732009, 417939004, 446945009, 449796001, 702315006, 715371006, 721459005, 721479002, 722658006, 838307002, 858580008, 68141000119109, 290241000119109, 346721000119108, 432001000124109, 434961000124102, 672501000119104, 690341000119100, 107441100011910З, 1099051000119104, 10962511000119102, 15733401000119101, 420718004, 1052326006, 1156019004, 1156857003, 10852901000119105, 1254911003, 1259069009, 1259830002, 1268815003, 1269360002, 35145002, 39745004, 40852003, 46885003, 49428008, 60146005, 63135006, 68574006, 73813001, 76916001, 78306007, 81285006, 84757009, 84937002, 89369001, 92820007, 95460007, 111480006, 128612007, 186567003, 230211002, 230231003, 230263009, 230562000, 246938006, 253146009, 267890003, 275324008, 307672008, 363498002, 371029002, 371804009, 373426005, 404633004, 404686001, 407675009, 416792008, 426107000, 426293000, 445404003, 698725008, 700507000, 718095000, 721391007, 721790006, 722996009, 723622007, 724065003, 724175002, 724548002, 724795000, 733418003, 737341006, 765216006, 766814006, 818967003, 16931000119102, 97391000119102, 122691000119106, 142851000119103, 431391000124106, 688151000119102, 10972861000119106, 15633361000119103, 15913031000119100, 16040071000119106, 54411001, 446958005, 1163470008, 1162304008, 1208720000, 1231200007, 1259536000, 1285065002, 4229009, 9412006, 10349009, 10552002, 19972008, 27097002, 43645004, 63986002, 72274001, 83752007, 92253009, 109478007, 111236008, 128613002, 186482006, 192966000, 204012009, 212149009, 230256009, 230307005, 230398009, 230696001, 238978007, 247099009, 253151003, 253154006, 275382005, 307365001, 398329009, 426266003, 429514007, 442536003, 702421006, 718772002, 719377004, 722389002, 737273002, 762346000, 770680004, 773989002, 782753000, 789585000, 107581000119103, 290131000119105, 291541000119104, 338091000119108, 338461000119109, 430821000124108, 436051000124109, 11718301000119106, 12240061000119108, 12367411000119102, 403787005, 284861000119104, 686461000119108, 1163063000, 1186799004, 1217047007, 1259075000, 1259829007, 1259640001, 1264275009, 1268716006, 1268825008, 1281839006, 297778421000119102, 13766008, 26520008, 34533008, 39648006, 52913008, 61830005, 90791004, 91956006, 92573003, 95468000, 126984006, 164091005, 192660009, 192897002, 200076006, 204139003, 209097000, 230753000, 233757000, 239869009, 240129002, 254974009, 257277002, 278857002, 371803003, 397763006, 397828008, 422666006, 429033009, 430047005, 442941002, 451037005, 457757008, 699262001, 713487008, 720864008, 721862000, 722436002, 724574008, 724729003, 724820006, 768555009, 838304009, 838389001, 9971000119105, 291981000119101, 309001000119104, 330271000119100, 431091000124108, 15632811000119100, 15637471000119109, 16016151000119105, 890396001, 1156966003, 10852941000119107, 1204176005, 1217040009, 1234782002, 1259029008, 1090511000119100, 1269390009, 1279883006, 5425007, 13151001, 15080006, 21095000, 23414001, 24777009, 25343008, 28778005, 31715000, 41764006, 49949003, 56530002, 58895005, 66046000, 80962007, 90623003, 95646004, 192737006, 194058007, 202677007, 230254007, 230423006, 237558008, 237667004, 240564002, 246565009, 254945000, 262721007, 262730004, 277532002, 302820008, 386766007, 394991004, 426853005, 698766008, 713327005, 719395001, 719985001, 720466001, 721245001, 724174003, 733044009, 733637001, 762457009, 771540008, 785722006, 787037000, 870550000, 311941000119104, 15738201000119109, 16000551000119102, 1082451000119102, 1003445004, 1156850001, 1176993001, 1172899000, 1208341008, 1259619002, 1260399002, 1263576001, 16576004, 31367003, 44201003, 58437007, 59818004, 84759007, 128192007, 129662007, 192709004, 192904000, 199297006, 201835007, 202676003, 230675008, 230804008, 235900003, 240123001, 248315005, 262692002, 314539001, 359624005, 371077002, 403408001, 403530003, 406587001, 417619001, 442685003, 713401006, 715340002, 718754008, 721255002, 721370004, 724817003, 732261005, 733029008, 735554004, 735562007, 737238004, 767254005, 860826006, 866051002, 68061000119109, 291791000119100, 307471000119109, 338051000119103, 1074371000119104, 1075811000119101, 15737161000119103, 15990391000119105, 15990431000119100, 16002151000119107, 16636821000119106, 12367461000119104, 1144968009, 1149278003, 1156409003, 683761000119107, 1187545003, 1163097007, 1234774002, 1259745001, 1268352008,

1268817006, 8563000, 33420007, 64309007, 64518008, 66499004, 75038005, 78458000, 81089005, 84568007, 94110001, 109913001, 126981003, 190502001, 193031009, 193178008, 193207007, 230457003, 238835001, 240845003, 247183001, 253138008, 254963000, 312944002, 371107004, 403539002, 702327009, 715665006, 721333008, 721461001, 721520003, 722978000, 725035001, 732247000, 735749005, 765100000, 782772000, 98541000119101, 99451000119105, 107571000119101, 292061000119100, 292701000119103, 318681000119106, 333241000119103, 432471000124108, 11718401000119103, 12240021000119103, 15678761000119105, 15863931000119108, 15968741000119100, 16016191000119100, 26291002, 128200000, 230438007, 1003463004, 890395002, 1172900005, 1162966002, 1197749008, 1208936008, 1220599002, 1260363007, 1263987001, 1285518004, 4945003, 14168008, 18653004, 27873005, 47371007, 52585001, 55776008, 78097002, 80599001, 81817003, 106018006, 109931000, 111337001, 128128003, 189488006, 191455000, 209987007, 212212005, 230229007, 230241000, 230624005, 233798007, 237697007, 237721006, 276321008, 277187008, 403571009, 403743001, 413100008, 417360004, 422340001, 424054000, 425961009, 718105008, 718606005, 719139003, 719378009, 721090002, 724804001, 765151006, 765212008, 771179007, 771306007, 840452004, 293861000119102, 332441000119107, 672471000119105, 674481000119100, 16057991000119109, 16058471000119101, 16260551000119106, 16716281000119108, 9133005, 403788000, 15678841000119100, 367241000119104, 1231204003, 1228860003, 1231675005, 1259073007, 1259880008, 6204001, 9236007, 10948005, 24321005, 26125006, 26568002, 30475004, 30753002, 33262002, 41115008, 45897005, 46263000, 50589003, 52125007, 77157004, 94131003, 94723004, 95641009, 192718002, 193195000, 201633002, 209196006, 212228000, 212336006, 230287006, 230723007, 238973003, 240070002, 262732007, 276278003, 277526007, 402374008, 403404004, 403505001, 438481007, 698726009, 715666007, 721514004, 721819001, 722111004, 723309006, 724988000, 733184002, 763718009, 763739002, 772129007, 840498003, 860843009, 9901000119100, 35111000119109, 135761000119101, 303241000119108, 308991000119104, 333071000119100, 452281000124106, 674131000119105, 16014071000119107, 110989007, 87921000119104, 1004090008, 1055297002, 880093002, 152148641000119104, 1187124004, 1172585006, 1176989007, 1197343006, 1231627000, 1231645000, 1259742003, 1259492003, 16639391000119106, 281004, 8480004, 16573007, 23869008, 32139003, 32735002, 33595009, 51277007, 73144008, 79840007, 94722009, 95691008, 111496009, 127026004, 163923007, 188325002, 201834006, 202932007, 209181007, 209962000, 230248006, 238027003, 253117002, 254986007, 262704003, 262705002, 262733002, 307359001, 307691008, 420146005, 423582009, 698404009, 698626001, 699812002, 703850002, 704079000, 720637005, 721471004, 721533009, 724682002, 724762006, 724995009, 736838001, 762358000, 765482002, 773305003, 782752005, 784371009, 816119002, 838395000, 840432000, 34101000119105, 302381000119101, 340521000119102, 347591000119101, 436011000124108, 15736801000119109, 16064811000119108, 266673001, 1055298007, 1231192000, 1263934001, 1279837000, 1279844009, 1285078002, 805002, 5120006, 8459005, 22255007, 25821008, 52448006, 70784009, 78028004, 95493003, 129135003, 164103002, 192918007, 201819000, 201857000, 206447006, 212284000, 213210005, 230580009, 230685009, 231456001, 231970007, 241996007, 253106003, 253543007, 359766000, 363055007, 403789008, 406570003, 421019006, 448563005, 698500002, 707608003, 715338007, 721377001, 722381004, 723154002, 724544000, 724558003, 725042001, 733168003, 764733009, 783558004, 788758003, 47351000119102, 93681000119102, 429181000124108, 434531000124104, 672511000119101, 679251000119107, 1074101000119106, 15997591000119103, 192791009, 237694000, 404159002, 16019351000119102, 878808006, 1187467000, 1187617004, 1162345000, 1217051009, 1217212009, 570951000124103, 1237514002, 1254650002, 1259083006, 1259106002, 1259733005, 1259570004, 1259603009, 18442000, 45265000, 52334005, 60189009, 73331006, 111465000, 126955002, 192926004, 212232006, 230405003, 230454005, 232435001, 233844002, 261808007, 262696004, 266126006, 267590006, 269144002, 278899001, 312786001, 312894000, 312945001, 371120001, 399233006, 403197009, 420788006, 445423005, 717013009, 717224002, 718179003, 721008000, 721409001, 721488006, 722139003, 722964001, 723126009, 724798003, 724996005, 733506009, 762672001, 770757004, 774208009, 786078008,

789431002, 870319003, 332341000119100, 332711000119103, 335841000119103, 341811000119103, 344471000119106, 347041000119103, 435341000124104, 1075831000119106, 1089701000119101, 16018831000119106, 1010230003, 890285006, 1145222005, 1156847004, 1259051000, 1259092009, 1259798003, 1260379008, 1263550001, 1269397007, 1478001, 3298001, 8209004, 13973009, 24700007, 32162001, 71044004, 73765005, 81363003, 88538005, 95404001, 126535008, 128126004, 191973007, 193329008, 196135008, 202664003, 202678002, 230658002, 232146005, 238889004, 253136007, 253145008, 417143004, 427975003, 457742001, 700313008, 703303004, 713325002, 713341001, 715908008, 719267003, 721468007, 722966004, 773699009, 783166000, 789016002, 870314008, 10701000119109, 200021000119109, 292201000119106, 292791000119109, 15648691000119108, 16206941000119101, 778060000, 15737881000119104, 1052244000, 684901000119107, 1163061003, 1162523002, 345571000119106, 1204418008, 1259044009, 39191000087108, 8443005, 10327003, 10698009, 24057006, 32022003, 37650008, 54422002, 65720003, 73663008, 74434009, 78572006, 90755006, 111505001, 191504009, 192814005, 206396002, 212273007, 230422001, 240075007, 253107007, 254092004, 285602007, 363058009, 373650004, 397961002, 402339007, 403529008, 414848001, 425879009, 702433001, 703861005, 722293005, 723129002, 765045003, 773700005, 860815000, 290381000119108, 294111000119107, 347481000119103, 429171000124105, 690321000119106, 15633441000119109, 15992631000119101, 16018911000119107, 82300000, 1153332006, 1156470000, 1187468005, 10962631000119106, 1186876007, 1204145004, 346661000119106, 10941651000119102, 4070001, 12869004, 14055002, 16631009, 51854002, 60576007, 73119000, 75122001, 81896006, 83664006, 87095001, 92050000, 95675005, 127302008, 191501001, 193022009, 193411004, 195168007, 204006004, 206448001, 212207009, 212300008, 230354009, 230644004, 230749002, 235038002, 254976006, 264633009, 398256009, 399076001, 403513000, 403851005, 421731000, 429436007, 445116003, 449728008, 698847000, 715406003, 722488009, 723405001, 724783000, 724818008, 724999003, 725907002, 770625006, 827047007, 827048002, 860868006, 292821000119101, 434371000124108, 1074161000119107, 1092311000119106, 12398201000119102, 15675641000119109, 16206261000119105, 198302001, 254196004, 724779000, 890287003, 1157062009, 682231000119100, 1163499003, 10962591000119106, 340541000119108, 1231692001, 1234778004, 1259048007, 1259545004, 1281844004, 23740006, 38941006, 41733009, 43982006, 47339000, 52994003, 75612006, 80849007, 88405003, 91570002, 94156009, 95650006, 116288000, 188290004, 209978003, 212317001, 230360009, 230502003, 230571009, 230614002, 230677000, 237719001, 271700006, 426510004, 430042004, 698624003, 703051004, 715794009, 719944006, 721366007, 721435000, 766709000, 768473009, 789675009, 127551000119100, 182961000119101, 292031000119108, 1074431000119108, 10677711000119101, 15722441000119106, 15930301000119107, 16054471000119105, 45814002, 230387008, 302862001, 1153337000, 1156417006, 1163508000, 1163070000, 1197306009, 18121009, 21244005, 21523006, 22973003, 24326000, 33316007, 44423001, 50776006, 78879009, 85561006, 88687001, 94245002, 94648005, 111850006, 195155004, 247055000, 271569006, 302226006, 352818000, 403632002, 410042009, 425476007, 699942000, 713844000, 715366004, 716051003, 717960003, 719800009, 723160002, 723937009, 724655005, 724668002, 763352005, 772224009, 838323002, 838324008, 313821000119103, 318641000119101, 442241000124102, 16206701000119101, 16270221000119108, 20415001, 64146000, 230269008, 1208883006, 1208338004, 1231186006, 1237342004, 1237462006, 1237578005, 1259691003, 1260362002, 1260371006, 1269363000, 578968971000119100, 36025004, 74885006, 75663006, 79997005, 80935004, 94267001, 109925003, 163920005, 192929006, 212205001, 230456007, 230751003, 236446001, 238705003, 271547004, 302507002, 308757003, 402780003, 406559005, 421973002, 698687007, 698691002, 700307004, 703050003, 712733005, 713624009, 721381001, 722626001, 722628000, 724152009, 782754006, 782886007, 783151003, 788969005, 838321000, 860883001, 870285004, 77841000119108, 311961000119100, 341471000119109, 1089791000119107, 15705201000119109, 15737121000119108, 16014231000119100, 16637761000119107, 768792007, 1156968002, 1157061002, 684941000119109, 1237413006, 1259028000, 1260101003, 1259630004, 1260344006, 1264193000, 45655002, 54739001, 55999004, 63684002, 67279004, 70931000, 77575005, 78914008, 91962001,

192686004, 202796002, 213383004, 230175007, 230334008, 230778001, 230791000, 246540007, 253182004, 307698002, 315352000, 359844001, 398640008, 400945000, 403417001, 404074003, 404075002, 712504007, 717257000, 718210003, 718556007, 721380000, 722435003, 733472005, 765751002, 770682007, 773393001, 785809005, 124391000119108, 290881000119103, 444451000124106, 15632851000119104, 15649941000119102, 315019000, 1156414004, 350631000119102, 1186830008, 345341000119107, 1217077001, 1230143001, 1263544006, 16916421000119100, 14434006, 17974002, 20301004, 42740008, 50122000, 62371005, 66760008, 76886005, 91646005, 109945006, 126991009, 191508007, 191953002, 196475005, 200073003, 204011002, 229665008, 230601003, 230637008, 239088003, 240460008, 254942002, 254959007, 276648002, 363469001, 402782006, 403262004, 403519001, 403572002, 403811007, 409501004, 703041000, 714027006, 715436007, 721233005, 721486005, 723621000, 723823004, 724696008, 724789001, 735557006, 764095005, 765202001, 770758009, 771473004, 773735007, 783554002, 840511005, 866090003, 866094007, 866133000, 361000119103, 1074531000119107, 43477006, 128082002, 1052238004, 680501000119105, 859422751000119101, 1162808000, 1197524007, 1258987002, 1264138006, 5645008, 8837000, 15241006, 23854007, 44547005, 44621005, 50855007, 54314008, 55031000, 73413009, 75241009, 79348005, 79631006, 81211007, 163602004, 186791009, 188301005, 189015008, 193610005, 205750001, 212271009, 230462002, 230686005, 231933003, 237611007, 254192002, 262735009, 307697007, 445497004, 710046001, 716997004, 718774001, 721076000, 763067000, 778070003, 788955005, 860813007, 286011000119101, 291771000119101, 432561000124100, 1074481000119109, 14070001000004105, 15631011000119102, 15731521000119105, 15986231000119101, 17262008, 1197212001, 1259696008, 4645000, 12912004, 21098003, 26594006, 27040004, 37429009, 45294007, 49605003, 52008007, 67725009, 71003000, 71473008, 75885003, 83759003, 95878009, 127341005, 172069000, 188283009, 191494004, 201279004, 201315001, 204005000, 209083008, 212178005, 230148000, 230176008, 230363006, 230465000, 230719004, 231931001, 239832006, 240459003, 246549008, 262693007, 262738006, 264534009, 270881008, 282742004, 283999006, 363034005, 402490007, 415176004, 421821002, 425349008, 427488005, 429759002, 445307009, 449906000, 451036001, 698192005, 698764006, 711158005, 719103009, 719979008, 723830005, 724690002, 724778008, 733049004, 733185001, 733455003, 763827002, 770683002, 870337002, 18541000119100, 68711000119109, 122141000119109, 292811000119108, 295071000119101, 332851000119107, 1077941000119102, 16000431000119109, 16238181000119101, 40354009, 56852002, 398040009, 1157141006, 1163149009, 1163119007, 1186815004, 1217652001, 1260102005, 1263578000, 1269369001, 39231000087102, 19210000, 26610007, 30023002, 38115001, 42376006, 58244001, 58629009, 59748008, 61003004, 62009002, 89498001, 90162006, 91268007, 95569006, 110981005, 123759007, 129618003, 192816007, 206454000, 230160004, 230162007, 230270009, 230542008, 230692004, 233761006, 237674009, 237706000, 238048001, 246826003, 253130001, 299726008, 402361002, 403418006, 406573001, 441688003, 445339002, 609221008, 609641005, 721376005, 722972004, 723366001, 724802002, 735598004, 770941005, 773576000, 870555005, 290541000119106, 292501000119108, 315631000119100, 341831000119108, 678601000119100, 10901691000119101, 15710441000119102, 15733041000119109, 15735801000119102, 16058151000119102, 16476681000119105, 652287331000119104, 1144905009, 1172590009, 822031000000101, 1259499007, 1260374003, 1260397000, 11049006, 14204003, 14447001, 46966000, 50620007, 66104007, 192792002, 200064006, 201645006, 206226005, 208035001, 230227009, 230322000, 230337001, 230540000, 237698002, 238712007, 267390005, 276649005, 312215006, 315049003, 359565003, 403406002, 417183007, 421725003, 433183000, 448248006, 698835006, 702465003, 717822006, 721754004, 723578001, 724563004, 724823008, 726616006, 762337006, 788909008, 303541000119105, 329701000119107, 347251000119109, 437931000124100, 15990111000119104, 12246008, 1145244000, 496369931000119104, 1255323007, 1259515002, 1260364001, 1268702001, 291491000119108, 1383008, 21860004, 28394000, 60503003, 60703000, 67761004, 69347004, 72103000, 83991006, 87185006, 89576007, 94847003, 95774001, 129633006, 163991000, 188324003, 192647003, 193504001, 200061003, 204148008, 229247004, 230430000, 230560008, 232049001, 232142007, 237683004, 240464004, 246758003,

262688009, 262700007, 361132001, 402354001, 420342002, 423084001, 698839000, 721433007, 721483002, 733167008, 762345001, 767659008, 771238004, 771471002, 771476007, 860809000, 100721000119109, 112981000119107, 344621000119108, 430061000124109, 16002111000119106, 16016631000119106, 16583361000119101, 49692006, 231944006, 1163137008, 1163058004, 1172586007, 1162302007, 10973081000119104, 1217046003, 1260388004, 1263977000, 8247009, 13310005, 22306000, 22811006, 25933002, 57838006, 59178007, 62732009, 65017003, 92276007, 95467005, 95643007, 126944002, 194057002, 201852006, 206444004, 212321008, 230168006, 230517004, 231918006, 233672007, 239865003, 247390002, 253118007, 253119004, 253175003, 281222009, 302883007, 400983004, 402719006, 403815003, 404234000, 404240007, 424911007, 457712004, 609593002, 715799004, 720850008, 721517006, 721791005, 724816007, 772791006, 773649005, 787206005, 860884007, 21831000119109, 33441000119100, 290321000119109, 323251000119103, 678841000119102, 16014031000119109, 74351001, 595899961000119100, 1145225007, 1148911004, 86553761000119103, 1172841001, 1228871002, 336491000119103, 330661000119109, 3446000, 13733009, 19448008, 48163001, 76156000, 93557001, 94068003, 188323009, 208067003, 212171004, 212325004, 230690007, 231446000, 231448004, 237707009, 237709007, 238783008, 238850005, 247057008, 254973003, 361216007, 405754008, 699220008, 715951007, 717012004, 718212006, 718579008, 721608001, 722703002, 723082006, 724840004, 733091002, 762287001, 762669008, 770664009, 771272007, 838347009, 866099002, 870295006, 870549000, 122021000119104, 293831000119105, 329371000119101, 329641000119104, 344541000119103, 678901000119109, 1075021000119104, 15737361000119100, 16010951000119105, 16040031000119108, 16058431000119104, 204061006, 1003430006, 1003465006, 679891000119101, 1156849001, 1163101003, 1217048002, 1197277003, 1263501007, 1269226006, 19109004, 30858001, 49422009, 66688008, 75507000, 80758005, 86444004, 89742000, 94767002, 95434006, 126993007, 128197001, 191512001, 193188009, 212347004, 230358007, 230466004, 232001007, 232021008, 240165007, 254960002, 277188003, 363059001, 402454000, 715374003, 717336005, 721403000, 722557007, 724281002, 725033008, 725132001, 762293009, 770094004, 787174003, 870270005, 870294005, 303881000119102, 330011000119102, 348351000119100, 15732961000119104, 16076171000119109, 71791000119104, 890430009, 1187128001, 1201950008, 1259731007, 1260359000, 1260386000, 9386003, 16061002, 35141006, 46721000, 48721008, 51212009, 72180004, 95800001, 187148002, 193950006, 195165005, 203992004, 209145009, 212223009, 230235007, 230684008, 231453009, 247061002, 253239009, 254180002, 267970006, 276220007, 276286003, 307685006, 362975008, 363057004, 402563000, 404690004, 409660009, 422504002, 473372009, 609493004, 700304006, 702344008, 715491000, 722654008, 723133009, 725001004, 732948003, 734987002, 764998005, 766710005, 766934006, 771263004, 773645004, 870267006, 290251000119106, 318661000119102, 435101000124104, 15933901000119104, 16039831000119107, 16206341000119100, 16594841000119106, 15782007, 192705005, 292211000119109, 15680281000119103, 146251000146109, 1173035001, 1163079004, 1231225003, 1259457006, 1259056005, 1279890001, 1284865006, 292291000119100, 1412008, 6348008, 15638008, 18708008, 21328003, 31785008, 36222008, 40449001, 42021008, 43532007, 44031002, 50330009, 59651006, 61536007, 84647002, 95208000, 95477007, 164208001, 192707002, 203980004, 204003007, 204128001, 208056004, 212279006, 230170002, 230177004, 230404004, 237665007, 237731004, 240465003, 276597004, 314008009, 402734000, 417651000, 445059005, 445477009, 699058000, 715797002, 715988005, 716278005, 717825008, 719271000, 722719009, 724769002, 725296006, 733191004, 762331007, 762436001, 792908009, 830275007, 860847005, 134831000119103, 187931000119106, 292401000119107, 330291000119104, 678621000119109, 1074061000119108, 1078031000119108, 16206781000119109, 111504002, 193093009, 237684005, 398229007, 1052318003, 897310009, 168747591000119109, 682241000119109, 759291281000119108, 1177178005, 1204334005, 332051000119100, 1208991001, 1231209008, 1231746006, 1234828008, 1236751004, 1259060008, 1259125004, 1259720004, 1260365000, 1260407007, 1263554005, 1269380001, 11800000, 57373003, 59523007, 61144001, 67754003, 81677009, 83159006, 86266009, 94782007, 186816007, 192805000, 192815006, 199095005, 212188006, 230416005, 230648001, 230757004,

231451006, 237868006, 238698004, 402684002, 403774004, 405764004, 425642008, 445967004, 702419001, 715574002, 720609003, 721399009, 721815007, 722908001, 723023003, 723819007, 735608003, 782718007, 866125001, 91471000119102, 292261000119107, 329651000119102, 332091000119105, 333161000119105, 432141000124105, 1078801000119105, 12239141000119103, 16057391000119108, 21404001, 67155006, 428857003, 1010229008, 1156822001, 1177014004, 1186844002, 1259124000, 1260342005, 1260354005, 9366002, 12454008, 14363008, 17940001, 25044007, 25277000, 29524003, 42986003, 51887003, 64664008, 76435008, 79267007, 80180004, 95671001, 109927006, 128329001, 186499007, 192782005, 201337008, 204044008, 206455004, 206470003, 230215006, 230419003, 230558006, 230651008, 240130007, 277190002, 403255008, 404676002, 406567002, 419378004, 424276002, 425756000, 444957003, 699060003, 699702003, 716380002, 716684004, 717258005, 722056009, 724171006, 724176001, 773729007, 788884002, 838441009, 34181000119102, 318721000119100, 1074521000119109, 1088261000119103, 12050001000004101, 15710281000119108, 15737641000119108, 15737801000119107, 16024151000119105, 43486001, 403786001, 733452000, 1088271000119109, 16036111000119103, 1156965004, 1186883000, 1163065007, 1186882005, 1231206001, 1222659003, 1259129005, 1259805000, 1259198005, 1263523006, 21664006, 22361007, 26298008, 48128006, 62106007, 68926002, 72983001, 77103006, 109789004, 201643004, 212331001, 212343000, 212362002, 230541001, 230627003, 233762004, 237874006, 240133009, 262731000, 363056008, 428964000, 442351006, 700063005, 703541007, 716704007, 718609003, 721508006, 724721000, 725048002, 735534000, 735609006, 735747007, 770751003, 290511000119107, 290741000119102, 290911000119103, 434511000124105, 450431000124108, 1074071000119102, 16076131000119106, 16077291000119100, 897312001, 1156266006, 1157064005, 16019391000119107, 1172684002, 1187215002, 1197523001, 1208620009, 1259081008, 1260380006, 1269458003, 9633006, 13225007, 14637005, 29504002, 56871000, 79554005, 80868005, 84138006, 95793004, 123758004, 126966009, 186707002, 193004006, 195160000, 198901003, 206238001, 212174007, 212289005, 230428002, 232144008, 238826008, 246552000, 254955001, 277921008, 281509000, 302811004, 399088004, 402899000, 404038007, 409555000, 415713009, 429420005, 442735001, 700219009, 703053001, 719717006, 720498007, 724819000, 763798008, 765757003, 766987006, 116811000119106, 145741000119101, 146371000119104, 147101000119108, 290711000119101, 344211000119104, 672551000119100, 679191000119105, 403252006, 1156461003, 1196902006, 1269393006, 10310006, 28534004, 60525009, 82346000, 109924004, 128614008, 129615000, 163922002, 195176009, 202771000, 230278002, 232034009, 237733001, 237899007, 247219000, 253158009, 254194001, 299965004, 307355007, 307680001, 405755009, 410015008, 410509003, 413099000, 432211007, 446028001, 698741009, 703536004, 721240006, 721456003, 721847002, 722866000, 723122006, 723131006, 732932004, 770622009, 770898002, 780827006, 833321003, 866060005, 292661000119105, 292871000119100, 303211000119109, 24800001000004103, 322112361000132104, 1153361007, 1179387001, 1231149005, 1217079003, 1208512000, 1259724008, 12208801000119106, 1263959005, 14357004, 28550007, 43977004, 44295002, 63157004, 111532006, 190472002, 192673008, 192723002, 202658005, 209101009, 209175004, 209179005, 230425004, 230437002, 246759006, 262689001, 262707005, 268612007, 285761007, 361272001, 399244003, 402166000, 402624000, 403407006, 406599008, 429297003, 709415008, 716996008, 719838008, 722027009, 722457005, 724383002, 724764007, 725000003, 725419003, 737230006, 766722008, 770437002, 770791000, 784391002, 87131000119104, 292471000119102, 323321000119100, 15731641000119103, 16000391000119104, 16279021000119106, 18756002, 42530008, 238069004, 262949005, 399040002, 10901411000119108, 1156780000, 1172631001, 1163553003, 1217643005, 1259104004, 20876004, 21086008, 38228000, 50967008, 75837004, 77659000, 79389008, 83405000, 87476004, 92030004, 127300000, 192003008, 193577007, 209163009, 212277008, 229662006, 230667002, 240104008, 247186009, 268962004, 276283006, 302897004, 388871003, 403310003, 406590007, 418572009, 420030000, 424801004, 444229001, 702321005, 702359002, 713704004, 715628009, 719517009, 762516004, 763829004, 763869003, 765774003, 771475006, 860803004, 866062002, 122051000119107, 347991000119105, 15737281000119100, 15737681000119103, 15985831000119107, 16057951000119104, 16650341000119107,

68637004, 80098002, 81780002, 89392001, 15710321000119103, 1177168007, 1186834004, 1567560100011 9107,
1231227006, 1217070004, 1599779100011 9105, 1217136003, 1237418002, 8239009, 8635005, 19206003,
24473007, 33548005, 33772003, 39838007, 62206004, 92294001, 93793002, 93815003, 95815000,
109939003, 126534007, 126973004, 128078004, 135884009, 163192006, 191457008, 191511008, 208007000,
209864000, 212224003, 230377009, 230520007, 231967008, 234510005, 239842008, 243338005, 253113003,
262691009, 302902003, 398660000, 403260007, 438113009, 700326007, 703285005, 715780008, 719988004,
721248004, 721507001, 722606000, 724702008, 733200004, 735610001, 762437005, 763803004, 774150004,
789170003, 8291000119107, 10049100011 9103, 30430100011 9101, 31984100011 9107, 32968100011 9109,
329991000119104, 434881000124100, 19886006, 1186879000, 1186878008, 1208727002, 1260041009,
1259617000, 1269236003, 1269248000, 1269381002, 33832002, 35640008, 37615000, 56253000, 56573006,
56921004, 63362009, 76474001, 83090005, 92051001, 109914007, 126960003, 126967000, 126977003,
164074006, 164097009, 188302003, 202689009, 203996001, 204021005, 204068000, 204941003, 206397006,
212266002, 230359004, 230361008, 230500006, 232124002, 237672008, 237717004, 246538002, 254179000,
308742005, 359629000, 399120006, 402460000, 406581000, 609417004, 698021005, 700324005, 715952000,
718608006, 720749004, 721454000, 721518001, 722632006, 722763000, 722987009, 724067006, 724801009,
763797003, 838320004, 5661000124106, 290361000119104, 292231000119104, 306131000119100, 690231000119102,
12240101000119106, 15633481000119104, 15732881000119102, 15738161000119104, 16661971000119104,
27971006, 230299004, 446311006, 880066000, 686431000119100, 1162462009, 1163078007, 1231846001,
1251447008, 1255252008, 1255268002, 1260352009, 19903002, 25822001, 34209003, 41022003, 74644004,
84947004, 92296004, 94242004, 94246001, 109588005, 129612002, 206235003, 212254008, 212259003,
212268001, 212335005, 230365004, 230433003, 231455002, 236548002, 247209006, 282355000, 301770000,
312116004, 359683002, 359686005, 373661007, 443142001, 447332005, 702427005, 713488003, 715528001,
717938001, 719255000, 719394002, 719520001, 719989007, 722558002, 722616008, 722982003, 723124007,
724543006, 732246009, 737241008, 840501003, 860799000, 860845002, 8551000119100, 97751000119108,
104461000119104, 304321000119105, 321341000119107, 329731000119100, 344891000119109, 438421000124109,
457551000124104, 1075841000119102, 11718351000119105, 15633521000119104, 15737961000119101,
16052471000119103, 41713005, 403257000, 682481000119100, 302661000119103, 1208887007, 1259037000,
1259538004, 1263525004, 1112003, 2736005, 34938008, 55481000, 55489003, 69763009, 77994009,
187094001, 193609000, 212291002, 230311004, 230443000, 237867001, 239840000, 253132009, 255112006,
298119007, 302813001, 307363008, 403653006, 416123004, 426055002, 429691007, 450378005, 450418003,
703302009, 709143008, 719380003, 721246000, 721396002, 722107005, 724542001, 724796004, 763717004,
771307003, 778027003, 782721009, 784341001, 432151000124107, 674161000119102, 10750951000119106,
10962431000119108, 15738321000119106, 16026951000119102, 16039191000119102, 236407003, 341081000119102,
87141000119108, 728667671000119108, 868431021000119100, 1259080009, 1263582003, 1268966009,
1269052009, 20801005, 27148008, 39058009, 41075004, 43105007, 59531002, 62240004, 64077000,
77873007, 78968003, 92099001, 95700006, 109944005, 111297002, 192721000, 195209007, 212363007,
230181004, 230459000, 253109005, 253120005, 253189008, 262724004, 276836002, 277340007, 277950001,
281865000, 311890007, 314006008, 402351009, 403405003, 403652001, 405810005, 428268007, 428438000,
473392002, 609594008, 702383005, 717964007, 719102004, 719432000, 721367003, 723137005, 724675001,
735753007, 762595001, 764732004, 778042000, 782719004, 21651000119104, 29951000119107, 291411000119104,
291831000119106, 292921000119109, 332481000119102, 347461000119107, 433931000124109, 12237231000119107,
15738561000119107, 5963005, 1163490004, 2992000, 9520006, 20897003, 29297002, 30483005, 31882001,
32909009, 35403005, 64741003, 65389002, 67841000, 83168008, 85638002, 89654006, 193064009,
195769008, 204046005, 209164003, 230257000, 230313001, 235042004, 237606005, 237666008, 238023004,
239880009, 254181003, 262687004, 264556007, 278512001, 298130005, 403435005, 403648001, 409661008,
416165005, 430888006, 449020009, 472697007, 697991001, 700332002, 709585003, 710212007, 715755008,

715905006, 721250007, 721383003, 721390008, 722548001, 723146004, 723441001, 723675006, 724679007, 733199002, 733422008, 762361004, 783705006, 290331000119107, 291401000119102, 303281000119103, 429851000124107, 678501000119101, 1084691000119100, 15737761000119104, 16008271000119106, 20343006, 82108004, 460681000124100, 874943000, 1148924004, 1163059007, 1163056000, 1204174008, 1231725002, 1269047004, 1269271003, 1269391008, 1281831009, 9537004, 10325006, 40218008, 68158006, 70350007, 76843005, 84455002, 92451008, 111861003, 188318009, 193756007, 194131002, 195205001, 212311000, 212319003, 229749000, 230563005, 230792007, 232306005, 236384008, 240434007, 253133004, 253139000, 253167009, 255129009, 314766004, 367460001, 386781001, 402353007, 424099008, 425659003, 426128009, 432065003, 441512004, 442512002, 450569000, 698836007, 716701004, 717934004, 718685006, 719164000, 719266007, 720576001, 723557004, 723927004, 724654009, 724992007, 726606003, 733072002, 762295002, 762356001, 771236000, 773548008, 784377008, 792844003, 838339009, 71771000119100, 135781000119105, 141091000119105, 337091000119107, 432521000124106, 678831000119106, 15675561000119107, 15730001000119102, 16024231000119109, 16376191000119109, 403633007, 1003461002, 1003464005, 1187618009, 1208412003, 1179359005, 1259047002, 1268814004, 8231007, 22471005, 28466007, 42074003, 65189006, 75875004, 85423005, 90099008, 94225005, 127309004, 190309006, 198990007, 209082003, 212213000, 212256005, 230511003, 230577008, 230595006, 230603000, 231951002, 253199003, 282747005, 302886004, 312788000, 445166009, 445257004, 720523006, 722066001, 722281001, 722675000, 724431008, 724775006, 735904003, 774147002, 23931000119104, 68091000119102, 124091000119105, 1089821000119104, 10962551000119101, 12241671000119104, 15704201000119105, 15731801000119104, 16002431000119105, 16217941000119102, 1003462009, 15704281000119102, 1145216001, 1187537008, 1177006005, 1197059004, 1187229001, 1260366004, 1263974007, 1268961004, 1269042005, 1279991001, 2040007, 26241001, 45133009, 48780006, 51274000, 52713000, 53748002, 63081009, 67876003, 72556006, 77897003, 89807005, 92427003, 109933002, 128421002, 192927008, 193187004, 195142000, 204074000, 230346007, 230399001, 230565003, 230678005, 230785002, 237610008, 239133004, 262744005, 275362006, 276550000, 371050006, 397809001, 410073008, 428241007, 431266005, 446220004, 446948006, 702326000, 719826004, 721501000, 722970007, 735754001, 737272007, 764456001, 771148008, 775907000, 783161005, 789053008, 827117008, 833326008, 840447009, 866067008, 870343000, 141151000119101, 308941000119107, 336991000119108, 434221000124100, 12237951000119105, 15632971000119103, 15633091000119107, 15648771000119103, 15733201000119102, 15737241000119105, 16046631000119100, 16058031000119107, 428638009, 1010668008, 874937002, 1153334007, 1163055001, 1204417003, 1197751007, 1234779007, 1259089005, 5262007, 7990002, 33217001, 43647007, 45600000, 48090007, 54767005, 67315001, 69233001, 71831005, 126959008, 193183000, 193508003, 229688009, 230297002, 238841008, 238957009, 240312009, 246542004, 264693006, 276651009, 277802001, 302910002, 416659006, 419207008, 425522009, 433083002, 441887006, 699331002, 719207000, 719405005, 719972004, 721072003, 721219005, 721406008, 721410006, 722702007, 722968003, 723553000, 724605002, 724800005, 763403007, 768926005, 778029000, 780822000, 787172004, 816120008, 290891000119100, 329401000119103, 430811000124100, 678611000119102, 10901371000119107, 16818591000119108, 13194006, 68618008, 91952008, 206459005, 420614009, 1186839009, 16917161000119101, 1259086003, 1259723002, 1260391004, 16907601000119106, 2177002, 8511007, 45659008, 48956000, 63942007, 65545003, 68382005, 79855003, 83501007, 95724003, 129179000, 190331003, 190645001, 193118002, 230252006, 230285003, 230289009, 232118008, 266196000, 312898002, 416265003, 423753005, 428668000, 429673002, 444977005, 445072006, 703150000, 703429003, 713425003, 715533002, 721768006, 722907006, 724785007, 733031004, 735751009, 737271000, 762508008, 763136000, 770678005, 778047006, 782824007, 782946000, 783203003, 788981009, 838330008, 27751000119104, 290201000119107, 292341000119102, 348801000119105, 349161000119106, 15639481000119102, 15978711000119106, 44553005, 421415007, 1156833003, 1156837002, 1169361002, 1230005002, 1208961006, 1231663000, 1259128002, 1259783000, 1259467001, 1259478009, 1260336006, 1268965008, 16903441000119104,

5855005, 6836001, 11701009, 18615009, 19276002, 31487001, 41799005, 48210000, 54736008, 59457001, 65172003, 65455002, 66158003, 74012004, 75046006, 84327002, 84590007, 85861002, 194068002, 201837004, 229656001, 229690005, 230417001, 230638003, 230698000, 253101008, 262737001, 277475006, 282787000, 310640003, 361267005, 371805005, 395502002, 402683008, 410060009, 427216002, 698633001, 718851007, 721473001, 722615007, 722630003, 724357007, 733318004, 734434007, 764995008, 782951006, 838392002, 292721000119107, 1074151000119105, 1089801000119108, 15632931000119101, 15675801000119108, 16076011000119100, 193413001, 238026007, 1156456000, 15731321000119101, 1231686007, 1259090001, 1259692005, 25858008, 39134007, 40980002, 45740001, 51604006, 53332000, 53898005, 59714007, 61565001, 86042009, 94784008, 129131007, 129664008, 230305002, 230795009, 237808008, 247176008, 253159001, 253166000, 276230003, 276837006, 277369003, 282785008, 302810003, 403693009, 410471004, 420436000, 427296003, 432616009, 448054001, 448254007, 721092005, 721392000, 721490007, 724684001, 726031001, 726405008, 737227004, 770604006, 783065004, 838371002, 846680003, 140621000119106, 291971000119104, 309031000119106, 331841000119100, 349111000119108, 433911000124103, 678581000119109, 1074391000119103, 1086061000119109, 10901531000119104, 16002071000119104, 80432009, 312991009, 1259770001, 112691000112107, 16637351000119109, 17949000, 38116000, 53776005, 72893007, 79534009, 94248000, 94788006, 111032003, 111522004, 128213006, 128215004, 129617008, 189017000, 191478006, 191492000, 195186005, 195254008, 206588008, 230213004, 230803002, 232269003, 247177004, 254183000, 254940005, 262959006, 268971008, 285601000, 302884001, 331652006, 363482009, 421075007, 698949001, 719189006, 724676000, 732930007, 737338002, 782757004, 23871000119106, 291801000119104, 308971000119100, 329431000119105, 15665521000119105, 15680201000119106, 28368009, 703535000, 1142031005, 1145223000, 1187639002, 1204415006, 16022431000119102, 1217663003, 1208840000, 1231257002, 1196948002, 1231624007, 1259614007, 1263491001, 1285319006, 16023951000119109, 16650161000119108, 11265003, 16195001, 16279005, 17819003, 19091006, 19466003, 25004000, 38086007, 59798009, 75659004, 77547008, 111199007, 111208003, 193615000, 203990007, 230249003, 230539002, 235046001, 237687003, 237696003, 238879003, 255128001, 262958003, 267604001, 282357008, 282791005, 307681002, 371109001, 403820003, 427411009, 710359002, 719827008, 721369000, 722575008, 722980006, 724991000, 2421000119107, 32641000119107, 331781000119101, 338841000119108, 679711000119102, 679911000119104, 19090001000004101, 23772009, 83304000, 314537004, 420756003, 712637001, 10899671000119109, 1145213009, 1156406005, 470721000124103, 1179282009, 1208908003, 1208939001, 1259066002, 1259071009, 1260104006, 1263536006, 1263970003, 1269364006, 1269365007, 14808008, 27270004, 44913001, 54200006, 60268006, 61407009, 79665007, 80499003, 86558004, 91954009, 92453006, 189487001, 191505005, 192138007, 208000003, 208001004, 208042001, 212217004, 212221006, 223176004, 230283005, 230553002, 237730003, 246528007, 285145004, 297278001, 398327006, 698194006, 702354007, 721211008, 721536001, 722147003, 722602003, 722998005, 724822003, 733491005, 771308008, 773425000, 838373004, 866069006, 870554009, 981000119108, 9721000119107, 290721000119108, 291731000119104, 329421000119107, 343861000119104, 688121000119105, 688161000119100, 15648731000119101, 15705481000119102, 15921731000119106, 16035591000119105, 53868003, 784343003, 880065001, 10976711000119104, 1162525009, 1217230002, 23119003, 39502009, 49292002, 52947006, 54515008, 73297009, 73590005, 81412002, 93658002, 109917000, 110150000, 193193007, 206228006, 212177000, 212296007, 230744007, 230752005, 237898004, 253187005, 398432008, 403538005, 409556004, 410041002, 419039007, 420918009, 446995005, 447292006, 715802008, 719990003, 721449009, 724994008, 733092009, 733187009, 764730007, 771146007, 782756008, 783179009, 860812002, 22881000119100, 147121000119104, 292001000119101, 346731000119106, 431411000124106, 434231000124102, 10901731000119108, 12242071000119103, 16066731000119102, 16650241000119101, 70099003, 230272001, 1003431005, 1187533007, 1163533002, 1172705006, 1177067004, 1259041001, 1259121008, 11538006, 22722001, 23880008, 38046004, 53243005, 77486005, 94331006, 95633009, 111501005, 193227008, 201925000, 204745000, 212290001, 230460005, 230745008, 231917001,

238956000, 239873007, 240054004, 253104000, 253188000, 254975005, 277456001, 402722008, 403766000, 425054007, 443153007, 446993003, 698744001, 712820006, 713260006, 715800000, 721463003, 721792003, 723824005, 724097003, 724440007, 725097006, 778003000, 784348007, 788935009, 838383000, 866047004, 103761000119107, 292831000119103, 346231000119100, 461431000124108, 1573688100119101, 88518009, 253186001, 1003429001, 352921000119108, 1229946007, 1208971008, 1231630007, 1251449006, 1259063005, 1259694006, 1263494009, 1269356000, 11442006, 31112008, 38023001, 45281005, 47374004, 54502004, 86125000, 87194000, 192772009, 202683005, 230564004, 230725000, 230747000, 238021002, 239085000, 253238001, 254241004, 255200003, 262576002, 262720008, 282790006, 307694000, 408749000, 416154000, 416913007, 420683009, 425957003, 427907007, 442511009, 443352005, 715822007, 716108004, 718573009, 724761004, 735560004, 763344007, 771234002, 778030005, 782747000, 840454003, 107561000119107, 318731000119102, 329451000119104, 346341000119103, 432031000124101, 1081501000119107, 1240561000000108, 10901491000119104, 16080411000119103, 16206741000119104, 55009008, 81682002, 240048000, 398187000, 1142056004, 1254877009, 1259734004, 1259685000, 30031007, 38453000, 53933002, 76418009, 163925000, 204919003, 208014003, 212233001, 212282001, 212346008, 229694001, 237677002, 238998002, 253149002, 253203003, 253359006, 308680003, 367475009, 402685001, 446496001, 698955006, 715647007, 715902009, 719510006, 720750004, 721249007, 721480004, 722517009, 724782005, 734020000, 737168002, 764969006, 766992008, 768642000, 770679002, 79341000119107, 87101000119106, 110181000119105, 194091000119102, 290091000119108, 329621000119105, 348081000119109, 430771000124100, 431071000124107, 1598403100119101, 360361000, 679871000119102, 1186856001, 1186849007, 1216942009, 1259099000, 1260331001, 2772003, 2951009, 5444000, 9678009, 16561007, 28835009, 38205001, 51258005, 54397000, 74360009, 79366000, 93928006, 109919002, 110995008, 111033008, 126992002, 186498004, 192859002, 192907007, 206453006, 209146005, 212326003, 230179001, 230472004, 230546006, 230635000, 230672006, 236549005, 238823000, 246544003, 262702004, 298125006, 403817006, 404108003, 404658009, 425687007, 426399003, 441722007, 445322004, 445869000, 699328003, 702437000, 703359009, 712986001, 717949007, 719980006, 723304001, 724766009, 726107008, 768554008, 771145006, 773200006, 48601000119107, 104981000119104, 124051000119100, 133981000119106, 137791000119100, 288461000119105, 292381000119107, 337601000119109, 1078791000119109, 15733161000119107, 15182000, 26929004, 86345004, 791000124107, 1089811000119106, 1163541002, 592513751000119104, 1217674007, 1259085004, 1259722007, 5127009, 39574006, 43019009, 47664006, 61386002, 67653003, 69482004, 82339009, 84160009, 94491007, 109534000, 190923000, 196476006, 201275005, 209883007, 212272002, 230282000, 230432008, 230530003, 230629000, 236447005, 237729008, 238949006, 239828000, 268673009, 307212006, 309520003, 387800004, 400077000, 403570005, 414950005, 428259005, 446940004, 450521003, 609595009, 713625005, 719253007, 719680009, 720863002, 721400002, 721465005, 721776008, 723162005, 723306004, 732264002, 733195008, 762329003, 763722004, 765170001, 766044005, 774151000, 774206008, 782725000, 789018001, 860614004, 860804005, 11471000224106, 367041000119108, 15986311000119103, 16023991000119104, 16052671000119100, 16709811000119106, 78141002, 239087008, 408751001, 783787000, 1172602000, 1186829003, 10972901000119100, 1197746001, 1237623009, 1254751008, 1259042008, 1259078003, 822021000000103, 1263529005, 9473008, 17498002, 19233004, 42769004, 43100002, 92047003, 126957005, 128189008, 194134005, 197321007, 201634008, 201831003, 209174000, 230284004, 230556005, 230606008, 230669004, 231928002, 237688008, 237897009, 276630006, 277508009, 281560004, 315047001, 403400008, 403812000, 429458009, 698761003, 715754007, 719252002, 721904001, 724692005, 724767000, 733729003, 734947007, 762335003, 769065000, 27931000119107, 139011000119104, 143521000119103, 309041000119102, 431971000124106, 672451000119101, 274100004, 15984951000119108, 1156796002, 16845341000119105, 1177169004, 293671000119109, 1230414002, 1251456000, 1260375002, 1269383004, 1269516003, 3072001, 14535005, 30337003, 40714009, 49644006, 56943007, 67157003, 72275000, 72986009, 73371001, 82547005, 95610008, 193600001, 194060009, 202794004, 208029001, 212142000, 212249004, 235070002, 238900004, 239833001, 246537007, 247142004,

253135006, 262745006, 300920004, 413101007, 418944009, 423279000, 423361002, 428359008, 442413005, 448135004, 702320006, 707442002, 724561002, 724788009, 725464001, 782771007, 783258000, 846660004, 10899831000119105, 12237151000119108, 418480000, 107557061000119108, 1137357005, 1187046005, 1204125007, 1229876001, 1259773004, 1260449002, 1263971004, 1263978005, 1829003, 14232007, 24408004, 25129008, 25133001, 27642008, 44551007, 51495008, 53482009, 53785005, 73097000, 73706008, 76309006, 84942005, 86073008, 92257005, 94243009, 95702003, 109626000, 110983008, 128212001, 192727001, 204127006, 208045004, 212261007, 230238009, 232058008, 238877001, 246529004, 253125000, 254149007, 262952002, 271764004, 277530005, 278849000, 281223004, 428507003, 442360003, 710349003, 713543002, 715483009, 720852000, 724429004, 724444003, 724547007, 739665005, 763314009, 770594005, 789020003, 10521000119108, 291621000119109, 291951000119108, 677391000119100, 32219008, 32422007, 123763000, 420452002, 428175000, 1155991005, 1259749007, 1259647003, 1260324000, 1260355006, 16638861000119102, 1384002, 5900006, 14763005, 19133005, 23132008, 50722006, 55712002, 87698003, 92071005, 95686007, 126994001, 128193002, 162808005, 186561002, 188327005, 192787004, 212180004, 212239002, 230396008, 230453004, 230659005, 232051002, 237703008, 254820002, 278465006, 371313002, 402612007, 403514006, 413638006, 414927004, 424435009, 426598005, 713706002, 715825009, 718847005, 720856002, 724426006, 724787004, 726607007, 726628003, 762512002, 765191009, 765325002, 860850008, 870350001, 103771000119101, 338441000119105, 434201000124105, 672421000119109, 677401000119103, 678491000119108, 12239101000119100, 15730921000119105, 16057431000119103, 13950001, 27937002, 52281009, 702356009, 1156768008, 182960891000119101, 58173271000119101, 1217042001, 1259114008, 1269343007, 1281835000, 17787002, 28318001, 29093005, 37936001, 52002008, 64228003, 85438006, 163606001, 188292007, 190828008, 193069004, 202680008, 209191001, 211236007, 212202003, 230393000, 230476001, 230707007, 233749003, 239837000, 253170008, 276222004, 276281008, 276829003, 416834004, 427432001, 431468008, 447396006, 702319000, 703543005, 709518008, 713529007, 721823009, 723676007, 725898002, 732927000, 732959007, 733032006, 733082001, 735561000, 735604001, 770596007, 771147003, 771267003, 784346006, 860807003, 22621000119103, 101421000119107, 343191000119102, 430711000124109, 457181000124101, 12398361000119105, 15710481000119107, 35545005, 193839007, 1156472008, 1163532007, 1187567002, 1187195007, 1229938006, 790731321000119100, 1237417007, 1259817003, 1259677009, 1279840000, 23276006, 23853001, 25297005, 27819004, 39944005, 40620001, 46138007, 51978008, 60937000, 64855000, 78569004, 91240008, 94108003, 111500006, 193795008, 204010001, 230209006, 240064008, 278510009, 361123003, 373606000, 388980004, 398963001, 403303009, 698736001, 713693008, 715796006, 715989002, 716868003, 719296002, 720815000, 721330006, 721439006, 721515003, 722390006, 723152003, 735649003, 764522009, 766881008, 773308001, 830033003, 1581000119101, 5731000119108, 68721000119102, 291591000119107, 292741000119101, 336081000119106, 432451000124103, 446531000124106, 12237031000119103, 12241631000119102, 16206181000119108, 16583321000119106, 31235003, 230266001, 422136003, 1157249003, 1179294000, 1163136004, 1163468004, 1177045000, 1187232003, 682461000119109, 1251351006, 2432006, 15523002, 24210004, 35323005, 37481009, 38281008, 47515009, 56271007, 58762006, 74333002, 88425004, 90429009, 92114009, 126949007, 126990005, 229704003, 230320008, 230600002, 240067001, 240473007, 254964006, 256321009, 276595007, 703526007, 713887002, 715737004, 720638000, 721469004, 721903007, 763374004, 771342004, 788911004, 789019009, 870264004, 31081000119101, 295541000119102, 15731841000119102, 1010465001, 313881000119104, 1144942002, 1177122009, 1220595008, 16039471000119108, 1231641009, 1231706000, 1259109009, 1263556007, 16644541000119106, 2065009, 24911006, 28111002, 32728005, 34359005, 67724008, 69664005, 89182000, 111936002, 126965008, 128331005, 164018003, 193067002, 193590000, 201826000, 201858005, 204042007, 208049005, 230207008, 230575000, 230717002, 232014008, 237704002, 246773002, 260912008, 262706001, 287080001, 299727004, 312936002, 312943008, 389088001, 423771003, 431265009, 705128004, 718607001, 721843003, 726669007, 762320004, 765047006, 770624005, 788898005, 789674008,

35091000119101, 47311000119103, 107881000119100, 124681000119104, 290261000119108, 10938941000119103, 36179005, 1148741008, 1186880002, 1163527006, 16039591000119108, 1231178006, 1208483002, 1231190008, 1259812009, 1259556004, 1259568008, 1269379004, 1269385006, 1279976003, 11422002, 29212009, 31283000, 41486008, 46229002, 50921008, 54364001, 59636002, 64631008, 73876000, 82598004, 88877002, 92256001, 93726004, 109947003, 129663002, 191495003, 192794001, 192971007, 202735001, 204036008, 230317000, 230593004, 231908001, 237668009, 237875007, 238711000, 240720008, 249945007, 253103006, 254779008, 276632003, 307690009, 309789002, 313304007, 359621002, 403525002, 403647006, 403803002, 426373005, 429672007, 716243005, 719300001, 723821002, 724545004, 725148000, 762325009, 771336003, 785810000, 860802009, 866104001, 1511000119107, 47391000119107, 291761000119107, 292841000119107, 304261000119109, 347471000119101, 1081471000119101, 10901611000119105, 12237111000119107, 15978831000119108, 128084001, 312956001, 768794008, 897534000, 1186841005, 1172689007, 1187230006, 1231660002, 1231678007, 1255723009, 1259197000, 1259513009, 1263496006, 1263522001, 1268816002, 1269367004, 39171000087109, 18869009, 20110000, 22819008, 43427008, 46578006, 56509006, 62999006, 77835008, 85402004, 86842008, 123708006, 163588007, 191952007, 192649000, 208066007, 212297003, 230448009, 230507009, 239862000, 246928001, 396779001, 413924001, 423488006, 443490000, 713839007, 717812000, 718771009, 721485009, 721524007, 721537005, 722209002, 733331009, 770643005, 771421000, 781129002, 782739000, 789120001, 838314000, 866053004, 3331000119108, 290231000119100, 292091000119107, 293931000119101, 678591000119107, 90584004, 897031002, 1153623003, 1172624000, 341461000119103, 1254868002, 875001, 9909003, 20899000, 20908003, 31429000, 40946000, 53021009, 61145000, 73856006, 75491005, 80328002, 87614000, 92904001, 93767009, 94452002, 95829004, 197367007, 202755000, 203955004, 212208004, 212283006, 230348008, 230625006, 231443008, 238634000, 253181006, 271986005, 278287000, 281110002, 311827003, 402735004, 403253001, 406579002, 424699007, 428544003, 429245005, 704272003, 717014003, 720816004, 721451008, 722064003, 722986000, 723999009, 773395008, 788983007, 870269009, 24991000119103, 290391000119106, 290871000119101, 306191000119101, 318691000119109, 690351000119103, 1075821000119108, 1076401000119108, 1092691000119109, 10999281000119103, 14380001000004102, 15978551000119104, 15989311000119107, 192795000, 230279005, 431011000124103, 15704321000119107, 682451000119107, 684921000119103, 1186734006, 1172630000, 1163076006, 1208478005, 1228876007, 1259055009, 1268888008, 1275625009, 4260009, 4473006, 7199000, 8183009, 8217007, 34663006, 40632002, 57602001, 58329000, 70655008, 73020009, 95652003, 126980002, 164110008, 192905004, 206224008, 209180008, 230333002, 230636004, 230640008, 230656003, 238829001, 238843006, 240042004, 240060004, 240065009, 246775009, 262709008, 279982005, 371756009, 400140006, 403631009, 698004001, 698767004, 703300001, 711406009, 713626006, 715627004, 718605009, 723132004, 724716003, 724776007, 732931006, 733638006, 763597000, 765758008, 768641007, 770655004, 773495009, 838348004, 840442003, 846622008, 870305003, 870553003, 5611000124108, 288861000119108, 315551000119105, 347151000119102, 348501000119108, 435321000124106, 15680241000119108, 15722521000119101, 15997631000119103, 16002231000119106, 16891151000119103, 193570009, 251770561000119107, 1163105007, 1187210007, 1186796006, 1179386005, 1217622009, 675341000119102, 1231659007, 1254936009, 1259130000, 1259732000, 1259816007, 1259679007, 10501004, 16997008, 36803009, 41009006, 52186006, 58749007, 74006009, 123950001, 128470003, 203969004, 209048009, 212320009, 213267001, 230611005, 237689000, 238700008, 240071003, 253179009, 402464009, 415525005, 698760002, 715425000, 719136005, 719278006, 721197001, 721288009, 722604002, 724274009, 724689006, 724770001, 735613004, 774153002, 783737007, 870268001, 290121000119107, 292371000119109, 10973901000119108, 12239621000119103, 15633011000119103, 15989351000119108, 15992711000119104, 16052111000119100, 109921007, 403783009, 874938007, 10861291000119109, 1172636006, 1260128008, 1259625003, 2032001, 21461006, 24225004, 39367000, 48518008, 49386006, 58263000, 59572000, 61819007, 78146007, 89629006, 192993002, 200990006, 202694009, 204008003, 230683002, 238718006, 239950003, 262694001, 262695000, 363061005,

402488006, 416960004, 421735009, 427232004, 448863000, 702393003, 720983002, 721462008, 722493007, 722707001, 735605000, 770404004, 771237009, 777999008, 784347002, 786076007, 295591000119105, 10752641000119102, 10861211000119100, 15631331000119105, 16015391000119105, 41590007, 56989000, 68779003, 82144009, 1141661004, 1187170006, 1220573009, 1259061007, 1259068001, 1260134001, 1259673008, 1263503005, 5934007, 6400008, 7052005, 7125002, 7806002, 23645006, 40933005, 44268007, 56795008, 62660000, 66010009, 76562003, 77600008, 81042008, 83982007, 92905000, 93747000, 109680008, 109902004, 189164002, 203363008, 206395003, 212192004, 230790004, 237664006, 240047005, 240313004, 361199007, 398726004, 402779001, 414875008, 425365009, 426167003, 442942009, 445359003, 699704002, 721247009, 721752000, 722547006, 722623009, 763135001, 782755007, 101571000119107, 122061000119109, 306211000119100, 1074441000119104, 1078041000119104, 15648601000119102, 15733361000119105, 16058391000119109, 56120004, 142971000119102, 1003921006, 1156407001, 1162342002, 1220598005, 1237475006, 1260368003, 1263489009, 8840000, 13706005, 25913001, 28155008, 29618004, 32515001, 36217008, 40640008, 49562005, 50888000, 61687004, 63649001, 66451004, 69316005, 72092001, 77706007, 82344002, 82696006, 92682003, 94786005, 129620000, 163601006, 164467006, 171131006, 192685000, 202696006, 203994003, 209050001, 212269009, 212345007, 230544009, 239876004, 246717005, 246768008, 254217002, 265622002, 271674009, 277321001, 282788005, 307177001, 307687003, 307692001, 396326008, 443369007, 444748003, 445475001, 449305009, 713491003, 715819005, 721229003, 721431009, 721455004, 722378009, 722380003, 724803007, 733113002, 827050005, 870591003, 131411000119107, 291641000119103, 292611000119107, 15699081000119101, 15731001000119108, 15738241000119106, 15989991000119102, 16056231000119104, 49497005, 87161000119107, 1208747005, 570901000124102, 1259057001, 1259503001, 1259663006, 1260271009, 1263547004, 15978511000119100, 563789641000119101, 3449007, 21615003, 23024003, 40725000, 51881002, 53318002, 59671002, 61200008, 67682002, 73462009, 75143000, 87607002, 88923002, 93828005, 94153001, 194132009, 206223002, 230474003, 230581008, 230641007, 233759002, 237608006, 241992009, 254153009, 276322001, 276596008, 372441001, 419096002, 450886002, 699655001, 717286002, 721395003, 724807008, 734099007, 764849002, 764859001, 768934004, 783174004, 860866005, 292711000119100, 347541000119109, 348581000119100, 690271000119104, 15933941000119102, 10901451000119109, 655081461000119101, 16039671000119106, 15731241000119101, 1222710008, 1237470001, 1281810006, 4598005, 10706006, 12394009, 33346005, 33871004, 52274002, 57948007, 74810008, 80976008, 92808004, 94766006, 95150005, 95399003, 95638000, 106014008, 109942009, 188297001, 193009001, 197659005, 209162004, 209902003, 212302000, 229738006, 230445007, 230468003, 230714009, 237691008, 238025006, 238842001, 239871009, 240062007, 240167004, 253142006, 262747003, 286933003, 311825006, 387602005, 387891008, 426394008, 429408002, 698617004, 700336004, 716723000, 717939009, 718759003, 719987009, 722282008, 722556003, 724720004, 733330005, 763370008, 764453009, 782390002, 785345002, 866056007, 292361000119103, 435331000124109, 684911000119105, 16026631000119103, 36524008, 237695004, 292411000119105, 1003375005, 1003884001, 16018991000119103, 1145217005, 1156772007, 1156842005, 687361000119101, 1186868005, 1177053008, 1204175009, 1230026002, 1259769002, 1251457009, 1263492008, 264008, 20447006, 35204001, 39427000, 45912004, 48662007, 57803009, 58557008, 64634000, 64678009, 73181007, 79520009, 80304004, 89449005, 89525009, 93559003, 95499004, 95663000, 95703008, 126954003, 127311008, 128533009, 129606007, 129666005, 192706006, 193078005, 203941007, 208057008, 208065006, 212299000, 213054005, 236404005, 236412002, 237692001, 238702000, 240049008, 240462000, 262719002, 314407005, 361273006, 363222009, 426187002, 609609004, 719816006, 723497003, 726609005, 773990006, 785299009, 870340002, 119001000119108, 292671000119104, 345021000119104, 672541000119102, 15632891000119109, 7259005, 204062004, 282364005, 421827003, 1172593006, 1186888009, 1208859003, 1208833001, 1208886003, 1197587003, 1217209006, 1236807002, 1251371002, 1259067006, 1264527003, 1279886003, 16638971000119103, 16650301000119105, 3577000, 8528005, 15662003, 26016002, 27362007, 28634005, 31076000, 40135004,

49778009, 57182000, 62440002, 75299005, 76675006, 78675000, 95709007, 95710002, 126986008, 127299008, 193148004, 203985009, 208034002, 230208003, 230308000, 230329009, 230763008, 236552002, 237680001, 237710002, 239174004, 240076008, 240431004, 253143001, 312924008, 361133006, 402733006, 419737003, 420996007, 424728002, 449221001, 609383005, 703627005, 713489006, 715463008, 718572004, 719282008, 721527000, 723359002, 724283004, 724806004, 733489002, 734989004, 735907005, 870342005, 138371000119104, 292121000119106, 292431000119100, 323311000119107, 339481000119109, 1620691000119103, 418531007, 1003424006, 1163103000, 1163546007, 1208901009, 302711000119108, 1197280002, 1259110004, 1269373003, 3548001, 29315008, 42069006, 43602006, 46251005, 54203008, 62878007, 64764001, 87513003, 95502000, 95794005, 186479001, 188280007, 193174005, 201277002, 209067008, 212298008, 231454003, 237708004, 238905009, 253177006, 253742004, 262708000, 267576008, 370891001, 371561008, 424941009, 715820004, 718174008, 720410001, 722990003, 733089005, 763622006, 765089003, 782679002, 783701002, 788756004, 788881005, 789676005, 840449007, 291781000119103, 346711000119101, 16019471000119104, 10394003, 66108005, 416780008, 421597001, 1149028005, 1157060001, 1187466009, 1186853009, 1197476009, 1208880009, 1208960007, 1217008005, 1231634003, 1259307004, 1259579003, 291421000119106, 1670004, 14521008, 16006009, 23150001, 29003001, 32739008, 63198006, 66022007, 85505000, 90448008, 94807005, 94808000, 94833002, 94868002, 95647008, 109419009, 187253003, 193114000, 206194008, 209921006, 209940000, 230187000, 230243002, 230247001, 230309008, 230364000, 230455006, 230781006, 238901000, 246550008, 254949006, 262740001, 262742009, 281365008, 310387003, 422471006, 444860006, 448314007, 717003001, 718551002, 721476009, 721825002, 722902000, 722981005, 724680005, 726404007, 763068005, 763802009, 292251000119105, 349061000119107, 677381000119103, 688101000119101, 16376241000119109, 21007002, 29145002, 1153362000, 352731000119109, 1186656004, 10625871000119105, 1259814005, 1268813005, 1268957005, 1268964007, 1674008, 1852004, 30736009, 31430005, 33927004, 45170000, 45299002, 88755007, 93749002, 111868009, 191486006, 203948001, 204086002, 207998001, 209947002, 212274001, 230347003, 230549004, 230630005, 232013002, 238649008, 239838005, 246551007, 253140003, 262711004, 276589006, 279953009, 280955001, 348083004, 403822006, 429419004, 442224005, 473373004, 700316000, 713450000, 713837009, 715668008, 716305005, 717042001, 762324008, 764736001, 773230003, 782826009, 866050001, 8041000119108, 290731000119106, 292861000119106, 295471000119105, 306531000119108, 323261000119101, 338681000119102, 347211000119108, 347521000119103, 672381000119100, 679171000119109, 679931000119109, 1074421000119105, 1074471000119106, 15692681000119101, 15933861000119105, 16000351000119109, 16039391000119100, 70694009, 1148444001, 687351000119103, 313871000119102, 1259127007, 1259750007, 1259632007, 1260396009, 1263531001, 1269257006, 24067001, 72513001, 76349003, 79897009, 87058001, 90496008, 126972009, 127346000, 128283000, 190492009, 193209005, 200071001, 203954000, 204047001, 209956005, 230143009, 233758005, 240072005, 240125008, 254227008, 254947008, 281240008, 307356008, 314540004, 402616005, 403573007, 404095008, 404647000, 418871005, 609640006, 698625002, 698782009, 699656000, 717259002, 721510008, 735537007, 758664007, 764272007, 765195000, 770939009, 778046002, 782887003, 788913001, 788956006, 789046004, 789657008, 2701000124105, 5471000124102, 105151000119105, 678881000119107, 15985111000119106, 420658009, 1153340000, 1156789004, 1156969005, 759950981000119101, 1187619001, 1163540001, 1222658006, 1259496000, 1263511000, 965003, 5102002, 29590001, 29740003, 39194005, 59754009, 71981002, 75468006, 78157002, 127307002, 180234006, 200982006, 201069009, 203392007, 212200006, 230523009, 230602005, 237872005, 238030005, 238605006, 247384001, 254147009, 254948003, 297944001, 301766008, 307689000, 314787009, 402721001, 403699008, 403802007, 405753002, 698755001, 713506004, 713956000, 717894000, 718214007, 719208005, 721749008, 722576009, 724674002, 733198005, 762321000, 860846001, 860878009, 866131003, 870288002, 290401000119108, 291581000119109, 292511000119106, 451971000124105, 16000471000119107, 30371007, 1144966008, 1163471007, 15709601000119109, 16917241000119108, 1222662000, 1259108001, 1260338007, 1263507006, 1279891002, 38403006,

40308000, 48713002, 49001006, 53675004, 86263001, 95454007, 111390002, 123614006, 193073001, 202662004, 212215007, 212286003, 230242007, 230350000, 237517001, 237705001, 238061001, 238975005, 282801005, 285758006, 402614008, 403284004, 423086004, 426134002, 444855007, 449797005, 700294000, 703174002, 713869000, 715629001, 715807002, 716724006, 716994006, 717975006, 718845002, 719254001, 723390000, 723825006, 724687008, 762360003, 762362006, 762509000, 763458005, 763770005, 770565003, 96981000119102, 313501000119105, 431461000124109, 1081541000119109, 1099041000119101, 10901771000119106, 15986191000119105, 16008191000119104, 16026991000119107, 688141000119104, 1144625005, 682251000119106, 1177011007, 1172838005, 1204351003, 1204463009, 1231267007, 1237194006, 1237370002, 1263559000, 1263579008, 1263976009, 16637571000119106, 16639981000119109, 26738009, 52677002, 76670001, 90768003, 93657007, 110988004, 190481008, 191972002, 192648008, 192877007, 231929005, 232061009, 233753001, 239825002, 240050008, 240051007, 249174002, 267602002, 282786009, 298137008, 307200007, 312216007, 371040005, 402778009, 403669004, 428870007, 443502000, 451034003, 712883005, 713826008, 716335003, 719190002, 721529002, 722909009, 723408004, 724719005, 725413002, 766752000, 772126000, 785302009, 290151000119104, 291121000119103, 303041000119104, 333091000119104, 349281000119109, 1074141000119108, 1074401000119101, 15738361000119101, 16023911000119108, 16024111000119109, 16036071000119105, 16039151000119107, 16206541000119108, 479003, 80651009, 230706003, 367528006, 303861000119106, 16019311000119103, 1217371005, 1222655009, 1208871009, 1237129009, 1260117009, 151004, 385006, 4463009, 17602002, 18200000, 23011003, 23798003, 24630008, 27830001, 31681005, 36603006, 39960009, 40673001, 43659006, 46549005, 95830009, 123711007, 123798002, 191510009, 192796004, 192979009, 193039006, 194069005, 201724008, 206439006, 208028009, 230519001, 230712008, 230718007, 231915009, 238156001, 238896002, 262726002, 275361004, 403821004, 404225005, 702439002, 709977002, 712537009, 718849008, 719185000, 721243008, 722601005, 735461003, 762515000, 763669001, 293951000119107, 307451000119100, 15737081000119106, 16058351000119104, 27529000, 1163141007, 1177060002, 1229943004, 1237515001, 1260129000, 1259554001, 1259584009, 1263514008, 1269372008, 3308008, 16170002, 55382008, 59026006, 67081008, 67854007, 68863007, 69820004, 70835005, 71286001, 79705001, 128117002, 191447007, 192655005, 209147001, 212312007, 230589005, 230657007, 237662005, 237669001, 237725002, 253153000, 255123005, 270907008, 275434003, 298126007, 311789006, 313259008, 362967002, 363235000, 363471001, 447655002, 448218008, 699688008, 703301002, 713417000, 718555006, 721534003, 723156000, 763387005, 788932007, 816068000, 866059000, 18301000175107, 329741000119109, 434551000124106, 12242111000119105, 15984991000119103, 16002511000119104, 5158005, 52165006, 95330001, 1153376004, 346674811000119104, 1186872009, 1172703004, 1197494003, 1230017000, 1259059003, 1259111000, 12208841000119108, 1260328002, 1269050001, 1269376006, 1269412004, 568005, 3900008, 30871003, 42365007, 51443000, 58942006, 65457005, 71460009, 72655000, 78071008, 88092000, 92683008, 95485003, 118951003, 129611009, 192894009, 201642009, 212197005, 230159009, 230182006, 230366003, 231995008, 234021009, 253114009, 253183009, 255522009, 269255007, 276277008, 276284000, 370483001, 426599002, 437871001, 609590004, 717223008, 721492004, 721609009, 721824003, 722900008, 723147008, 724424009, 762322007, 773552008, 785307003, 113041000119108, 291821000119108, 292941000119103, 330461000119107, 15675761000119100, 21634003, 30278004, 874940002, 1144894004, 1187279003, 1163096003, 1163459001, 1208513005, 1260340002, 1263505003, 1263535005, 1268821004, 2415007, 12166008, 14289006, 53857003, 90302003, 93768004, 95631006, 192714000, 193214009, 194177006, 198438009, 202692008, 230323005, 230326002, 230617009, 254961003, 276282001, 402493009, 403661001, 428233007, 442183006, 449799008, 472698002, 715770009, 721165001, 721244002, 721436004, 724677009, 762513007, 783622001, 860886009, 290171000119108, 290901000119101, 293811000119100, 313911000119104, 335831000119107, 434291000124103, 16054391000119102, 16419741000119105, 1010604007, 849579281000119106, 1172701002, 1186837006, 1187231005, 1259045005, 1259588007, 1264280000, 1269049001, 2169001, 6537000, 15445004, 25397008, 28193003, 37001003, 39951001,

47185005, 55637002, 62904009, 72052003, 93660000, 186509002, 186624004, 192713006, 201637001, 201646007, 229681003, 230639006, 230664009, 247187000, 253124001, 254956000, 254980001, 269496008, 275470000, 307405000, 372310001, 420920007, 422240004, 424151006, 432504007, 451038000, 698291007, 700295004, 702381007, 715437003, 718702003, 719205008, 721252004, 721371000, 721484008, 723820001, 739666006, 763455008, 764993001, 860808008, 866130002, 84221000119101, 87111000119109, 98811000119103, 332831000119101, 431111000124104, 680781000119108, 15731041000119105, 421920002, 890216002, 890350009, 1148694009, 1153395006, 1156800008, 686441000119109, 1172606002, 1179297007, 1231244005, 570931000124105, 1208481000, 212455011000119105, 1255271005, 1087891000119107, 1260377005, 1263583008, 1269359007, 1269392001, 21202004, 29733004, 29807001, 31249007, 31524007, 39807006, 46829007, 50320000, 63900005, 67437007, 69195002, 95883001, 111897007, 128194008, 193184006, 209079008, 212262000, 230440002, 238155002, 239841001, 240875008, 392662004, 397751001, 422724001, 429437003, 443001001, 700338003, 715990006, 720830009, 720956003, 722455002, 722624003, 722627005, 722901007, 724763001, 725163002, 735615006, 773280009, 783158009, 785305006, 860805006, 2611000119103, 20121000119105, 112991000119105, 140281000119108, 304341000119104, 330711000119104, 434101000124100, 434491000124104, 434541000124109, 436041000124107, 16236661000119100, 403258005, 890434000, 1177012000, 1177001000, 1217665005, 1197152005, 1208486005, 1231689000, 1263573009, 1279730009, 39211000087107, 46795000, 51689003, 56384000, 60914006, 69116000, 91483004, 91637004, 93746009, 94601008, 192712001, 194070006, 212225002, 212227005, 213208008, 230312006, 230408001, 230413002, 230703006, 233956002, 240432006, 262961002, 264906008, 308634000, 312372003, 403506000, 403630005, 424578002, 425237009, 426566004, 429306002, 443333004, 449203004, 699520009, 700293006, 713747004, 715534008, 715752006, 719379001, 721475008, 723151005, 770434009, 770727008, 771305006, 5571000124103, 368521000119107, 432291000124105, 436611000124103, 15682841000119106, 15736841000119106, 16320871000119102, 239881008, 939885431000119109, 1172595004, 1172793008, 1186884006, 339971000119103, 1197363004, 1217228004, 1231631006, 1259597006, 1260357003, 1275631007, 16819301000119102, 10877007, 16652001, 17944005, 19373007, 25957002, 51514006, 66924005, 72758005, 86907008, 111633007, 126982005, 129613007, 163665004, 191466007, 195167002, 204022003, 209092006, 209194009, 212270005, 213197002, 230332007, 231445001, 237870002, 315353005, 403393000, 431520004, 703305006, 718219002, 719521002, 721373002, 721407004, 721481000, 721748000, 723936000, 737582007, 762357005, 773415005, 773648002, 84161000119100, 105421000119105, 292451000119106, 293971000119103, 330411000119109, 344461000119100, 431061000124100, 431431000124100, 432501000124101, 15710601000119102, 15733001000119107, 16046951000119104, 16057351000119103, 56231002, 1003374009, 1010610007, 1010643006, 1010644000, 890432001, 1176991004, 1259248003, 101631000119108, 1269051002, 10575009, 26595007, 41497008, 42295001, 49238001, 68116008, 77015008, 77759009, 94086000, 94669007, 95478002, 129593003, 163664000, 163910000, 191449005, 195141007, 203997005, 212203008, 230370006, 230488004, 230631009, 230689003, 231517009, 237702003, 247062009, 282799007, 400190005, 405722004, 419476001, 421455009, 444197004, 449903008, 715777007, 716700003, 716999001, 719210007, 719455002, 721818009, 724227000, 732952003, 738276008, 762340006, 763130006, 764688002, 765434008, 782743001, 782825008, 784380009, 788984001, 860834000, 17761000119109, 53521000119106, 61291000119103, 291511000119103, 308961000119106, 318711000119107, 433691000124104, 434091000124106, 15705361000119101, 15960661000119107, 16005191000119106, 16008111000119108, 16053991000119103, 50801008, 1075751000119105, 16019431000119102, 1187045009, 1186861004, 10852861000119104, 16005311000119104, 1290801000000102, 1264565005, 4062006, 23713006, 31646008, 42345000, 44087005, 46995009, 73840001, 79591004, 85007004, 90224001, 93433003, 126978008, 128120005, 188287005, 193602009, 194065004, 203986005, 212236009, 229676007, 230153005, 230345006, 230521006, 230561007, 231442003, 231898006, 254946004, 262701006, 262957008, 276722003, 315057000, 398170002, 445478004, 609612001, 716667005, 718176005, 721231007, 732251003, 762336002, 763533003, 840721004, 866058008, 113011000119109, 124041000119102,

134821000119101, 292071000119106, 303941000119103, 311221000119103, 343841000119103, 429831000124100, 436771000124102, 15639441000119107, 15731281000119106, 1515008, 21921000119103, 1172696009, 561258721000119104, 1231682009, 1269371001, 1284855000, 4183003, 5361003, 27908001, 29159009, 43763009, 48376004, 50866000, 50933003, 53622003, 60644007, 63627007, 69206000, 73431005, 85592008, 189167009, 191503003, 193030005, 201278007, 204040004, 209165002, 212186005, 230336005, 230594005, 239863005, 254941009, 276599001, 277186004, 282746001, 302225005, 367137004, 387893006, 399993004, 402617001, 425805004, 425988004, 609591000, 699701005, 703522009, 708013001, 717942003, 718848000, 719836007, 721440008, 721470003, 788931000, 789014004, 789588003, 816984002, 866068003, 870284000, 292461000119108, 337461000119108, 345611000119102, 349301000119108, 15637191000119103, 16076051000119104, 16476641000119100, 1010653007, 15629411000119106, 1156841003, 511452481000119102, 1187149000, 1231168008, 1196839006, 5724005, 10629009, 14405000, 15938005, 17141001, 56409008, 60750009, 71047006, 74728003, 80734006, 123709003, 126976007, 203934001, 209189009, 212295006, 224186005, 230286002, 230470007, 230616000, 230708002, 231997000, 232054005, 235037007, 240061000, 253011004, 253160006, 254966008, 307176005, 313257005, 314009001, 389098007, 428141001, 428687006, 698619001, 715803003, 717826009, 721523001, 721531006, 735599007, 737239007, 765744006, 765773009, 768556005, 788632005, 838331007, 860951008, 126571000119107, 678671000119105, 1074171000119101, 10846221000119106, 12367361000119109, 253185002, 422089004, 16207161000119102, 1156855006, 1187643003, 1187565005, 1172843003, 1186836002, 1236804009, 1259046006, 1259070005, 1237425009, 1260341003, 1263980004, 1269555002, 7713009, 9551004, 65260001, 80965009, 88269008, 89601008, 92255002, 93946000, 93964007, 129132000, 190470005, 202776005, 230384001, 230739000, 231909009, 277326006, 282793008, 285596009, 402731008, 719183007, 722988004, 724567003, 737229001, 785300001, 870556006, 292781000119106, 323171000119101, 433711000124101, 1074571000119105, 15648641000119100, 15697321000119105, 16052711000119101, 72284000, 206456003, 446312004, 1145240009, 1157068008, 1172683008, 1186827001, 1231240001, 1263957007, 1263972006, 4040006, 4510004, 4619009, 8939001, 26155004, 53810004, 74772000, 76938004, 79161002, 82058009, 126963001, 192720004, 193206003, 195189003, 202675004, 208031005, 209102002, 212315009, 230314007, 230666006, 230673001, 233754007, 238899009, 240078009, 246548000, 249763000, 254983004, 267612009, 278285008, 286914008, 298117009, 301765007, 363470000, 422873003, 428965004, 713482002, 715795005, 716198008, 722617004, 723183004, 724814005, 737231005, 762663009, 770623004, 783180007, 785304005, 827049005, 142001000119106, 217791000119107, 337951000119101, 688171000119106, 1075001000119108, 10901571000119101, 15740881000119102, 15960381000119109, 16236701000119107, 48982009, 1156455001, 1187506008, 1163091008, 1186813006, 1231256006, 1275624008, 330221000119101, 10196009, 17258002, 37075008, 38073003, 39832008, 42012007, 42998008, 75019001, 84062004, 118940003, 123615007, 190471009, 192813004, 201631000, 212323006, 230321007, 232053004, 233716007, 235928009, 237711003, 237869003, 238704004, 238948003, 240566000, 246545002, 248915001, 275269004, 406594003, 422061002, 427796004, 457652006, 609599003, 719817002, 721368008, 721530007, 722584008, 723404002, 723721007, 723928009, 725012004, 838529004, 866126000, 291531000119108, 292051000119102, 431101000124102, 15685281000119108, 15710561000119102, 16005111000119102, 56453003, 770095003, 672441000119103, 879937000, 1230343006, 1230273004, 1264139003, 991898981000119108, 1845001, 10076004, 15228007, 17140000, 37991008, 39181008, 43521007, 46826000, 58193001, 74116004, 87065009, 90531003, 94224009, 127324008, 193576003, 230338006, 230579006, 230761005, 235043009, 267592003, 267601009, 278855005, 314427006, 403286002, 425868004, 441519008, 609461007, 702464004, 710357000, 720512007, 723125008, 723828008, 724549005, 736432009, 762511009, 782935003, 783062001, 783064000, 788899002, 860796007, 291651000119101, 292771000119108, 323301000119109, 431401000124108, 678511000119103, 10846261000119101, 15968581000119104, 16035391000119102, 915141931000119109, 1230376005, 1208998007, 1259762006, 1260398005, 1279884000, 17585008, 27741009, 49420001, 57981008, 68216000, 74460005, 91955005, 94646009, 230251004,

230429005, 238642004, 238717001, 246937001, 254943007, 274659008, 276458009, 281363001, 371031006, 403170002, 426536006, 426592006, 428190001, 433493000, 715662009, 715727009, 718755009, 719021005, 721535002, 735603007, 735607008, 737240009, 763065008, 771141002, 788757008, 788882003, 432541000124104, 16039631000119108, 229654003, 230194002, 768793002, 898941951000119108, 1163542009, 1230308005, 1259079006, 1259740006, 1263960000, 1264528008, 15742000, 19883003, 30310000, 66074005, 94855005, 94963005, 95689000, 109922000, 111878007, 191476005, 212220007, 230324004, 231924000, 237283007, 240569007, 254954002, 254968009, 371024007, 404109006, 698734003, 702628006, 706891008, 712647003, 717732009, 718224004, 719043002, 721445003, 722451006, 723158004, 726704006, 734986006, 737057000, 765093009, 784289008, 788883008, 789119007, 836301008, 870341003, 2851000119101, 330331000119105, 347071000119105, 432491000124109, 674361000119104, 1074381000119101, 10899791000119104, 12266161000119102, 15731881000119107, 15749161000119108, 15987431000119100, 16013711000119104, 16026551000119108, 1172840000, 1260369006, 1260405004, 1269342002, 7379000, 23168003, 28143002, 31568009, 52522001, 59979003, 68859000, 68900007, 82800008, 87888006, 192958009, 193010006, 193400007, 209085001, 229655002, 230378004, 230392005, 230734005, 230801000, 237120002, 238951005, 239084001, 253169007, 254977002, 280816001, 307693006, 406572006, 406589003, 423441007, 424340000, 425576009, 441526008, 609613006, 703524005, 713847007, 716200002, 722432000, 724349009, 735558001, 763865009, 766932005, 771271000, 866093001, 100511000119108, 285151000119108, 303921000119109, 337711000119100, 347561000119108, 15639401000119105, 16002031000119102, 16864831000119104, 39925003, 237693006, 237960000, 1156835005, 1163550000, 1222649004, 675331000119106, 335801000119100, 1259473000, 1259665004, 1263545007, 1269382009, 8663007, 16350000, 19593003, 26808007, 29963001, 58638006, 70328006, 80544005, 190480009, 190823004, 191465006, 191519005, 192659004, 193247000, 201924001, 212199008, 229664007, 230607004, 230633007, 231912007, 233817007, 240132004, 284449005, 311826007, 312792007, 313307000, 314950006, 392481002, 416813006, 448989001, 449793009, 698072001, 715439000, 719431007, 721242003, 721382008, 721428008, 722965000, 724847001, 732926009, 735206008, 770564004, 783724009, 866123008, 26261000119109, 135321000119107, 290491000119102, 344091000119101, 1085361000119104, 16052631000119103, 16206301000119102, 47362003, 75834006, 770098001, 890286007, 1156790008, 517253051000119105, 1186724002, 1186819005, 1163549000, 1208621008, 1217655004, 1231228001, 1259112007, 39241000087108, 1538006, 6607004, 15488007, 40122008, 42452002, 51044000, 61484000, 63983005, 65144005, 74231003, 95108005, 111503008, 192812009, 193129003, 204171005, 222158003, 230152000, 230327006, 230586003, 232221001, 246539005, 253171007, 267384006, 301455002, 359837005, 404239005, 413807008, 418143002, 424497004, 443876008, 609559001, 698837003, 721388007, 722453009, 722943000, 723364003, 733068001, 734988007, 735555003, 737234002, 773627004, 774070008, 99131000119108, 128211000119102, 291871000119109, 303961000119104, 307371000119102, 323821000119107, 335531000119109, 338771000119109, 436591000124109, 672531000119106, 1083141000119100, 10629311000119107, 10901651000119106, 15633401000119107, 16006591000119109, 1142207008, 682261000119108, 957319791000119104, 1162839003, 1217370006, 1230397006, 1259581001, 1259658007, 1593000, 4381006, 5842009, 10649000, 26135000, 31839002, 36696005, 42341009, 43398005, 51723007, 62985007, 94968001, 109923005, 126989001, 128080005, 128191000, 163595003, 188317004, 203975008, 209094007, 210039000, 230355005, 262697008, 274945004, 276588003, 281899002, 282795001, 298282001, 360414000, 398148000, 400946004, 403398009, 420006002, 424549003, 426936004, 445900004, 448148000, 715429006, 722377004, 722961009, 722973009, 724508005, 725047007, 789017006, 792842004, 860814001, 866066004, 866114005, 84171000119106, 290621000119101, 290761000119103, 304121000119108, 313921000119106, 342821000119103, 346401000119108, 368581000119106, 442161000124105, 722762005, 1156763004, 1187563003, 1177034003, 15709361000119103, 337661000119105, 1259087007, 1284863004, 4223005, 19726003, 27277001, 38528001, 41510006, 52441000, 77645007, 87235005, 87842000, 129596006, 192650000, 193185007, 197661001, 206398001, 212173001, 230371005, 247204001, 253115005, 277130004, 307684005, 307756005, 397928009, 413921009,

425219008, 702363009, 709489006, 713911007, 715484003, 721430005, 721457007, 721474007, 770630005, 783242003, 866045007, 10481000119108, 292111000119104, 292971000119105, 303361000119103, 330521000119103, 336221000119104, 431131000124105, 436031000124102, 442151000124108, 1078081000119109, 1089781000119109, 13015821000119105, 15737321000119105, 204740005, 735387004, 431121000124107, 15628611000119101, 16019031000119105, 367411000119102, 1187470001, 1177054002, 1598991100011910 6, 1208614008, 1196844004, 1230019002, 1217383007, 1259531005, 1259565006, 6481005, 16026008, 26714005, 45339001, 45864009, 60389000, 63903007, 80495009, 84873005, 111005008, 128123007, 193212008, 204149000, 209900006, 230444006, 230806005, 237718009, 238977002, 247174006, 402487001, 404659001, 406569004, 418961007, 421931005, 429998004, 448788000, 699258007, 702398007, 711151004, 713831005, 715341003, 715565004, 717185008, 717859007, 720517001, 721015008, 721207002, 721472006, 721487001, 726083008, 733065003, 733650000, 763367009, 773501006, 773503009, 3371000119106, 291931000119102, 295511000119101, 347121000119105, 688191000119107, 1074111000119109, 24654003, 45639009, 421023003, 1148738004, 1149095004, 1179508005, 1217638003, 1231636001, 1259519008, 1259990004, 2109003, 4069002, 11197005, 11371006, 13920009, 24003004, 32112006, 34004002, 37960002, 40720005, 47417003, 62950007, 77436004, 92406008, 112107000, 186476008, 193230001, 196017002, 230357002, 237723009, 253168004, 277505007, 280871000, 302882002, 373587001, 403169003, 403671004, 445217002, 698621006, 700305007, 719276005, 719403003, 719430008, 719511005, 721504008, 724540009, 724546003, 763861000, 766764008, 766977007, 768666006, 771514002, 838528007, 19191000119106, 84201000119105, 302311000119107, 340941000119101, 349121000119101, 429861000124109, 432281000124107, 16054431000119107, 421998001, 1003444000, 1003448002, 1156836006, 1187147003, 1197757006, 1259469003, 1259675001, 21454007, 25690000, 37331004, 54359000, 78085003, 85039006, 85972008, 88219000, 109250009, 126945001, 126970001, 192819000, 195213000, 203991006, 230415009, 230441003, 238031009, 247131008, 254243001, 302904002, 392559009, 399081005, 403399001, 403401007, 403819009, 410014007, 415347009, 428703001, 441892008, 442668000, 447575003, 715422002, 719824001, 721384009, 724841000, 734065009, 735756004, 737340007, 762344002, 762351006, 762355002, 773394007, 773498006, 785301002, 866127009, 90811000119100, 135801000119109, 290521000119100, 291961000119105, 306591000119107, 329631000119108, 366951000119109, 15631251000119109, 16001911000119103, 16206381000119105, 128209004, 1003434002, 25521000119106, 683791000119100, 1172698005, 1197758001, 1217207008, 343001000119109, 1208615009, 1208970009, 1231628005, 1259667007, 1279834007, 20725005, 24500009, 26360005, 28055006, 37895003, 65242001, 95488001, 95723009, 111028009, 188298006, 188315007, 191454001, 195216008, 201276006, 203927003, 205583005, 212209007, 212253002, 230188005, 230458008, 230802007, 233756009, 233788001, 240568004, 253174004, 262727006, 276631005, 303061003, 312720005, 313915006, 372477008, 402628002, 403670003, 420351005, 441894009, 442701004, 443410001, 445496008, 449822000, 702441001, 719808002, 721458002, 724728006, 725407006, 735602002, 770721009, 771144005, 783009008, 783016009, 838350007, 82351000119105, 292991000119106, 329461000119102, 15697241000119105, 39912006, 360434004, 874939004, 1156035006, 1187047001, 1163072008, 1254960008, 1259787004, 1259628001, 1259652008, 1263518006, 1264189006, 1268342009, 5280003, 11862006, 13456001, 16726004, 19350004, 19871000, 54509008, 61778004, 72452005, 87043009, 95498007, 188296005, 192644005, 193003000, 209176003, 209845006, 212337002, 230504002, 230796005, 231913002, 239082002, 254978007, 312217003, 399100005, 402466006, 402610004, 406580004, 426570007, 428561000, 443353000, 457772007, 702375004, 709118002, 719986000, 721397006, 721429000, 721438003, 763069002, 763369007, 773622005, 82561000119101, 106021000119105, 340961000119102, 434151000124101, 434501000124107, 1081511000119105, 15681801000119103, 313791000119106, 1260049006, 12217441000119107, 1260381005, 2689001, 10277002, 20544001, 32552001, 50490005, 61165007, 67975003, 77817004, 95670000, 95699005, 128116006, 192715004, 212172006, 212260008, 229691009, 230226000, 230304003, 230418006, 230805009, 231439009, 239874001, 240463005, 268722008, 275945006, 315348000, 359619007,

372063002, 392133001, 402613002, 405758006, 425492002, 428700003, 442439008, 697944008, 721375009, 721511007, 722967008, 724990004, 734023003, 735597009, 764734003, 770431001, 773665006, 782670003, 789220007, 295571000119109, 1469001000004105, 1569281000119107, 62201009, 190615004, 230271008, 422126006, 1004092000, 878889000, 1156471001, 1156761002, 384430101000119103, 1163130005, 1186886008, 342971000119107, 1196898000, 1236805005, 1259656006, 1263985009, 1264188003, 4168008, 15059000, 17093002, 19334007, 22443004, 23501004, 36739006, 43132002, 43497001, 44219007, 54794009, 57148006, 62745008, 80980003, 86269002, 106170009, 127305005, 134193000, 186118006, 188340000, 192643004, 193837009, 204072001, 206446002, 208020002, 212355006, 230292008, 230584000, 233718008, 238636003, 254962005, 267889007, 399041003, 429347004, 609530001, 703284009, 717632002, 719947004, 721364005, 721386006, 722580004, 724226009, 724706006, 726702005, 763347000, 770755007, 771261002, 783616005, 783764008, 789002007, 870266002, 292241000119108, 292561000119109, 302391000119103, 322461000119108, 10901331000119109, 10943471000119107, 10999321000119108, 15676001000119105, 15692641000119106, 16002191000119102, 29422001, 1177177000, 1187547006, 1231189004, 1231727005, 1260330000, 16517004, 17496003, 45163000, 60404007, 79410001, 110030002, 127024001, 201849003, 212219001, 212264004, 230450001, 262698003, 262703009, 309246000, 425561001, 427571000, 446474007, 719209002, 721512000, 782884005, 870283006, 342581000119102, 348601000119109, 428051000124108, 690241000119106, 16058311000119100, 368251000119101, 1156473003, 1156823006, 1172588008, 1186871002, 1186718008, 1186828006, 1259708007, 1263449003, 8656007, 19735005, 26206000, 37770007, 39662004, 41564009, 42448002, 52289006, 60217008, 70073005, 95109002, 110270004, 126961004, 127298000, 192845009, 193101001, 194059004, 200985008, 202671008, 203949009, 206233005, 208059006, 229658000, 230149008, 230180003, 230609001, 232057003, 236403004, 237699005, 238972008, 253137003, 253172000, 282354001, 373590007, 402718003, 403449001, 403577008, 406616008, 408537003, 416071001, 426983002, 703286006, 707364007, 721374008, 721437008, 724001005, 724989008, 733469003, 735646005, 773330000, 783060009, 783094006, 840461004, 7761000119106, 17151000119108, 17701000119108, 103931000119102, 120401000119101, 291091000119102, 294011000119108, 341941000119108, 432171000124102, 1076391000119106, 15639281000119101, 16376391000119106, 1145219008, 1259101007, 1260387009, 1279563000, 17796002, 28864000, 34907007, 110990003, 111558006, 202688001, 206399009, 212280009, 212294005, 230295005, 230325003, 230691006, 253192007, 254185007, 254197008, 262743004, 276590002, 276592005, 277480002, 278286009, 309760004, 404233006, 417075004, 421847006, 422217003, 425556003, 429565004, 442024001, 609557004, 698783004, 711487002, 721221000, 721365006, 722795004, 724146008, 733033001, 734017008, 762305005, 771516000, 783010003, 838390005, 870287007, 291111000119105, 336941000119100, 15710401000119104, 16039071000119104, 16058271000119100, 25362006, 49049000, 239872002, 1010664005, 1163074009, 1187543005, 15665801000119104, 341411000119101, 1222586005, 682441000119105, 1237366005, 1259456002, 1263517001, 849000, 1534008, 6267005, 8829008, 12677003, 14786003, 93923002, 123706005, 127308007, 195230003, 202730006, 203033005, 203935000, 212293004, 212344006, 230186009, 230230002, 230349000, 230524003, 230597003, 230699008, 230797001, 246877005, 252794008, 253226005, 254187004, 254939008, 277478008, 389101006, 403536009, 403801000, 429743002, 442481002, 443265004, 444606000, 445897006, 699318007, 721372007, 721448001, 722108000, 723826007, 782744007, 783697000, 784352007, 789015003, 789063000, 13691000119103, 318651000119104, 338781000119107, 12239661000119108, 16058231000119103, 4863002, 43959009, 679881000119104, 1153378003, 1231824009, 1263524000, 1269368009, 16637291000119109, 16638481000119109, 40816002, 45795007, 50639002, 55016009, 57138009, 60676002, 69078007, 85672005, 95720007, 109381003, 109920008, 126969002, 128205005, 163590008, 191507002, 202733008, 230146001, 230620001, 230773005, 231452004, 240433001, 361130009, 408386002, 427789001, 703095005, 716112005, 716663009, 717016001, 719515001, 721196005, 722581000, 723644002, 724672003, 763348005, 782726004, 866096009, 1591000119103, 125081000119106, 313901000119102, 690261000119105, 368151000119108, 1173998003, 1260047008, 1264190002, 2438005,

10068001, 46960006, 49398001, 67531005, 68107009, 82401000, 82999001, 85076009, 92029009, 192722007, 202663009, 203951008, 206458002, 208009002, 212206000, 229659008, 229686008, 230225001, 230779009, 238836000, 238942002, 239868001, 246531008, 253737007, 262728001, 267574006, 282792003, 302877009, 387802007, 420585007, 425919003, 427027005, 439072001, 444024002, 708463003, 713739004, 718180000, 719810000, 723612001, 724686004, 725043006, 725590001, 763345008, 771419005, 788980005, 827172005, 140881000119109, 323161000119107, 1074131000119104, 1092321000119104, 15686041000119102, 15992671000119103, 16056271000119101, 110997000, 402446002, 1144223007, 1156340006, 686451000119106, 1163152001, 1186802008, 1187145006, 1186838001, 1197151003, 1231282002, 1259480003, 1259642009, 1260332008, 800703961000119100, 2584003, 26039008, 38467009, 42658009, 53889007, 71178008, 74737003, 193002005, 201627006, 201921009, 230261006, 230733004, 246536003, 247064005, 253348002, 267718000, 297147009, 298124005, 313437008, 361212009, 388872005, 397568004, 398102009, 403402000, 403663003, 418435001, 709144002, 719513008, 719516000, 763743003, 765481009, 773331001, 783257005, 785726009, 35531000119107, 112961000119103, 291471000119107, 294051000119109, 346701000119104, 12265261000119102, 15648821000119105, 15705521000119102, 15986111000119101, 15986151000119100, 77956009, 79385002, 420774007, 10954821000119108, 1148739007, 1187538003, 1179379002, 1163552008, 1177071001, 16008151000119109, 1231644001, 1259524006, 1260400009, 1263584002, 1269233006, 16639181000119106, 9491003, 14046000, 21018002, 39003006, 41783004, 47683004, 77173005, 77355000, 93931007, 95456009, 129616004, 189198006, 229693007, 230144003, 230157006, 230695002, 238861002, 240069003, 253225009, 277522009, 277523004, 285597000, 285600004, 309587003, 314840009, 360358001, 443971004, 700317009, 715464002, 715817007, 722944006, 724746000, 763127004, 772225005, 783244002, 295551000119100, 432511000124103, 1089711000119103, 12367511000119101, 16006631000119109, 16035831000119109, 78468005, 1163062005, 1172628002, 1217059006, 16039511000119104, 1259661008, 27152008, 37711000, 58766009, 73221001, 88032003, 90493000, 91959004, 93727008, 95653008, 95776004, 111510002, 126953009, 190644002, 192761004, 201829007, 208037009, 230262004, 230410004, 230716006, 231919003, 238974009, 246927006, 255124004, 278852008, 282741006, 311891006, 403531004, 445252005, 449420002, 698292000, 698616008, 715345007, 720518006, 721379003, 721661009, 722456001, 722718001, 724812009, 735391009, 771302009, 782949007, 783008000, 838530009, 840489001, 866057003, 870317001, 84191000119107, 89441000119109, 135311000119100, 142011000119109, 291921000119100, 292331000119106, 337701000119103, 341451000119100, 368961000119107, 29610001000004107, 1156459007, 1156964000, 352771000119107, 1263557003, 1263961001, 1269361003, 1285322008, 16638551000119101, 10491005, 11023001, 22607003, 33147008, 38353004, 48233004, 60490008, 69880002, 71325002, 72858000, 83605009, 87132004, 93929003, 95696003, 127310009, 163594004, 195373009, 202693003, 212201005, 212314008, 229682005, 230748005, 231972004, 238632001, 253164002, 276703007, 298118004, 299966003, 312787005, 386572005, 389271000, 402458002, 427943001, 431043000, 442097001, 702949005, 715768000, 717054001, 721228006, 721331005, 722285005, 722625002, 722659003, 722931001, 724604003, 724811002, 762352004, 765403009, 768939009, 773644000, 774069007, 782917007, 787408008, 860851007, 866061009, 870540001, 18801000119100, 341861000119100, 431161000124101, 672371000119103, 15699161000119109, 193687000, 713060000, 890118006, 1186875006, 1197298003, 101611000119103, 1263984008, 5134006, 11807002, 23849003, 33941008, 42594001, 46963008, 50026000, 74469006, 75968004, 77827005, 109911004, 130990006, 209195005, 212248007, 212263005, 230191005, 230382002, 230394006, 230439004, 230769007, 231440006, 240052000, 277798002, 282796000, 287062005, 328681008, 403569009, 408750000, 414400006, 422527005, 445158000, 445779008, 698618009, 719140001, 722127006, 722671009, 722977005, 723822009, 763349002, 783243008, 12551000132107, 90011000119105, 94601000119107, 130121000119104, 151161000119102, 339651000119100, 431141000124100, 434761000124100, 10962471000119106, 15992311000119100, 16026511000119107, 16039111000119106, 1010464002, 1010712009, 344261000119101, 679111000119101, 1145224006, 1167373005, 1187552001, 1208972001, 1269414003, 16637971000119104,

4412009, 19378003, 29753000, 33411003, 85262003, 91327001, 126987004, 193179000, 199093003, 203957007, 209920007, 212235008, 212322001, 230316009, 230559003, 232281004, 237714006, 237873000, 247138002, 313434001, 363476006, 370502006, 398760006, 403809003, 713703005, 715645004, 716099003, 718611007, 719184001, 723161003, 724813004, 726617002, 766753005, 773688007, 778011005, 783703004, 292271000119101, 303901000119100, 321311000119108, 429271000124103, 1074121000119102, 1239828100011910‌5, 16839401000119104, 66695004, 127294003, 1162319009, 1259494002, 1259600007, 813925041000119103, 828066161000119106, 640003, 26037005, 34601006, 38576000, 49823009, 71104009, 92503002, 192687008, 192818008, 193072006, 195518005, 205294008, 209051002, 209093001, 212255009, 230344005, 237670000, 238782003, 247400008, 253227001, 267581004, 274156000, 276285004, 281364007, 285764004, 298133007, 402679008, 418254004, 439567002, 448250003, 450362008, 698762005, 700314002, 715462003, 718850008, 720460007, 721746001, 722003007, 724141003, 724747009, 724998006, 735616007, 737006004, 763310000, 763834000, 764944006, 771182002, 774205007, 778021002, 782309008, 788631003, 870291002, 5181000119108, 303191000119108, 341141000119106, 1082781000119102, 12236641000119103, 13015741000119105, 16005271000119104, 781641005, 15705401000119105, 1003380001, 1003422005, 1162362005, 1259586006, 1263540002, 22386003, 32916005, 49255002, 49840000, 51399001, 63864004, 74107003, 77420001, 79633009, 89437009, 95644001, 95655001, 95660002, 109477002, 206191000, 212222004, 212306002, 212358008, 229685007, 230246005, 230808006, 231968003, 238706002, 262710003, 262736005, 404238002, 406598000, 418072004, 427916006, 441735003, 715827001, 716998009, 721387002, 721447006, 763889002, 766821006, 770759001, 783550006, 785808002, 788417006, 838384006, 391000119105, 113021000119102, 147131000119101, 313891000119101, 342741000119103, 431081000124105, 687571000119101, 688131000119108, 1074511000119102, 15720481000119102, 16002271000119109, 890288008, 1186873004, 335851000119101, 1236975007, 1260008006, 1279707003, 1767005, 7611002, 9740002, 42944009, 46091002, 55702009, 62564004, 66637005, 86022000, 191394000, 195210002, 206188000, 209098005, 212185009, 230189002, 239143001, 246532001, 277303004, 308143008, 402723003, 457732003, 699299001, 700315001, 700333007, 712645006, 716281000, 717968005, 719187008, 721432002, 722969006, 724683007, 726622002, 762332000, 768639006, 773497001, 840496004, 106241000119108, 122731000119104, 290671000119100, 311191000119109, 313801000119107, 329721000119103, 347261000119106, 451041000124103, 15740841000119107, 15968701000119102, 15992271000119100, 40956001, 72820004, 766834007, 1156825004, 1186907003, 1230014007, 15665881000119107, 1263493003, 2041006, 10256000, 17996008, 26609002, 29702001, 31204009, 37605006, 68504005, 81442004, 94725006, 94986003, 95701005, 109682000, 127312001, 192982004, 194119004, 208071000, 229687004, 230264003, 230670003, 239026002, 239079007, 247184007, 249405005, 308756007, 361268000, 422736007, 428609006, 609553000, 702753003, 707510005, 716169009, 719818007, 721516002, 721528005, 725408001, 737339005, 783005002, 290191000119109, 291631000119107, 294121000119100, 309011000119101, 690281000119101, 15685201000119100, 69742007, 238044004, 445198003, 1172692006, 15997831000119104, 1259465009, 1279967005, 843004, 7916009, 10651001, 15733007, 16300007, 50805004, 69646003, 74267005, 81634008, 188293002, 194842008, 209193003, 212242008, 229689001, 230192003, 231920009, 238018004, 363483004, 442181008, 443524000, 703217005, 720825005, 721442000, 721842008, 722583002, 723021001, 733417008, 733926004, 735750005, 737236000, 770405003, 838322007, 860842004, 116821000119104, 140911000119109, 292011000119103, 343471000119108, 429841000124105, 1077931000119106, 16013351000119104, 271730003, 1156852009, 1217055000, 1234781009, 1259485008, 1260353004, 1279838005, 721251000124100, 4805006, 8526009, 23186000, 30057005, 46303000, 59252009, 95240001, 129573006, 191499009, 192965001, 193956000, 195211003, 197294009, 203946002, 204081007, 208003001, 212359000, 230427007, 230447004, 230623004, 230777006, 232113004, 238898001, 253111001, 267613004, 277949001, 359634001, 361151007, 406596001, 420675003, 448227009, 450376009, 702314005, 711443006, 717813005, 718602007, 722600006, 765747004, 770761005, 773643006, 788880006, 40121000119107, 336201000119108, 87555007, 90036004, 193141005, 672461000119104, 1149087003, 1156457009, 1186855002, 1163529009, 1162256002,

1179455003, 1197365006, 1179360000, 1237420004, 1251417006, 1254861008, 1259529001, 1259605002, 1734006, 2665008, 9015001, 38599001, 38837006, 41250005, 58170007, 67869008, 84438001, 87536007, 92136007, 92247009, 163919004, 192876003, 192949002, 193409008, 198299001, 209066004, 212278003, 212285004, 212333003, 230310003, 230406002, 230508004, 230634001, 230807001, 237595008, 240046001, 253163008, 308758008, 360527003, 402469004, 403265002, 711162004, 721363004, 721434001, 721973006, 722619001, 724466005, 724717007, 782723007, 782742006, 783055005, 284881000119108, 329361000119107, 433681000124102, 435361000124100, 680801000119107, 15985791000119101, 15990471000119102, 16207031000119109, 16641561000119107, 237512007, 1003408005, 1186835003, 1162826002, 1204350002, 1197593006, 1231233002, 1222656005, 1263549001, 3652007, 15552004, 33000001, 33559001, 37340000, 46855008, 50541007, 71105005, 75068001, 86044005, 95640005, 95697007, 111681000, 126958000, 129137006, 129608008, 191451009, 191480000, 202686002, 208051009, 229684006, 230319002, 230397004, 230610006, 237578000, 240055003, 268146009, 268650001, 371121002, 371129000, 403804008, 429466000, 449902003, 715482004, 721398001, 721482007, 722379001, 722518004, 722997000, 723153008, 723580007, 723992000, 771262009, 771304005, 774154008, 782727008, 785306007, 2691000124105, 303261000119107, 329391000119100, 436601000124101, 15685121000119100, 15704361000119102, 15731601000119100, 15737921000119106, 15972821000119108, 15985031000119102, 16016511000119102, 16026471000119103, 16419781000119100, 17570001000004102, 31848007, 89062002, 270505009, 312927001, 350841000119107, 682471000119103, 1187191003, 1186800000, 1177002007, 1208933000, 1259827009, 19833008, 23728006, 40307005, 50076003, 50808002, 56529007, 66073004, 68761002, 69501006, 93962006, 95375003, 95662005, 111197009, 188286001, 188295009, 191509004, 192811002, 192906003, 193180002, 193203006, 198991006, 204043002, 209049001, 230724001, 246764005, 248260009, 253152005, 276878008, 307767006, 370987005, 404241006, 414662006, 429432009, 716706009, 719188003, 721466006, 723452007, 724673008, 778002005, 783148005, 783726006, 789021004, 116401000119105, 329691000119107, 343251000119100, 344231000119109, 434141000124103, 688111000119103, 1090231000119107, 1149280009, 239965291000119107, 1186887004, 1186721005, 10972941000119103, 1217382002, 1237619001, 1254909007, 1259088002, 1259611004, 1269237007, 16024071000119102, 3458000, 12286000, 15123008, 41283003, 42429001, 67747009, 69875006, 77375009, 81854007, 82331007, 85761009, 95722004, 128431009, 129583005, 187080002, 193589009, 205425003, 208017005, 212303005, 230155003, 230185008, 230318005, 230381009, 230608009, 239135006, 239883006, 241994005, 275462005, 276674008, 307766002, 361117008, 403419003, 428768008, 702316007, 718103001, 722622004, 723862008, 724562009, 726618007, 733094005, 763377006, 764939004, 771448004, 773398005, 773769008, 782680004, 860825005, 860844003, 67771000119102, 84211000119108, 335871000119105, 432021000124104, 434321000124107, 10999361000119103, 15699241000119102, 16024271000119107, 78631004, 79268002, 403784003, 733173009, 429161000124103, 1172584005, 1163537001, 1163482004, 1186728004, 15997751000119100, 1237365009, 1269362005, 1269378007, 16636451000119101, 13044008, 13694005, 28736004, 41133002, 48611009, 52702003, 59103002, 62260007, 109936005, 163930001, 190646000, 190842000, 191459006, 191502008, 192964002, 193196004, 195185009, 200984007, 202670009, 203942000, 206443005, 209078000, 212258006, 230161000, 230302004, 230414008, 230760006, 240461007, 253224008, 267638009, 277461004, 290653008, 299737009, 402470003, 404243009, 423628002, 423683008, 442155009, 449916008, 699190008, 720990007, 721389004, 721453006, 763688008, 766049000, 773583007, 866065000, 290221000119103, 292041000119104, 292221000119102, 292571000119103, 333881000119109, 1078061000119100, 15675721000119105, 15710521000119107, 15960141000119102, 792004, 680491000119103, 894125007, 1179028000, 1163151008, 1187251009, 1162864000, 1217381009, 1208340009, 1251446004, 1259039002, 1259425005, 1260231004, 1260335005, 6807001, 7180009, 21350002, 30005005, 30085007, 42440009, 47673003, 49765009, 51603000, 52330001, 53406005, 89261000, 94317006, 111497000, 129609000, 186378005, 209827006, 230255008, 230590001, 230784003, 237663000, 240178007, 240448007, 253876004, 294705005, 302894006, 307683004, 403394006, 406568007,

418054005, 419051009, 423023005, 423341008, 426634003, 702423009, 709584004, 711403001, 712857000, 715504003, 715981004, 721505009, 721821006, 724565006, 724685000, 724765008, 724780002, 724792002, 724993002, 735564008, 762297005, 764959000, 771076003, 773668008, 788933002, 866251007, 8423100019103, 124061000119103, 292391000119105, 302331000119102, 330751000119103, 431151000124103, 679131000119106, 691461000119103, 1074561000119104, 15692601000119109, 24285001, 92824003, 352911000119101, 1163548008, 10973041000119109, 1208346003, 1259739009, 1259815006, 1260043007, 16732921000119107, 1259689006, 1263506002, 1264131000, 1284864005, 2231001, 17393007, 54280009, 63819001, 69093006, 72669000, 76896001, 164122006, 188312005, 191714002, 193880000, 195212005, 201927008, 230216007, 230421008, 230535008, 237671001, 238092004, 238878006, 239870005, 254132000, 254188009, 267664003, 291665000, 313258000, 398900000, 402465005, 403294009, 403856000, 406564009, 430959006, 441622000, 700380004, 712858005, 717008005, 721450009, 721846006, 724707002, 724768005, 762327001, 763534009, 870338007, 117891000119100, 348431000119100, 680991000119108, 1082511000119102, 12237911000119109, 15686081000119107, 15960581000119102, 16013871000119104, 16238221000119109, 16415361000119105, 426470009, 15705601000119107, 10899711000119108, 1144916002, 1155933006, 1173036000, 1186833005, 1163071001, 302681000119107, 1234772003, 1259043003, 1263562002, 1269366008, 7680008, 9112001, 14266003, 15316002, 20610004, 31097004, 43972005, 44434003, 51075009, 56177003, 60317006, 63600008, 66521008, 68390005, 78689005, 95651005, 127301001, 128124001, 212187001, 212246006, 230368002, 230407006, 230509007, 230787005, 230788000, 262713001, 268054009, 270487001, 278467003, 402467002, 424334007, 424358006, 440443001, 445135003, 446644006, 609600000, 700318004, 703537008, 716722005, 718177001, 723123001, 724566007, 724670006, 763376002, 771142009, 783555001, 787177005, 789001000, 33581000119109, 308951000119109, 343481000119106, 679211000119106, 15710361000119108, 1172605003, 1177043007, 1177062005, 1204170004, 1204144000, 1208477000, 1254937000, 1263552009, 23056005, 25053000, 27195007, 46975003, 52866005, 76889003, 77097004, 77489003, 79220008, 83465002, 83886009, 188326001, 193138001, 193466003, 203936004, 230174006, 230660000, 236405006, 238946004, 301764006, 302885000, 312429005, 402692006, 403392005, 418402006, 421468001, 428061005, 447253004, 709285002, 719250005, 721521004, 722979008, 723501008, 733194007, 737199001, 737233008, 762339009, 762506007, 770626007, 133951000119104, 290371000119105, 330301000119103, 330721000119106, 1082421000119105, 15997671000119100, 16052591000119103, 1003378007, 1144902007, 1172844009, 1186806006, 15675681000119104, 1259765008, 1263512007, 4088009, 4103001, 13196008, 26538006, 28192008, 32798002, 34139004, 34476008, 41222005, 49384009, 49776008, 51984006, 58795000, 63814006, 95664006, 109912006, 123670005, 127025000, 191496002, 197315008, 206471004, 208043006, 212313002, 230166005, 230518009, 230587007, 232143002, 237685006, 238091006, 239877008, 239878003, 240122006, 254972008, 297945000, 402435000, 419868009, 442733008, 449795002, 713454009, 713571008, 715344006, 720632004, 723138000, 723155001, 724000006, 724725009, 726608002, 771223000, 788930004, 830032008, 870293004, 106071000119106, 348131000119109, 378007, 64383006, 1156813002, 1222661007, 1208935007, 1231197006, 1237511005, 1259100008, 1259828004, 2776000, 11413003, 21590003, 27982003, 32875003, 48248005, 54404000, 65120008, 73495003, 80420005, 86923008, 111242007, 194043004, 202657000, 209068003, 212183002, 212226001, 230190006, 282789002, 402459005, 403259002, 404653000, 419197009, 420662003, 424271007, 427787004, 428027008, 444856008, 609531002, 698073006, 698737005, 698838008, 713895003, 715317001, 720634003, 721393005, 724225008, 762282007, 763776004, 765091006, 765197008, 770430000, 770724001, 782914000, 788529004, 101461000119102, 285141000119106, 346691000119104, 15722961000119101, 128083007, 186591007, 926304005, 1172691004, 1186870001, 302701000119105, 1197357008, 1259511006, 7794004, 25772007, 25999001, 40425004, 46459009, 49793008, 80690008, 81308009, 88845000, 89019003, 91953003, 111527005, 129136002, 203981000, 209069006, 209173006, 209188001, 230274000, 232050001, 251002009, 253150002, 262960001, 268143001, 284571002, 302881009, 371562001, 402611000, 402730009, 404040002, 422426003, 445151006, 609529006,

698846009, 715801001, 715980003, 721401003, 725392005, 773672007, 778001003, 778048001, 782388003, 866097000, 870286003, 21601000119103, 295621000119107, 313931000119109, 329711000119105, 431961000124104, 16026751000119100, 4338008, 1052237009, 680511000119108, 1186932001, 1163551001, 1186842003, 817968741000119105, 1259123006, 1259522005, 1263495005, 10613001, 10736002, 18834007, 22383006, 40571009, 41083005, 41362007, 67873006, 79745005, 81774005, 81981006, 94448002, 109932007, 192976002, 195032004, 201639003, 202695005, 230621002, 232052009, 233748006, 237673003, 253190004, 254191009, 271969004, 276593000, 281864001, 314130008, 371116000, 403816002, 416903004, 438583008, 449901005, 609601001, 700335000, 715507005, 715630006, 715824008, 716379000, 721402005, 721525008, 724777003, 733467001, 737225007, 770725000, 782695002, 785723001, 789430001, 290141000119101, 330781000119105, 343101000119108, 1074491000119107, 16039231000119106, 16058071000119105, 404906000, 698279003, 1186710001, 1254890002, 1263541003, 1264191003, 6729006, 7033004, 17169009, 37660004, 40802007, 51493001, 52897009, 54593000, 70534000, 134209002, 186480003, 186863009, 192874000, 193119005, 195190007, 212193009, 212308001, 230250003, 230652001, 234188002, 237612000, 238606007, 262955000, 307688008, 370509002, 402449009, 402948006, 403640008, 403689003, 442191002, 444978000, 449905001, 717332007, 721200000, 723139008, 725168006, 763350002, 763460007, 771303004, 871646007, 18321000175101, 67661000119100, 87091000119101, 293901000119108, 311231000119100, 344551000119101, 431421000124103, 431471000124102, 432571000124107, 15990031000119108, 163605002, 447262002, 1052210007, 1090881000119109, 1148923005, 1153331004, 1177069001, 1187278006, 1231284001, 15704401000119106, 1231202004, 1231184009, 1237337007, 1259832005, 1259638006, 1268341002, 1285519007, 26718008, 50325005, 51169003, 61927004, 64815002, 69071001, 91950000, 92048008, 94971009, 95171000, 193134004, 195206000, 212230003, 231444002, 231449007, 231948009, 233763009, 239867006, 246602002, 253148005, 254950006, 298278003, 312942003, 313003009, 363474009, 402447006, 402450009, 406571004, 413808003, 428960009, 441604003, 441991000, 447232008, 698870008, 703622004, 715840006, 717827000, 719512003, 721443005, 721532004, 724172004, 773275000, 773664005, 789187001, 336331000119108, 342831000119100, 348051000119102, 432461000124101, 672561000119103, 1074211000119104, 16583281000119101, 879919001, 1163092001, 1201961000, 1231226002, 1196860000, 1231737000, 1259102000, 1259488005, 1269493005, 13386009, 19445006, 22451001, 50582007, 51568001, 54099005, 55713007, 64749001, 93748005, 192781003, 212231004, 229666009, 230288001, 230293003, 230713003, 231934009, 234139002, 238837009, 238897006, 246769000, 250978003, 254756007, 301009006, 402952006, 403507009, 403649009, 405759003, 427111000, 446835008, 698754002, 717052002, 721503002, 723163000, 723940009, 724681009, 724821005, 735606004, 762281000, 765331004, 770720005, 773554009, 773663004, 302321000119100, 304281000119100, 690251000119108, 16002471000119108, 71253000, 73751008, 75739009, 1144895003, 1148888004, 1187549009, 1163060002, 1186852004, 1172626003, 1259743008, 1260343000, 1263560005, 18191000, 18871009, 21237001, 30385002, 32390006, 44759006, 54304004, 60342002, 63510008, 73013002, 111000003, 111395007, 128119004, 129667001, 200078007, 206469004, 230296006, 232280003, 233955003, 235041006, 236444003, 246771000, 247181004, 249892007, 285599002, 309279000, 398925009, 406620007, 427945008, 431769004, 442617003, 713705003, 716699004, 719833004, 762514001, 771470001, 309021000119108, 335351000119105, 679691000119100, 1090871000119106, 12275031000119106, 15705561000119107, 15732921000119109, 15737041000119101, 15930341000119109, 16891111000119104, 350621000119100, 1187048006, 1163469007, 1234780005, 1259636005, 1263665007, 1269389000, 20022000, 26554009, 34406007, 35786001, 42369001, 77797009, 111406002, 127295002, 191990001, 192724008, 202672001, 203382004, 212234007, 212257001, 230154004, 230214005, 230431001, 230626007, 236528009, 238703005, 307649006, 398071000, 399336001, 424353002, 441951003, 703216001, 711482008, 722655009, 723930006, 762617003, 792799000, 866063007, 134811000119108, 340251000119108, 428351000124105, 1099031000119105, 12398321000119100, 15630241000119104, 16019511000119108, 16414611000119100, 1187620007, 1187205008, 1197429000, 1231423008, 336421000119100, 1260360005,

9526000, 32415004, 35777006, 38513001, 43898003, 52888005, 67144006, 72494008, 83942000, 95002008, 111389006, 163591007, 188322004, 192871008, 194064000, 230693009, 230780007, 240063002, 240429008, 249411008, 254193007, 282797009, 299967007, 300992002, 305719002, 310202009, 402615009, 402626003, 403775003, 413102000, 422088007, 432991000, 442077006, 698948009, 700325006, 713692003, 713861002, 724576005, 734066005, 763375003, 778044004, 782879004, 866095008, 870321008, 134891000119104, 333661000119101, 347581000119104, 15930381000119104, 15675961000119102, 1153408000, 1163068009, 1163099005, 1231683004, 1231757001, 1260044001, 5387003, 7689009, 12589008, 15244003, 16341002, 21258007, 35691006, 41119002, 44248001, 51174006, 82793005, 91960009, 95792009, 109915008, 111488004, 128195009, 186738001, 192701001, 212218009, 230585004, 253194008, 301086002, 307696003, 403503008, 403542008, 403595006, 417658006, 428753007, 609218006, 698620007, 698765007, 709203006, 715798007, 715984007, 716197003, 717010007, 718769009, 763373005, 782696001, 784342008, 866129007, 97381000119100, 291351000119109, 291861000119103, 292901000119100, 293921000119104, 308981000119102, 431641000124107, 678681000119108, 15705241000119106, 16005231000119102, 2355008, 1142203007, 1179295004, 1217379007, 1220597000, 1258934002, 514793471000119104, 5251007, 18842008, 20385005, 33033006, 45021001, 79442007, 92052008, 94309003, 126950007, 127011001, 128328009, 162702000, 189484008, 198037006, 201630004, 209080006, 230576004, 235044003, 238822005, 238867003, 240458006, 246541006, 312839008, 402491006, 444163006, 698742002, 703052006, 721814006, 724206005, 724730008, 734031008, 782746009, 788934008, 838363001, 21391000119102, 141991000119109, 290481000119100, 292911000119102, 431991000124107, 1074081000119104, 16276361000119109, 16320831000119100, 1010642001, 682221000119103, 1187049003, 1179380004, 1177176009, 1169367003, 15704441000119108, 1220574003, 1260337002, 2593002, 9441002, 13141002, 14784000, 16319002, 21263006, 21954000, 38645004, 42653000, 95635002, 123760002, 127306006, 128190004, 192454004, 192717007, 192915005, 193225000, 206578007, 208048002, 230475002, 237678007, 247146001, 247155003, 262734008, 278284007, 312838000, 312915004, 315046005, 423144007, 429668008, 430977001, 444991004, 445947008, 702317003, 709073001, 721253009, 721332003, 721467002, 724693000, 733096007, 736321008, 737232003, 763793004, 765401006, 767448007, 770438007, 771418002, 773577009, 788958007, 840440006, 840465008, 82501000119102, 332101000119100, 16846261000119106, 1010630006, 732330391000119107, 1187465008, 1186803003, 345161000119108, 1217660000, 15665841000119102, 1259097003, 1281838003, 3456001, 14900002, 18358003, 19598007, 21233002, 29774004, 37739004, 38428003, 63576006, 82178003, 92072003, 93249003, 109937001, 110999002, 111506000, 126947009, 190869004, 204004001, 212327007, 230389006, 230534007, 230591002, 230741004, 232076001, 233755008, 235045002, 237900002, 240131006, 275468009, 278834008, 286552001, 304838007, 308232009, 314780006, 371026009, 423234004, 445006008, 699259004, 702379005, 703304005, 709411004, 719981005, 721460000, 721500004, 724091002, 782720005, 783175003, 838319005, 838387004, 838391009, 290691000119104, 292321000119108, 303551000119107, 311951000119102, 16002351000119105, 16637451000119103, 420554003, 447378002, 1084801000119107, 1142100001, 1144893005, 1187043002, 1259103005, 1263958002, 1259003, 3261002, 10340008, 16901001, 22719003, 23291008, 36071006, 38795005, 53050002, 58856009, 59017008, 59292006, 74934004, 75111000, 111569009, 191452002, 192744002, 192900009, 203982007, 203987001, 230570005, 230618004, 262723005, 277333006, 307695004, 312899005, 371041009, 402457007, 403515007, 403682007, 406578005, 407472000, 414063002, 419900000, 427055009, 441794001, 702442008, 715821000, 717943008, 718756005, 721362009, 722995008, 724718002, 724797008, 726610000, 763351003, 763457000, 765746008, 770723007, 783618006, 784201008, 789000004, 840471002, 18761000119108, 102831000119104, 292931000119107, 323291000119108, 338321000119109, 432271000124109, 434991000124105, 448701000124105, 680791000119106, 1074201000119102, 15730041000119100, 230552007, 275884002, 1003922004, 1172591008, 16917201000119106, 570911000124104, 16415791000119104, 1263539004, 12195004, 24950005, 48638002, 55234004, 60935008, 163604003, 186478009, 195217004, 202661006, 209084002, 230612003, 240077004, 246533006, 274158004, 314538009,

371076006, 372062007, 389089009, 398446008, 417395001, 418541005, 420499009, 432733005, 444657001, 707221002, 718720007, 721873007, 722607009, 724786008, 724799006, 725139005, 735914007, 762326005, 764686003, 783698005, 785303004, 788908000, 838276009, 838388009, 846621001, 866140004, 18311000175105, 292801000119105, 336501000119105, 15699121000119104, 58756001, 123534001, 191471000, 442344002, 771184001, 1148631003, 1153428001, 849488701000119104, 1186798007, 1208890001, 1201863001, 1204173002, 1231203009, 1231185005, 1259622000, 1260329005, 1281833007, 16064691000119103, 24392008, 25094008, 34361001, 38692000, 41142009, 43300008, 44600005, 47924005, 49176002, 53194005, 63702009, 80116000, 86188000, 88697005, 94720001, 111521006, 111851005, 128196005, 192970008, 193177003, 202684004, 212148001, 229680002, 230273006, 230537000, 230645003, 230646002, 230647006, 232072004, 237351003, 238024005, 240074006, 276064006, 276826005, 276828006, 304831001, 307357004, 312917007, 428705008, 449253005, 699813007, 717011006, 720636001, 720855003, 721254003, 723929001, 724669005, 735757008, 860810005, 860840007, 9631000119102, 102781000119107, 291941000119106, 293871000119108, 330261000119106, 334051000119107, 334651000119101, 335331000119104, 433961000124100, 434211000124108, 15631291000119104, 9936001, 446957000, 791000119109, 851365731000119106, 1169358003, 1187249005, 1208876004, 1259062000, 1259095006, 1259831003, 1263534009, 1279566008, 14415006, 16873003, 21524000, 30491001, 52620003, 53267002, 56194001, 89704006, 93807001, 95203009, 202729001, 212360005, 225655006, 230794008, 254150007, 262729009, 266133006, 285760008, 363467004, 421182009, 446495002, 609528003, 702343002, 703156006, 722212004, 724072002, 724726005, 724781003, 724793007, 725394006, 733604003, 783160006, 781000119106, 4211000119108, 290311000119102, 294131000119102, 345831000119106, 688181000119109, 16035991000119101, 193410003, 204741009, 237961001, 403785002, 1156413005, 1186709006, 1172629005, 1196837008, 15709761000119102, 1234819007, 1259764007, 1259621007, 1264130004, 195772021000119105, 7339009, 27382006, 29322000, 32009006, 39020005, 43424001, 82959004, 93743001, 95582003, 109474009, 128491006, 186217006, 191962000, 193415008, 193598007, 200981004, 204125003, 213209000, 230306001, 230513000, 230572002, 253725005, 253743009, 254255007, 254981002, 263179007, 274157009, 277922001, 303063000, 402492004, 404689008, 414667000, 416069001, 441529001, 448705004, 448995000, 702450004, 713320007, 715487005, 721444004, 721446002, 721478005, 732949006, 763837007, 763895001, 838312001, 866085001, 866124002, 866143002, 14401000119109, 291101000119107, 306371000119100, 339261000119101, 341481000119107, 678821000119108, 1074191000119100, 16039311000119109, 16219341000119105, 129665009, 416975007, 733193001, 1172694007, 1162642006, 1255639008, 1259730008, 1259818008, 1259595003, 330731000119109, 25604001, 35055000, 41870008, 56684007, 64370005, 73877009, 82893001, 94451009, 109941002, 188339002, 192967009, 193213003, 201855008, 203003003, 204788008, 212275000, 230649009, 237701005, 238832003, 238845004, 254152004, 277879009, 282794002, 403309008, 403723007, 413758000, 415147002, 441698009, 445512009, 699866005, 700238000, 700342000, 718576001, 719162001, 719249005, 720522001, 721394004, 722294004, 763821001, 784573006, 866128004, 870565004, 284871000119105, 291991000119103, 330741000119100, 451361000124102, 15722561000119106, 230291001, 1177165005, 1177019009, 10852821000119109, 1222704008, 1201947005, 1237626001, 1259629009, 1259644005, 1264192005, 4275001, 11676005, 14070001, 18894005, 20900005, 27956007, 32599008, 36785009, 40259002, 48257004, 51928006, 68982002, 69909000, 70611002, 75962003, 82088000, 89410007, 92517006, 111200005, 200066008, 208022005, 230228004, 230328001, 230343004, 230758009, 236445002, 247054001, 248861000, 253108002, 254979004, 359824007, 363677007, 365258000, 397827003, 402448001, 438156004, 441630004, 703177009, 711409002, 713278001, 719251009, 722213009, 726615005, 762338001, 838357005, 68121000119103, 288631000119104, 311931000119108, 430851000124104, 454041000124101, 678891000119105, 12266201000119107, 15735721000119100, 36454001, 881694631000119107, 343431000119105, 1208934006, 1208974000, 1231677002, 12217401000119105, 1260385001, 7025000, 9031009, 16060001, 16644004, 18377001, 34184002, 40405003, 59918000, 62585004, 72100002, 74073002, 80577009, 84900008, 206442000,

209148006, 212211003, 212287007, 212318006, 230372003, 231927007, 232115006, 237675005, 238153009, 238979004, 254186008, 264443002, 277476007, 403627003, 417496004, 421029004, 447351004, 702318008, 703166003, 703215002, 721753005, 721826001, 722036008, 722599008, 722974003, 788967007, 789047008, 68151000119106, 113031000119104, 135491000119100, 135511000119105, 291741000119108, 292081000119109, 292101000119102, 292141000119100, 292691000119103, 329671000119106, 333171000119104, 429741000124103, 430871000124109, 10899751000119109, 15733081000119104, 15985351000119105, 1156415003, 1156962001, 117776611000119102, 1237421000, 342101000119104, 1259560001, 1260126007, 1263497002, 1268719004, 6595006, 13289004, 22133005, 23697004, 39450006, 48732008, 51638003, 57406009, 58258004, 68764005, 75023009, 111299004, 230522004, 230650009, 230702001, 232459006, 253176002, 282803008, 282805001, 417872000, 420090003, 441460004, 442020005, 472835004, 702376003, 703862003, 716170005, 716662004, 719301002, 719514002, 724385009, 733190003, 735611002, 737228009, 788957002, 860849008, 870308001, 1531000119102, 100731000119107, 104431000119107, 342291000119102, 10861171000119102, 15636211000119100, 15983991000119100, 46808003, 231974003, 421315006, 430397002, 1155717003, 1163098002, 1177058004, 1163535009, 1162307001, 1230018005, 15709721000119107, 1259072002, 1260140008, 1259517005, 1259687008, 1263933007, 1268960003, 1275628006, 241006, 4724003, 4952001, 6118003, 28188001, 41454003, 66020004, 72620002, 73935008, 77949003, 81526008, 90182005, 95656000, 109930004, 109943004, 212216008, 230435005, 230473009, 230605007, 230746009, 233715006, 238895003, 246611002, 262714007, 262725003, 262746007, 275360003, 280994000, 301643003, 302913000, 313294001, 359611005, 373623009, 400944001, 402732001, 403254007, 403395007, 422338006, 425882004, 447012002, 451039008, 716107009, 717893006, 719297006, 720819006, 723503006, 735389001, 788873009, 789005009, 838315004, 838394001, 870345007, 153091000119109, 292651000119108, 303171000119107, 336251000119107, 433671000124100, 681041000119106, 1085371000119105, 1092771000119104, 15705281000119101, 16039791000119101, 16052351000119101, 32680009, 188174841000119103, 1187621006, 1179376009, 1177020003, 1186863001, 1186850007, 1208968000, 1259706006, 1269048009, 29956001, 47999008, 54069001, 55051001, 68675004, 72836002, 80381005, 85641006, 90630009, 127304009, 192654009, 192743008, 195169004, 201274009, 208064005, 212356007, 230604006, 230793002, 232065000, 277496004, 277796003, 285755009, 371114002, 402805001, 404664002, 405765003, 419982005, 441717007, 442240008, 449873008, 609611008, 702347001, 718178006, 719302009, 721491006, 722006004, 765745007, 784344009, 838351006, 89511000119106, 348531000119101, 450921000124103, 15631051000119101, 16005071000119109, 25671008, 278065000, 10861251000119104, 1156848009, 1163104006, 1163100002, 1186807002, 1162338000, 1177055001, 1204349002, 1231274002, 1260390003, 1263581005, 15139001, 24059009, 35386004, 51806005, 64246009, 84170006, 87486003, 95390004, 109788007, 109946007, 111479008, 192072002, 192817003, 203950009, 212145003, 230383007, 230569009, 230798006, 232056007, 239866002, 254936001, 262741002, 276827001, 398106007, 402431009, 432249006, 702323008, 702445005, 702611008, 713503007, 718753002, 721405007, 724572007, 763218005, 765753004, 776087007, 778063003, 783014007, 789542009, 789704001, 870548008, 22381000119105, 339141000119108, 342751000119101, 344611000119101, 679701000119100, 1082621000119108, 16371781000119100, 21978005, 43405004, 58610003, 1157072007, 1187539006, 1187144005, 1187250005, 334931000119102, 611641000124103, 1231661003, 1259098008, 1259105003, 1259562009, 1263940008, 1263973001, 5202009, 10082001, 11426004, 14460007, 22193007, 25113000, 33332005, 33952002, 38632003, 46659004, 52201006, 63437008, 65433005, 73749009, 75572007, 82473003, 94622002, 95207005, 109918005, 188285002, 192679007, 192710009, 193116003, 202751009, 209903008, 212334009, 230260007, 230598008, 230701008, 246781001, 262748008, 276652002, 285762000, 286608004, 360353005, 363468009, 363473003, 399915007, 427086003, 441678004, 447464004, 712882000, 718362003, 723926008, 724228005, 724691003, 734021001, 763402002, 774152007, 784345005, 788864001, 838393007, 870364001, 8001000119106, 18751000119106, 291521000119105, 303481000119104, 331071000119101, 431731000124106, 690311000119104, 11018701000119109, 15637551000119104,

15699201000119104, 15733441000119104, 16058511000119105, 69463008, 418455000, 1163073003, 1228857005, 1208516002, 1208969008, 1259126003, 1260097007, 1279990000, 330231000119103, 10007009, 10970008, 36010004, 44359008, 55623006, 60258001, 68267002, 72585000, 81750007, 86365006, 89835009, 110982003, 111548007, 203928008, 213323001, 230538005, 230613008, 230762003, 232055006, 239946005, 247233000, 253131002, 389274008, 403520007, 406586005, 425082000, 698954005, 702429008, 713965007, 721506005, 722062004, 735390005, 762670009, 771074000, 783198006, 866055006, 139291000119101, 290181000119106, 290341000119103, 292151000119103, 294101000119109, 303471000119102, 342111000119101, 15739801000119100, 16000511000119103, 16024031000119100, 16845301000119108, 1162303002, 1260103000, 1259053002, 1260339004, 2198002, 6955002, 13847000, 28007006, 32358001, 75467001, 126979000, 203998000, 208023000, 212244009, 230369005, 230400008, 230412007, 230464001, 230700009, 240068006, 246767003, 254176007, 268160006, 276436007, 281411007, 298120001, 312837005, 363054006, 386537007, 389100007, 402720000, 406562008, 703313007, 715624006, 715753001, 721223002, 721489003, 721519009, 722437006, 722629008, 722653002, 773737004, 870262000, 290351000119101, 292731000119105, 330321000119107, 330451000119105, 331331000119101, 344671000119109, 347601000119108, 368601000119102, 432241000124101, 434311000124104, 1074461000119100, 16005991000119109, 16053951000119108, 16035631000119105, 1156018007, 1162336001, 1162261000, 1217676009, 1197745002, 1208973006, 1259725009, 1259501004, 1259634008, 16636321000119104, 539009, 13282008, 13980006, 15732002, 20484008, 30620003, 36905001, 42866003, 47032000, 54954004, 55004003, 57739006, 65705009, 66881004, 70476006, 76976005, 77553008, 81884004, 94471000, 95479005, 111369005, 191485005, 192981006, 204049003, 212351002, 230411000, 237724003, 238637007, 238719003, 238754005, 239089006, 247139005, 247182006, 267626000, 384993003, 396325007, 402352002, 402451008, 402453006, 403628008, 449794003, 462165005, 698641001, 698958008, 708728007, 718182008, 723450004, 724208006, 724835007, 726611001, 770628008, 788915008, 789703007, 5591000124102, 82381000119103, 84181000119109, 290631000119103, 302341000119106, 303001000119101, 348521000119104, 672521000119108, 1092351000119107, 16002391000119100, 16206661000119108, 230265002, 1237618009, 1260389007, 1263941007, 1268818001, 1279729004

Truveta:
1215926

##### 3.23 Paralysis

ICD10CM:
G81.00, G82.52, G82.54, G81.91, G81.13, G81.92, G83.14, G83.9, G81.1, G82.21, G83.11, G82.20, G83.22, G83.0, G04.1, G81.03, G83.23, G81.14, G82.50, G83.4, G83.3, G83.33, G81.94, G82, G83.13, G81.10, G83.12, G82.22, G81.11, G82.53, G81.01, G83.20, G83.31, G81.93, G82.51, G83.10, G83.32, G83.1, G81.90, G83.30, G81.12, G81.0, G83.24, G83.21, G11.4, G80.2, G81.02, G81.04, G83.2, G83.34, G80.1, G81.9, G81, G82.2, G82.5

ICD9CM:
343.3, 344.9, 342.9, 343.9, 342.00, 342.01, 344.0, 344.4, 344.40, 344.3, 342.91, 344.00, 342.80, 343.2, 334.1, 344.01, 344.02, 344.41, 343.1, 344.5, 342.92, 344.30, 344.61, 342.90, 342.02, 342.8, 342.81, 343.8, 343, 344.1, 342.1, 342.11, 344.42, 344.31, 344.2, 344.60, 343.0, 344.09, 342, 342.0, 344.03, 344.32, 344.04, 342.82, 344.6, 342.10, 342.12, 343.4

SNOMED CT:

17409003, 276221006, 441705005, 609554006, 698743007, 123535000, 1231632004, 732933009, 1156764005, 715776003, 1156858008, 702463005, 766767001, 208060001, 427065003, 723157009, 733086003, 330311000119100, 16171003, 28978003, 782945001, 128188000, 291081000119100, 1156853004, 44395000, 441980007, 11045000, 171822009, 294071000119100, 209182000, 294061000119106, 722033000, 763070001, 1052243006, 16110005, 46252003, 208078006, 860881004, 290931000119108, 340311000119104, 129614001, 209086000, 290461000119109, 345671000119105, 41455002, 29426003, 249944006, 5822000, 290921000119105, 294041000119107, 15729961000119108, 723159007, 1259038005, 9753004, 363477002, 714279000, 717266001, 290471000119103, 209197002, 732958004, 827115000, 1156856007, 314783008, 426934001, 430947007, 334711000119109, 1231212006, 29188005, 400947008, 442676003, 290411000119106, 371079004, 702315006, 1156857003, 78306007, 230263009, 723622007, 97391000119102, 446958005, 1231200007, 192966000, 724820006, 28778005, 41764006, 1208341008, 733029008, 702327009, 1208936008, 78097002, 672471000119105, 1231204003, 674131000119105, 764733009, 1187467000, 570951000124103, 111465000, 267590006, 371120001, 32162001, 232146005, 1052244000, 1259044009, 765045003, 290381000119108, 294111000119107, 15633441000119109, 1187468005, 724818008, 1231692001, 23740006, 95650006, 116288000, 230614002, 723160002, 80935004, 230778001, 240460008, 723621000, 723823004, 361000119103, 1052238004, 163602004, 49605003, 719103009, 733455003, 441688003, 723366001, 1172590009, 206226005, 702465003, 232142007, 771238004, 290321000119109, 48163001, 208067003, 230466004, 48721008, 715491000, 732948003, 86266009, 94782007, 425642008, 723819007, 12454008, 43486001, 290911000119103, 232144008, 724819000, 28534004, 698741009, 732932004, 24800001000004103, 1172631001, 1217643005, 38228000, 702321005, 1177168007, 290361000119104, 690231000119102, 15633481000119104, 446311006, 34209003, 84947004, 94242004, 732246009, 104461000119104, 15633521000119104, 41713005, 69763009, 674161000119102, 29951000119107, 278512001, 697991001, 290331000119107, 68158006, 84455002, 726606003, 15730001000119102, 1259047002, 724775006, 1197059004, 827117008, 441887006, 763403007, 787172004, 290891000119100, 40980002, 737227004, 331841000119100, 1231257002, 773425000, 22881000119100, 1259041001, 11538006, 698744001, 723824005, 103761000119107, 1231630007, 38023001, 716108004, 778030005, 782747000, 441722007, 48601000119107, 84160009, 230530003, 723162005, 782725000, 78141002, 92047003, 672451000119101, 442413005, 702320006, 725464001, 726607007, 672421000119109, 29093005, 64228003, 163606001, 427432001, 702319000, 703543005, 763374004, 246773002, 789674008, 54364001, 192971007, 249945007, 723821002, 77835008, 722209002, 278287000, 1186734006, 164110008, 246775009, 1217622009, 1260134001, 66010009, 212192004, 426167003, 163601006, 570901000124102, 690271000119104, 733330005, 763370008, 782390002, 1156842005, 48662007, 79520009, 609609004, 726609005, 1208886003, 1197587003, 75299005, 64764001, 95794005, 186479001, 733089005, 1187466009, 1208880009, 1670004, 230781006, 763068005, 764736001, 672381000119100, 764272007, 782887003, 405753002, 698755001, 718214007, 290401000119108, 723825006, 716335003, 291121000119103, 723156000, 329741000119109, 72655000, 722900008, 1187279003, 783622001, 290901000119101, 1259045005, 55637002, 372310001, 698291007, 702381007, 723820001, 1231244005, 570931000124105, 722901007, 785305006, 140281000119108, 1231631006, 19373007, 111633007, 431061000124100, 77015008, 764688002, 79591004, 337461000119108, 1156841003, 56409008, 1156855006, 1259046006, 126963001, 209102002, 278285008, 785304005, 1187506008, 1231256006, 75019001, 672441000119103, 1845001, 37991008, 58193001, 702464004, 426536006, 723158004, 192958009, 724349009, 8663007, 312792007, 290491000119102, 1156790008, 15633401000119107, 1593000, 298282001, 400946004, 1156763004, 307756005, 330521000119103, 1187470001, 6481005, 60389000, 733650000, 763367009, 1231636001, 186476008, 441892008, 442668000, 1231628005, 441894009, 39912006, 763069002, 763369007, 764734003, 782670003, 1156761002, 783764008, 690241000119106, 783094006, 291091000119102, 278286009, 442024001, 609557004, 291111000119105, 1534008, 723826007, 783697000, 230773005, 782726004, 690261000119105, 1173998003, 68107009, 230261006, 763743003, 294051000119109,

12367511000119101, 698292000, 427943001, 442097001, 672371000119103, 723822009, 344261000119101, 91327001, 313434001, 723161003, 778011005, 49823009, 79633009, 89437009, 86022000, 726622002, 106241000119108, 208071000, 609553000, 783005002, 294121000119100, 690281000119101, 442181008, 192965001, 230777006, 232113004, 420675003, 702314005, 672461000119104, 192949002, 782723007, 371129000, 771304005, 782727008, 1187191003, 23728006, 343251000119100, 129583005, 702316007, 763377006, 59103002, 192964002, 442155009, 430959006, 1259043003, 95651005, 763376002, 83886009, 290371000119105, 232143002, 442733008, 726608002, 1231197006, 80420005, 770724001, 101461000119102, 778048001, 782388003, 1052237009, 81774005, 192976002, 186480003, 212193009, 163605002, 447262002, 1187278006, 441991000, 717827000, 50582007, 54099005, 698754002, 770720005, 690251000119108, 20022000, 441951003, 230780007, 442077006, 763375003, 1231683004, 763373005, 97381000119100, 1217379007, 698742002, 782746009, 290481000119100, 278284007, 702317003, 16846261000119106, 1187465008, 275468009, 447378002, 75111000, 441794001, 726610000, 18761000119108, 102831000119104, 15730041000119100, 570911000124104, 163604003, 186478009, 725139005, 764686003, 783698005, 123534001, 1231203009, 192970008, 446957000, 1208876004, 21524000, 290311000119102, 294131000119102, 93743001, 404689008, 732949006, 291101000119107, 192967009, 4275001, 1231677002, 232115006, 702318008, 1259560001, 442020005, 425882004, 789005009, 441717007, 765753004, 230260007, 763402002, 18751000119106, 783198006, 290341000119103, 294101000119109, 281411007, 290351000119101, 330321000119107, 344671000119109, 726611001

#### 3.24 Peptic Ulcer Disease

ICD10CM:
K26.9, K27.9, K27, K27.5, K27.7, K28.9, K26.7, K27.6, K27.4, K25.7, K27.1, K27.2, K27.0, K25.9, K27.3, K28.7

ICD9CM:
534.90, 532.9, 534.7, 531.91, 532.90, 531.71, 532.70, 533.9, 534.91, 534.9, 531.9, 533.71, 533.90, 533.91, 532.7, 533.7, 531.90, 531.7, 531.70, 532.91, 532.71, 534.70, 533.70, 534.71

SNOMED CT:
72408002, 717892001, 31452001, 56461008, 307260009, 196775009, 109814008, 55483002, 1197691004, 23649000, 737190002, 23812009, 62465000, 307510004, 415623008, 16516008, 235650007, 235702004, 59913009, 722200003, 724523000, 16121001, 37442009, 77987006, 111350000, 724528009, 22511002, 276525003, 57940000, 196694009, 235688009, 60400003, 61300005, 235612001, 397825006, 717849006, 313425006, 708645007, 689991000119100, 12625009, 34580000, 2066005, 59356009, 35560008, 307190008, 5492000, 81142005, 762274007, 86983005, 47152002, 3023008, 439955006, 12274003, 723102007, 59515005, 12847006, 87756006, 53877005, 196652006, 62366003, 76078009, 196666001, 723105009, 250856961000119105, 66673003, 196719008, 724532003, 32490005, 444926003, 40214005, 24060004, 76181002, 64595005, 301007008, 717863000, 56776001, 360374006, 60531007, 24001002, 442805003, 88968005, 218541000119109, 36975000, 46523000, 89469000, 63137003, 6761005, 13200003, 15902003, 21759003, 173889005, 235652004, 43406003, 1567007, 17067009, 51847008, 221899751000119107, 68834009, 79118000, 62477005, 38365000, 60551006, 88169003, 308882008, 1197692006, 26221006, 62936002, 86895006, 57871005, 34602004, 54157007, 724520002, 724533008, 10389003, 10897002, 1144981002, 53337006, 196632005, 196671008, 738791006, 70418001, 81518000, 30183003, 56579005, 49232000, 51868009, 235609004, 4269005, 2783007, 1197714001, 245754007, 72219001, 1197690003, 39204006, 235608007, 724519008, 1197685001, 27281001, 72395008, 22620000, 723099007, 84124004,

85859006, 10699001, 81225008, 717848003, 95530000, 129141000119104, 196707000, 235610009, 235614000, 18169007, 78054007, 87796008, 55617001, 85787009, 89748001, 683509791000119109, 196658005, 12355008, 76338009, 76796008, 64121000, 8322005, 103691000119106, 196687006, 62341002, 75342000, 307512007, 723101000, 235689001, 41986000, 77661009, 865912001, 1086791000119100, 30514008, 81387001, 738792004, 18665000, 35681000, 840592004, 956321981000119108, 16694003, 36246001, 74341002, 80953005, 3483000, 34021006, 40719004, 48658001, 724529001, 34921009, 58711008, 111353003, 717879004, 500790491000119109, 96720031000119104, 31301004, 4911000119101, 235232008, 1197688004, 11818002, 723100004, 29755007, 77410006, 196712004, 782935003, 1197724009, 2807004, 724530006, 840591006, 89662003, 90489006, 698496004, 64094003, 733147004, 1197771003, 708644006, 196607008, 15115006, 66767006, 18367003, 46708007, 57748001, 307233002, 39344002, 173842008, 30811009, 42698006, 55746001, 717891008, 50663005, 86258000, 196682000, 235613006, 109813002, 67964002, 162033007, 235690005, 307259004, 360372005, 723103002, 79806007, 128286008, 724534002, 235651006, 54798007, 66636001, 713638002, 43694004, 781203005, 724531005, 314632001, 90257004, 196609006, 72486001, 1177073003, 23693000, 724521003, 128288009, 724522005, 41626001, 45485004, 89600751000119100, 28945005, 95529005, 287811008, 45640006, 1148555003, 12384004, 73481001, 128287004, 9829001, 196639001, 48974009, 235611008, 1163589006, 762276009, 53132006, 235648004, 17593008, 235692002, 58085004, 737189006, 90628007, 359574001, 57246001, 48336009, 415624002, 54053008, 84038009, 360377004, 62838000, 22157005, 723104008, 19850005, 49916007, 88700009, 61347001, 39755000, 43035002, 63954007, 64398008, 723884008, 724516001, 786068005, 35517004, 1197708008, 47064007, 307511000, 28082003, 35892001, 717878007

#### 3.25 Peripheral Vascular Disorders

ICD10CM:
I70.343, I70.431, I70.541, Z95.81, I70.491, I70.591, I70.645, I71.51, I71.52, I70.545, I70.61, I70.72, I70.319, I70.418, I70.321, I70.648, I70.749, I70.298, I70.562, I70.531, I70.713, I70.361, I70.413, I70.508, K55.032, I73.9, I70.398, I70.798, I70.344, I70.603, K55.8, I70.438, I70.691, I71.5, I70.338, I70.693, I71.12, I70.49, I70.444, I71.4, I70.24, I70.619, I70.40, I70.461, I70.56, I70.70, I67.1, I70.669, I70.449, I70.52, I70.641, I70.745, I70.202, I70.392, I70.644, I70.498, Z95.9, I71.41, I70.92, K55.039, I70.521, I70.429, I70.74, I70.341, I70.218, I70.243, I70.768, K55.012, I71.21, I70.248, I70.618, K55.0, K55.031, I71.011, I70.23, I70.548, I70.631, I70.53, I71.62, I70.342, I70.399, I70.568, I70.22, I70.90, I70.642, I70.535, I70.7, I73.8, I70.45, I70.698, I73.89, I70.633, I70.39, K55.31, K55, I70.26, K55.04, I70.33, I70.518, I70.60, I70.744, Z95.812, I71.60, I70.3, I70.513, I70.643, I70, I72.2, I71.10, I70.729, I71.30, I70.55, I70.362, I70.739, I70.509, K55.051, I72.1, K55.06, I70.213, I70.712, I70.332, I70.538, K55.03, K55.061, I70.512, I70.522, I77.1, I71.61, I70.519, I71.00, K55.029, I70.268, Z95.820, I70.212, I70.232, I70.799, I70.549, I70.734, I70.203, I70.761, Z95.818, I70.51, I70.539, I70.708, I71.02, I71.32, I70.201, I70.433, I70.735, I70.35, I70.462, I70.561, I70.221, I70.542, I70.42, I70.523, I70.533, I70.403, I70.529, I70.303, I70.445, I71.2, K55.02, I71.012, I70.331, I70.469, I71.01, I70.0, I70.263, I70.43, K55.1, K55.2, Z95.811, I70.262, I71.03, I71.43, I70.41, I70.503, I70.731, I70.228, I71.0, I72.0, I72.4, I70.493, I70.5, I70.621, I70.709, I70.769, K55.059, I70.668, K55.041, I70.209, I70.602, K55.019, I71.42, I70.223, I70.62, I71.23, I70.241, I70.322, I70.328, I70.434, I70.448, I70.421, I72.5, I70.663, I70.723, I70.234, I70.611, I70.391, I70.692, I70.702, I70.29, I70.335, I70.742, I71, I71.31, I70.231, I70.629, I71.9, K55.049, K55.05, I70.408, I70.442, I70.511, I70.638, I70.318, I70.44, I70.599, I70.468, I70.612, I70.20, I70.21, I70.293, I70.31, I70.36, I70.4, I70.66, I70.703, I72, I70.368, I71.11, I70.34, I70.443, I70.211, I70.793, I71.3, K55.33, I70.291, I71.8, I70.432, I70.2, I70.441, I70.649, I70.219, I70.245, I73.1, I70.632, K55.022, K55.062, I70.628, K55.30, I71.50, I70.25, I70.412, I70.721, I70.363,

I70.748, I70.222, I70.242, I70.439, I70.233, I70.64, I70.311, I70.528, I70.791, I70.261, I70.499, I70.722, I70.309, I70.593, I71.6, K55.20, I70.422, I70.532, I70.69, I70.763, I70.8, K55.011, K55.021, I70.329, I70.349, I70.543, I70.312, I70.393, I71.010, I70.502, I70.79, I72.6, I70.50, I70.269, I70.313, I70.54, I70.402, I70.534, I70.613, I70.699, I70.733, I70.738, I70.76, K55.01, I70.302, I70.46, I70.601, Z95.82, K55.069, I70.308, I72.8, I71.1, I70.238, I70.411, I70.711, I70.6, I70.30, I70.75, I70.323, I70.732, I71.019, I70.608, I70.91, I71.33, I71.13, I70.741, I70.229, I70.369, I70.661, K55.3, I70.592, I70.634, I70.719, I70.463, I70.622, I70.635, I70.299, K55.21, I70.249, Z95.8, I70.32, I70.762, K55.052, I70.1, I70.63, K55.042, I70.409, I79.0, I71.20, I70.301, I70.428, I70.563, I70.59, I70.718, I71.22, I70.345, I72.9, Z95.810, I70.609, I70.701, I70.244, I70.728, I70.743, K55.32, I70.208, I70.235, I70.339, I70.73, I71.40, I70.501, I70.292, I70.423, I70.623, I70.334, I70.348, I70.662, I70.71, K55.9, I70.333, I70.492, I70.792, I70.435, I70.544, I70.239, I70.419, I72.3, I70.598, I73.81, Z95.828, I70.401, I70.9, I70.569, I70.639, I70.65

ICD9CM:
440.21, 440.2, 443.2, 441.01, 440.20, 442.84, 442.8, 443.89, 440.0, 443.24, 443.23, 442.82, 443.29, 443.82, 442.81, 442.9, 441.03, 441.9, 441, 441.5, 442.0, 447.1, 440.23, 440.29, 442.83, 441.3, 442.1, 442.89, 441.1, 440.24, 440.3, 557.9, 443.21, 440.8, 440.30, 441.0, 443.81, 441.4, 441.6, V43.4, 557.1, 441.2, 441.7, 440.9, 440.22, 440.32, 443.1, 442.3, 440.31, 440.1, 440, 443.22, 441.00, 443.9, 440.4, 442.2, 441.02, 442, 443.8

SNOMED CT:
26214006, 29507009, 29899005, 72408002, 83351003, 93067003, 194843003, 204490005, 210789004, 233944003, 233968006, 253318000, 253527000, 297222002, 301899003, 373092008, 397187003, 400159008, 450306008, 459182005, 706913006, 713412006, 733225008, 432481000124106, 12245491000119103, 16006351000119106, 16065251000119109, 39291006, 45894003, 69357003, 88174006, 93329009, 213310008, 233977004, 234018007, 234060006, 234081006, 239939001, 253319008, 425448004, 425936006, 459160003, 699314009, 717043006, 735897000, 12236911000119107, 15648401000119104, 21290001000004104, 15966221000119102, 15970301000119109, 1144912000, 1209062008, 1254666000, 56267009, 61612001, 234129008, 236491005, 237286004, 253656007, 297135003, 427134009, 427776007, 443770005, 447690005, 448627003, 461376004, 699705001, 737331008, 143551000119106, 285271000119105, 285531000119105, 330791000119108, 16288421000119101, 1519002, 21542005, 329541000119100, 16007831000119105, 23637001, 28967006, 88922007, 234199000, 276697006, 721513005, 723869004, 5601000124105, 132221000119109, 286361000119101, 367431000119107, 433941000124104, 16837461000119106, 1204198001, 1204148002, 10645431000119107, 1279832006, 76598006, 128058007, 191306005, 210900007, 268187005, 308135003, 312593004, 402746001, 402860008, 403958000, 442298000, 444869007, 703331008, 713034001, 284811000119102, 285201000119100, 677781000119104, 16001671000119100, 254774003, 1145199001, 1187628000, 1197661009, 1179366006, 1208761000, 1209205004, 19474002, 27550009, 32450007, 38742007, 51070004, 61091005, 70936005, 79439001, 95459002, 200227005, 233957006, 238761009, 253674003, 403765001, 442421004, 448637008, 449513006, 702428000, 703244000, 707372009, 722929005, 870564000, 134971000119105, 15712001000119106, 16014391000119106, 897005004, 1144809002, 1197697000, 1231183003, 1208856005, 1255265004, 54538006, 93424000, 198663000, 200251000, 234058009, 234069007, 238788004, 247479008, 280973003, 282036005, 447929001, 697909006, 703369003, 721385005, 735831001, 871589004, 434891000124102, 15712281000119109, 1148916009, 11842891000119102, 1179298002, 1204187002, 1251566005, 1269053004, 145851000119109, 64009001, 95450003, 233947005, 233983001, 233996000, 238816001, 262716009, 277330009, 447365002, 447827003, 698352000, 711545001, 719678003, 735896009, 789682008, 11816541000119102, 15712441000119103, 16009351000119100, 54160000, 1204123000, 1208865003, 17024001, 53741008,

56097005, 94191001, 234033004, 274119009, 308187004, 312588002, 442693003, 447773008, 449904002, 461112001, 698997000, 699814001, 17441000119102, 455041000124100, 11799281000119102, 16006151000119108, 16899261000119106, 16012191000119104, 621041000124100, 6378004, 31295000, 36664002, 38877003, 42970005, 49465005, 91399006, 210908000, 230467008, 231847000, 234055007, 239200000, 277324009, 402855009, 405577005, 410022000, 418918009, 438646004, 448471006, 709587006, 724874000, 737158007, 782699008, 840580004, 15710201000119100, 16012951000119106, 16025871000119109, 1144435009, 1144807000, 1237564009, 23341007, 38385001, 39823006, 44101005, 55520003, 59204004, 62469006, 64638002, 80426004, 85081000, 86487001, 210845007, 234124003, 253321003, 253634009, 307223005, 312825008, 423674003, 429397000, 447772003, 448154004, 448326002, 448804008, 460954009, 460958007, 461387007, 472748007, 724857000, 651000119108, 679491000119103, 11759891000119108, 1187305006, 1196843005, 8214000, 17810004, 92977007, 200207009, 210835005, 230222003, 238765000, 253371000, 297713002, 312935003, 403650009, 403964007, 441574008, 448087002, 448872008, 473449006, 724002003, 291481000119105, 15960781000119108, 15967341000119100, 16009391000119105, 15966101000119102, 1144950006, 11824521000119108, 1254658009, 1254664002, 40283005, 65838008, 123642008, 234121006, 239203003, 253622001, 276794009, 405332009, 461408000, 697898008, 786054003, 871588007, 124171000119105, 434821000124104, 12237311000119105, 15663561000119101, 15972661000119104, 15988351000119101, 1148894007, 1197698005, 1234908005, 2213002, 4262001, 6956001, 8166000, 15531007, 34781003, 82275008, 91997004, 111298007, 128054009, 195396009, 234028003, 240469009, 277495000, 307398006, 307420006, 373135009, 447348006, 447664007, 448809003, 473005004, 697914005, 153821000119103, 285591000119109, 1085121000119107, 15710881000119109, 14493003, 16891191000119108, 1197742004, 1231614004, 1237130004, 1237611003, 2261009, 2929001, 10392004, 39785005, 52931009, 57534004, 65312002, 66620003, 66923004, 85890008, 95441000, 111294009, 233781007, 234057004, 238774003, 254784002, 266267005, 277198009, 303070000, 408546009, 447665008, 461435009, 472749004, 718096004, 720519003, 735576008, 435271000124103, 461441000124103, 12237351000119106, 18000001000004101, 29495008, 55589000, 61267008, 63040001, 75825001, 80383008, 93057008, 212365000, 239937004, 253342001, 297196004, 311798009, 313001006, 402856005, 405565000, 425007008, 439007008, 449626004, 461414007, 702792005, 737244000, 32201000119107, 142121000119101, 15973221000119103, 16014311000119102, 53485006, 1184601001, 30400005, 40250003, 50323003, 83799000, 195200006, 210871006, 247121001, 253310007, 307396005, 370480003, 405756005, 441322009, 449188001, 449897004, 472102006, 726490008, 737160009, 737196008, 762629007, 766959002, 781149006, 871603009, 7991000119102, 429231000124101, 11759931000119100, 697985003, 1156383000, 24272004, 74908007, 93384001, 195405004, 198711003, 234006003, 234017002, 234152007, 253650001, 312607004, 405554005, 443916000, 447571007, 448645003, 460880006, 697921005, 714180007, 721464009, 732230001, 767030009, 789544005, 291371000119100, 15649861000119105, 15711361000119101, 139971000119106, 59117006, 82986004, 95657009, 195234007, 204379001, 210867008, 234032009, 269438006, 276622005, 306854007, 308137006, 439731006, 447927004, 448630005, 461090007, 770582001, 840419005, 124001000119104, 15972701000119106, 16012631000119105, 1010730002, 11816501000119104, 1255258007, 3704008, 75971007, 77788005, 85448008, 195353004, 232025004, 247126006, 253197001, 276394007, 278533005, 303086007, 371071001, 427199002, 429434005, 448280008, 717765001, 722191003, 766951004, 783090002, 822975003, 823016000, 124081000119107, 11781601000119105, 15711201000119109, 15978471000119109, 697899000, 1010652002, 15952061000119106, 1260327007, 28608002, 50817002, 126739000, 195182007, 210865000, 210872004, 233960004, 235878005, 253309002, 271870002, 309735004, 410469004, 459064009, 460593008, 609465003, 609480009, 703207000, 41371000119100, 330111000119103, 446491000124106, 15960021000119107, 16009431000119100, 880000009, 11255009, 21848000, 37104009, 40276003, 127031002, 199900008, 232044006, 234000009, 234205007, 235844004, 253643000, 255022003, 276706004, 285738005, 306849003, 373094009, 431667009, 460913001, 461331004, 471288001, 720809000, 721979005, 735832008, 868230003, 25971000119100, 124071000119109,

285321000119103, 430781000124102, 450251000124102, 15960061000119102, 1197700001, 1204120002, 11831321000119100, 1231788002, 1264001009, 2066005, 31938009, 59356009, 77978002, 83940008, 234013003, 253644006, 253652009, 254883003, 297136002, 306853001, 447274003, 448097006, 457602002, 697904001, 703265006, 703356002, 717868009, 762630002, 870420005, 16012071000119101, 1010627004, 338061000119101, 15970341000119106, 1163244009, 1264235001, 15711321000119106, 33806008, 45549002, 87730004, 90507008, 234144009, 276500007, 370552002, 402661009, 403965008, 417544004, 461597002, 697906004, 703294004, 723857007, 737891000, 767127000, 781065009, 871590008, 871667007, 338891000119100, 350331000119106, 677651000119102, 15708041000119108, 15710921000119102, 1187126002, 1231779006, 1237607009, 52593001, 52711003, 67600007, 134335004, 239924002, 253620009, 280968009, 373138006, 402662002, 444569004, 447902006, 461424004, 609486003, 697907008, 697928004, 717824007, 766756002, 766945009, 5491000124101, 15997951000119102, 16009271000119101, 1204172007, 14336007, 71719003, 79553004, 192765008, 234030001, 313000007, 609449004, 722985001, 724446001, 724600007, 734326000, 766976003, 767309006, 16012791000119103, 15969021000119101, 47780009, 67656006, 195273007, 199901007, 200304004, 233981004, 234198008, 253316001, 253647004, 253675002, 307314009, 373093003, 402859003, 427919004, 446432002, 448078006, 449435001, 460965004, 461440001, 703243006, 703277001, 717867004, 783423004, 788893001, 860679004, 285261000119104, 285551000119104, 12236991000119103, 16012751000119108, 1003844005, 1156066000, 18322005, 46253008, 65340007, 73557005, 111323005, 204457005, 213049001, 232071006, 233935004, 234026004, 234206008, 266272001, 402659000, 439969006, 445543002, 447928009, 448595006, 460307002, 721459005, 766957000, 780842009, 860687003, 860820000, 5531000124101, 292621000119100, 436791000124101, 15711561000119107, 15712681000119106, 16026431000119101, 717267005, 1156019004, 1196905008, 1204377006, 10643431000119108, 1256043002, 1258896001, 27015006, 34704009, 63168004, 92266009, 186567003, 192764007, 197000003, 234066000, 234153002, 266254007, 271990007, 371029002, 371804009, 416792008, 439162001, 448577008, 450301003, 460957002, 461389005, 697915006, 788885001, 126011000119107, 132531000119105, 143371000119108, 285301000119107, 431391000124106, 1204196002, 1204127004, 1285065002, 10349009, 15386007, 19363005, 47468000, 58872001, 82453008, 93468003, 233941006, 234133001, 247099009, 254206003, 262945004, 271573009, 277315000, 285710002, 403966009, 405580006, 425453009, 448081001, 461104009, 699056001, 714818009, 724361001, 724427002, 724842007, 455941000124104, 15699361000119107, 698191003, 284861000119104, 1010680000, 13026000, 31110000, 195427008, 213308006, 371803003, 414297000, 424802006, 447813008, 448086006, 448726004, 461091006, 707420003, 786041005, 285521000119107, 12237791000119102, 15708481000119108, 15735201000119101, 15988391000119106, 1144919009, 1145181009, 1204176005, 7346000, 31570000, 59515005, 92997002, 235853006, 253364005, 253663007, 253717008, 307319004, 309750008, 314189001, 404669007, 404814007, 461382001, 473109007, 703183007, 717865007, 840438001, 128171000119104, 285641000119100, 434951000124104, 16066331000119101, 1087081000119102, 16697181000119105, 1144766004, 2367005, 56975005, 76019006, 195437003, 197812000, 204431007, 234044007, 235900003, 261482004, 277299009, 307362003, 433874008, 448329009, 458088003, 460969005, 461614001, 472100003, 723991007, 783418004, 871662001, 135531000119100, 293481000119109, 15649181000119109, 11844791000119100, 11798911000119109, 1268294009, 15023006, 17228008, 38230003, 40272001, 53666001, 77535002, 105373008, 128558004, 232890002, 233985008, 239945009, 251038002, 253670007, 253724009, 402808004, 447817009, 460510005, 460524009, 460606004, 473444001, 697901009, 703268008, 721461001, 722287002, 733124000, 789689004, 99451000119105, 285401000119104, 15966981000119108, 16697101000119102, 57484009, 109558001, 15708241000119104, 1201858000, 6098001, 18887007, 35095003, 77181006, 81817003, 94980009, 195199008, 254207007, 449486006, 461103003, 461433002, 718634003, 734963004, 735583001, 767311002, 778061001, 783629005, 868222004, 132611000119104, 153911000119104, 285451000119100, 1156032009, 20116006, 29950007, 111696001, 210837002, 230723007, 234008002, 234027008, 312997008, 413552002, 429098002, 430305000, 441024003,

448000003, 461557000, 472756005, 697912009, 721514004, 9901000119100, 101711000119105, 132161000119105,
679541000119107, 12237551000119104, 15711521000119102, 16007911000119106, 16014071000119107,
74561007, 16012231000119108, 10643631000119106, 1255624007, 63043004, 95691008, 195154000,
210860005, 210879008, 233994002, 239947001, 253719006, 302233006, 403511003, 445027003, 449014008,
473394001, 703158007, 703161008, 788894007, 116291000119103, 132551000119104, 15712081000119103,
16064811000119108, 1268295005, 1279844009, 15705007, 28840001, 59877000, 126729006, 195265003,
195410000, 210855006, 210901006, 232967006, 233997009, 234004000, 234140000, 253320002, 253711009,
254780006, 271432005, 311366001, 384677005, 461419002, 471276003, 710167004, 737184001, 762302008,
33041000119105, 429181000124108, 16019351000119102, 1142107003, 1208816008, 5753006, 20258000,
20696009, 45492009, 49584005, 53877005, 61401005, 64253000, 93305002, 128067007, 195327007,
195450002, 233844002, 238771006, 239941000, 239943002, 253673009, 266262004, 400047006, 402664001,
449493005, 472790001, 473445000, 713030005, 717880001, 783707003, 16011391000119108, 16018831000119106,
429241000124106, 344015141000119100, 1204202004, 1263550001, 44918005, 68237008, 81363003,
89330007, 93347003, 210829005, 232788006, 232962000, 400130008, 403517004, 447812003, 449085001,
461094003, 461095002, 471274000, 697910001, 703308008, 707371002, 712722007, 840420004, 142111000119108,
286411000119108, 433821000124102, 15648691000119108, 15711961000119102, 16011831000119101,
778060000, 1010645004, 1208842008, 45874007, 62366003, 76078009, 85564003, 128555001, 232045007,
234034005, 234083009, 275519006, 373095005, 445305001, 460312001, 734879002, 789666007, 840307003,
429811000124106, 16018911000119107, 897555003, 1204145004, 2571000112102, 1231671001, 1258933008,
16023671000119106, 11614003, 28366008, 56276002, 65084004, 95440004, 200204002, 210847004,
233966005, 235851008, 253651002, 276792008, 361209006, 417426005, 422001004, 445928005, 448104005,
449444000, 703205008, 704140009, 724783000, 767029004, 770687001, 783420001, 5521000124104,
132591000119109, 431601000124105, 15966861000119106, 16697021000119105, 28810001000004102,
1204153007, 16023751000119106, 5176003, 18296004, 26850007, 42861008, 43663004, 66673003,
75398000, 80820004, 83938003, 93353003, 95455008, 238813009, 245544005, 262937009, 447111007,
703226008, 707477002, 871536000, 15699401000119103, 16014551000119108, 1003874002, 1222665003,
10644431000119106, 23999003, 39402007, 54265003, 67362008, 77628002, 163989008, 195155004,
234190001, 234243005, 239927009, 253333008, 282348002, 448887003, 461093009, 697916007, 697919000,
697927009, 868215007, 64146000, 1155919006, 1204214007, 15663521000119106, 1231188007, 36025004,
67189007, 195235008, 233969003, 236777003, 277319006, 277328007, 314188009, 399980009, 402433007,
703198009, 721381001, 723858002, 724486009, 735595001, 737161008, 783630000, 11799811000119107,
15711481000119102, 16014231000119100, 343831000119107, 1153634000, 1208943002, 1208940004,
1237135009, 1264081007, 9058002, 34037000, 37796009, 59062007, 60190000, 88784003, 213385006,
232048009, 234201003, 253657003, 445349004, 703184001, 703197004, 721380000, 769036006, 124391000119108,
285511000119100, 11770251000119105, 15649941000119102, 15964661000119102, 15966541000119105,
1204213001, 11433004, 51536009, 56462001, 75138007, 76181002, 204300001, 241154007, 262947007,
302300004, 307323007, 428543009, 449527001, 460380002, 703357006, 707478007, 132111000119107,
436781000124104, 677791000119101, 679511000119108, 11760091000119104, 15930061000119106,
16662571000119103, 1148691001, 1155698001, 11831281000119105, 23854007, 31529002, 34736002,
55734000, 75473000, 111697005, 123661007, 230462002, 232968001, 253723003, 254787009, 307316006,
359938001, 371038000, 384680006, 399046008, 447825006, 721714001, 735853004, 770600002, 677601000119101,
15648241000119103, 15929861000119106, 16026111000119100, 1204121003, 968361541000119107,
31481000, 36422005, 43293004, 46286007, 49605003, 63934006, 82582006, 84272007, 92962004,
95445009, 195236009, 209083008, 210801007, 210878000, 230465000, 234009005, 234077006, 239931003,
253721001, 276620002, 414086009, 427184002, 447846002, 449337005, 449906000, 713081000, 716745004,
870336006, 7931000119101, 122141000119109, 11800451000119102, 79325000, 90383006, 191329002,

213266005, 232042005, 234195006, 238750001, 253627007, 300921000, 402833001, 442249009, 447275002, 447938004, 697923008, 734382000, 871901008, 285411000119101, 450641000124101, 11845231000119106, 15710441000119102, 1157162007, 11826981000119108, 1231255005, 1269238002, 44801000087106, 24001002, 38214006, 58358002, 128063006, 195412008, 200259003, 253297008, 253306009, 253637002, 277320000, 307407008, 425771001, 448305002, 448644004, 460609006, 461428001, 703258003, 763316006, 767169001, 783413008, 789036001, 871653003, 285491000119105, 437931000124100, 16014871000119101, 15710641000119100, 1148584007, 12065931000119105, 29149100011910 8, 10930001, 40627003, 65366005, 69352009, 93059006, 93940006, 195447000, 210864001, 233979001, 234036007, 234118009, 238806009, 253693001, 277301002, 361132001, 448099009, 459163001, 703206009, 714817004, 719475006, 722645005, 771476007, 218541000119109, 434831000124101, 679501000119105, 15649421000119102, 15711841000119105, 16002111000119106, 16006031000119103, 16711591000119103, 1204179003, 1204164005, 16606761000119102, 46523000, 88837005, 93469006, 210907005, 233971003, 251036003, 253193002, 253304007, 253544001, 254776001, 262941008, 275517008, 281717005, 402660005, 461101001, 698187008, 713078005, 5001000124101, 449821000124101, 15966941000119103, 16005391000119108, 16014031000119109, 1155689004, 17406005, 88898002, 186668002, 204378009, 210888004, 233209007, 233939005, 233995001, 234007007, 238781005, 238783008, 253623006, 403668007, 428031002, 448084009, 448105006, 448487009, 448500006, 460962001, 461386003, 461609000, 470760002, 703180005, 713542007, 766751007, 122021000119104, 292631000119102, 329641000119104, 11816581000119107, 1156983007, 11826941000119103, 11263005, 24805007, 33591000, 68109007, 80758005, 230740003, 233942004, 233958001, 233959009, 234059001, 302728008, 402858006, 428089008, 471292008, 471293003, 735572005, 783417009, 5581000124100, 27820001000004104, 234160008, 429821000124103, 876858006, 12253004, 15902003, 16061002, 60706008, 67486009, 81103005, 93062009, 93690007, 195414009, 206202008, 236120009, 253512000, 253577002, 253681005, 253730009, 297149007, 302880005, 416881003, 427213005, 448744007, 461629004, 734959006, 735911004, 762433009, 286081000119107, 332431000119103, 12237391000119101, 1197694007, 1197722008, 3090000, 42878004, 46516001, 66403007, 69390003, 206403000, 210836006, 232356000, 239944008, 307394008, 308692003, 764965000, 792908009, 860680001, 141631000119100, 15710961000119107, 16012471000119105, 332451000119109, 1155687002, 710006, 17067009, 22949006, 38996000, 126037009, 192770001, 213046008, 253308005, 425932008, 860686007, 871580000, 329651000119102, 1003841002, 16024311000119107, 1209172009, 12060551000119102, 14363008, 27986000, 50128001, 61959006, 69798007, 77930002, 79267007, 125963005, 195425000, 234039000, 234134007, 253655006, 402676001, 402851000, 446465005, 448303009, 718216009, 721664001, 783416000, 29721000119101, 11799731000119109, 15710281000119108, 15966381000119107, 1208430005, 64775002, 68926002, 70629007, 89135007, 111296006, 195276004, 210889007, 230732009, 234191002, 240485004, 253680006, 289923007, 312378004, 398274000, 439470001, 448113007, 448528000, 460581004, 700063005, 713029000, 762256003, 762286005, 762652006, 871637001, 15631651000119107, 402416000, 1010646003, 890220003, 16019391000119107, 1209203006, 331021000119102, 44781000087105, 28067008, 46085004, 58123006, 93396008, 95458005, 192755002, 195160000, 210788007, 210808001, 233949008, 234075003, 237343001, 277494001, 285749004, 306855008, 312585004, 373420004, 425600006, 442735001, 449451009, 460596000, 734396006, 762250009, 840439009, 1090361000119108, 890114008, 331041000119108, 1254662003, 8370004, 17709002, 192759008, 200249004, 239930002, 253633003, 314186008, 403544009, 447914003, 448180003, 448632002, 461412006, 721456003, 735899002, 34881000119105, 285281000119108, 350291000119100, 11770291000119100, 16004991000119105, 1163536005, 1285020006, 13290008, 33456005, 36477001, 52403007, 65198009, 73390009, 74315008, 95446005, 128066003, 175340007, 204448004, 209179005, 210852009, 233938002, 233946001, 253322005, 308882008, 403189002, 444819004, 448157006, 449427004, 703336003, 724063005, 132281000119108, 1149059007, 15663481000119106, 5230009, 10878002, 13943000, 26201005, 195363007, 195421009, 209163009, 253710005, 276793003, 312373008, 409641002, 429421009, 449426008, 449532000, 472758006, 707437005, 762537007, 762633000,

122051000119107, 450801000124102, 11816061000119107, 15972461000119101, 724437007, 15710321000119103, 11858661000119108, 10596002, 94196006, 195299000, 200254008, 200296007, 210824000, 232040002, 233991005, 234241007, 253329001, 363340006, 407529009, 441557008, 703221003, 703285005, 737333006, 5431000124100, 133411000119108, 434881000124100, 448661000124102, 1086951000119108, 16730001000004104, 19886006, 897593008, 1142085005, 1153612002, 1156017002, 1235006003, 1258882007, 11791001, 56253000, 57614009, 62207008, 64156001, 67992007, 85503007, 89400003, 111293003, 200226001, 204482005, 234130003, 253312004, 266520007, 300919005, 445874008, 450311005, 460619000, 460956006, 707436001, 712866001, 721206006, 341801000119101, 12237271000119105, 15649241000119107, 15978431000119106, 16025751000119105, 16780001000004106, 15966261000119107, 890376009, 15968861000119105, 1173099009, 14180000, 25114006, 37553001, 197244008, 210779003, 230223008, 237345008, 274101000, 285740000, 307406004, 310625000, 405545007, 448117008, 460952008, 840679006, 437911000124106, 11816141000119107, 15978071000119100, 16014671000119100, 1204184009, 302661000119103, 1209180002, 759556531000119104, 711161000124103, 7216000, 20981007, 71226004, 87438005, 236493008, 251042004, 280143008, 280972008, 280979004, 314268004, 415810004, 471277007, 724442004, 766950003, 783422009, 121881000119102, 284821000119109, 11760051000119109, 11799361000119102, 15707841000119101, 16662371000119109, 15970261000119101, 16662611000119107, 961792901000119102, 53337006, 55230008, 62240004, 79955004, 111873003, 195209007, 195394007, 238812004, 307400005, 314269007, 403390002, 405810005, 448782004, 449495003, 449533005, 460598004, 473392002, 703279003, 789667003, 285291000119106, 291411000119104, 680941000119100, 12237231000119107, 16025711000119109, 11798871000119106, 1268521006, 35239001, 37151006, 70418001, 73192008, 210890003, 232036006, 232038007, 232460001, 253303001, 279990005, 306852006, 312586003, 445208002, 448100001, 473443007, 709585003, 721383003, 733224007, 735115007, 735568006, 771515001, 871599009, 132151000119108, 285441000119102, 291401000119102, 329531000119109, 429851000124107, 16012871000119107, 20343006, 1204174008, 10646231000119103, 57662003, 57834008, 90539001, 93415009, 210887009, 210893001, 253196005, 253690003, 311808009, 312374002, 443904006, 448611005, 449896008, 461097005, 461384000, 722922001, 771236000, 783415001, 792844003, 11844951000119108, 724598001, 11815981000119107, 15968821000119100, 1269244003, 18932005, 28248000, 70668000, 90099008, 93050005, 210790008, 232354002, 234072000, 247475002, 253698005, 402837000, 425420004, 425527003, 441545004, 448092000, 459181003, 707413005, 724431008, 124091000119105, 132601000119102, 293491000119107, 86299006, 1010639007, 1197658008, 331031000119104, 776009, 1351002, 14482000, 24807004, 33490001, 51274000, 58780002, 58863009, 72565004, 95838002, 210883008, 230720005, 234132006, 238786000, 240567009, 306848006, 307402002, 447988007, 447997006, 448027004, 448728003, 460591005, 471287006, 709597002, 721663007, 724502006, 783161005, 12237951000119105, 15648771000119103, 15712041000119108, 1264234002, 55546004, 72242008, 77910001, 93055000, 199194006, 210899002, 218728005, 253621008, 253696009, 302910002, 312826009, 399094007, 404223003, 416377005, 703246003, 4611000119106, 5501000124109, 455031000124105, 16011791000119107, 13194006, 17993000, 21379009, 28574005, 48273005, 65498003, 80235008, 87343002, 204456001, 210849001, 238768003, 253703004, 429673002, 448842002, 449926001, 721662002, 733325006, 735854005, 293441000119104, 368351000119106, 12237831000119108, 16011351000119103, 7089008, 75053002, 128321003, 175476007, 200299000, 204189006, 213220000, 234136009, 234148007, 307313003, 308129003, 309407004, 371805005, 399932006, 439447009, 460583001, 737243006, 871669005, 12720001000004100, 16697571000119103, 1144797000, 1149218009, 1153544008, 1251390005, 330101000119101, 25878000, 66173002, 111694003, 234025000, 234067009, 249658006, 253699002, 253708008, 282785008, 308136002, 315033000, 399969009, 448181004, 449439007, 697924002, 726031001, 15649141000119104, 16002071000119104, 843433951000119102, 1187633001, 1254661005, 1263731004, 6331000, 21981000, 35162007, 37639005, 60601008, 83916000, 92124001, 123276005, 174802006, 195254008, 230735006, 234054006, 236490006, 238763007, 254745007, 429064006, 448330004, 448747000, 460963006, 707200002, 788868003, 329521000119106, 1155697006,

1208858006, 1237566006, 7505006, 10129005, 35265002, 60366008, 111199007, 195408002, 234157001, 285752007, 438773007, 871656006, 455841000124109, 10749961000119104, 21877004, 1144765000, 1145183007, 1197683008, 10938511000119106, 7169009, 21234008, 33997006, 44437005, 48868008, 175145006, 186893003, 210854005, 213045007, 231828003, 234016006, 253330006, 253625004, 253631001, 262940009, 427775006, 448328001, 461418005, 472757001, 762252001, 132131000119102, 285381000119104, 15640221000119106, 15648731000119101, 16007751000119109, 53868003, 81121301000119109, 1255256006, 5499009, 21112004, 24366001, 35979000, 76805007, 93392005, 128566008, 175339005, 186317009, 254790003, 300917007, 403389006, 422233000, 423462008, 448356006, 461345005, 461415008, 762651004, 133971000119108, 328511000119109, 431411000124106, 433971000124107, 15712201000119101, 16066731000119102, 314019007, 16011951000119104, 1204183003, 1197033002, 1208898008, 1236755008, 23913003, 26660001, 48520006, 86341008, 95461006, 123641001, 195279006, 200297003, 204306007, 230482003, 234142008, 234240008, 236757005, 253668003, 445222002, 445237003, 450308009, 461385004, 461427006, 471286002, 472128009, 703193000, 703281001, 703386009, 721463003, 723873001, 724097003, 724440007, 724599009, 735419009, 735574006, 735575007, 735577004, 766949003, 870566003, 123411000119106, 430721000124101, 461431000124108, 233948000, 104346171000119109, 10645231000119108, 1236986009, 11399002, 23953000, 45281005, 95578000, 174809002, 192772009, 195326003, 234194005, 253324006, 280111008, 281596000, 297143008, 307401009, 446491006, 723868007, 723878005, 871651001, 350341000119102, 15648361000119108, 15711801000119108, 16002751000119106, 16279401000119108, 1204188007, 13957003, 43164009, 89142007, 111021003, 119820001, 233945002, 234242000, 238984005, 447668005, 448622009, 460608003, 721413008, 430731000124103, 10760131000119108, 15712161000119106, 16001711000119101, 16014511000119107, 11842931000119105, 1204177001, 2495006, 36110001, 76975009, 209146005, 210838007, 234202005, 309408009, 402832006, 405560005, 445322004, 449146005, 697920006, 722020006, 124051000119100, 337421000119103, 450471000124106, 16218291000119100, 1204186006, 1196903001, 45456005, 59047005, 77278008, 239206006, 253305008, 426856002, 449452002, 449870006, 450313008, 461383006, 721465005, 722643003, 725138002, 286441000119107, 15631861000119105, 16709811000119106, 783787000, 1010625007, 1254751008, 62703006, 93694003, 94187006, 195258006, 204451006, 209174000, 237897009, 307312008, 410468007, 447926008, 448599000, 448646002, 460959004, 703307003, 721178001, 21000119103, 139011000119104, 285541000119101, 15649581000119104, 16025591000119107, 293671000119109, 17567001, 20735004, 23202007, 23819000, 76846002, 84884003, 91489000, 93699008, 95286004, 119797004, 200310004, 208029001, 210911004, 232966002, 233967001, 234119001, 243286001, 247136003, 253672004, 300920004, 312829002, 423279000, 446668003, 449576007, 459165008, 703278006, 703312002, 733326007, 735726008, 737200003, 783258000, 788896009, 31211000119101, 132141000119106, 136751000119108, 12237151000119108, 16012671000119108, 16711631000119103, 1179365005, 43658003, 58729003, 63795001, 64042000, 85859006, 86256001, 195317001, 233950008, 233975007, 278489004, 302878004, 371069001, 402869009, 428507003, 450315001, 460595001, 460923005, 609459003, 724429004, 724444003, 338041000119100, 289566561000119108, 1204217000, 14246007, 60091004, 195520008, 198057005, 200232006, 230722002, 234040003, 239940004, 253689007, 254757003, 266263009, 449467003, 460588005, 721689007, 724535001, 725157006, 733226009, 784353002, 870350001, 117051000119103, 285161000119105, 455831000124104, 15649781000119105, 15712601000119103, 27937002, 1235003006, 40198004, 61812003, 66858001, 90085009, 95557003, 194387002, 204461004, 211236007, 234002001, 371075005, 403968005, 429542009, 447703009, 447903001, 448651008, 461602009, 707412000, 840455002, 97761000119105, 293451000119102, 11800771000119100, 15710481000119107, 15967301000119102, 15972541000119104, 19830001000004106, 16528221000119104, 1153633006, 1204141008, 1208917003, 1237561001, 18058007, 30542005, 37153009, 93061002, 111354009, 111432008, 200311000, 210834009, 373606000, 443887000, 445284003, 461371009, 703208005, 703271000, 721515003, 285361000119108, 285391000119101, 285481000119107, 429221000124104, 1087161000119105, 12237031000119103, 31235003, 17890003,

46276006, 53751009, 54519002, 82985000, 111322000, 195422002, 200224003, 232043000, 234127005, 253718003, 280980001, 314187004, 314266000, 461434008, 721191000, 723875008, 735578009, 840608004, 860683004, 679531000119103, 11011871000119101, 16837521000119102, 895085001, 1145198009, 1156027008, 12853006, 18796000, 24596005, 38059007, 81577001, 93031005, 93056004, 94319009, 128608001, 186562009, 195272002, 195374003, 200218009, 210833003, 230717002, 234005004, 235846002, 280981002, 307395009, 373408007, 472779006, 776416004, 787162002, 15649301000119103, 15708441000119103, 16288341000119100, 896690007, 16023711000119105, 23115009, 41486008, 87796008, 128053003, 163052007, 200253002, 234156005, 238838004, 253700001, 285750004, 426711003, 431706008, 461619006, 472747002, 762253006, 11827061000119103, 12237111000119107, 15966461000119108, 15967181000119108, 1144957009, 1231140009, 40193008, 59600000, 85787009, 89748001, 95449003, 232041003, 285753002, 396779001, 428781001, 448096002, 717881002, 734383005, 762632005, 838314000, 12851000119104, 329561000119101, 429251000124108, 15978631000119109, 16012511000119101, 403674007, 16024351000119108, 1217645003, 1269245002, 7387004, 66131005, 91605000, 234074004, 261537001, 268190004, 307404001, 363261001, 424699007, 429245005, 722642008, 722986000, 868227005, 871650000, 338031000119109, 448841000124100, 15707921000119104, 16012711000119107, 3275009, 1204212006, 1209060000, 1269057003, 4473006, 8901009, 52156004, 125964004, 195289005, 210869006, 253669006, 285739002, 297151006, 302930003, 439388005, 447913009, 449514000, 449523002, 460885001, 461286001, 461409008, 697922003, 783011004, 840315000, 840441005, 871664000, 5611000124108, 153811000119105, 285371000119102, 15711601000119107, 16002231000119106, 16026351000119101, 16891151000119103, 420308006, 1230062001, 163018009, 204395001, 210870007, 234024001, 314014002, 400034001, 417303004, 448059006, 448631009, 449871005, 782698000, 786055002, 134961000119104, 285171000119104, 15631691000119102, 15711441000119107, 16006191000119103, 677621000119105, 15707961000119109, 876859003, 15986031000119105, 1254873008, 16662411000119105, 12617007, 73757007, 195340002, 200990006, 204467000, 235855004, 253683008, 306857000, 314184006, 403443000, 405556007, 447260005, 449185003, 449430006, 461567005, 721462008, 285581000119106, 15648561000119102, 16711031000119109, 16846341000119101, 1144805008, 610412651000119100, 1208768006, 903008, 54122009, 62583006, 195313002, 200222004, 234037003, 240484000, 425365009, 432461000, 446558004, 733126003, 122061000119109, 455851000124106, 677211000119101, 15648601000119102, 16025671000119105, 11816021000119102, 1209115004, 1269302007, 5534004, 26421009, 37787009, 61687004, 62727008, 72092001, 234082004, 234197003, 235000001, 239935007, 253678000, 312594005, 312913006, 449184004, 461438006, 461577007, 472751000, 721455004, 15649741000119100, 15712481000119108, 49497005, 16013191000119107, 878858004, 1149472000, 1153607003, 1208818009, 15978511000119100, 13316004, 25412000, 43259003, 54866009, 67682002, 89980009, 126757004, 195281008, 195426004, 233988005, 233989002, 234001008, 234147002, 253347007, 400017009, 402831004, 446140003, 711164003, 720626009, 721688004, 737159004, 773993008, 783730009, 846667001, 285351000119106, 13730001000004107, 15711761000119100, 16026071000119107, 94081000119107, 1155723008, 1251350007, 44647001, 53312001, 73173006, 86252004, 87394009, 128584005, 195319003, 230468003, 237347000, 252986008, 253682003, 399972002, 444816006, 448479008, 449350006, 709602009, 717864006, 735725007, 10759311000119104, 15648861000119100, 239921005, 16018991000119103, 1155699009, 16012311000119106, 11858741000119104, 1204175009, 331061000119107, 1237610002, 682004, 17337006, 37139009, 43306002, 52757001, 58309001, 195342005, 208057008, 232039004, 234084003, 253660005, 297152004, 312828005, 445486004, 447939007, 448065006, 449516003, 718614004, 721226005, 737324009, 15711081000119107, 16002551000119103, 16624001000119104, 1153545009, 98067641000119101, 1269056007, 49778009, 62893006, 72150009, 93817006, 95579008, 233936003, 247213004, 280144002, 361133006, 424728002, 443210003, 448179001, 449494004, 460582006, 15711641000119109, 15712641000119101, 16006071000119100, 1204215008, 302711000119108, 1220630007, 677721000119103, 1237133002, 7369002, 10451007, 31996006, 79917009, 128218002, 209067008, 233937007, 234068004, 238773009, 238785001, 238810007, 306851004, 385442006,

403606008, 409506009, 449398002, 450302005, 460592003, 471299004, 473364007, 724602004, 783701002, 143661000119109, 11858381000119105, 16019471000119104, 285661000119101, 1204126008, 291421000119106, 6536009, 17920008, 26900001, 59568004, 62335009, 95242009, 199192005, 253712002, 360455002, 371051005, 403516008, 447901004, 460966003, 703355003, 717003001, 733127007, 7071000119102, 16012911000119105, 421895002, 1153546005, 1204219002, 58361001, 70602002, 75409000, 81387001, 88755007, 92002004, 95471008, 204464007, 253664001, 254782003, 266329009, 312375001, 359557001, 399866003, 432119003, 442224005, 472781008, 699073004, 734964005, 870637009, 136771000119104, 290731000119106, 15710681000119105, 15966901000119100, 16007631000119106, 16014431000119101, 1153631008, 1209061001, 331011000119109, 1269257006, 3681008, 20656007, 89420002, 200983001, 234041004, 253685001, 312496009, 414391001, 448742006, 707668000, 5471000124102, 132291000119106, 133421000119101, 11830361000119103, 14640001000004103, 16014631000119103, 1254663008, 52496006, 82196007, 93471006, 111319002, 194821006, 200258006, 213048009, 233961000, 238775002, 253315002, 399228007, 416770009, 449015009, 460607008, 703176000, 840434004, 77731000119106, 132321000119103, 15631781000119105, 16289381000119103, 1197687009, 15711281000119101, 188001, 17355002, 22883003, 33700007, 58142005, 195277008, 195423007, 200286003, 204427001, 239202008, 253728007, 262944000, 400054000, 402648009, 444855007, 447860000, 448060001, 449651003, 460605000, 460899002, 461422000, 471298007, 698363002, 712582008, 725002006, 733223001, 735567001, 770599000, 782773005, 5451000124107, 5511000124107, 145611000119107, 285631000119109, 343811000119102, 15708281000119109, 16012591000119105, 16011911000119100, 1204185005, 1204191007, 1251423001, 5050001, 56349004, 77892009, 195343000, 234078001, 239204009, 247135004, 277322008, 297176007, 308134004, 371040005, 403510002, 427490006, 428870007, 443502000, 712578006, 714181006, 734298005, 765055004, 789672007, 5651000124109, 16711071000119107, 878857009, 1156031002, 16019311000119103, 972604701000119104, 10645631000119105, 1237129009, 11844671000119105, 11617005, 16694003, 51315008, 53330008, 61099007, 74341002, 126737003, 127317007, 193039006, 194892009, 253624000, 255190002, 271985009, 445296005, 446836009, 449025004, 471297002, 473393007, 724428007, 285471000119109, 16006271000119105, 1204216009, 15997991000119107, 1231662005, 10057001, 15920007, 43331001, 55382008, 67081008, 87239004, 128062001, 204423002, 234207004, 312380005, 312998003, 404668004, 425435003, 717866008, 721412003, 721706007, 840313007, 285311000119105, 15711161000119104, 16013151000119102, 15969701000119108, 1208844009, 10643231000119109, 16418006, 28545001, 46126003, 62827000, 65785006, 210811000, 210841003, 210862002, 210894007, 233986009, 234021009, 234052005, 253313009, 253697000, 254744006, 448332007, 723874007, 435281000124100, 11798951000119105, 15649821000119100, 16007871000119108, 359789008, 399957001, 13886001, 15971009, 16972009, 195476002, 200209007, 206421002, 236488005, 253323000, 426270006, 426948001, 448620001, 461110009, 711546000, 723867002, 734965006, 765750001, 789690008, 132301000119107, 1208760004, 1209119005, 10643831000119105, 95620003, 193379006, 233170003, 233990006, 234049002, 247212009, 253646008, 297137006, 312601003, 699876008, 710944000, 331801000119102, 455901000124101, 15712241000119104, 16711551000119108, 856856651000119109, 1254665001, 7368005, 34492003, 85719003, 93064005, 192753009, 210827007, 243410003, 277329004, 405557003, 427508005, 428674000, 448063004, 449560003, 457597009, 461102008, 461407005, 721328009, 27631000146108, 436041000124107, 11830441000119109, 15649541000119109, 15711401000119105, 15972581000119109, 16005471000119100, 16715971000119108, 16520041000119104, 56384000, 66877004, 75145007, 129212008, 195448005, 213131003, 232965003, 233956002, 234126001, 234141001, 234193004, 308805008, 399884001, 402958005, 439160009, 460597009, 703245004, 721703004, 448831000124105, 16006111000119107, 399868002, 890209003, 1155686006, 15968901000119104, 1162761001, 1197363004, 44791000087107, 16652001, 53190001, 63077009, 77773000, 209194009, 232357009, 280976006, 297162006, 402840000, 403393000, 448614002, 721411005, 788454002, 789671000, 789700005, 431061000124100, 431431000124100, 10759761000119100, 15710601000119102, 16025551000119102, 16624151000119101,

56231002, 1208875000, 1237609007, 1055001, 22750001, 28670008, 46956008, 72127003, 75543006, 195475003, 210828002, 210853004, 230224002, 233964008, 253314003, 308546005, 309783001, 459180002, 460906001, 460960009, 472138004, 871498004, 15960661000119107, 16477701000119105, 16019431000119102, 1204182008, 14534009, 15191001, 16750002, 41740005, 58711008, 210848009, 210851002, 233980003, 297150007, 312827000, 400042000, 448016001, 448120000, 448745008, 5251000119108, 124041000119102, 135061000119104, 285331000119100, 429831000124100, 436771000124102, 15648481000119107, 1162504009, 11858501000119106, 34233009, 40136003, 45650007, 193030005, 210786006, 235052000, 253704005, 302946006, 307322002, 448098001, 472705001, 699706000, 703267003, 733282009, 762648006, 15710241000119103, 1231168008, 1251415003, 20934006, 54636000, 94461002, 200288002, 209189009, 233815004, 253636006, 267284008, 285711003, 297146000, 427419006, 430332005, 438483005, 460387004, 703335004, 971000119105, 132541000119101, 455791000124106, 1082711000119108, 15649621000119104, 15708201000119101, 15711681000119104, 15712121000119101, 16026231000119100, 879814007, 1153608008, 1220652003, 677731000119100, 1269555002, 56515006, 95448006, 134367001, 234208009, 238787009, 268284006, 276392006, 402861007, 405585001, 461420008, 860682009, 285571000119108, 454031000124106, 15648641000119100, 1149060002, 1204128009, 1204378001, 11800411000119103, 45237002, 84150000, 134399007, 195341003, 210846008, 234065001, 253614008, 253726006, 306850003, 419944002, 421365002, 459179000, 461390001, 472702003, 697897003, 702385003, 713033007, 723363009, 455801000124107, 15960381000119109, 15973181000119106, 1263548009, 1279533009, 20956009, 21344007, 92994009, 112819006, 200215007, 232024000, 239925001, 297153009, 302968001, 403518009, 448727008, 450305007, 460961008, 698627005, 703266007, 871596002, 329491000119109, 15710561000119102, 15710761000119109, 195295006, 254775002, 876856005, 99591000119101, 16662451000119106, 8069005, 80606009, 111321007, 195323006, 402675002, 442267002, 447318008, 460437005, 461592008, 472755009, 609489005, 697905000, 720512007, 840306007, 871598001, 871614007, 96141000119105, 136781000119101, 431401000124108, 15648281000119108, 16011751000119102, 1149069001, 11795201000119108, 1204181001, 331051000119105, 1269054005, 1269239005, 2477008, 51496009, 57981008, 68092007, 94857002, 127318002, 238778000, 253729004, 301432000, 304026004, 440441004, 445371009, 447528003, 448721009, 697903007, 702617007, 783419007, 840713005, 15709841000119107, 16007791000119104, 16012271000119106, 44761000087104, 12075007, 15258001, 15742000, 20059004, 192769002, 210881005, 210902004, 266253001, 266330004, 276475003, 461410003, 455931000124109, 13720001000004105, 15712521000119108, 16013711000119104, 78862003, 11858621000119103, 1204197006, 10645031000119104, 15708321000119104, 1034003, 9177003, 28773001, 37876005, 55976003, 66829004, 230734005, 232750009, 234070008, 234073005, 239928004, 275446004, 297141005, 297218007, 423062001, 445440001, 447269006, 448975006, 459164007, 461439003, 860677002, 285151000119108, 285251000119101, 293461000119100, 677801000119100, 11844991000119103, 16002031000119102, 16004911000119101, 95501007, 1156246002, 1204195003, 1208942007, 1285323003, 2818004, 67815006, 70405009, 93318005, 126738008, 210892006, 233817007, 233976008, 233982006, 253658008, 403560008, 448783009, 459183000, 472760008, 706870000, 709582000, 721382008, 722852005, 724441006, 838275008, 868231004, 5481000124104, 27651000146102, 32711000119101, 132251000119101, 293471000119106, 343801000119100, 430861000124102, 15708401000119100, 1204220008, 1255264000, 9250002, 10365005, 51096002, 63739005, 75878002, 85514005, 90858003, 92961006, 200255009, 230220006, 254211001, 419310003, 428171009, 445106006, 609457001, 703311009, 721388007, 436591000124109, 15710801000119101, 361278002, 1230397006, 31455004, 51723007, 70153002, 72866009, 128191000, 164626005, 199193000, 253649001, 262939007, 277316004, 297138001, 301755001, 312822006, 414024009, 420006002, 449433008, 460964000, 461111008, 472759003, 472792009, 132241000119103, 285611000119104, 455811000124105, 677201000119104, 677811000119102, 15966501000119108, 16415501000119100, 16837401000119105, 1148583001, 1284863004, 6715005, 21591004, 72466000, 112815000, 195321008, 233972005, 253722008, 275700003, 445350004, 461572001, 471290000, 472706000, 703334000, 705066004,

721457007, 770581008, 868221006, 871665004, 329571000119107, 350321000119108, 436031000124102, 16019031000119105, 1156984001, 11858701000119101, 1209117007, 23808003, 29931007, 38739001, 40929003, 77558004, 235843005, 240483006, 254998002, 270510008, 307399003, 447663001, 461105005, 471285003, 703295003, 16012831000119109, 16711111000119100, 698190002, 1204180000, 20427003, 32112006, 36523002, 52674009, 72930009, 193474002, 239207002, 253640002, 280871000, 373096006, 408666009, 447930006, 472101004, 840422007, 429861000124109, 1087811000119103, 15735321000119104, 876857001, 1197432002, 1237565005, 46708007, 65617004, 73990001, 76402003, 87224000, 111290000, 195213000, 195324000, 195397000, 233978009, 264258007, 264557003, 271376002, 281597009, 297160003, 432293006, 442105001, 445176007, 447110008, 457592003, 713825007, 721384009, 783729004, 870544005, 329631000119108, 677821000119109, 16006311000119105, 1010655000, 15970021000119103, 880044007, 1260127003, 1268522004, 11029002, 14540002, 52984008, 70607008, 76097009, 111028009, 198750004, 212372004, 234012008, 234158006, 302857002, 307403007, 312937006, 442701004, 460944005, 697902002, 721458002, 770112002, 430841000124101, 677681000119109, 239922003, 880043001, 1148587000, 1231666008, 1237532002, 1255187002, 16934004, 25263003, 37325002, 79671001, 93312006, 128052008, 232046008, 234200002, 285709007, 402850004, 404667009, 447832002, 448472004, 449232001, 450304006, 471268000, 765812004, 868223009, 106021000119105, 449691000124103, 677591000119108, 12237471000119109, 15699321000119102, 16014351000119101, 1208916007, 11824561000119103, 213304008, 237346009, 253628002, 267283002, 315348000, 371067004, 428110002, 442439008, 446029009, 447823004, 448510002, 450300002, 460375006, 461107002, 461326001, 697917003, 720583008, 721511007, 723877000, 788455001, 789668008, 5461000124109, 285421000119108, 1172565007, 1204189004, 35830007, 38261007, 74725000, 134193000, 210880006, 236094005, 247124009, 280964006, 428479004, 429561008, 438647008, 444036008, 445209005, 447849009, 447924006, 449119000, 697908003, 703284009, 707414004, 721386006, 722004001, 734880004, 838364007, 11858421000119101, 16002191000119102, 16279441000119105, 10644031000119103, 1260330000, 845006, 8435000, 26150009, 38905006, 44902004, 76276001, 82153002, 86003009, 163024003, 196999001, 200301007, 297148004, 304090004, 403391003, 431545001, 433068007, 461388002, 461401006, 721512000, 21631000119105, 285461000119103, 455911000124103, 15648921000119102, 16012551000119100, 54034003, 1251387004, 1254657004, 56238008, 175142009, 212364001, 233940007, 234051003, 254778000, 314265001, 447418009, 447702004, 449650002, 472791002, 473362006, 699135001, 703286006, 721705006, 723876009, 285501000119103, 450781000124101, 15978151000119108, 879816009, 1156029006, 16288381000119105, 1237560000, 52576001, 59879002, 90458007, 111295005, 230731002, 237342006, 312592009, 312914000, 403619007, 448723007, 460538002, 461413001, 698783004, 713265001, 735565009, 783421002, 78741000119103, 15710401000119104, 16025511000119103, 11800211000119104, 7438000, 24638001, 198650006, 200290001, 253632008, 280971001, 285759003, 302945005, 425963007, 447661004, 461293002, 733128002, 860676006, 143561000119108, 286391000119108, 12237631000119109, 15631821000119100, 15712561000119103, 16066531000119106, 16289221000119108, 1156729001, 1208854008, 1255189004, 1269246001, 4692001, 50751005, 50821009, 109381003, 128060009, 128064000, 175144005, 195180004, 248741001, 360578005, 446890001, 448304003, 449551006, 460590006, 715364001, 724436003, 734327009, 860688008, 285561000119102, 432261000124102, 679521000119101, 15966701000119103, 15967141000119103, 310701003, 15952021000119101, 879815008, 26743002, 36184004, 44622003, 59093008, 74883004, 82889007, 230225001, 230728003, 234045008, 253341008, 253715000, 269451001, 297175006, 461352007, 473446004, 723336008, 723859005, 786050007, 789097007, 132201000119100, 450771000124104, 11816101000119105, 15707881000119106, 16014471000119103, 232030000, 1078223005, 1208919000, 26954004, 60555002, 74218008, 195271009, 204460003, 210840002, 230733004, 247210001, 252036000, 253326008, 280974009, 306847001, 371160000, 403957005, 445285002, 461366002, 763615003, 764100007, 100751000119101, 286431000119103, 291471000119107, 10835781000119105, 15648821000119105, 880037003, 1204150005, 12065811000119106, 16023631000119108, 34742003, 42486006, 95456009, 230730001, 234154008, 247127002, 249633001,

280966008, 280969001, 314000002, 429304004, 430301009, 440444007, 443971004, 445167000, 449547009, 461399000, 763714006, 90971000119103, 1092941000119109, 11816181000119102, 11845191000119102, 12237871000119106, 15708521000119108, 16012031000119104, 1230182004, 1208762007, 1208855009, 1255621004, 36675003, 58144006, 63363004, 64933009, 87825006, 88032003, 90493000, 95447001, 95653008, 127014009, 192761004, 210826003, 230716006, 234071007, 247100001, 314902007, 403508004, 427368007, 448061002, 460603007, 460937001, 461417000, 472761007, 735573000, 1395000100004103, 15649361000119102, 253514004, 1144956000, 16662531000119101, 1197364005, 1209174005, 1269243009, 1279575005, 195373009, 200260008, 233965009, 297155002, 426651005, 433844000, 698816006, 725958000, 766960007, 783733006, 838308007, 25981000119102, 15708001000119106, 15966341000119102, 1149219001, 1234823004, 88065006, 93385000, 195382003, 206576006, 233954004, 399923009, 403545005, 403569009, 403976007, 447700007, 460531008, 703299009, 734397002, 737325005, 846684007, 860689000, 871671005, 15997911000119103, 16697061000119100, 1204147007, 1196852001, 4240001, 11516009, 22323009, 84216001, 88223008, 210868003, 213218003, 213309003, 234131004, 302879007, 403616000, 403618004, 405539004, 422166005, 448281007, 449434002, 11830521000119104, 1732000100004103, 1255623001, 14223005, 15842009, 23687008, 75955007, 80901002, 87837008, 195518005, 209093001, 213198007, 234047000, 238782003, 361264003, 402657003, 402836009, 448067003, 460953003, 721105004, 721160006, 781126009, 455821000124102, 690791000119107, 16014791000119105, 32890004, 32916005, 46395002, 65593009, 95644001, 95655001, 95660002, 112813007, 234203000, 253198006, 253343006, 280977002, 439129009, 445109004, 448116004, 460930004, 472780009, 699301008, 715644000, 721387002, 737191003, 781130007, 785808002, 840436002, 433951000124102, 450791000124103, 677771000119102, 11795281000119100, 16002271000119109, 16011431000119103, 16624051000119100, 1144955001, 677711000119105, 16750471000119102, 1269058008, 16066851000119103, 13507004, 54798007, 93033008, 123803006, 195210002, 195232006, 209098005, 232369001, 233973000, 234061005, 247137007, 251024009, 253195009, 253707003, 267280004, 285708004, 431530008, 735855006, 783716004, 871582008, 8051000119105, 122731000119104, 286381000119105, 329581000119105, 332421000119101, 350301000119104, 431571000124103, 451041000124103, 12237431000119106, 1342000100004101, 1144914004, 1153543002, 1204156004, 1197417009, 1231726001, 26780008, 57363000, 61574004, 64832003, 68504005, 195183002, 195268001, 233970002, 234043001, 234125002, 276667003, 302213007, 384681005, 416703007, 427567003, 428609006, 447814002, 707476006, 709687000, 710864009, 720575002, 721516002, 840308008, 94091000119105, 286331000119109, 15711241000119106, 2582000100004100, 69742007, 1222546002, 1255188007, 1258884008, 85664008, 128952006, 194842008, 204299009, 204391005, 235842000, 239938009, 253727002, 265506006, 371894001, 443264000, 446712002, 447498008, 448722002, 449442001, 724597006, 762049002, 783409001, 868228000, 870323006, 429841000124105, 16013351000119104, 23186000, 28578008, 37943007, 38921001, 64959007, 129573006, 195211003, 195339004, 204501003, 253615009, 371158002, 402867006, 449120006, 450307004, 458039003, 460585008, 460890003, 461411004, 718602007, 789681001, 871605002, 135071000119105, 16002711000119105, 46041001, 402842008, 31268005, 49956009, 57035005, 402834007, 417444002, 447970004, 450309001, 460438000, 699529005, 722867009, 734299002, 132171000119104, 284881000119108, 16007591000119106, 16026391000119106, 57348003, 16013071000119105, 1187508009, 1197664001, 1255257002, 8281003, 41017003, 68478007, 78129009, 200208004, 238811006, 251045002, 253648009, 253676001, 253677005, 253691004, 254208002, 307397001, 408665008, 413577001, 448278002, 448968005, 703329004, 735767003, 840500002, 677831000119107, 1144964006, 1149473005, 1237131000, 16606801000119105, 7930004, 21604008, 50808002, 60787001, 75533008, 90257004, 95452006, 195233001, 195438008, 204463001, 204493007, 209049001, 210912006, 234062003, 236492003, 253311006, 253642005, 307315005, 312377009, 312916003, 447840008, 448635000, 460586009, 461400007, 707665002, 721466006, 721704005, 735834009, 15711921000119107, 1155688007, 471651000124106, 51147005, 55275006, 67119003, 119762001, 127276009, 195411001, 204433005, 210814008, 232035005, 234159003, 253654005, 314267009, 361210001,

402844009, 403651008, 447689001, 447861001, 449443006, 449831000, 461096001, 472768001, 723862008, 735420003, 430831000124106, 15929941000119103, 329501000119102, 1237569004, 11454006, 51551000, 60106004, 91304009, 128211008, 204296002, 209078000, 230721009, 234063008, 239205005, 402656007, 423683008, 445286001, 447322003, 461624009, 722705009, 763747002, 871647003, 15710521000119107, 15960141000119102, 16013111000119103, 1148690000, 1153630009, 57357009, 71890008, 111497000, 128059004, 253630000, 253653004, 286959000, 403659005, 444744001, 449271003, 450314002, 471291001, 709584004, 124061000119103, 677221000119108, 677611000119103, 11798991000119100, 12237511000119100, 1010641008, 1162974001, 1208675002, 7870007, 15400003, 51677000, 56272000, 67843002, 72252007, 88244008, 90958004, 112812002, 128065004, 195320009, 200284000, 233943009, 234020005, 282664001, 297140006, 403856000, 697925001, 723871004, 43691000119105, 11795321000119105, 12237911000119109, 15960581000119102, 16013871000119104, 1156016006, 302681000119107, 1237558002, 11242008, 12232008, 29934004, 54687002, 56780006, 69954004, 210866004, 233183002, 233987000, 234192009, 236759008, 285712005, 427592000, 432294000, 460324008, 697896007, 840437006, 860699005, 871578006, 10759191000119106, 15710361000119108, 15711721000119105, 1204170004, 1204144000, 2534005, 19472003, 36304008, 52539004, 95451004, 128609009, 198029003, 213044006, 238764001, 257812009, 308140006, 402847002, 445351000, 449125001, 721414002, 735131004, 132191000119103, 16011991000119109, 16696981000119105, 1197695008, 10645831000119109, 11781801000119106, 1255266003, 486006, 12530001, 32413006, 34173009, 68087005, 194883006, 195474004, 253328009, 253706007, 254209005, 275510005, 285751000, 309405007, 313347007, 402841001, 427602004, 10754331000119108, 16013031000119107, 16662331000119106, 45965006, 307816004, 1254660006, 20702009, 26373009, 40450001, 48248005, 64586002, 73699003, 75879005, 111289009, 128056006, 190815001, 210843000, 234150004, 254788004, 262715008, 262943006, 281595001, 444856008, 449536002, 707667005, 135001000119100, 135011000119102, 150721000119102, 285141000119106, 15648441000119102, 302701000119105, 1209120004, 1748006, 25772007, 36665001, 55168004, 73067008, 195318006, 200206000, 210787002, 210896009, 234056008, 239926000, 242001005, 302881009, 439407002, 445436005, 461109004, 733129005, 764521002, 126531000119109, 15966781000119106, 51696001, 1251567001, 16023831000119107, 41114007, 45640006, 59631007, 90520006, 91993000, 210897000, 449010004, 713032002, 713193007, 716590006, 733263000, 735566005, 735898005, 871663006, 286451000119109, 15711001000119104, 943003, 6729006, 10058006, 10964002, 64583005, 68708005, 80829003, 86191000, 195280009, 200217004, 210844006, 253307000, 253716004, 306856009, 313350005, 447660003, 448005008, 448499002, 449905001, 471289009, 697926000, 703636009, 735132006, 286071000119109, 431421000124103, 15944861000119108, 253515003, 1052210007, 11844831000119106, 1237559005, 7014009, 47432005, 56683001, 83330001, 95474000, 119798009, 127316003, 195206000, 213306005, 253720000, 307320005, 402843003, 405564001, 448517004, 449445004, 460967007, 472754008, 705129007, 734960001, 15648521000119107, 16014591000119103, 1196861001, 1237254001, 14977000, 32477003, 35191005, 43910005, 48974009, 70933002, 84939004, 88527005, 93054001, 95236005, 163022004, 195229008, 210839004, 233999007, 371414009, 423894005, 448158001, 460471001, 721238001, 782823001, 789064006, 680841000119109, 16007711000119108, 16289261000119103, 19431000, 1204218005, 11816461000119104, 21030008, 88722002, 91109007, 192771002, 233955003, 253327004, 253635005, 307408003, 307409006, 315295003, 444549009, 460955005, 461359003, 461436005, 609458006, 697918008, 789669000, 40241000119109, 691471000119109, 11799771000119107, 697900005, 471661000124108, 15985991000119104, 1208676001, 1254659001, 1255569006, 5245002, 32194006, 62702001, 66816004, 70995007, 71362000, 93328001, 163055009, 195325004, 232974001, 234155009, 307318007, 312383007, 359936002, 373434004, 447701006, 448628008, 449425007, 460587000, 460589002, 703385008, 703533007, 838311008, 860678007, 132271000119105, 285191000119103, 329481000119106, 436021000124100, 455921000124106, 15710841000119104, 16019511000119108, 1197429000, 26865008, 57734001, 76211000, 92965002, 110283003, 128557009, 200237000, 234048005, 253679008, 277493007, 307317002, 403775003,

405768001, 460584007, 460610001, 735833003, 1196899008, 1197696009, 44771000087108, 2079001, 3506008, 21258007, 45669002, 52303006, 87050008, 91445000, 128055005, 174808005, 210898005, 234151000, 253194008, 276594006, 307221007, 413218001, 426907004, 460370001, 460604001, 461425003, 472788002, 763867001, 766948006, 291351000119109, 350311000119101, 15972501000119101, 16012151000119109, 16025791000119100, 16289341000119108, 2085000100004108, 26146002, 1153632001, 1258934002, 15946006, 17593008, 35350000, 61599003, 83256006, 195406003, 233962007, 280967004, 280978007, 314185007, 425414000, 438961001, 458432002, 459066006, 460517008, 472103001, 697913004, 762649003, 868216008, 1001000119102, 12237711000119106, 15649101000119101, 15711041000119102, 1169367003, 1204178006, 1208822004, 1255622006, 1367008, 10460004, 18919000, 31019002, 71444005, 87937009, 200308001, 232973007, 233984007, 234050002, 253714001, 363315004, 417019000, 447925007, 448095003, 448153005, 448486000, 448905005, 702374000, 721467002, 735114006, 737157002, 870579007, 124161000119104, 11800811000119100, 15649041000119103, 15708361000119109, 16025831000119106, 1153611009, 58341661000119101, 128129006, 163986001, 200233001, 230741004, 234128000, 237900002, 239929007, 253358003, 254997007, 438510004, 448746009, 448965008, 721460000, 723870003, 737332001, 783731008, 788290007, 9611000119107, 285341000119109, 15648321000119103, 15966741000119101, 16705611000119108, 1204162009, 1204160001, 1217005008, 1208852007, 5166001, 5946006, 43696002, 47040006, 54771008, 59292006, 64672005, 94755004, 155441006, 195404000, 199195007, 200216008, 239297008, 248742008, 253629005, 275518003, 297156001, 308133005, 371041009, 399216004, 438476003, 448028009, 449009009, 449458003, 459173004, 460594002, 766955008, 871661008, 133031000119106, 286341000119100, 1208847002, 5201002, 28790007, 39905002, 41991004, 200303005, 233993008, 275359008, 287731003, 312584000, 444657001, 447691009, 448079003, 448115000, 704142001, 724856009, 783773000, 285431000119106, 336191000119105, 10690551000119101, 11781561000119105, 15709881000119102, 16012111000119108, 1204173002, 1237132007, 1260329005, 674571000119104, 24768002, 48601002, 49176002, 57246001, 111291001, 111458008, 197712008, 234019004, 234145005, 251047005, 270512000, 404670008, 407530004, 438477007, 443095000, 460612009, 764452004, 9631000119102, 16012991000119101, 791000119109, 1269055006, 62023000, 67962003, 93388003, 195275000, 210804004, 210859000, 234123009, 262717000, 402655006, 448480006, 448843007, 459065005, 697911002, 713036004, 718217000, 722858009, 32361000119104, 15648201000119100, 1153638002, 1208853002, 20593006, 29322000, 45256007, 239201001, 251046001, 253645007, 253667008, 253725005, 264503005, 275363001, 277325005, 280970000, 448724001, 450310006, 461381008, 470749006, 470759007, 472133008, 711435001, 713035000, 770111009, 15640181000119103, 15760351000119105, 1204193005, 1237134008, 3168002, 60993004, 118955007, 210809009, 234161007, 236489002, 238769006, 268185002, 413758000, 438581005, 441739009, 445512009, 448629000, 472789005, 284871000119105, 285621000119106, 285671000119107, 451361000124102, 11799401000119106, 11799851000119108, 15648981000119103, 1010678006, 1144953008, 1209204000, 1237568007, 14070001, 17263003, 25559009, 28931004, 39987008, 59282003, 61605005, 62838000, 92517006, 210820009, 230221005, 234003006, 234122004, 234196007, 253317005, 253641003, 253692006, 276393001, 307321009, 313254003, 402839002, 448501005, 449559008, 450312003, 460365008, 461562004, 787044009, 840310005, 139741000119102, 285601000119102, 286351000119103, 430851000124104, 15964701000119109, 1010729007, 1177015003, 1196859005, 1251422006, 17127005, 47058000, 95234008, 95580006, 109380002, 128556000, 192760003, 234076002, 234086001, 448478000, 449521000, 460611002, 460968002, 722930000, 723872006, 734041006, 773992003, 786049007, 186971000119106, 329671000119106, 11830601000119109, 15631741000119100, 15710721000119104, 16014711000119101, 897004000, 11858461000119106, 1237567002, 20001006, 29995000, 60265009, 111299004, 213305009, 238780006, 239936008, 239948006, 402658008, 402663007, 438785004, 447850009, 449288005, 460545002, 461421007, 735082004, 773996000, 10760081000119108, 46557031000119105, 15985951000119109, 1231273008, 95235009, 95656000, 128072003, 128187005, 200238005, 200294005, 234149004, 297154003, 403708002, 403959008, 421327009, 446655006, 473356001,

609497003, 699257002, 734384004, 868219001, 333281000119108, 1082631000119106, 11827021000119108, 15649681000119100, 890115009, 590005, 28104001, 54682008, 63954007, 93354009, 208064005, 238772004, 253331005, 253688004, 280975005, 297157005, 307224004, 442240008, 444325005, 449027007, 449873008, 473395000, 722006004, 724439005, 782724001, 860681002, 2831000119107, 15711121000119109, 82385007, 314015001, 1003848008, 1144932004, 1145200003, 11795241000119105, 11781841000119108, 28398002, 35386004, 58474006, 73369001, 95237001, 197001004, 210850001, 234029006, 234046009, 234079009, 253340009, 253513005, 280831005, 301433005, 432249006, 449016005, 461587006, 734374000, 789704001, 16371781000119100, 21978005, 43405004, 1156070008, 621031000124105, 1269240007, 52089001, 64290004, 75884004, 94202009, 195335005, 213037002, 238762002, 262942001, 271967002, 277196008, 311845003, 360573001, 371039008, 399982001, 400131007, 408664007, 438580006, 447876009, 788895008, 870364001, 8001000119106, 367531000119106, 455011000124104, 11018701000119109, 1230344000, 1258883002, 7305005, 24624008, 35709005, 43935004, 70195006, 204470001, 210816005, 233974006, 234143003, 238776001, 239946005, 253695008, 266519001, 313346003, 405588004, 447968008, 461423005, 472750004, 472753002, 713965007, 734961002, 771235001, 132261000119104, 285651000119103, 286421000119101, 347011000119102, 1086041000119105, 7600001000004104, 16025471000119107, 1204158003, 1252750008, 1256099004, 29057008, 50570003, 58008004, 88744007, 88859007, 128551005, 129582000, 200291002, 210895008, 230464001, 232037002, 234010000, 268184003, 313348002, 395218007, 410065004, 426568003, 449589001, 132091000119104, 1156018007, 1204192000, 1208819001, 3827008, 13867009, 42866003, 60989005, 85461008, 200252007, 210856007, 234011001, 253638007, 253687009, 267282007, 307393002, 384993003, 402835008, 403724001, 427356003, 472752007, 700151002, 724208006, 724435004, 789703007, 5591000124102, 15649481000119103, 15972621000119109, 1208673009, 1237608004

Truveta:
1215970, 1215941

#### 3.26 Psychoses

ICD10CM:
F24, F31.2, F20.9, F28, F25.0, F20.81, F20, F20.8, F25.9, F20.3, F23, F30.2, F20.89, F25.1, F25.8, F20.1, F20.2, F31.5, F20.0, F20.5, F29, F22, F25

ICD9CM:
293.8, 295.1, 295.21, 295.8, 298.3, 295.64, 296.14, 295.73, 295.52, 295.04, 295.75, 295.7, 295.44, 295.5, 295.22, 295.00, 297.3, 295.20, 295.03, 295.6, 295.94, 295.93, 295.72, 295.12, 295.30, 295.4, 297.8, 295.51, 295.43, 295.60, 295.50, 295.2, 295.45, 295.32, 295.65, 298.9, 295.13, 295.23, 295.14, 295.84, 295.54, 295.10, 293.83, 295.82, 297.1, 296.44, 295.92, 298, 295.41, 295.24, 295.81, 297.0, 293.81, 295.70, 298.1, 295.85, 295, 297, 295.11, 295.35, 296.54, 295.90, 295.83, 298.8, 298.0, 295.71, 293.89, 295.40, 295.9, 298.2, 295.15, 295.33, 295.80, 295.95, 293.84, 295.01, 295.63, 295.25, 295.05, 295.31, 293.82, 295.91, 295.62, 295.61, 295.34, 297.9, 298.4, 297.2, 295.74, 295.02, 295.42, 295.53, 296.04, 295.55, 295.0, 295.3

SNOMED CT:
231450007, 712824002, 1231503005, 14291003, 42344001, 162313000, 765176007, 698951002, 70546001, 191475009, 10760461000119107, 31658008, 68995007, 786120041000132108, 191483003, 268624000, 712850003, 1220650006, 270901009, 762507003, 724755002, 111483008, 762510005, 357705009, 58647003, 5510009, 247804008, 441704009, 1973000, 5464005, 238978007, 441833000, 51133006, 268617001,

724729003, 30336007, 191572009, 191623007, 238973003, 76566000, 427975003, 64905009, 75122001, 191527001, 724655005, 65179007, 302507002, 27387000, 724696008, 17262008, 191494004, 724759008, 68890003, 111484002, 191531007, 129602009, 21831000119109, 288751000119101, 111482003, 723901004, 6348008, 61144001, 231451006, 88975006, 719717006, 12939007, 84760002, 268612007, 14144000, 724702008, 1231846001, 723899008, 724675001, 191574005, 723927004, 191569002, 1204417003, 237352005, 191548004, 762508008, 1259128002, 36158005, 85861002, 33323008, 191478006, 280949006, 15921731000119106, 231437006, 78640000, 191680007, 54502004, 70814008, 191554003, 55009008, 307417003, 47664006, 69482004, 191668004, 191565008, 50722006, 61831009, 31081000119101, 762325009, 63395003, 274953007, 724689006, 7052005, 231489001, 18573003, 63649001, 89618007, 33871004, 231438001, 191486006, 191667009, 31373002, 724674002, 60123008, 238975005, 762509000, 191672000, 473452003, 724719005, 191447007, 191563001, 51443000, 83746006, 278853003, 724756001, 129604005, 723936000, 64731001, 69322001, 50933003, 29599000, 71103003, 54761006, 63204009, 47447001, 371024007, 60401000119104, 63983005, 723900003, 724758000, 65971000052100, 268622001, 26714005, 238977002, 77475008, 737340007, 191570001, 191484009, 26472000, 32552001, 191525009, 191542003, 191547009, 724706006, 18260003, 238972008, 7761000119106, 63181006, 278506006, 38368003, 416340002, 238974009, 278852008, 35218008, 38295006, 191567000, 191571002, 10760421000119102, 4926007, 42868002, 735750005, 191499009, 55736003, 191670008, 191577003, 58214004, 724673008, 191678001, 762327001, 723913009, 231487004, 737225007, 724760003, 231449007, 19445006, 191555002, 278508007, 48500005, 403595006, 408858002, 20385005, 79204003, 39610001, 371026009, 35252006, 271428004, 724718002, 762326005, 191471000, 191526005, 237351003, 30491001, 79866005, 7025000, 238979004, 16990005, 191676002, 301643003, 26025008, 32358001, 191485005

#### 3.27 Pulmonary Circulation Disorders

ICD10CM:
I27.20, I27.23, I27, I28.0, I26.02, I27.1, I27.29, I27.81, I27.89, I26.90, I27.21, I27.22, I28.9, I27.82, I26.92, I26.93, I27.9, I26.99, I26.09, I26.94, I26.01, I27.0, I26.9, I27.83, I27.8, I28.8, I26.0, I27.2, I26, I27.24

ICD9CM:
415.1, 416.1, 417.0, 415.19, 415.13, 416.8, 417.8, 415.0, 415.11, 416, 416.2, 416.9, 415.12, 416.0, 417.9

SNOMED CT:
233944003, 450306008, 69357003, 286361000119101, 268187005, 448637008, 449513006, 703244000, 707372009, 697909006, 233947005, 447827003, 253321003, 253634009, 448087002, 253622001, 276794009, 461408000, 697898008, 447664007, 697914005, 2261009, 39785005, 95441000, 303070000, 55589000, 447571007, 697921005, 697899000, 609480009, 447274003, 697904001, 1010627004, 1163244009, 697906004, 253620009, 697907008, 697928004, 200304004, 446432002, 448078006, 449435001, 703243006, 111323005, 204457005, 233935004, 445543002, 448595006, 460307002, 1258896001, 27015006, 697915006, 15386007, 233941006, 271573009, 447813008, 448086006, 458088003, 697901009, 461433002, 472756005, 697912009, 49584005, 449493005, 472790001, 68237008, 93347003, 449085001, 471274000, 697910001, 703308008, 707371002, 460312001, 840307003, 11614003, 276792008, 445928005, 449444000, 707477002, 697916007, 697919000, 697927009, 67189007, 15964661000119102, 460380002, 703357006, 707478007, 36422005, 253627007, 447275002, 697923008, 253637002, 703258003, 10930001, 93059006, 88837005, 253304007, 713078005, 233939005, 253623006, 448084009, 448105006, 448487009, 448500006, 233942004, 762433009, 860680001, 871580000, 27986000, 58123006, 233949008, 449451009, 460596000, 253633003,

447914003, 74315008, 204448004, 233938002, 233946001, 253322005, 276793003, 472758006, 707437005, 200296007, 441557008, 450311005, 251042004, 79955004, 449495003, 449533005, 445208002, 735568006, 57834008, 253690003, 210790008, 707413005, 95838002, 448027004, 448728003, 253621008, 703246003, 17993000, 204456001, 200299000, 697924002, 1187633001, 37639005, 60366008, 438773007, 253625004, 253631001, 472757001, 5499009, 128566008, 133971000119108, 328511000119109, 445237003, 450308009, 233948000, 11399002, 233945002, 447668005, 36110001, 697920006, 253305008, 449452002, 1010625007, 447926008, 448599000, 703307003, 76846002, 200310004, 446668003, 449576007, 459165008, 233950008, 460595001, 253689007, 449467003, 66858001, 447903001, 707412000, 200311000, 111322000, 461434008, 776416004, 449514000, 697922003, 448059006, 204467000, 878858004, 86252004, 87394009, 128584005, 449350006, 10759311000119104, 52757001, 447939007, 49778009, 233936003, 449494004, 233937007, 95242009, 703355003, 89420002, 707668000, 200286003, 447860000, 448060001, 735567001, 77892009, 194892009, 253624000, 445296005, 789690008, 7368005, 449560003, 461407005, 460597009, 703245004, 22750001, 448120000, 45650007, 200288002, 233815004, 253636006, 267284008, 460387004, 879814007, 472702003, 697897003, 448727008, 447318008, 697905000, 840306007, 68092007, 445371009, 447528003, 697903007, 78862003, 447269006, 459164007, 285251000119101, 448783009, 472760008, 706870000, 51096002, 445106006, 449433008, 472759003, 447663001, 11029002, 697902002, 447832002, 448472004, 253628002, 371067004, 448510002, 460375006, 697917003, 1172565007, 280964006, 445209005, 447924006, 697908003, 707414004, 38905006, 82153002, 200301007, 233940007, 472791002, 879816009, 237342006, 200290001, 253632008, 360578005, 446890001, 449551006, 879815008, 82889007, 83291003, 723859005, 74218008, 445167000, 449547009, 472761007, 233954004, 88223008, 449434002, 87837008, 26780008, 447814002, 707476006, 720575002, 840308008, 286331000119109, 447498008, 870323006, 449120006, 450307004, 450309001, 251045002, 253691004, 234062003, 447840008, 707665002, 447861001, 449443006, 472768001, 11454006, 253630000, 200284000, 233943009, 697925001, 29934004, 697896007, 871578006, 449125001, 194883006, 111289009, 707667005, 59631007, 10964002, 448499002, 697926000, 703636009, 286071000119109, 95236005, 721238001, 253635005, 609458006, 697918008, 697900005, 32194006, 70995007, 449425007, 703385008, 460370001, 26146002, 697913004, 1001000119102, 200308001, 447925007, 448486000, 239297008, 253629005, 286341000119100, 39905002, 200303005, 447691009, 448079003, 251047005, 270512000, 460612009, 764452004, 697911002, 251046001, 450310006, 234161007, 472789005, 25559009, 39987008, 59282003, 253692006, 449559008, 460365008, 286351000119103, 15964701000119109, 448478000, 449521000, 200294005, 54682008, 253688004, 860681002, 360573001, 129582000, 200291002, 449589001, 253638007, 724208006

Truveta:
1215970

##### 3.28 Rheumatoid Arthritis

ICD10CM:
M05.319, M06.3, M08.841, M08.919, M05.659, M08.95, M08.959, M12.37, M05.46, M06.042, M05.37, M05.611, M06.249, M08.879, M08.26, M08.40, M08.851, M12.079, M35.08, M05.19, M05.23, M05.722, M06.359, M08.461, M08.93, M12.311, M33.92, M12.361, M05.7, M12.321, M08.03, M06.832, M08.21, M08.862, M05.511, M06.03, M35.07, M05.51, M06.011, M06.821, M08.0, M05.012, M05.342, M08.832, L94.3, M05.442, M05.661, M05.85, M08.931, M05.32, M05.60, M05.851, M06.2, M08.469, M08.922, M33.21, M46.84, M06.0A, M05.74, M05.811, M06.9, M12.021, M45.5, M05.132, M05.679, M05.71, M06.041, M08.23, M08.962, M08.80, M05.07, M12.349, M32.1, M05.55, M06.02, M06.269, M08.042, M08.3, M12.339, M46.8, M05.539, M12.052, M12.08, M33.09, M34.81, M35.0C, M05.242, M05.45,

M33.0, M45.9, M45.A6, M05.712, M06.25, M12.312, M12.342, M33.20, M05.059, M05.642, M05.751, M06.839, M06.869, M08.04, M08.08, M08.449, M06.24, M05.362, M06.231, M06.251, M08.041, M08.05, M05.052, M06.4, M06.861, M08.221, M08.459, M12.329, M32.11, M46.90, M05.472, M08.019, M12.319, M45.A4, M35.0A, M05.211, M06.339, M08.44, M06.08, M06.312, M12.371, M35.3, M08.94, M08.942, M32.10, M05.89, M08.09, M12.069, M33.90, M05.80, M05.57, M08.421, M45.3, M06.039, M06.22, M06.872, M08.98, M05.872, M34.89, M05.532, M35.00, M05.271, M05.541, M05.70, M06.369, M05.759, M30, M33.01, M05.512, M08.99, M05.179, M05.65, M05.672, M06.87, M12.332, M46.86, M05.542, M05.641, M06.262, M06.37, M08.412, M08.222, M12.362, M05.52, M05.76, M06.071, M06.342, M08.97, M30.3, M35.09, M05.019, M08.27, M33.13, M45.4, M05.31, M05.622, M08.471, M05.212, M05.221, M05.649, M06.23, M08.232, M34.0, M35.0B, M05.73, M06.259, M08.249, M34.82, M45.A3, M45.A5, M35.05, M05.332, M12.031, M05.04, M05.61, M06.01, M46.92, M05.071, M05.412, M08.43, M12.30, M05.129, M05.17, M05.359, M08.819, M12.019, M35.9, M05.142, M06.0, M45.A7, M05.479, M05.822, M08.251, M08.411, M35.4, M05.039, M05.149, M05.421, M05.771, M05.879, M08.242, M08.911, M08.921, M31.31, M05.371, M08.971, M12.322, M05.8A, M31.2, M46.93, M05.549, M05.571, M05.842, M12.359, M08.2A, M06.879, M08.821, M12.33, M12.372, M35.03, M46.94, M05.139, M05.529, M05.739, M05.869, M08.419, M05.562, M05.152, M05.171, M05.461, M05.719, M08.429, M08.939, M05.251, M05.349, M05.569, M05.662, M08.021, M08.071, M05.122, M05.329, M05.772, M08.48, M08.822, M12.351, M46.9, M05.072, M05.35, M06.85, M08.872, M05.131, M08.46, M35.01, M05.429, M05.612, M06.242, M06.361, M06.362, M08.01, M08.062, M46.88, M08.0A, M35.06, M05.341, M05.41, M06.871, M08.842, M12.34, M30.8, M34.9, M45.A, M05.331, M05.369, M05.44, M06.29, M08.241, M05.011, M05.4, M05.559, M05.579, M05.731, M06.371, M45.A0, M08.269, M12.0, M05.051, M05.422, M05.83, M08.00, M08.069, M08.07, M12.039, M31.10, M06.059, M06.311, M08.871, M05.079, M05.66, M05.862, M46.96, M05.112, M05.59, M06.271, M08.272, M12.071, M12.36, M46.95, M46.98, M05.312, M05.50, M06.229, M06.829, M05.119, M05.279, M05.77, M06.222, M06.341, M08.85, M08.932, M12.05, M05.39, M05.711, M06.8, M08.262, M05.27, M05.431, M05.54, M05.72, M05.779, M06.00, M45, M05.24, M05.321, M05.379, M06.1, M06.20, M05.859, M06.349, M08.039, M08.42, M12.341, M32.15, M08.25, M08.4, M08.472, M05.351, M05.452, M06.061, M35.5, M05, M05.269, M05.272, M05.3, M06.26, M12.31, M12.38, M32.13, M05.631, M05.84, M05.042, M05.849, M06.012, M08.059, M08.829, M12.059, M05.86, M08.949, M05.259, M06.351, M05.67, M05.752, M06.05, M08.02, M08.91, M08.28, M08.41, M08.422, M05.43, M05.471, M06.812, M08.239, M08.279, M05.05, M05.06, M05.56, M12.02, M33.11, M45.8, M05.36, M08.229, M46.81, M05.34, M06.211, M06.89, M46.87, M05.252, M05.651, M05.852, M06.07, M08.952, M32.8, M05.552, M08.92, M05.162, M05.26, M06.069, M06.822, M08.45, M08.961, M12.041, M30.0, M05.062, M05.621, M06.079, M06.851, M08.442, M08.452, M12.09, M05.03, M05.352, M06.322, M08.2, M08.8, M08.88, M33.9, M08.839, M33.22, M34, M05.121, M06.062, M31.30, M34.1, M05.9, M08.072, M12.00, M08.29, M05.239, M06.36, M34.83, M05.25, M05.761, M06.27, M06.862, M31.3, M33, M05.49, M33.00, M33.91, M06.241, M06.819, M12.072, M35.2, M08.252, M33.12, M35.89, M05.769, M06.052, M05.13, M05.372, M06.319, M06.842, M08.011, M08.079, M08.852, M45.2, M06.321, M06.83, M08.219, M08.96, M45.0, M45.A8, M05.029, M05.262, M05.832, M12.06, M31.1, M35.0, M05.1, M06.022, M06.232, M33.02, M05.222, M05.229, M06.019, M08.212, M08.441, M45.7, M05.459, M33.93, M05.7A, M06.8A, M05.871, M06.379, M30.1, M05.031, M05.249, M08.051, M46.99, M05.79, M08.06, M08.969, M05.339, M06.221, M06.852, M12.03, M12.35, M05.021, M12.331, M45.A1, M05.10, M06.34, M06.81, M08.9, M12.042, M05.64, M05.75, M05.821, M08.031, M08.061, M08.951, M05.111, M08.47, M08.869, M05.01, M06.252, L94.0, M05.049, M08.20, M08.84, M05.22, M08.859, M05.041, M35.7, M05.032, M08, M12.369, M05.632, M06.331, M06.372, M06.841, M06.86, M08.912, M05.069, M05.741, M05.841, M06.352, M08.052, M05.411, M05.819, M05.141, M05.322, M05.839,

M06.272, M08.451, M32.12, M32.14, M05.531, M05.62, M08.029, M08.211, M05.2, M05.21, M05.159, M05.232, M08.22, M08.431, M08.861, M12.07, M05.231, M05.449, M05.762, M08.83, M12.352, M31.11, M08.261, M08.462, M32, M05.20, M05.33, M05.8, M06.04, M12.051, M33.2, M05.829, M05.87, M08.032, M08.81, M05.0, M05.721, M06.279, M46.91, M05.419, M05.5, M06.021, M08.479, M06.239, M08.231, M08.89, M05.061, M05.729, M06, M06.09, M06.261, M08.022, M08.439, M05.361, M05.432, M06.029, M06.28, M31.0, M45.6, M05.11, M05.619, M05.82, M06.38, M35.8, M05.521, M06.811, M08.271, M08.812, M33.1, M46.82, L94.1, M06.21, M06.859, M08.811, M33.29, M46.83, M46.85, M05.572, M12.029, M12.3, M05.12, M05.169, M05.629, M06.031, M06.332, M06.831, M35.6, M05.15, M06.31, M06.35, M35.1, M35.81, M08.941, M05.151, M06.032, M06.329, M08.012, M08.972, M05.522, M08.90, M35.02, M05.53, M05.69, M12.01, M05.161, M05.639, M05.81, M06.051, M08.831, M45.A2, M05.439, M05.462, M05.6, M06.33, M08.86, M12.379, M35, M06.32, M05.732, M06.049, M06.072, M06.39, M06.82, M05.451, M06.212, M06.88, M08.049, M34.8, M35.04, M12.04, M12.062, M30.2, M05.241, M05.441, M05.671, M05.749, M12.012, M45.1, M06.06, M32.0, M05.02, M05.29, M12.032, M05.40, M06.849, M33.99, M12.32, M05.09, M05.47, M08.24, M32.9, M08.9A, M08.929, M12.011, M33.19, M46.89, M05.742, M05.812, M33.03, M05.561, M12.39, M05.519, M08.432, M33.10, M08.4A, M08.82, M05.022, M06.84, M32.19, M05.14, M05.172, M05.551, M05.669, M05.831, M12.049, M31.19, M05.00, M05.652, M46.1, M05.30, M06.30, M08.1, M12.022, M12.061, M05.261, M06.80, M05.16, M05.861, M08.259, M34.2, M46.97, M45.AB, M05.219, M05.311, M05.469, M08.87, M08.979, M05.63, M05.42, M06.219, M08.849, M46.80

ICD9CM:
714.32, 711.20, 719.37, 711.2, 720.89, 710.1, 446, 714.33, 725, 446.5, 446.20, 719.32, 711.22, 719.35, 714.0, 714.89, 714.30, 720.8, 714.1, 714.31, 719.31, 719.38, 711.21, 719.36, 719.34, 720.2, 710.3, 728.89, 711.29, 710.4, 714.3, 446.2, 711.27, 719.3, 446.1, 720.1, 711.28, 446.7, 710.9, 446.3, 719.33, 711.25, 711.24, 711.23, 446.6, 714.4, 719.30, 720, 714.9, 710.0, 714.2, 719.39, 446.21, 446.0, 446.29, 710.8, 701.0, 710.2, 720.81, 714, 446.4, 714.8, 720.9, 729.30, 711.26, 728.5, 714.81, 720.0

SNOMED CT:
1073801000119100, 15690361000119104, 1144912000, 52661003, 15691801000119104, 1144931006, 201768005, 319151000119103, 201805000, 423486005, 239886003, 38877003, 201776007, 104681000119106, 156370009, 239792003, 307261000119104, 128461001, 201781003, 15673361000119100, 236502006, 82275008, 4676006, 82939003, 86261000119104, 1073751000119106, 236744002, 402428008, 439007008, 1073391000119100, 57160007, 16050071000119108, 319091000119107, 441322009, 306561000119100, 15691161000119108, 1162331006, 239890001, 195353004, 239808004, 306551000119102, 15691321000119101, 300961000119108, 1264001009, 15687321000119109, 1212005, 62382002, 239805001, 427770001, 786077003, 239812005, 16426601000119103, 319071000119106, 201436003, 15691721000119102, 196138005, 735600005, 202744000, 397856003, 201777003, 238936001, 267971005, 1073761000119108, 16044751000119106, 724606001, 201775006, 698694005, 1156779003, 239803008, 1162355001, 126729006, 1156774008, 310501000119101, 239943002, 78946008, 402339007, 143441000119108, 1162330007, 1231671001, 239800006, 239927009, 459911000124100, 201771002, 402433007, 201767000, 201783000, 398640008, 1073701000119107, 19682006, 202666001, 16606681000119101, 11013005, 201772009, 402434001, 1073681000119109, 201770001, 1144977002, 238806009, 281358000, 305961000119100, 196133001, 11055151000119108, 15686281000119101, 287007001, 410802003, 1073611000119103, 1156777001, 318961000119108, 89681000119101, 319081000119109, 295111000119108, 373420004, 319121000119106, 128460000, 201780002, 239930002, 287006005, 319131000119109, 1222679006, 65323003, 77522006, 304301000119101, 319841000119107, 239912009, 318861000119101, 1073741000119109, 428218004,

1073821000119109, 202667005, 86219005, 232460001, 15685961000119107, 73286009, 724502006, 319011000119103, 715401008, 302935008, 410801005, 1162292003, 239815007, 1149218009, 55464009, 201736002, 1073691000119107, 74391003, 1073731000119100, 1074971000119103, 15673521000119101, 10713006, 239799007, 1144920003, 429298008, 784332006, 201789001, 724782005, 236503001, 15691881000119107, 1162364006, 54867000, 201774005, 712547007, 15687201000119107, 724767000, 310561000119100, 402432002, 299276009, 15630971000119102, 310521000119105, 402712002, 1073601000119101, 2304001, 721272009, 295101000119105, 15686001000119104, 50442003, 202700003, 410799007, 318901000119107, 410800006, 431236003, 126766000, 1153391002, 15691241000119101, 15692001000119101, 301051000119101, 398726004, 319031000119108, 235000001, 239935007, 287008006, 402425006, 712548002, 201807008, 15691081000119100, 1073791000119101, 201048007, 318881000119105, 1153372002, 25380002, 201784006, 201791009, 309762007, 15691121000119103, 202457009, 400054000, 712524006, 1162677006, 1156778006, 33719002, 774082004, 459921000124108, 403488004, 444556003, 1073721000119103, 87442008, 427967009, 318871000119107, 196137000, 128458002, 239802003, 319061000119100, 51156002, 403487009, 15687841000119108, 202752002, 15972981000119101, 318891000119108, 15686321000119106, 81573002, 735599007, 239798004, 129563009, 201773004, 239804002, 239811003, 239925001, 403635000, 202748002, 239887007, 201568008, 201796004, 1153369009, 201443009, 239928004, 59165007, 319051000119102, 870281008, 310081000119100, 318931000119100, 89155008, 162930007, 720363008, 68815009, 89030000, 15691921000119100, 201769002, 403521006, 69896004, 295121000119101, 76521009, 723116002, 9350004, 306821000119108, 1073771000119102, 1144925008, 201778008, 724603009, 239810002, 202465007, 318851000119103, 403522004, 15691761000119107, 1153410003, 318911000119105, 200956002, 318971000119102, 22784002, 60555002, 281371002, 410502007, 58144006, 1153411004, 239806000, 396230008, 1149219001, 1156819003, 371082009, 399923009, 387801000, 402427003, 443872005, 773333003, 319941000119103, 319141000119100, 1162319009, 28880005, 1162362005, 95644001, 429555005, 232369001, 308143008, 402338004, 58961005, 77753005, 110041000119104, 318951000119106, 1156766007, 201764007, 239938009, 398049005, 83901003, 239915006, 201051000, 202461003, 239797009, 1073381000119103, 78129009, 202756004, 402426007, 31848007, 1144964006, 1033009, 239809007, 429192004, 699302001, 783711009, 319871000119100, 15691961000119105, 55146009, 1073781000119104, 239889005, 298951000119104, 310511000119103, 310571000119106, 712549005, 15685921000119102, 781206002, 318841000119100, 7607008, 239926000, 298285004, 444133002, 1073831000119107, 52042003, 16426641000119101, 239791005, 1073711000119105, 16024431000119108, 410796000, 319111000119104, 1156765006, 304311000119103, 311671000119100, 310491000119108, 701003, 67224007, 15691841000119102, 319041000119104, 1162313005, 410798004, 1156817001, 201779000, 202668000, 239795001, 774080007, 319861000119106, 36402006, 239807009, 239796000, 239901009, 239929007, 155441006, 201766009, 403486000, 438476003, 15692041000119104, 238935002, 713777005, 718217000, 724781003, 1074981000119100, 1073811000119102, 410797009, 16426681000119106, 239936008, 54072008, 201785007, 84801008, 310551000119102, 58474006, 402431009, 318941000119109, 319951000119101, 235066005, 86365006, 318921000119103, 1162303002, 202463000, 239888002, 116441000119107, 9631008, 1162336001, 202460002, 762303003, 16606721000119107

##### 3.29 Solid Tumor without Metastasis

ICD10CM:
C50, C50.311, C72.21, C69.12, C10.0, C14.0, C72.40, C69.41, C57.01, C22.4, C25.4, C40.01, C47.1, C49, C16.8, C18.0, C40.02, C50.921, C72.30, C76.52, C25.8, C41.4, C38.1, C69.01, C11.1, C38.2, C43.39, C49.1, C50.021, C76.3, C50.522, C43.72, C43.5, C50.61, C50.621, C60.8, C68.1, C62.02, C34.8, C51.9, C75.1, C25, C26, C40.22, C76, C57.00, C69.02, C75, C10.4, C21.2, C43.4, C26.9, C45.1, C50.212, C62.01, C46.4, C50.419, C50.81, C50.91, C75.2, C18.4, C18.8, C50.019, C50.511,

C52, C74.9, C67.2, C69.21, C22.2, C43.62, C46.2, C50.62, C34.02, C40.11, C43.12, C63.01, C23, C40, C60.0, C64.9, C74.10, C34.80, C50.4, C50.52, C56, C40.81, C49.0, C72.20, C48.0, C50.122, C41, C41.1, C45.7, C49.8, C76.7, C69.50, C21.8, C50.119, C63.1, C18.5, C21, C50.912, C63.00, C67.6, C75.5, C40.3, C48.1, C50.629, C34.01, C40.9, C75.9, C46, C55, C71.5, C17.9, C34.10, C50.819, C63.9, C67.7, C46.5, C46.9, C49.3, C69.9, C71, C50.21, C71.3, C30.1, C40.0, C69.20, C72.59, C15.8, C32, C46.50, C40.12, C50.42, C48, C50.811, C66.0, C16.5, C69.4, C10.8, C43.2, C50.219, C45.2, C50.92, C72.3, C72.41, C31.3, C62.91, C64.0, C76.40, C63.8, C69.92, C50.121, C62, C74.12, C75.4, C18.3, C34.30, C68.0, C43.7, C74.1, C40.80, C50.221, C43.112, C50.12, C50.422, C67.1, C68.8, C57.22, C69.00, C69.1, C20, C43.111, C47.22, C76.0, C46.1, C56.0, C49.A4, C70.9, C34.81, C63, C62.9, C66.1, C32.1, C56.2, C69.52, C71.6, C22.7, C31, C31.9, C40.91, C50.211, C63.2, C50.0, C50.519, C54.9, C24.1, C50.5, C67.9, C69, C50.11, C72.9, C65.0, C47.9, C22.1, C34.0, C56.9, C69.32, C34.90, C40.1, C43.61, C50.611, C71.0, C13.1, C69.11, C74.01, C50.312, C10, C47.6, C50.82, C57.02, C57.9, C61, C62.10, C63.12, C16.6, C69.42, C69.61, C17, C50.929, C76.8, C49.A1, C50.51, C72.50, C47.10, C51, C69.10, C50.029, C24, C43.8, C46.3, C50.529, C21.0, C22, C34.11, C40.92, C32.0, C74.90, C18.7, C43.59, C43.71, C16.4, C34.00, C50.512, C50.612, C76.51, C26.0, C58, C69.62, C72.31, C57.4, C75.8, C46.0, C65.1, C70, C15.5, C47.11, C30.0, C50.829, C13.8, C40.20, C50.012, C54.1, C57.2, C10.3, C16, C18.6, C43.122, C50.521, C24.8, C39.9, C43, C67.8, C69.31, C72.32, C49.A5, C50.412, C30, C57.11, C63.02, C31.1, C43.1, C43.20, C49.22, C69.91, C70.0, C13, C43.60, C72.1, C74.02, C34.92, C67.0, C31.2, C34.2, C50.01, C74.0, C40.21, C49.9, C50.02, C50.022, C66.2, C14.8, C40.82, C46.52, C50.129, C17.0, C50.429, C15, C47.12, C50.111, C69.5, C72, C74.00, C16.0, C17.8, C56.1, C64, C16.9, C69.82, C75.0, C41.2, C53.9, C12, C43.30, C60.9, C72.22, C10.9, C32.3, C33, C69.90, C76.50, C22.0, C32.9, C39, C41.0, C43.0, C50.919, C69.0, C17.3, C57.0, C63.10, C11.9, C38.8, C40.2, C51.1, C74.11, C76.1, C24.9, C40.32, C43.10, C47.4, C54.0, C47.0, C50.329, C50.6, C50.619, C50.9, C57.21, C63.11, C72.2, C47.2, C62.11, C26.1, C68.9, C32.2, C49.A0, C49.A9, C65, C18.9, C34.31, C47.3, C50.922, C57.8, C34.3, C57.1, C16.2, C25.7, C13.2, C50.421, C53.8, C34.91, C49.2, C50.1, C67.4, C48.2, C74, C76.41, C18.2, C69.8, C34.32, C11.8, C49.6, C50.31, C50.821, C69.30, C71.1, C15.3, C38.4, C43.21, C53.0, C49.4, C49.A2, C50.2, C50.41, C62.12, C15.4, C34.12, C49.12, C50.319, C50.911, C54.8, C67.3, C43.51, C50.011, C47.20, C50.222, C51.2, C41.3, C67, C69.3, C49.A, C40.10, C50.622, C53.1, C11, C18, C24.0, C63.0, C75.3, C15.9, C34.1, C46.7, C71.4, C53, C57, C69.51, C54.2, C43.22, C43.31, C39.0, C71.2, C13.0, C25.0, C31.8, C34, C50.3, C60.2, C62.90, C22.3, C49.11, C60.1, C18.1, C40.31, C46.51, C50.321, C65.2, C69.6, C72.4, C49.10, C76.4, C25.1, C38.0, C67.5, C73, C74.91, C43.121, C63.7, C17.2, C25.2, C56.3, C14, C40.30, C72.42, C10.1, C14.2, C11.2, C45.0, C50.229, C11.0, C50.812, C57.10, C57.3, C69.22, C74.92, C38.3, C41.9, C64.1, C69.2, C10.2, C43.3, C50.22, C57.7, C76.2, C76.5, C49.20, C50.112, C51.8, C54.3, C47, C48.8, C50.322, C54, C66, C38, C40.90, C45.9, C19, C32.8, C64.2, C21.1, C16.1, C22.9, C31.0, C43.52, C45, C22.8, C49.A3, C71.7, C62.92, C37, C57.12, C62.00, C50.32, C69.80, C43.6, C69.81, C76.42, C13.9, C40.8, C68, C69.40, C17.1, C34.9, C11.3, C25.9, C47.8, C51.0, C69.60, C72.5, C25.3, C49.5, C50.411, C65.9, C72.0, C43.70, C47.21, C50.8, C60, C16.3, C50.822, C47.5, C49.21, C57.20, C62.1, C66.9, C70.1, C34.82, C40.00, C43.9, C62.0, C71.8, C71.9, C43.11

ICD9CM:
156.8, 164, 143, 145.5, 159.8, 147, 151.1, 161.3, 149, 161.2, 143.1, 160.9, 170.3, 147.8, 151.0, 171, 145.9, 146.5, 150, 140.1, 154.0, 157.2, 146.2, 163.9, 141.3, 170.1, 146.9, 163, 159.0, 162.8, 170.7, 160, 161, 151.6, 140.3, 158.8, 146.0, 156.2, 172.1, 172.5, 141.6, 172.4, 140.8, 144.1, 162.4, 161.8, 140.5, 148.8, 150.9, 163.8, 155, 141.0, 152, 153.0, 151.4, 158.9, 171.6, 172.7, 142.1, 151.3, 145, 172.2, 154.3,

145.8, 150.0, 153.9, 142.8, 153.5, 156, 170.8, 170.9, 145.0, 146.8, 147.2, 152.1, 147.0, 162.2, 157.3, 160.3, 159, 149.1, 158, 146.4, 148.2, 160.0, 172, 159.1, 165.8, 170.5, 154.8, 160.1, 172.8, 146, 171.7, 148, 162.9, 172.6, 150.8, 160.5, 141.1, 141.2, 165, 141.5, 146.6, 162.3, 150.5, 171.2, 145.4, 153.3, 153, 164.2, 152.9, 156.0, 170, 152.0, 162, 162.5, 148.1, 150.1, 156.9, 172.3, 141.9, 150.4, 153.8, 157.8, 144.9, 157.9, 157.4, 143.0, 146.1, 151.2, 151.5, 172.9, 140.4, 142.2, 143.9, 149.9, 146.7, 152.2, 171.5, 153.7, 154, 145.3, 161.1, 147.3, 170.4, 148.0, 152.3, 160.4, 164.0, 164.3, 144, 141, 141.8, 171.4, 144.8, 164.9, 170.0, 163.0, 148.3, 159.9, 165.0, 171.8, 153.1, 142, 142.0, 160.2, 170.6, 160.8, 148.9, 155.1, 147.9, 172.0, 140.0, 150.3, 163.1, 145.1, 145.6, 161.0, 171.9, 162.0, 157.0, 157, 143.8, 153.4, 140, 152.8, 155.0, 151.8, 155.2, 165.9, 171.0, 144.0, 151.9, 156.1, 145.2, 150.2, 157.1, 171.3, 153.2, 146.3, 149.0, 141.4, 147.1, 151, 170.2, 142.9, 154.1, 158.0, 164.8, 140.9, 149.8, 154.2, 140.6, 153.6, 161.9, 164.1

SNOMED CT:
94002000, 94069006, 94077005, 94946006, 109882003, 126513000, 188073001, 188159008, 254766004, 254987003, 371969003, 400010006, 448372003, 448674007, 707383000, 707429002, 716855006, 722687007, 833285007, 87151000119105, 681601000119101, 16583001000119104, 69408002, 109385007, 414780005, 352751000119103, 689441000119102, 1196876009, 1255190008, 1255898004, 1259406007, 1259446001, 1259502006, 1259512004, 1259590008, 1259637001, 1260094000, 1268393000, 1268485001, 91998009, 187947006, 255031003, 255037004, 427038005, 446939001, 449386007, 722444002, 764855007, 1652721000119108, 16906561000119108, 1259367002, 1259653003, 1254727003, 1268532006, 1268638003, 1268955002, 1284829001, 92350005, 92355000, 93911008, 94450005, 94571002, 94572009, 94869005, 109370001, 126570005, 189406008, 254443004, 254716005, 302837001, 400092004, 403899005, 426336007, 447886005, 449219006, 699357004, 716653001, 721579004, 763131005, 771080008, 785879009, 816205008, 838346000, 840502005, 1080341000119105, 15635721000119108, 15959061000119101, 94719007, 402911002, 1255808008, 1259477004, 1259711008, 8090002, 92097004, 92101008, 92362009, 92393005, 94129007, 94627008, 126782004, 188502002, 188666005, 188780006, 189399009, 235001002, 275396004, 277617004, 371968006, 449166001, 699852000, 721644006, 722445001, 767444009, 773775004, 99111000119103, 323691000119105, 460231000124103, 10708511000119108, 16909081000119101, 1157159009, 682041000119100, 350781000119106, 1259449008, 1260045000, 1263654002, 93141006, 93160004, 93689003, 93865007, 93930008, 94000008, 94294002, 94518002, 94639000, 94972002, 95711003, 126759001, 126971002, 187708004, 188010006, 254659009, 254733003, 255081007, 372005006, 393563007, 400099008, 403958000, 403978008, 404141000, 444869007, 448317000, 448865007, 704252008, 707392002, 733346002, 130371000119101, 184861000119102, 348711000119100, 93543003, 1079581000119107, 1081361000119102, 681691000119107, 685981000119100, 1255652003, 1259314002, 1268387008, 53633000, 91554004, 92111001, 93872008, 94087009, 94230009, 95005005, 109264009, 109876001, 126563005, 126564004, 202442002, 242862004, 254840009, 277549009, 400118002, 419738008, 442537007, 448666006, 707627009, 766046007, 870318006, 113241000119104, 109962001, 685571000119105, 690141000119103, 1162282006, 1255809000, 1259390007, 1259721000, 1260077002, 31047003, 93638003, 93803002, 94041003, 94539009, 94781000, 94948007, 95174008, 109270003, 109887009, 126575000, 188060000, 188321006, 188674006, 188805008, 254713002, 254818000, 255096006, 276739001, 302848008, 314976006, 372111007, 402553008, 403918002, 405546008, 713483007, 735916009, 767812003, 291911000119107, 11307961000119106, 1079881000119105, 1153347002, 683241000119101, 689251000119101, 1197276007, 1080901000119107, 1259695007, 1236943009, 1237467000, 1268539002, 92283000, 92361002, 94001007, 94949004, 94987007, 126741004, 188051008, 188507008, 188976006, 202581004, 254506004, 255029007, 271449003, 402499008, 404094007, 423158009, 448267005, 448435005, 449803009, 703387000, 1079371000119103, 11305561000119109, 16905141000119107, 685951000119107, 1255112004, 1255656000, 1259372006, 1259420000, 1259530006, 1259583003, 1268502003, 92713007,

92751007, 93133006, 93645003, 93798006, 93993007, 94082003, 94122003, 94191001, 94443006, 94453007, 95015004, 95148002, 188857008, 254401006, 307601000, 400122007, 403878007, 698043002, 721568009, 722677008, 739301006, 203047009, 16782691000119109, 1254844005, 1259370003, 1259410005, 1259472005, 1260057009, 1268459000, 92000007, 92171007, 93744007, 93991009, 94016005, 94465006, 95104007, 109821008, 109832008, 126794008, 187989003, 188648000, 189815007, 254805008, 254982009, 372112000, 449637003, 707673006, 721637002, 722666002, 763477007, 1079861000119101, 16908561000119107, 1079071000119101, 16636101000119105, 687841000119105, 1222705009, 1259594004, 1260071001, 1268701008, 93146001, 93667002, 94264008, 94342002, 94454001, 94567000, 94716000, 94840001, 94915007, 127002001, 187972006, 188091006, 188133005, 188781005, 202575007, 239096008, 254811006, 255017003, 274902006, 277626001, 402529002, 403946000, 404037002, 413441006, 423424005, 447991007, 707388009, 707674000, 785807007, 788972003, 87121000119102, 293801000119103, 15956181000119102, 16091171000119106, 110005000, 203051006, 897649003, 684931000119100, 685941000119105, 1187305006, 1196843005, 1255798008, 1259744002, 1259785007, 1263460007, 8214000, 76813008, 91860005, 93814004, 93927001, 93944002, 94022001, 94170009, 95050007, 95072009, 126507009, 187613005, 187622006, 187926007, 188256008, 189001006, 189084001, 254764001, 254938000, 312935003, 371992003, 372108006, 403964007, 403979000, 404161006, 422833009, 449632009, 721605003, 762458004, 770402000, 130891000119106, 93890009, 404135007, 1156416002, 1156460002, 683041000119105, 1179377000, 1197268005, 1259819000, 1260070000, 1260074009, 34739009, 91986004, 92335001, 92344000, 95081003, 109282007, 126521006, 126610006, 238528004, 254632001, 254715009, 274087000, 285769009, 369568004, 371134001, 371978009, 402520003, 403714009, 417264005, 423673009, 449578008, 698045009, 707391009, 733470002, 460271000124100, 1080551000119100, 1162354002, 1254805002, 1260063000, 91855006, 91997004, 92425006, 94011000, 95214007, 109940001, 126488004, 187942000, 188122007, 188135003, 369557004, 399990001, 404105000, 722515001, 734522002, 2961000119106, 113121000119101, 1082901000112103, 93541001, 109970006, 359631009, 1078981000119108, 1080581000119107, 15955141000119104, 1254803009, 91854005, 92297008, 92386006, 92400002, 93652001, 93925009, 93957005, 94015009, 94107008, 94343007, 94415002, 94990001, 95041005, 126602005, 187601000, 188963000, 238863004, 255109008, 303055001, 423463003, 707581004, 721302006, 721576006, 774066000, 783704005, 789721005, 350761000119102, 351661000119103, 682031000119109, 683421000119106, 1259376009, 1260039008, 91975001, 92723003, 93473009, 93787005, 94043000, 94115006, 94969009, 94970005, 95128009, 110986000, 126655004, 126975006, 126983000, 188506004, 254650008, 254990009, 255070009, 255072001, 314420008, 404117003, 404170009, 416921001, 703228009, 721545000, 765136002, 133531000119104, 460041000124103, 92212001, 1156795003, 16527601000119105, 352391000119109, 689791000119102, 1196911006, 1255795006, 1255796007, 1236954004, 92144007, 92719006, 94323001, 94395004, 94674004, 187633009, 188635004, 402873007, 403889000, 416351002, 721716004, 723845003, 1079271000119106, 93159009, 188568000, 685701000119105, 939595491000119108, 1163357003, 1186631005, 352551000119106, 1260040005, 1237487004, 1268526001, 92341008, 92391007, 92457007, 93788000, 94728008, 95058000, 109926002, 109938006, 126597003, 202448003, 254404003, 254700008, 277942005, 286894008, 371966005, 402509003, 403873003, 404056007, 413843002, 448931007, 699356008, 90831000119105, 354031000119102, 109966003, 123731000119106, 39295002, 92122002, 93775003, 95226002, 109837002, 188631008, 402503002, 402522006, 423295000, 449034009, 723182009, 765095002, 822969007, 840424008, 107601000119107, 348261000119107, 460021000124105, 1078881000119102, 1082281000119105, 11312881000119107, 1081291000119104, 1082241000119100, 15951701000119100, 1259794001, 91999001, 92322005, 92474007, 92725005, 92736003, 93816002, 94014008, 94097000, 94932006, 95037006, 188017009, 202451005, 254874008, 277185000, 404008001, 424078005, 449248000, 707397008, 707590006, 713609000, 721577002, 1681000119102, 11312801000119104, 16301091000119107, 1080561000119103, 1186615002, 1197315002, 1196856003, 1260018001, 1260091008, 1237520001, 1251466008, 92200000, 93757004, 94581003, 94780004, 94961007,

126496009, 126503008, 126532006, 126739000, 188634000, 189308005, 254408000, 254428001, 254442009, 254797000, 314408000, 369448007, 400132000, 423195009, 441313008, 447100004, 715904005, 718220008, 721561003, 721672004, 735678002, 782697005, 866072004, 101621000119105, 139671000119100, 15635761000119103, 16091411000119109, 1197332003, 1255799000, 1259401002, 1259662001, 1259697004, 21848000, 93687001, 93701008, 93891008, 94640003, 126611005, 126780007, 127031002, 201161009, 254717001, 254765000, 254786000, 254886006, 255022003, 277569004, 363477002, 402497005, 404113004, 415174001, 423468007, 448867004, 704244005, 724805000, 683731000119104, 11996441000119108, 13351431000119102, 16594881000119101, 895358004, 15958021000119103, 687331000119109, 1254906000, 1259330005, 1259532003, 1259563004, 1259693000, 1260115001, 1251495004, 39843000, 92228006, 94154007, 94595000, 109275008, 109292004, 127001008, 188503007, 189138008, 254883003, 369572000, 373082000, 402500004, 403205000, 403929003, 448146001, 448354009, 449630001, 707407001, 715664005, 764735002, 1651000119109, 67821000119109, 681621000119105, 16538831000119103, 1196854000, 1236957006, 1269044006, 92354001, 93190006, 93809003, 93996004, 94120006, 94244003, 94473002, 94486006, 94658009, 94797005, 94875001, 126652001, 187927003, 187945003, 188065005, 188269007, 202583001, 255186004, 403965008, 447712006, 703294004, 721580001, 722385008, 762396004, 792907004, 116711000119103, 681131000119101, 890528009, 1081351000119104, 16068091000119101, 685641000119102, 1255725002, 1259491005, 92383003, 93161000, 93714009, 94112009, 94540006, 94762008, 95069002, 109271004, 126566002, 126622009, 134335004, 187681002, 253001006, 254414007, 276751004, 277574007, 304990002, 363365008, 372014001, 373081007, 424454009, 449065000, 698646006, 707470000, 713574000, 721544001, 722513008, 68261000119102, 16347601000119109, 188029000, 1010624006, 897554004, 1079011000119109, 1079051000119105, 1148891004, 1156469001, 1254825003, 1259672003, 1263911005, 92631001, 92709000, 93145002, 93684008, 93842002, 94035005, 94463004, 94943003, 95186006, 109831001, 187864008, 188121000, 188180002, 254631008, 363496003, 707408006, 823017009, 61301000119102, 96901000119105, 107791000119107, 1079281000119109, 11309441000119106, 93496009, 1082181000119103, 689851000119102, 1186614003, 1260059007, 31704005, 93760006, 94195005, 94478006, 94860009, 109287001, 126520007, 233719000, 254392009, 254405002, 254637007, 254984005, 278433008, 314975005, 372105009, 402526009, 404005003, 12311000132101, 107771000119106, 16314711000119100, 1010681001, 1078971000119105, 1156066000, 1156410008, 16774591000119105, 688061000119104, 1196998001, 1197285007, 1259435004, 1259813004, 1260032004, 92235003, 92287004, 93650009, 93776002, 94493005, 94950004, 188033007, 200721005, 269515006, 275491008, 402868001, 404154007, 404156009, 448371005, 449628003, 713189001, 764952009, 122961000119105, 350871000119100, 353441000119103, 683441000119100, 1080981000119104, 404157000, 1081721000119106, 15955941000119102, 15958101000119106, 16782241000119104, 1196905008, 1259486009, 1259830002, 1254735000, 1268486000, 1268538005, 91856007, 92266009, 92321003, 92388007, 92397006, 93958000, 94494004, 94812006, 109290007, 109991003, 110006004, 126526001, 127255004, 187862007, 188103003, 188943007, 188947008, 189324006, 254732008, 255074000, 369536007, 402555001, 403915004, 403933005, 404137004, 707359008, 721700001, 733162002, 786894002, 818967003, 117211000119105, 54411001, 188512009, 188672005, 1010656004, 16908601000119107, 687421000119101, 688081000119108, 1259340008, 1259580000, 1259643004, 1259700000, 1268534007, 1269123008, 92268005, 93200005, 93468003, 93655004, 93862005, 93912001, 94113004, 94255003, 94809008, 95099006, 95169000, 109992005, 187700006, 254455005, 254669003, 371984007, 372017008, 400062008, 403199007, 403966009, 709204000, 840514002, 107581000119103, 117091000119108, 1156404008, 1156806002, 16907791000119106, 351951000119107, 686461000119108, 1196831009, 1259829007, 1268716006, 91965004, 93770008, 94098005, 94327000, 94344001, 94873008, 95187002, 126984006, 187969004, 188862009, 254625005, 254792006, 276728004, 277627005, 312104005, 363383000, 363421004, 370511006, 402946005, 403924008, 403995000, 423793008, 448730001, 724467001, 830029005, 458321000124102, 460071000124106, 1081021000119109, 1081561000119108, 16905581000119107,

687971000119108, 689831000119108, 1255661003, 1259528009, 1260072008, 1237461004, 1254729000, 1268540000, 93692004, 93772000, 93945001, 94231008, 109276009, 109348004, 126784003, 188000002, 313250007, 363227003, 363441009, 373168002, 403200005, 404087009, 448774004, 698766008, 713327005, 734015000, 762457009, 822970008, 107591000119100, 1079101000119105, 351251000119103, 1259660009, 1254731009, 1268545005, 56975005, 93801000, 93837005, 93878007, 93917007, 94084002, 94625000, 94631002, 94877009, 95034004, 95062006, 188492005, 188883003, 246881005, 254409008, 254453003, 269533000, 307596009, 403940006, 404091004, 404118008, 707349007, 707421004, 707452003, 716652006, 763309005, 4521000119105, 188513004, 1010603001, 896745008, 1144252005, 1156409003, 683761000119107, 689371000119102, 1196900003, 1259391006, 1259745001, 1268352008, 1268905005, 62557001, 92399009, 93228004, 93998003, 94110001, 94182000, 94560003, 94635006, 95095000, 109367000, 126500006, 126584000, 126981003, 187608006, 188062008, 189155007, 269581007, 276962007, 403913006, 403930008, 449309003, 707377003, 707472008, 789689004, 107571000119101, 1076961000119100, 188589009, 1080531000119106, 350751000119104, 1217020002, 1259423003, 1251492001, 92401003, 92712002, 93229007, 93665005, 94024000, 94194009, 94273000, 94367004, 94548004, 94594001, 94675003, 94800007, 94980009, 109870007, 109931000, 126746009, 127003006, 237064007, 254691005, 276874005, 363461003, 371962007, 403874009, 403972009, 423284006, 733834006, 737312002, 783183009, 91181000119105, 113181000119102, 116381000119105, 127961000119108, 1080111000119108, 203045001, 254727007, 898197001, 350681000119101, 352481000119103, 16764101000119100, 1255813007, 1259332002, 1259374007, 1259645006, 1259716003, 1268639006, 1268720005, 92281003, 94003005, 94131003, 94204005, 94285008, 94590005, 94690004, 94723004, 95177001, 126807001, 253021007, 254551004, 286887005, 302816009, 363351006, 369539000, 372096000, 400138001, 404071006, 404127009, 404169008, 707457009, 721698005, 722678003, 766758001, 766979005, 71111000119109, 681651000119102, 880093002, 1081591000119101, 1156452003, 1156810004, 1197343006, 1259455003, 1259742003, 1260019009, 92790006, 94067008, 94140004, 94551006, 94722009, 187865009, 187906008, 188325002, 254426002, 254708001, 254986007, 255086002, 280959007, 303017006, 371975007, 404019003, 446241005, 707390005, 1078931000119107, 1080701000119109, 15956701000119109, 16637101000119107, 1229871006, 1259357007, 1259523000, 1259543006, 1236968006, 1251489000, 1254732002, 1268705004, 1268710000, 1268904009, 91996008, 94265009, 109852009, 123313007, 234140000, 254424004, 277572006, 363369002, 403485001, 403934004, 707342003, 713897006, 733362008, 138011000119105, 445269007, 1080051000119105, 1080721000119100, 15977551000119100, 352331000119105, 352701000119102, 688011000119102, 1259733005, 1259753009, 1260083004, 1269413009, 92108002, 92762007, 93653006, 93796005, 93947009, 93974005, 94046008, 94142007, 94637003, 94718004, 95021000, 109272006, 126874009, 254798005, 276799004, 281702006, 371983001, 394726009, 404121005, 420788006, 422572002, 428100006, 449268006, 721607006, 724554001, 1010679003, 895345000, 897646005, 10976631000119101, 352381000119106, 1187197004, 1259798003, 1237528008, 92365006, 92749008, 93771007, 93892001, 94051002, 94296000, 94545001, 94553009, 95038001, 95053009, 95079000, 126571009, 126653006, 126747000, 164420007, 188795008, 202453008, 254663002, 255020006, 276860003, 302841002, 307592006, 369522002, 372025005, 403826009, 403916003, 404055006, 446076000, 448221005, 449217008, 449318001, 707426009, 721311006, 722685004, 788874003, 79281000119106, 91061000119100, 113171000119100, 11304881000119103, 12238731000119100, 93522004, 1010607000, 1082271000119107, 684901000119107, 1162351005, 1259381000, 1259684001, 78572006, 92566008, 93222003, 93846004, 93916003, 93971002, 94364006, 95091009, 126869001, 188030005, 188064009, 188110009, 254654004, 373090000, 403941005, 404069006, 404144008, 423535002, 427685000, 445305001, 448220006, 448448009, 707596000, 721718003, 354671000119106, 93514006, 93523009, 110007008, 203054003, 1010674008, 879956005, 1148904003, 681751000119106, 352581000119104, 1259382007, 1268715005, 92026002, 92366007, 92781004, 93710000, 93812000, 94052009, 94297009, 95139007, 109879008, 126515007, 126601003, 126630005, 187658004, 190818004, 254437001, 254498009,

254808005, 278491007, 363366009, 414250001, 702476004, 707431006, 710209009, 733356003, 764940002, 836274002, 866070007, 1701000119104, 36831000119103, 107691000119101, 457721000124104, 683401000119102, 1079301000119108, 1079361000119109, 1080211000119103, 1082311000119107, 15710001000119105, 110004001, 302261000119101, 1080071000119101, 1157062009, 682231000119100, 145298771000119104, 15951421000119100, 1259434000, 45061000087102, 93871001, 94156009, 94192008, 95060003, 95070001, 95141008, 109996008, 126524003, 187604008, 187631006, 187948001, 188020001, 202452003, 254799002, 276954004, 281498002, 281565009, 363386008, 397011009, 398670003, 403922007, 404149003, 423325007, 444712000, 447710003, 707423001, 724058006, 724060008, 133881000119100, 351961000119109, 1079321000119104, 1079391000119102, 423829008, 1156417006, 682541000119106, 302251000119103, 1268962006, 92744003, 93216007, 93899005, 94458003, 94559008, 94591009, 94648005, 94688000, 109296001, 126508004, 187992004, 255184001, 276975007, 363385007, 371996000, 372121004, 403876006, 413842007, 418372008, 448983000, 449292003, 723301009, 725390002, 733345003, 121951000119101, 1080911000119105, 11305921000119109, 187752007, 255114007, 1079701000119109, 1237578005, 1259705005, 1259759008, 1259799006, 1260110006, 16705281000119104, 36025004, 92363004, 92717008, 93763008, 93956001, 94058008, 94102002, 94619004, 95193005, 95264000, 109293009, 109848009, 109925003, 126772000, 187697007, 187976009, 188718006, 189814006, 254440001, 254863004, 403198004, 415287001, 427186000, 448554008, 773624006, 860829004, 1144945000, 1157061002, 684941000119109, 1259350009, 1259514003, 1237491009, 1240346000, 91962001, 93186009, 93977003, 94145009, 94259009, 94366008, 94513006, 94535003, 94582005, 94998008, 109279002, 276821000, 371971003, 372013007, 404017001, 698287002, 703197004, 704253003, 707487003, 721623006, 721626003, 763884007, 1079681000119106, 109967007, 414166008, 1082261000119101, 1156414004, 350631000119102, 689491000119105, 1259589004, 1236955003, 1263652003, 91969005, 92347007, 94426008, 94544002, 94686001, 94711005, 95082005, 95152002, 109945006, 110980006, 126873003, 126991009, 188637007, 254795008, 363378008, 403998003, 404020009, 449101009, 699459002, 707582006, 707705008, 733349009, 771473004, 866094007, 91171000119107, 123631000119103, 123781000119107, 188590000, 143291000119104, 15931021000119108, 1196881000, 1268396008, 1268537000, 1268712008, 92006001, 94139001, 94422005, 94476005, 94485005, 94561004, 95013006, 109985000, 126494007, 126512005, 126658002, 187694000, 188001003, 188662007, 254434008, 254652000, 254787009, 278453007, 403858004, 403925009, 423005002, 445406001, 448465000, 707339009, 724468006, 778066006, 838355002, 352791000119108, 460051000124101, 460241000124108, 15632011000119105, 16908281000119101, 1259366006, 1259498004, 1259810001, 1260042002, 1237518004, 1251497007, 92424005, 92722008, 93225001, 93745008, 93994001, 94093001, 126639006, 127259005, 188090007, 189042009, 254429009, 254762002, 310526005, 372128005, 448319002, 449206007, 708921005, 721557009, 733136006, 773629001, 788982002, 870327007, 351761000119106, 352201000119105, 354131000119101, 1081751000119103, 89987007, 1156783003, 1157141006, 681991000119104, 685611000119101, 1255885008, 1259327003, 1259484007, 1259521003, 1259577001, 1263467005, 1268897007, 61493004, 92073008, 93651008, 94474008, 109351006, 188997007, 254462001, 278050001, 400058002, 419889000, 448144003, 712750007, 723366001, 786710002, 830025004, 351231000119109, 354101000119108, 1082291000119108, 687101000119100, 1082121000119102, 1157162007, 130941000119107, 682311000119102, 685851000119104, 1196815001, 1255890006, 1259520002, 1254750009, 1264498003, 1268901001, 92106003, 92478005, 92728007, 93952004, 94006002, 94168000, 94306005, 94400003, 94411006, 94562006, 94573004, 188733003, 254626006, 254856004, 369538008, 399931004, 404014008, 415110002, 415286005, 417647002, 425048006, 425688002, 448549004, 707464006, 725437002, 460111000124103, 109979007, 15951621000119102, 1079201000119101, 16823941000119108, 1255892003, 1259633002, 1259719005, 1254755004, 1268702001, 1268898002, 93784003, 93902008, 93940006, 188071004, 188089003, 188324003, 231827008, 254695001, 255011002, 313427003, 399911003, 399929008, 402819001, 404128004, 449497006, 704245006, 719475006, 721556000, 722527003, 766978002, 100721000119109, 118613001, 302181000119103, 1082171000119101,

1153393004, 15950141000119105, 353811000119104, 1197299006, 1197279000, 1254847003, 1255812002, 1263475004, 1268460005, 1268488004, 1268518009, 91995007, 92232000, 93469006, 94121005, 94529003, 94584006, 127000009, 188741003, 188803001, 189040001, 254412006, 254776001, 276953005, 307576001, 363464006, 371967001, 389272007, 398028009, 403815003, 447705002, 447730004, 448548007, 723854000, 724645006, 735917000, 840423002, 33441000119100, 105111000119109, 292591000119102, 16347551000119107, 1259427002, 1259437007, 1259803007, 1263466001, 20224008, 71404003, 92109005, 92119004, 93220006, 93712008, 93755007, 93854002, 93894000, 94068003, 94169008, 94495003, 94565008, 94839003, 95097008, 109843000, 126607004, 187994003, 188323009, 254451001, 254973003, 372116002, 404173006, 423619003, 710195004, 716649003, 716657000, 723843005, 733355004, 733598001, 735919002, 763666008, 99101000119101, 460281000124102, 94930003, 118617000, 1003465006, 1259251005, 1259447005, 1251471001, 92351009, 93729006, 94274006, 94371001, 126993007, 188454009, 202589002, 254425003, 372010005, 403884005, 403895004, 421249001, 428089008, 609515005, 707353009, 722954005, 763479005, 830031001, 866176003, 91141000119100, 236811000119101, 351851000119104, 1080101000119105, 1080671000119108, 11873701000119107, 16906881000119101, 16906921000119108, 203046000, 895354002, 1079141000119107, 1259727001, 1259731007, 1260021004, 1268536009, 92364005, 92455004, 93690007, 93972009, 94029005, 94512001, 94919001, 127271004, 254555008, 254706002, 254736006, 254758008, 276873004, 281564008, 314191009, 402563000, 403992002, 409660009, 448298007, 448738008, 460091000124107, 16594841000119106, 110985001, 1082231000119109, 1259439005, 1259457006, 1259535001, 1259626002, 1259726005, 93715005, 93777006, 94527001, 95077003, 95116001, 95546008, 126654000, 187693006, 188154003, 240163000, 254638002, 255004001, 307625008, 312115000, 371986009, 403540000, 404043000, 723554006, 133871000119103, 1076941000119104, 92511007, 682241000119109, 1259561002, 1259720004, 1268641007, 92282005, 92288009, 94118008, 94556001, 94782007, 95209008, 188649008, 274901004, 302851001, 363502009, 424849005, 448450001, 702785000, 707346000, 717922007, 721673009, 767814002, 830024000, 860819006, 68291000119109, 354351000119105, 11996401000119106, 1082051000119106, 1144928005, 1259399008, 1259623005, 92258000, 92330006, 93196000, 93900000, 93961004, 93985007, 94369001, 94752001, 109927006, 126624005, 187952001, 188004006, 188072006, 188729005, 254724000, 254730000, 277619001, 285607001, 285631006, 398943008, 400074007, 402813000, 404003005, 424276002, 424487008, 429570006, 443488001, 448147005, 703626001, 717736007, 721696009, 770685009, 773774000, 460081000124109, 188537004, 1080541000119102, 1156403002, 15977631000119105, 145194251000119106, 1255644001, 1259680005, 1259805000, 92359006, 92441007, 93693009, 94103007, 94208008, 94831000, 95000000, 109836006, 254629004, 254668006, 707424007, 682761000119103, 1080201000119101, 11315001000119103, 188516007, 404122003, 10737781000119101, 1081611000119106, 1144891007, 1157064005, 16527481000119100, 243971001000119103, 688891000119103, 1197340009, 1259334001, 1259408008, 1259429004, 1259715004, 1240351006, 1263653008, 1269458003, 1284830006, 92771003, 93169003, 94049001, 94089007, 94423000, 126753000, 127266006, 187641009, 188609000, 236019001, 254955001, 281563002, 307502000, 312359004, 372007003, 402541009, 403866008, 404038007, 433067002, 116811000119106, 147101000119108, 1081571000119102, 274897005, 890534002, 1156461003, 15951061000119102, 1217490003, 1259537009, 1236962007, 1269122003, 92714001, 93677000, 93901001, 102606000, 109924004, 126871001, 187861000, 254449000, 254505000, 363377003, 448377009, 448669004, 721641003, 721669006, 723077004, 773537001, 830028002, 866079008, 116871000119103, 1010669000, 1080021000119102, 1162286009, 1197324006, 1197260003, 1259387001, 1259400001, 1259412002, 1259724008, 1259748004, 1260116000, 1251494000, 13370002, 92704005, 93857009, 93888008, 188003000, 188500005, 254731001, 254748009, 255131000, 402544001, 402944008, 403917007, 408643008, 422782004, 449636007, 698048006, 707491008, 707592003, 722689005, 766048008, 87131000119104, 184801000119103, 254898001, 878799003, 1156780000, 12241031000119108, 145501000119108, 16908881000119100, 689451000119100, 1162584006, 1259368007, 1259585005, 1259776007, 1269045007, 92345004, 92529002, 93695002, 94687005, 94835009,

94900006, 95074005, 109378008, 118611004, 126497000, 126510002, 126596007, 188005007, 254432007, 254547001, 254851009, 307624007, 414553000, 448217003, 448731002, 709517003, 709830006, 716648006, 721547008, 128041000119107, 130351000119105, 460221000124101, 71101000119106, 1081321000119107, 1081701000119102, 400375351000119102, 1197266009, 1259474006, 1260067004, 1240352004, 92776008, 93193008, 93793002, 93815003, 93849006, 93966009, 94005003, 94196006, 94557005, 94764009, 94895007, 109267002, 109939003, 126744007, 126973004, 187941007, 188016000, 254445006, 254761009, 281367000, 285603002, 372133009, 398660000, 402518001, 402530007, 403935003, 448376000, 448993007, 449418000, 698011002, 703285005, 704255005, 707471001, 735680008, 783541009, 830026003, 184891000119109, 1079111000119108, 19886006, 424779008, 1078951000119101, 1079641000119101, 1080711000119107, 16527441000119105, 681611000119103, 1197328009, 1259325006, 1259433006, 1259822003, 1260031006, 1268355005, 92230008, 93943008, 94073009, 94117003, 94419008, 94612008, 109994006, 126918002, 126967000, 126977003, 188641006, 188946004, 189004003, 189080005, 231829006, 254714008, 255191003, 315036008, 369554006, 402515003, 402913004, 707662004, 722672002, 773641008, 783155007, 1079191000119104, 1079121000119101, 686431000119100, 1197294001, 1196865005, 1259403004, 1259567003, 1259823008, 48543002, 93826009, 94189009, 94236003, 94242004, 94386007, 126919005, 188002005, 254399000, 254635004, 254699007, 254726003, 363373004, 363398003, 371991005, 414676007, 447989004, 707703001, 713516007, 733134009, 766247009, 830030000, 15978071000119100, 16909041000119106, 1153349004, 351041000119109, 682481000119100, 688981000119107, 689281000119108, 1162441008, 1254781000, 1259394003, 1259763001, 1259780002, 1237483000, 1268640008, 92571001, 93679002, 93806005, 94133000, 94206007, 94537006, 95080002, 126527005, 188737002, 188746008, 188950006, 269469005, 303056000, 363352004, 363505006, 369460008, 403541001, 403893006, 413442004, 424190005, 445304002, 708504008, 715903004, 721563000, 721643000, 764938007, 122951000119108, 1080941000119109, 15951741000119103, 87141000119108, 1197283000, 1259402009, 1268966009, 1269052009, 92099001, 93915004, 94111002, 94359007, 94876000, 109842005, 109944005, 187652003, 188168005, 237468000, 254402004, 254465004, 254771006, 372003004, 402872002, 403886007, 404000008, 404104001, 448952004, 715426004, 721549006, 723847006, 723852001, 93495008, 94407000, 188579005, 277550009, 353801000119102, 1259395002, 1259489002, 1259790005, 1260092001, 1268533001, 92126004, 92396002, 95086008, 126574001, 126870000, 187640005, 187932002, 235002009, 239124001, 254452008, 447783007, 449260004, 710212007, 722676004, 723853006, 838344002, 431911000124102, 459411000124109, 10737561000119103, 1081891000119103, 1259337008, 1259760003, 1269047004, 92451008, 93201009, 94521000, 94632009, 255129009, 301873005, 307647008, 314419002, 369555007, 372124007, 397014001, 402540005, 402551005, 403973004, 425230006, 448436006, 723079001, 1076951000119102, 1082071000119102, 16906801000119109, 201101007, 15956661000119102, 1162346004, 92814006, 93868009, 189003009, 202425008, 254609000, 277573001, 403932000, 415283002, 425869007, 448447004, 123661000119106, 16301131000119109, 1197334002, 1259747009, 1268961004, 1269042005, 92384009, 93898002, 94200001, 94420002, 94645008, 109933002, 189120005, 372136001, 400174005, 440173001, 448378004, 449053004, 449259009, 707492001, 707528007, 712526008, 721636006, 722686003, 352311000119100, 1080261000119100, 353511000119101, 10737521000119108, 1081341000119101, 682361000119104, 688811000119107, 1179390007, 1259801009, 1259802002, 1268634001, 1268909004, 40312006, 92014007, 92479002, 93213004, 93705004, 94042005, 94841002, 95121003, 126590001, 126617009, 187688008, 187804005, 188917006, 188995004, 254416009, 277643003, 372027002, 372107001, 397015000, 402532004, 403902008, 403937006, 404138009, 405945003, 410812005, 420519005, 427285006, 723076008, 726721002, 733144006, 768926005, 353561000119103, 1079871000119107, 188576003, 402910001, 139681000119102, 16091371000119108, 16905461000119104, 681701000119107, 688731000119109, 16907241000119105, 1259450008, 1259481004, 1259569000, 1259723002, 1260076006, 91990002, 94012007, 94425007, 94655007, 94940000, 188013008, 188726003, 202584007, 231964001, 254457002, 255030002, 255066001, 369452007, 369454008, 369537003, 394728005,

403861003, 425657001, 448387008, 703429003, 707343008, 707432004, 707587000, 722683006, 724536000, 724552002, 733064004, 29431000119108, 682381000119108, 688931000119106, 16782771000119104, 1259341007, 1259428007, 1259615008, 1259783000, 1268965008, 92138008, 92143001, 92730009, 93202002, 93906006, 94039004, 109853004, 187866005, 277589003, 302855005, 313354001, 369540003, 399932006, 403892001, 424408000, 441559006, 448388003, 707484005, 721621008, 786038001, 117061000119101, 133841000119105, 1080241000119104, 1081741000119100, 1156456000, 15956741000119106, 352961000119103, 688751000119103, 1259392004, 1259692005, 1259736002, 1259786008, 1237477003, 1263997005, 1268351001, 92061008, 92104000, 92334002, 92352002, 94081005, 94301000, 95052004, 109283002, 127264009, 188156001, 189924002, 254837009, 255026000, 277545003, 302828001, 303012000, 369573005, 369590004, 385430001, 399969009, 402506005, 403857009, 424286001, 448254007, 449416001, 707385007, 96281000119107, 681161000119109, 93505001, 16906481000119103, 353081000119103, 1197263001, 1255649006, 1259566007, 1259770001, 1259782005, 112691000112107, 6331000, 92028001, 92124001, 92373002, 92432002, 93209006, 93686005, 93876006, 93941005, 94132005, 94416001, 94620005, 109368005, 187937008, 188102008, 188993006, 189288009, 254734009, 254745007, 254940005, 255025001, 255071008, 307617006, 363384006, 402504008, 402521004, 707451005, 722827008, 123701000119104, 353521000119108, 1082191000119100, 11313401000119108, 15959101000119103, 109965004, 203050007, 1079841000119100, 1079611000119100, 351051000119106, 688801000119109, 689781000119100, 1179389003, 1255393002, 1254734001, 1268958000, 92471004, 92748000, 94028002, 94412004, 95260009, 109277000, 109374005, 126613008, 255090000, 255128001, 307591004, 363745004, 372139008, 402539008, 402820007, 403467008, 403820003, 404013002, 448386004, 449629006, 717735006, 723887001, 116741000119104, 683691000119106, 898198006, 1156406005, 681971000119100, 1259396001, 1260058004, 1268391003, 39795003, 92453006, 92595004, 92628002, 93986008, 94030000, 94268006, 94337005, 95176005, 126511003, 188177003, 189317005, 231828003, 363466008, 385428003, 397012002, 402871009, 404078000, 445105005, 449220000, 733348001, 866069006, 98981000119103, 688121000119105, 688161000119100, 1079651000119104, 188517003, 1081761000119101, 10976711000119104, 1153425003, 16775311000119107, 16905541000119102, 16908321000119106, 688031000119107, 689401000119104, 302171000119101, 1255811009, 1259321002, 1259551009, 1259758000, 1237476007, 1254747006, 93658002, 93834003, 94017001, 94896008, 109359008, 109917000, 126591002, 126648001, 202444001, 202449006, 254790003, 255021005, 277571004, 278054005, 307341004, 312111009, 371990006, 397013007, 402495002, 403538005, 403927001, 404172001, 424887002, 446189008, 449377002, 870355006, 94591000119100, 15664241000119100, 1080471000119105, 894171571000119107, 1197265008, 1259409000, 1259582008, 1259735003, 1259777003, 1260061003, 1263465002, 1263912003, 91963006, 94079008, 94331006, 94570001, 95134002, 187661003, 239123007, 254975005, 286897001, 363391009, 371486001, 371993008, 372127000, 402498000, 448384001, 707360003, 721555001, 721629005, 723879002, 735679005, 16583041000119102, 1010675009, 16527561000119105, 352921000119108, 353061000119107, 1196874007, 1259451007, 1259497009, 1268908007, 92346003, 93913006, 94219006, 94278009, 94304008, 94379004, 94704006, 126562000, 126580009, 126875005, 189024004, 189371000, 254241004, 254701007, 254917006, 255032005, 363487003, 371989002, 402507001, 404011000, 448709005, 724578006, 681721000119103, 687891000119102, 689821000119105, 985355341000119101, 1255801002, 1259734004, 91987008, 93221005, 93820003, 93880001, 93981003, 93989001, 94124002, 94135007, 94241006, 94708009, 109998009, 126528000, 187829004, 202587000, 254501009, 277632006, 363493006, 363503004, 371481006, 372140005, 403201009, 404046008, 404062002, 422607004, 707467004, 711414003, 716593008, 721602000, 722517009, 766045006, 838343008, 87101000119106, 1079401000119100, 1081581000119104, 188515006, 15951021000119107, 16527521000119100, 1177173001, 1197327004, 1259326007, 1259353006, 1259438002, 1259702008, 68979007, 93681000, 93928006, 94116007, 94221001, 95036002, 95085007, 109269004, 109847004, 109919002, 109989006, 126499002, 126992002, 127262008, 187944004, 188050009, 188668006, 189258000, 202446004, 254843006, 255001009, 255068000, 307603002,

313356004, 363381003, 372101000, 400175006, 403817006, 404002000, 404108003, 417234002, 422541001, 443487006, 699354006, 707628004, 707660007, 722680009, 104981000119104, 1082331000119102, 895356000, 897648006, 1081901000119104, 15955901000119104, 1196903001, 1255192000, 1259722007, 1237527003, 1269043000, 92811003, 93778001, 94263002, 109288006, 126604006, 126642000, 254563009, 255148008, 304524009, 414950005, 417181009, 447738006, 448775003, 722665003, 725138002, 838342003, 11471000224106, 24111000119105, 460031000124108, 11305601000119109, 1082141000119108, 1237057000, 1237521002, 92047003, 92729004, 93694003, 94187006, 94233006, 94498001, 94525009, 126519001, 188982009, 254844000, 254852002, 255185000, 281560004, 285642002, 372020000, 403729006, 403870000, 403920004, 404120006, 448451002, 714463003, 866052009, 118601006, 1148851002, 16845341000119105, 1255753002, 1259356003, 1259419006, 1259448000, 1259821005, 92472006, 93194002, 93699008, 93978008, 94123008, 110013004, 187940008, 189289001, 277587001, 363462005, 402945009, 403712008, 423361002, 707593008, 733352001, 733359005, 82291000119105, 91151000119103, 683741000119108, 1080661000119102, 1082111000119109, 1187464007, 1259773004, 92739005, 93187000, 94092006, 94353008, 94912005, 95133008, 126651008, 126748005, 126802007, 187950009, 187978005, 188074007, 188944001, 202437005, 202909008, 231831002, 254559002, 255027009, 403891008, 404015009, 446352002, 447390000, 702391001, 707337006, 707466008, 721620009, 723174009, 353661000119102, 423294001, 143401000119106, 1080691000119109, 1099571000119100, 1259749007, 1259768005, 1268350000, 19133005, 92003009, 92309002, 92377001, 93781006, 94522007, 109369002, 126994001, 187801002, 188327005, 254431000, 254567005, 254670002, 254820002, 271943005, 274085008, 277601005, 302849000, 312112002, 369461007, 449732002, 698044008, 700488005, 721635005, 788979007, 7391000119103, 21851000119103, 291901000119109, 897647001, 1079061000119107, 1144924007, 16774671000119108, 685671000119109, 1259343005, 1259375008, 1259678004, 1259738001, 1237034006, 1268484002, 1268503008, 1268718007, 92225009, 92234004, 92459005, 92734000, 93217003, 93636004, 93713003, 93740003, 93908007, 93920004, 94511008, 94654006, 126582001, 126790004, 187824009, 188061001, 188498009, 255008003, 276829003, 277625002, 403968005, 404039004, 443136000, 449269003, 113141000119107, 350911000119102, 1079831000119109, 1081841000119106, 1156472008, 689501000119103, 1208460000, 1259422008, 1259817003, 1260082009, 1260085006, 94019003, 94032008, 94108003, 95043008, 188022009, 254768003, 269580008, 275397008, 363488008, 372087000, 404093001, 703271000, 352321000119107, 687121000119109, 15951821000119103, 109972003, 255007008, 1081281000119102, 1153397003, 685621000119108, 682461000119109, 1231639008, 1254806001, 1255629002, 1259313008, 1237032005, 1237519007, 92389004, 93711001, 93969002, 94341009, 94564007, 94613003, 94952007, 95100003, 126491004, 126698000, 126917007, 126949007, 126990005, 188063003, 202579001, 202588005, 254703005, 277618009, 363375006, 403202002, 404103007, 426217000, 448401007, 717921000, 866102002, 101351000119105, 456911000124108, 459391000124109, 1078901000119100, 40467008, 402881008, 15632131000119104, 685831000119105, 687821000119104, 1251445000, 1268389006, 1268637008, 92337009, 93671004, 93782004, 94083008, 94319009, 94444000, 94464005, 126560008, 188272000, 188501009, 188974009, 241861008, 371995001, 373083005, 404034009, 404072004, 418686001, 721558004, 723849009, 766248004, 830151004, 61471000119100, 351211000119104, 353421000119109, 15664361000119105, 109964000, 203052004, 1156418001, 681781000119104, 685721000119101, 1208483002, 1259509002, 1259602004, 1259704009, 1259812009, 92256001, 92392000, 92745002, 92795001, 93182006, 93707007, 93726004, 93830007, 94050001, 94094007, 94508007, 94588009, 95194004, 109840002, 109947003, 126504002, 187635002, 188996003, 189058007, 202428005, 255019000, 271476003, 302842009, 398903003, 402556000, 403203007, 404111002, 425941003, 449173006, 707531008, 721640002, 116841000119105, 208041000119100, 292581000119100, 354701000119107, 681631000119108, 123761000119103, 683131000119100, 1197311006, 1255723009, 1259378005, 1259462007, 1259820006, 1237048006, 1263468000, 1268462002, 1268698000, 92001006, 92064000, 92227001, 93893006, 94672000, 109839004, 126583006, 126996004, 202590006,

202592003, 252997008, 255028004, 277613000, 301872000, 314951005, 363382005, 372019006, 403862005, 403961004, 403999006, 404162004, 423121009, 423278008, 449319009, 449649002, 698013004, 699659007, 188569008, 733627006, 78461000119105, 688681000119100, 1237340007, 1259453005, 1259466005, 1259493008, 1268956001, 93767009, 94253005, 238904008, 254694002, 255018008, 302819002, 369588000, 403869001, 404115006, 430338009, 440422002, 449153001, 722953004, 723173003, 724647003, 139661000119106, 203053009, 359640008, 682451000119107, 684921000119103, 1259442004, 1259518000, 1268888008, 7199000, 92623006, 94569002, 95054003, 126608009, 126980002, 187973001, 237063001, 254441002, 254645002, 254815002, 255146007, 371988005, 402505009, 402511007, 402513005, 404067008, 424666005, 447781009, 449204005, 449254004, 713038003, 719045009, 722512003, 735735001, 1148845007, 15950221000119108, 687861000119109, 688411000119109, 681921000119101, 1254801006, 1255754008, 1259732000, 1259816007, 1268461009, 92142006, 92291009, 93139005, 93751003, 94352003, 94483003, 94678001, 189509003, 231834005, 254397003, 255069008, 369558009, 397009000, 707583001, 707595001, 717731002, 721541009, 764990003, 9541000119105, 67841000119103, 302271000119107, 109921007, 188547001, 1156807006, 1157136003, 15949941000119101, 352461000119107, 685901000119108, 1197342001, 1254691000, 1254733007, 1268633007, 1268699008, 92284006, 92614008, 93654000, 94033003, 94148006, 95089001, 187811009, 187869003, 187997005, 188261005, 231824001, 314987003, 369556008, 403207008, 403894000, 449108003, 721540005, 735332000, 135091000119106, 302221000119106, 683451000119103, 109392002, 1141661004, 353791000119103, 689471000119109, 1231697007, 1255645000, 1255888005, 1259384008, 1259775006, 1237033000, 1268544009, 1268563005, 92398001, 93135004, 93747000, 94020009, 94163009, 94334003, 94391008, 188613007, 188627002, 188778000, 254561006, 254710004, 269467007, 277602003, 403824007, 403939009, 449308006, 699704002, 707485006, 707493006, 716788007, 722668001, 737359009, 61311000119104, 133861000119109, 1156812007, 682291000119101, 1217301006, 1259452000, 1259668002, 1259755002, 1259796004, 62727008, 92024004, 92353007, 93963001, 94205006, 94496002, 94499009, 94586008, 126774004, 187653008, 254398008, 254410003, 707395000, 735921007, 1079331000119101, 1079691000119109, 11312841000119102, 15956341000119105, 359648001, 878858004, 689771000119103, 1255658004, 1259756001, 1254730005, 93742006, 93828005, 94153001, 94305009, 95055002, 95230004, 109828002, 126531004, 126752005, 126757004, 187809000, 188076009, 231833004, 234147002, 254909003, 255005000, 255012009, 370468006, 372103002, 403954003, 422479008, 424993006, 445448008, 707460002, 716859000, 722543005, 734099007, 768934004, 830017001, 459401000124106, 1080121000119101, 12240951000119107, 93525002, 404143002, 123721000119108, 1079021000119102, 1080571000119109, 354391000119100, 1259344004, 1259825001, 1268546006, 39629007, 92371000, 93983000, 94013002, 94176003, 94610000, 94861008, 95150005, 109942009, 126619007, 187870002, 254817005, 285605009, 285616002, 302845006, 387891008, 399972002, 403863000, 404119000, 440525003, 707355002, 713290004, 733608000, 68271000119108, 145831000119103, 460261000124107, 684911000119105, 1076921000119105, 15956381000119100, 188511002, 1082131000119104, 1156495004, 1156772007, 1156808001, 1157071000, 16899941000119102, 16907691000119100, 687361000119101, 1186616001, 1259369004, 1259479001, 1259769002, 1236942004, 92367003, 92758001, 93214005, 94099002, 94429001, 95413004, 109857003, 126589005, 188868008, 188937005, 254415008, 254444005, 254655003, 254718006, 402525008, 402882001, 404147001, 413389003, 425178004, 449063007, 707453008, 707489000, 721699002, 15957181000119104, 353111000119108, 1281812003, 92292002, 93700009, 93817006, 93845000, 94075002, 94587004, 94776009, 95094001, 95102006, 126986008, 188487008, 188754005, 188779008, 253003009, 276822007, 277551008, 277568007, 277579002, 313353007, 372008008, 404063007, 427056005, 448315008, 449177007, 449221001, 449419008, 724649000, 724806004, 733603009, 680821000119103, 109968002, 398838000, 895355001, 1080041000119108, 1153348007, 1259336004, 1259424009, 1259500003, 1254723004, 1268349000, 1268704000, 93663003, 93825008, 94193003, 109274007, 126506000, 187977000, 189057002, 238905009, 254794007, 286899003, 372126009, 372135002, 403859007, 424229006, 424260006, 449174000, 710196003,

723081004, 723164006, 733357007, 11312921000119100, 895111002, 1157060001, 15951181000119107, 16908961000119102, 1196864009, 1259475007, 1264115008, 23150001, 94833002, 94868002, 95028006, 126811007, 187660002, 187884007, 188042000, 188055004, 189002004, 202434003, 236513009, 254430004, 254435009, 254995004, 269617008, 307651005, 363388009, 402510008, 408642003, 697993003, 699955004, 707575007, 717003001, 721306009, 721567004, 772992009, 1079171000119100, 11304841000119108, 109386008, 188679001, 16907641000119108, 352731000119109, 689231000119107, 1186630006, 1201780008, 1255887000, 1259414001, 1259458001, 1259789001, 1259814005, 1236964008, 1268957005, 1268964007, 92002004, 93749002, 94479003, 94626004, 95056001, 109366009, 109371002, 126650009, 189401003, 237065008, 254782003, 255188003, 402552003, 403822006, 707486007, 722514002, 770684008, 866050001, 352001000119100, 11010461000119101, 16314751000119104, 1079781000119101, 1082091000119101, 687351000119103, 1259750007, 1259797008, 1268499002, 92724009, 93709005, 94322006, 94351005, 94629006, 95123000, 126972009, 127229000, 188009001, 254539001, 363374005, 363399006, 373088001, 402538000, 403901001, 404140004, 448468003, 721670007, 722511005, 866073009, 897558001, 1081651000119107, 1259388006, 1259779000, 1237471002, 1237526007, 92369000, 93471006, 93640008, 93789008, 93995000, 94034009, 94408005, 94549007, 94579000, 126588002, 126867004, 187929000, 254712007, 254948003, 254993006, 277615007, 313248004, 704247003, 707468009, 707579001, 724553007, 323621000119108, 349361000119109, 457331000124100, 1079231000119108, 683031000119101, 91211000119109, 1259454004, 1259761004, 1260084005, 92643000, 93675008, 93850006, 93889000, 94314004, 94709001, 126568001, 126893006, 187857006, 300988009, 307636001, 403911008, 403938001, 416510003, 426134002, 448229007, 449156009, 723850009, 96981000119102, 113191000119104, 681171000119103, 12240991000119102, 274142002, 404136008, 688141000119104, 1153377008, 682251000119106, 1163347004, 1231267007, 1237474005, 93648001, 93657007, 93970001, 94172001, 188100000, 254888007, 269516007, 372023003, 372106005, 404052009, 404068003, 448560008, 721548003, 830027007, 188566001, 878857009, 1081331000119105, 16229011000119100, 1255648003, 1259322009, 92358003, 92375009, 93185008, 94027007, 94574005, 94666000, 94813001, 95188007, 109830000, 109835005, 126737003, 127256003, 187956003, 188458007, 188734009, 202441009, 254707006, 275265005, 277570003, 281562007, 402516002, 403821004, 404058008, 415172002, 422599000, 713249002, 1079241000119104, 109969005, 1197359006, 1217617009, 1259312003, 1254725006, 1254745003, 1263469008, 92317005, 92477000, 92642005, 94025004, 94197002, 94399005, 94409002, 94805002, 94951000, 187683004, 187881004, 188152004, 254423005, 254667001, 255123005, 314966008, 402508006, 402537005, 424952003, 440527006, 444596001, 448712008, 449224009, 449627008, 707389001, 733360000, 764961009, 783771003, 91081000119109, 353671000119108, 354591000119108, 1079801000119102, 1079901000119107, 15974541000119108, 188538009, 16909161000119109, 10737601000119103, 1141621000, 685731000119103, 687911000119100, 1259698009, 1268911008, 1269050001, 1269412004, 92164002, 92610004, 93144003, 93909004, 94292003, 94424006, 94530008, 94555002, 94802004, 94991002, 94992009, 109978004, 188138001, 188994000, 254436005, 254744006, 269544008, 280960002, 286895009, 363456000, 372119009, 372142002, 402818009, 403208003, 403484002, 418590005, 422968005, 423494003, 449631002, 707539005, 113101000119105, 16314671000119104, 93531004, 188632001, 10737401000119100, 1080451000119101, 249389151000119103, 353841000119100, 1259426006, 1259714000, 1259717007, 92191000, 92747005, 93218008, 93733004, 93768004, 94157000, 94210005, 94418000, 109285009, 126773005, 128519009, 187606005, 187991006, 188075008, 188095002, 254722001, 255052006, 312114001, 402494003, 404131003, 721310007, 721569001, 789690008, 860923004, 871843002, 18121000119104, 94291000119103, 117111000119100, 878805009, 1081631000119101, 683121000119103, 689361000119108, 354211000119101, 354191000119102, 1259318004, 1269049001, 92039003, 92376005, 93637008, 93660000, 93821004, 94804003, 94882002, 189156008, 189291009, 254980001, 326072005, 363435002, 369458006, 369487007, 402527000, 419977003, 424151006, 425127006, 707379000, 721642005, 765740002, 87111000119109, 99121000119105, 354171000119103, 686441000119109, 1162855005, 1259320001, 1259352001, 1259421001,

1259682002, 1259690002, 92224008, 92417009, 93843007, 93959008, 94384005, 94942008, 94996007, 109823006, 109878000, 109885001, 254908006, 255006004, 274084007, 277614006, 278044006, 307226002, 307610008, 372115003, 402550006, 402817004, 447953007, 707357005, 707394001, 712849003, 721582009, 722455002, 136641000119101, 53435002, 255115008, 688671000119103, 428487561000119102, 1254845006, 1255895001, 1268519001, 1268707007, 46795000, 92031000, 92128003, 93189002, 93697005, 93746009, 93905005, 94018006, 94502008, 110000005, 126492006, 202580003, 254729005, 277616008, 286896005, 314512009, 363376007, 372000001, 399884001, 403898002, 403926005, 404082003, 427642009, 443333004, 699358009, 704249000, 707399006, 713747004, 721562005, 721632008, 721697000, 722673007, 1080641000119101, 15957141000119109, 352421000119102, 681821000119109, 686981000119108, 1172793008, 1197275006, 1237479000, 16652001, 60620005, 93934004, 94104001, 94838006, 95092002, 126982005, 187949009, 188274004, 254698004, 271467005, 285634003, 363437005, 372137005, 396198006, 402840000, 423032007, 707409003, 707591005, 722510006, 726652005, 789700005, 350921000119109, 93487009, 404124002, 1081671000119103, 1156804004, 1157157006, 15951341000119104, 1197259008, 1255905007, 101631000119108, 1269051002, 92737007, 93670003, 93723007, 93867004, 94008001, 94086000, 94339008, 94638008, 94669007, 94676002, 94911003, 95068005, 126493001, 188770007, 310605004, 373091001, 403311004, 716588005, 733163007, 61291000119103, 354491000119109, 1079551000119100, 1081921000119108, 354461000119102, 354731000119100, 1162361003, 1208741006, 1259505008, 1268487009, 93156002, 93646002, 93730001, 94105000, 94302007, 94893000, 109291006, 126690007, 126793002, 126978008, 187662005, 188505000, 277604002, 372015000, 372018003, 403888008, 403914000, 403951006, 403955002, 404041003, 420835009, 449307001, 449635006, 1079151000119109, 188586002, 1080751000119108, 16905421000119109, 681711000119105, 1255395009, 1259310006, 1259349009, 1259358002, 1237035007, 78745000, 92090002, 92443005, 92705006, 93948004, 94484009, 94554003, 94605004, 254796009, 254904008, 254915003, 255015006, 372016004, 387893006, 403887003, 404007006, 703531009, 707406005, 715901002, 765328000, 830196006, 860831008, 866068003, 113231000119108, 93532006, 93550004, 1010653007, 1081851000119108, 1144922006, 1196839006, 1259345003, 92395003, 93722002, 94026003, 94461002, 94477001, 126623004, 126656003, 126976007, 127257007, 187814001, 187996001, 285309005, 314952003, 402502007, 449067008, 723009000, 726019003, 12301000132103, 67831000119107, 122981000119101, 15951661000119107, 681591000119108, 688091000119106, 16907161000119103, 1259329000, 1259445002, 1259674002, 92255002, 92307000, 93134000, 93210001, 93783009, 93946000, 93964007, 94941001, 126586003, 187757001, 202432004, 254407005, 254586002, 363433009, 402542002, 403883004, 404114005, 422706003, 423691004, 424938000, 707580003, 722828003, 735918005, 827186009, 351151000119105, 1157068008, 15977591000119105, 683981000119101, 1197296004, 1255191007, 1259669005, 1259707002, 1260087003, 1236953005, 1268711001, 92070006, 92360001, 92716004, 93708002, 94143002, 94158005, 94517007, 94933001, 126556005, 126963001, 237457000, 254651007, 254838004, 254850005, 254900004, 254983004, 402517006, 402546004, 403881002, 418439007, 449633004, 707356001, 723846002, 723851008, 724648008, 113251000119102, 1156455001, 1156797006, 1179375008, 1255651005, 1259444003, 1259767000, 1251493006, 93137007, 93754006, 93922007, 109874003, 109880006, 126625006, 126657007, 188147009, 188744006, 188983004, 254622008, 277622004, 285312008, 310600009, 423114009, 717734005, 721625004, 78411000119107, 459371000124108, 460061000124104, 12246641000119104, 681661000119100, 1197307000, 1255800001, 1259443009, 1259592000, 1259772009, 1259806004, 92189008, 92316001, 93833009, 94059000, 94670008, 94842009, 95048004, 126770008, 187767006, 188077000, 202429002, 254393004, 363372009, 363440005, 402523001, 402545000, 403945001, 36851000119109, 96141000119105, 139651000119109, 109390005, 123751000119100, 354331000119104, 1187519007, 1197262006, 1230318000, 1255631006, 1259762006, 1236944003, 1254753006, 12008008, 13048006, 93140007, 93151007, 93903003, 94338000, 94398002, 94510009, 94630001, 94646009, 94857002, 126594005, 126779009, 188972008, 195469007, 254543002, 254809002, 278697001, 281566005, 372141009, 404021008, 404089007, 443261008, 707465007,

714251006, 766980008, 148911000119107, 15709841000119107, 1010677001, 1141622007, 15950061000119105, 1197267000, 1197355000, 1259440007, 1259740006, 1237052006, 1268721009, 93203007, 93741004, 93766000, 93804008, 94855005, 95130006, 95154001, 109922000, 126592009, 187803004, 202582006, 254466003, 254503007, 254812004, 277623009, 312820003, 314954002, 369591000, 372009000, 385429006, 404109006, 431396003, 444910004, 449289002, 707347009, 707402007, 709214009, 721628002, 722451006, 722682001, 722713005, 723165007, 12281000132104, 1079761000119105, 1156453008, 687961000119102, 1172840000, 1162348003, 1230023005, 1231381006, 16782731000119102, 1259627006, 1236945002, 1268697005, 92046007, 92379003, 93886007, 93979000, 94053004, 94277004, 94372008, 94401004, 94737008, 95071002, 95263006, 109289003, 126522004, 126577008, 127004000, 188796009, 189417009, 202578009, 255180005, 255195007, 276870001, 307637005, 363370001, 398679002, 403823001, 404010004, 404064001, 404153001, 414785000, 423280002, 444374006, 448609001, 707396004, 707686002, 721762007, 733471003, 866093001, 302301000119109, 15664281000119105, 1079601000119103, 1153423005, 1162799008, 1259260002, 1259613001, 1237489001, 92434001, 93682007, 93932000, 94177007, 109833003, 126738008, 188006008, 188971001, 254739004, 254829001, 277473004, 363494000, 371977004, 426964009, 449064001, 449634005, 709031009, 724650000, 773995001, 113201000119101, 1079771000119104, 1082221000119106, 11312361000119106, 307691000119108, 1080731000119102, 1259380004, 1254740008, 1268541001, 92068002, 92429000, 92605003, 93885006, 94023006, 94321004, 94373003, 109349007, 109858008, 188748009, 202438000, 240204004, 254770007, 255003007, 307646004, 425749006, 707405009, 707479004, 722453009, 733353006, 783706007, 865954003, 96291000119105, 99131000119108, 354041000119106, 1082301000119109, 16301051000119102, 315058005, 372131006, 10737821000119106, 682261000119108, 689801000119101, 1255907004, 1259373001, 1259389003, 1259800005, 1260088008, 1263476003, 1268548007, 1268642000, 1268706003, 92290005, 92710005, 93797001, 93808006, 94385006, 94431005, 94667009, 109923005, 126593004, 126606008, 126989001, 254841008, 363394001, 363490009, 423987006, 426642002, 427658007, 445476000, 449223003, 449306005, 707469001, 721639004, 722377004, 816201004, 15636991000119103, 231830001, 897650003, 1081861000119105, 15938781000119103, 1196841007, 93942003, 94376006, 94390009, 94715001, 95090005, 188493000, 188952003, 189277003, 255000005, 255093003, 363463000, 372004005, 372065009, 403908007, 403974005, 447687004, 722542000, 61321000119106, 91041000119104, 113071000119101, 133131000119107, 460291000124104, 1079731000119102, 895361003, 1148847004, 16527641000119107, 16907741000119103, 1196844004, 1217383007, 1254848008, 1259507000, 1251467004, 1263786003, 93716006, 93982005, 94109006, 94382009, 94806001, 94947002, 95065008, 109886000, 126572002, 127225006, 202595001, 254448008, 254535007, 254696000, 254893005, 254912000, 254998002, 276952000, 297134004, 369513003, 372028007, 404139001, 446807009, 702446006, 703295003, 782667002, 184781000119102, 452291000124109, 307608006, 16908681000119104, 16909121000119104, 16218761000119102, 1259351008, 1259655005, 1268717002, 92422009, 92463003, 93142004, 93183001, 93753000, 93951006, 94488007, 94543008, 95022007, 109294003, 126517004, 126775003, 201091002, 202427000, 277505007, 307597000, 310524008, 363363001, 404054005, 404066004, 702786004, 707458004, 708971008, 721604004, 721617001, 721671006, 19191000119106, 67811000119102, 354051000119108, 1079631000119105, 354301000119106, 1259564005, 92229003, 92378006, 92726006, 92794002, 93223008, 94433008, 94487002, 94542003, 95023002, 95143006, 109273001, 126897007, 126920004, 126970001, 187659007, 188273005, 188725004, 254243001, 254450000, 254658001, 285608006, 363414004, 403206004, 403819009, 404045007, 414666009, 447951009, 707348004, 707422006, 713293002, 722509001, 830055006, 90811000119100, 130381000119103, 352071000119105, 354071000119104, 683271000119108, 1079261000119100, 1080151000119109, 105041000119109, 351841000119101, 683791000119100, 689311000119105, 1162353008, 1197032007, 1197323000, 1196846002, 1260065007, 1260100002, 1236947005, 1254752001, 92332003, 92551005, 92630000, 93152000, 93704000, 93918002, 94184004, 94576007, 94852008, 109281000, 126771007, 188132000, 188151006, 188949006, 254570009, 302857002, 307599002, 312937006, 363492001,

372011009, 402548003, 403912001, 403989001, 403994001, 420890002, 422378004, 449216004, 698042007, 709202001, 789004008, 838350007, 1153336009, 1156809009, 16909001000119109, 1255906008, 1259468006, 1259587002, 1259751006, 1259754003, 1259787004, 1260089000, 1237053001, 1268501005, 1269046008, 92011004, 92060009, 92390008, 93227009, 94040002, 94299007, 94617002, 94650002, 95224004, 109286005, 126489007, 188015001, 188633006, 254802006, 277580004, 399934007, 403987004, 429661002, 707538002, 723855004, 726654006, 763064007, 460251000124105, 1078961000119104, 15710041000119107, 1196868007, 1259316000, 1259510007, 1259575009, 1237485007, 1268632002, 1268703006, 93659005, 93874009, 94482008, 109278005, 126587007, 187692001, 187999008, 202593008, 254456006, 275394001, 307219002, 399970005, 404133000, 404145009, 415112005, 422508004, 423447006, 447990008, 448561007, 449294002, 707393007, 707576008, 722674001, 733350009, 1079211000119103, 11309921000119106, 1148551007, 1156471001, 1208461001, 1196898000, 1197354001, 1259371004, 1259791009, 92169007, 92357008, 93230002, 93933005, 94038007, 109356001, 126702001, 128466006, 187993009, 188669003, 233718008, 254720009, 274900003, 369450004, 369455009, 403909004, 404150003, 703230006, 703284009, 709205004, 724644005, 767544007, 1080161000119106, 109971005, 192681000119104, 1080681000119106, 1081911000119101, 1082211000119104, 1162264008, 1255641009, 1255642002, 1237472009, 93143009, 93215006, 93761005, 94057003, 94380001, 94768007, 94888003, 95044002, 109875002, 109975001, 188675007, 254460009, 255137001, 271568003, 278042005, 285633009, 372130007, 404012007, 419317000, 424302003, 449733007, 707430007, 707473003, 716774008, 860830009, 116691000119101, 351311000119107, 351461000119100, 352161000119100, 1079221000119105, 11311721000119107, 203049007, 1078941000119103, 1079181000119102, 682411000119106, 354151000119107, 1259648008, 1260068009, 1237424008, 1251473003, 92016009, 92440008, 92699002, 92746001, 93470007, 93683002, 93875005, 93939009, 94333009, 94577003, 127218004, 187709007, 233940007, 254831005, 255024002, 373089009, 419240004, 421246008, 423349005, 443936004, 447782002, 698200003, 703286006, 707403002, 718604008, 722519007, 722670005, 733469003, 737308008, 764845008, 80071000119103, 15978151000119108, 187716008, 1080931000119100, 1251475005, 1264066001, 1268959008, 92741006, 92817004, 94472007, 94593007, 94995006, 109383000, 126618004, 188163001, 188990009, 202424007, 254413001, 254582000, 269616004, 286891000, 369535006, 402524007, 403458008, 448231003, 449176003, 703625002, 707378008, 715950008, 721624000, 722524005, 722795004, 724587002, 349381000119100, 353951000119106, 1080651000119104, 1153346006, 683711000119109, 682441000119105, 1197349005, 1259436003, 1259456002, 1259548002, 92184003, 92544001, 93923002, 94010004, 94207003, 94558000, 94575006, 94652005, 94913000, 94944009, 95066009, 126579006, 126778001, 254750001, 255136005, 313249007, 403875005, 404018006, 404130002, 449163009, 699318007, 699657009, 702368000, 707483004, 721574009, 721645007, 722530005, 78501000119105, 94682004, 16636051000119105, 16774551000119100, 350851000119109, 687771000119106, 1196883002, 1259333007, 1259459009, 1259737006, 1259792002, 1260056000, 92315002, 92558004, 92789002, 93737003, 93936002, 94036006, 94061009, 94062002, 94190000, 94421003, 94592002, 109920008, 126502003, 126969002, 189151003, 313357008, 372113005, 402514004, 402564006, 404057003, 413537009, 423106003, 423615009, 721559007, 764847000, 780844005, 789051005, 838341005, 866096009, 82761000119108, 91161000119101, 15974581000119103, 1079591000119105, 689211000119102, 1196853006, 1259417008, 1259441006, 1259601006, 1268902008, 92313009, 92572008, 93759001, 94507002, 95168008, 109834009, 126516008, 188099008, 188736006, 188951005, 202435002, 254693008, 255108000, 369453002, 369571007, 372143007, 385431002, 400001003, 403910009, 404016005, 404155008, 416842003, 423464009, 423506005, 425376008, 707400004, 709210000, 713739004, 716650003, 721618006, 722712000, 724577001, 135931000119106, 302211000119104, 353431000119107, 1081811000119107, 231835006, 420302007, 686451000119106, 1259383002, 1259596002, 1259681009, 92394004, 92449009, 92731008, 93661001, 93725000, 93984006, 94387003, 126728003, 187614004, 264561009, 302838006, 363504005, 372021001, 403897007, 404070007, 404129007, 422691006, 424422000, 442528007, 445238008, 448607004, 707588005, 722679006, 764100007,

354361000119107, 890529001, 1080501000119104, 1153379006, 1259616009, 1259624004, 1268900000, 92576006, 93931007, 94509004, 94795002, 95051006, 95063001, 126792007, 188934003, 189259008, 189280002, 254395006, 285307007, 369592007, 372001002, 403949007, 422676009, 448214005, 448667002, 707411007, 707454002, 733358002, 1080231000119108, 128875000, 1148906001, 688861000119105, 1197344000, 1259418003, 91959004, 92078004, 93703006, 93727008, 93987004, 94125001, 94624001, 95040006, 109280004, 187995002, 188642004, 188777005, 254992001, 255124004, 363348004, 363491008, 712525007, 713718006, 722718001, 762315004, 816204007, 94144008, 1079661000119102, 1156459007, 11717361000119108, 15957261000119102, 352771000119107, 91191000119108, 1163568002, 16832981000119109, 1259347006, 1259541008, 1259664000, 92718003, 93219000, 93647006, 93929003, 93975006, 126576004, 127261001, 188263008, 188361007, 189279000, 231825000, 371987000, 372002009, 372120003, 402652009, 403742006, 442348004, 448015002, 707354003, 707358000, 707481002, 724866001, 764951002, 769168000, 770601003, 866101009, 84811000119107, 101371000119101, 354641000119104, 92656007, 402912009, 105051000119106, 1079621000119107, 1080741000119106, 1153421007, 16782321000119104, 16905501000119104, 350881000119102, 1197298003, 1259612006, 1260081002, 101611000119103, 1236960004, 1268348008, 1268354009, 26904005, 92645007, 93836001, 94228007, 109911004, 189006001, 313355000, 369529006, 403971002, 416901002, 419842002, 426248008, 704248008, 830036006, 94601000119107, 460101000124101, 1080131000119103, 1080191000119104, 118614007, 687881000119100, 1196852001, 1231180000, 1259774005, 1259826000, 1268631009, 1269414003, 92226005, 93792007, 93858004, 93883004, 94329002, 94578008, 94732002, 95175009, 95231000, 126614002, 126777006, 126987004, 187988006, 188155002, 188745007, 254634000, 254833008, 285310000, 302836005, 363368005, 373080008, 404051002, 413656006, 733347006, 15632091000119101, 240531002, 1082061000119108, 681831000119107, 91231000119104, 1187209002, 1196820001, 1208451008, 681221000124106, 92503002, 93184007, 93674007, 93818001, 109295002, 188612002, 188939008, 254803001, 314955001, 372026006, 372100004, 402547008, 403880001, 403942003, 420524008, 448296006, 448707007, 723848001, 724591007, 184741000119107, 683251000119104, 15638331000119104, 780821007, 781641005, 15956261000119100, 682321000119109, 1259342000, 1259525007, 1259688003, 1236961000, 1251458004, 1263601000, 1268561007, 92236002, 92342001, 94072004, 94090003, 94238002, 94261000, 94430006, 94710006, 95146003, 126605007, 126627003, 188125009, 202585008, 363434003, 369589008, 371012000, 372094002, 402880009, 404059000, 404110001, 448273006, 448300007, 448868009, 449222008, 707380002, 707384006, 721633003, 733351008, 147131000119101, 350731000119105, 350901000119100, 352231000119103, 681141000119105, 688131000119108, 188524002, 308121000, 1079031000119104, 1153344009, 355141000119106, 1162349006, 1255193005, 1236969003, 91858008, 93642000, 93644004, 93769007, 93997008, 94159002, 94999000, 95073004, 126573007, 188973003, 254549003, 275395000, 277629008, 281366009, 285308002, 372006007, 372022008, 372098004, 399985004, 444604002, 448665005, 707584007, 707664003, 717968005, 764694005, 353741000119106, 680831000119100, 1082701000112100, 15635801000119106, 109350007, 351741000119107, 455733521000119103, 1197310007, 1259416004, 1259516001, 1259578006, 1259593005, 1259795000, 36982001, 58961005, 92231007, 92569001, 92742004, 93138002, 93226000, 93863000, 94007006, 94165002, 94725006, 109856007, 126615001, 126785002, 126865007, 276418007, 403928006, 403956001, 416864007, 422736007, 424132000, 449141000, 721578007, 352301000119103, 1079161000119106, 1080091000119100, 1080181000119102, 109389001, 878798006, 1080621000119107, 688691000119102, 1259315001, 1259650000, 1259788009, 1260079004, 1237478008, 1263474000, 1264497008, 83448006, 93662008, 93968005, 94589001, 95156004, 126567006, 126795009, 188445006, 253042009, 254418005, 254438006, 277575008, 311779007, 363360003, 385427008, 399954008, 423607006, 448259002, 448672006, 448994001, 722425009, 733926004, 766757006, 866098005, 116821000119104, 1141627001, 1148852009, 16906601000119108, 16908921000119107, 687801000119108, 688321000119107, 1259659004, 1259804001, 1259808003, 92818009, 94056007, 94180008, 94945005, 95047009, 95129001, 126585004, 126629000, 126876006, 188265001, 188504001,

240207006, 254557000, 277651000, 312949007, 363393007, 372123001, 402635005, 402867006, 403981003, 423421002, 423708008, 426370008, 429114002, 448449001, 704246007, 707495004, 707704007, 709471005, 46041001, 93451002, 109977009, 402842008, 1003406009, 16699241000119100, 1259338003, 1259348001, 1259553007, 1259666003, 1268906006, 44721000087109, 84449007, 92136007, 93204001, 93973004, 94047004, 94060005, 94181007, 94653000, 126581008, 126646002, 126671004, 187675005, 187925006, 254421007, 255102004, 275399006, 281499005, 302856006, 363380002, 404044006, 707398003, 707589002, 721560002, 722664004, 723844004, 724056005, 838349007, 352411000119109, 1081781000119105, 1259533008, 1259676000, 1259686004, 1263651005, 92176002, 92534003, 92680006, 93756008, 93882009, 94054005, 94532000, 94550007, 109976000, 118612006, 126498005, 126796005, 187810005, 187974007, 202430007, 278043000, 363400004, 363436001, 402531006, 403868009, 423973006, 722518004, 723889003, 733344004, 764856008, 348791000119109, 15930861000119100, 188558005, 123741000119102, 1081271000119100, 682471000119103, 686591000119102, 1259620008, 1259827009, 1237469002, 1264364008, 308006, 92045006, 92059004, 92445003, 92611000, 93720005, 93831006, 93884005, 93960003, 93962006, 94044006, 94497006, 94608002, 95147007, 118618005, 187637005, 188153009, 239147000, 254504001, 255132007, 285645000, 370987005, 403936002, 404116007, 704251001, 721622001, 762665002, 1671000119100, 91251000119105, 688111000119103, 1197297008, 16899901000119104, 1255646004, 1259415000, 1259728006, 1268500006, 1268881002, 92074002, 93851005, 93914000, 93990005, 95096004, 95103001, 126530003, 187760008, 188489006, 235966007, 254860001, 271323007, 402549006, 402557009, 418134006, 423050000, 423099001, 707404008, 723281005, 726653000, 736322001, 67771000119102, 16906841000119106, 878806005, 1081311000119100, 16068051000119106, 690121000119109, 208008, 93953009, 94276008, 94566009, 95045001, 95064007, 95067000, 95225003, 109284008, 109936005, 126509007, 187968007, 254446007, 254664008, 254702000, 277461004, 277567002, 315230006, 397008008, 403943008, 404004004, 449208008, 699028006, 707490009, 707672001, 721709000, 723856003, 133891000119102, 11996361000119102, 109388009, 188514005, 1081691000119102, 16228971000119104, 1162352003, 1259425005, 1259431008, 1236956002, 1237466009, 92269002, 92370004, 92374008, 92674003, 92715000, 93724001, 93860002, 93861003, 94317006, 94492000, 94960008, 126578003, 126603000, 254769006, 269464000, 307598005, 363358000, 363395000, 403919005, 432328008, 443439001, 447109003, 448139005, 722529000, 737313007, 830060005, 351871000119108, 92824003, 1080591000119105, 352911000119101, 763101411000119100, 1217489007, 1259377000, 1259739009, 1259815006, 62497000, 92066003, 93839008, 93955002, 94128004, 95101004, 126490003, 126620001, 187917009, 187970003, 188049009, 188665009, 201325006, 202576008, 254612002, 314974009, 372024009, 403856000, 419498005, 443937008, 707344002, 712858005, 737268008, 866109006, 91421000119103, 302191000119100, 354681000119109, 1078891000119104, 1081301000119103, 1153381008, 350771000119108, 681941000119107, 685601000119104, 793197981000119106, 1259598001, 1259651001, 1260093006, 93879004, 94045007, 94055006, 94596004, 95261008, 127258002, 188204000, 253018005, 255023008, 286890004, 363465007, 369569007, 369593002, 399660006, 403950007, 423896007, 426124006, 716655008, 33581000119109, 234941000119100, 242951000119108, 15664401000119101, 15709961000119104, 15937821000119109, 687941000119101, 688911000119101, 1197309002, 1259407003, 1259483001, 94066004, 94076001, 94909007, 126501005, 126609001, 188268004, 188326001, 188861002, 254619006, 254810007, 286889008, 310525009, 371974006, 403274000, 403711001, 706970001, 709285002, 721619003, 724059003, 1082101000119106, 126836001, 188548006, 404123008, 353211000119103, 1222581000, 1255883001, 1259309001, 1259328008, 1259354000, 1259765008, 1264369003, 91861009, 92348002, 92796000, 93649009, 93698000, 93864006, 94240007, 94490008, 94583000, 109266006, 109912006, 126745008, 254209005, 369456005, 402841001, 403713003, 413587002, 418529003, 448213004, 448663003, 700057001, 715344006, 866082003, 113211000119103, 432131000124100, 12241071000119106, 15950101000119108, 16260431000119104, 688901000119104, 1255662005, 1259559006, 1259709004, 1259781003, 1259828004, 1251496003, 92105004, 92110000, 92203003, 92372007, 93224002, 93924008,

94037002, 94707004, 126649009, 187957007, 188107002, 188728002, 202436001, 253038006, 254417000, 254788004, 314514005, 369570008, 403993007, 404092006, 707427000, 707475005, 707532001, 718200007, 833295000, 113081000119103, 136651000119104, 15632051000119106, 15931061000119103, 354721000119103, 689481000119107, 1255886009, 1259404005, 1259599009, 92277003, 92381001, 92769003, 93848003, 95003003, 95124006, 109854005, 109855006, 254403009, 255084004, 269476000, 278189009, 307622006, 400084008, 403970001, 404040002, 404090003, 404152006, 422886007, 423435000, 707455001, 707585008, 866097000, 348341000119102, 12235601000119105, 1079571000119109, 353761000119105, 687991000119109, 1259346002, 1259461000, 1268543003, 1284849005, 91993000, 92768006, 94080006, 94167005, 109932007, 126514006, 188998002, 369553000, 371997009, 372064008, 372125008, 403816002, 448257000, 448311004, 448562000, 448732009, 449207003, 716590006, 721305008, 724537009, 763796007, 12291000132102, 354741000119109, 92512000, 1010631005, 1153356005, 1156420003, 16260391000119109, 1259379002, 1259393009, 1259495001, 1259557008, 1260062005, 6729006, 92299006, 92565007, 93731002, 94796001, 95140009, 188019007, 188021002, 254389005, 254458007, 254749001, 363439008, 372092003, 387837005, 403468003, 403953009, 416753001, 448408001, 721630000, 866080006, 87091000119101, 91031000119108, 113091000119100, 122901000119109, 140051000119109, 1079291000119107, 1079341000119105, 1082321000119100, 254692003, 403947009, 890374007, 16905381000119100, 681681000119109, 808558991000119105, 1259504007, 1259832005, 1254748001, 92158009, 92720000, 93827000, 93887003, 94085001, 95171000, 110002002, 127254000, 188070003, 254454009, 254807000, 254950006, 274898000, 277474005, 307623001, 722714004, 107751000119102, 352191000119107, 1079561000119103, 1099581000119102, 352341000119101, 1196861001, 1196860000, 1259470002, 1259710009, 93732009, 93748005, 93802007, 93832004, 94226006, 94232001, 94585007, 94982001, 95210003, 109838007, 109948008, 126565003, 187769009, 187822008, 187939006, 187975008, 188068007, 234139002, 277624003, 277628000, 363485006, 370967009, 425327002, 449058008, 765331004, 353501000119104, 683431000119109, 1079741000119106, 352471000119101, 1255659007, 1259324005, 1259386005, 1259712001, 1259743008, 1259778008, 126866008, 187967002, 253020008, 363507003, 399897004, 403896003, 403944002, 404009009, 720487006, 721575005, 721725005, 735450006, 763063001, 763129001, 767813008, 84831000119102, 683221000119107, 253017000, 350621000119100, 1255815000, 1259487000, 1259544000, 1264495000, 1268635000, 92813000, 93835002, 94939002, 118615008, 126621002, 126742006, 127265005, 187723009, 187938003, 254709009, 275490009, 285432005, 307609003, 363389001, 372012002, 372129002, 402528005, 402554002, 402623006, 423189008, 423384009, 423600008, 447416008, 707425008, 707671008, 717916003, 721308005, 722229001, 838311008, 1131232100011910, 1156811000, 16414611000119100, 16845381000119100, 1162280003, 1259641002, 1259752004, 1260069001, 1236958001, 1254746002, 92743009, 93717002, 93728003, 94004004, 94269003, 95019005, 95083000, 126781006, 126998003, 187701005, 188184006, 188322004, 254747004, 254906005, 255153003, 310599006, 371970002, 371976008, 400173004, 403775003, 423700001, 721603005, 734066005, 763220008, 866071006, 866095008, 72891000119103, 130911000119108, 1081551000119106, 12238771000119102, 683721000119102, 1153382001, 681911000119108, 685971000119103, 1162284007, 1196899008, 1255752007, 1259335000, 1259413007, 91960009, 92732001, 94071006, 94078000, 94303002, 94428009, 95049007, 95093007, 109915008, 126561007, 126595006, 128516002, 187634003, 189062001, 253096008, 254649008, 254763007, 254806009, 276738009, 278052009, 302815008, 363379000, 371980003, 400026007, 404001007, 408649007, 423038006, 425148008, 447784001, 449699003, 698765007, 699818003, 707410008, 707456000, 707482009, 709203006, 721546004, 721634009, 722681008, 871802002, 350861000119106, 682701000119104, 11305881000119102, 277654008, 685861000119102, 1259311005, 1259571000, 1264257008, 92684002, 92707003, 93672006, 93910009, 94138009, 94254004, 95160001, 109824000, 127066000, 188955001, 285776004, 372093008, 449100005, 707586009, 721638007, 733135005, 122881000119107, 354851000119101, 352431000119104, 682221000119103, 685921000119104, 687401000119105, 1259635009, 1259757005, 1254724005, 1268542008, 76621000, 92738002, 93734005, 93799003, 93800004, 94300004, 94536002,

94726007, 95192000, 126701008, 127243005, 231832009, 274903001, 307650006, 402533009, 402536001, 404053004, 404148006, 415175000, 422399001, 424632001, 444597005, 707529004, 767448007, 788674000, 82501000119102, 101321000119102, 352121000119105, 1079131000119103, 895359007, 1145245004, 15955101000119101, 688711000119104, 1255194004, 1259323004, 1259385009, 1259464008, 92349005, 92480004, 92735004, 92797009, 93188005, 93643005, 93980002, 94644007, 94714002, 95412009, 109937001, 109982002, 126783009, 254427006, 254653005, 254997007, 255187008, 285604008, 285839005, 369457001, 369520005, 402496001, 402879006, 404006002, 723265000, 724091002, 733361001, 737309000, 761958009, 766981007, 788290007, 816202006, 838340006, 350951000119101, 15664881000119106, 92513005, 285420006, 1080631000119105, 16782651000119104, 1197350005, 1259331009, 1237055008, 92172000, 92223002, 92740007, 93696001, 94048009, 94375005, 94383004, 94546000, 94552004, 94568005, 94755004, 94843004, 94870006, 109357005, 126569009, 253000007, 254633006, 255035007, 372030009, 393573009, 402636006, 403931007, 403986008, 404142007, 424970000, 699251001, 707535004, 766935007, 782722002, 683261000119102, 404106004, 16091131000119108, 1251343008, 1259718002, 1236966005, 93676009, 93786001, 94021008, 94134006, 94439007, 94844005, 109841003, 109988003, 126799003, 254804007, 271481007, 280356005, 363392002, 722516000, 782827000, 91131000119109, 226521000119108, 354651000119102, 685811000119100, 15709881000119102, 16091251000119109, 1208459005, 1259319007, 1268885006, 88172005, 92010003, 92185002, 94096009, 94234000, 94547009, 95057005, 126776002, 190030009, 254459004, 254988008, 255016007, 255110003, 274905008, 276826005, 276828006, 310504009, 423807009, 428733008, 440397000, 721606002, 735757008, 737310005, 766930002, 450521000124109, 11996481000119103, 1172704005, 15951461000119105, 1259463002, 1259618005, 1259746000, 1259831003, 1254726007, 1268636004, 1268899005, 25050002, 93211002, 93738008, 93807001, 93824007, 94272005, 94504009, 94524008, 94712003, 95076007, 188157005, 254497004, 254656002, 254999005, 371999007, 372095001, 402519009, 404126000, 445266000, 449648005, 704250000, 32361000119104, 184871000119108, 683991000119103, 1081711000119104, 285422003, 10737361000119109, 1156413005, 1172629005, 1259432001, 1259555000, 1259764007, 1237516000, 1264496004, 92270001, 92382008, 93743001, 93764002, 93907002, 94101009, 94229004, 94533005, 95059008, 187946002, 254981002, 277642008, 312819009, 400063003, 403877002, 415177008, 448995000, 449472007, 707386008, 721695008, 722832009, 767818004, 133851000119107, 352251000119109, 1079351000119107, 1080171000119100, 10737641000119101, 1080611000119100, 688781000119105, 1259730008, 1259741005, 1259771002, 1259818008, 1268547002, 1268714009, 94451009, 94738003, 109941002, 126808006, 254916002, 399490008, 403952004, 404151004, 449257006, 702405001, 707345001, 707494000, 716647001, 723499000, 735920008, 1661000119106, 184881000119106, 11313361000119104, 188578002, 16906521000119103, 687811000119106, 1186629001, 1255797003, 1259573002, 1259629009, 1259833000, 1268458008, 82546001, 91857003, 92601007, 93641007, 94427004, 94747006, 94749009, 94908004, 95087004, 254390001, 254697009, 254767008, 254819008, 254979004, 276797002, 312113007, 314418005, 314515006, 402543007, 402839002, 403904009, 423425006, 449059000, 722112006, 860828007, 354201000119104, 1079381000119100, 109253006, 1010635001, 307771000119103, 688941000119102, 1196859005, 1255894002, 1259657002, 1237056009, 1268700009, 93680004, 93721009, 93844001, 94166001, 95075006, 109980005, 126495008, 126612003, 126626007, 126997008, 202586009, 254388002, 254419002, 254553001, 254723006, 254994000, 403825008, 403860002, 404134006, 415284008, 423318000, 447708000, 448670003, 702977001, 113131000119103, 302291000119108, 350891000119104, 394727000, 192701000119101, 878807001, 1080001000119106, 1156415003, 15950021000119100, 15957221000119107, 16905341000119105, 15951381000119109, 1259539007, 1260034003, 1268528000, 1268719004, 1269347008, 92217007, 92380000, 92413008, 93706003, 93739000, 93967000, 95061004, 109851002, 126786001, 134312002, 254492005, 254839007, 275492001, 402501000, 403865007, 404060005, 404107008, 415285009, 445227008, 721313009, 733343005, 735082004, 783006001, 865951006, 100731000119107, 1076971000119106, 12246601000119101, 93549004, 109391009, 276876007,

1079991000119101, 1081731000119109, 1179762006, 1197286008, 1208457007, 1255814001, 1255899007, 1259411009, 1259546003, 1259639003, 1259766009, 1268960003, 92012006, 92077009, 92387002, 92590009, 93136003, 93904009, 93976007, 109930004, 109943004, 126525002, 188266000, 188645002, 188863004, 254447003, 254800003, 277782009, 285606005, 285612000, 363401000, 371998004, 372110008, 403879004, 403882009, 403885006, 707537007, 707670009, 707697002, 716274007, 769169008, 354621000119105, 15951261000119109, 1187521002, 1197258000, 1254800007, 1259317009, 1259405006, 1269048009, 92069005, 92233005, 92525008, 93150008, 93829002, 93926005, 94440009, 94673005, 94775008, 109984001, 126750002, 188069004, 188264002, 202577004, 254661000, 255091001, 277653002, 281339009, 403867004, 404047004, 404664002, 425225007, 448145002, 448996004, 704254009, 712527004, 29421000119105, 1076931000119108, 1080601000119103, 1255902005, 1259339006, 1259398000, 1259549005, 1259784006, 92727002, 93773005, 94126000, 94227002, 94332004, 109788007, 109946007, 126868009, 126999006, 202439008, 254406001, 254754005, 254996003, 276827001, 303057009, 359780007, 369449004, 398768004, 400022009, 403864006, 403923002, 404073009, 699355007, 707401000, 713573006, 722684000, 737311009, 830023006, 840573001, 459421000124101, 188559002, 1010633008, 1156070008, 1157070004, 1157072007, 1196817009, 1259355004, 46659004, 92812005, 93669004, 94063007, 94202009, 94523002, 95027001, 95084006, 109918005, 202574006, 254711000, 255101006, 276750003, 278060005, 369459003, 400131007, 402534003, 404112009, 702369008, 722533007, 732201008, 860832001, 101601000119101, 15930901000119106, 15930941000119108, 354421000119107, 682271000119102, 686991000119106, 1254846007, 1259610003, 1259713006, 1259793007, 1259807008, 92188000, 93205000, 93779009, 93950007, 93992002, 95078008, 126523009, 126557001, 126749002, 127263003, 187644001, 187943005, 188945000, 231826004, 254725004, 369451000, 371963002, 371981004, 398623004, 402512000, 403204001, 403988009, 444911000, 698041000, 716586009, 722528008, 771235001, 134871000119100, 1079091000119100, 1079721000119100, 1081601000119108, 1082251000119103, 15956221000119105, 16845301000119108, 682421000119104, 685691000119105, 91221000119102, 1255897009, 1259526008, 1259631000, 1259646007, 1259824002, 94256002, 94636007, 95098003, 109384006, 126505001, 126628008, 127260000, 188676008, 188732008, 232075002, 239137003, 254989000, 274899008, 286893002, 371994002, 372104008, 400057007, 403996004, 422893006, 448258005, 733354000, 12235561000119105, 285421005, 686601000119109, 1259725009, 1236967001, 1237621006, 92442000, 93841009, 94282006, 94516003, 94642006, 109822001, 126756008, 188768003, 254394005, 277597005, 278055006, 363364007, 372122006, 403900000, 404077005, 413847001, 721701002, 866048009, 92516002, 313351009, 16908641000119109, 1148931000, 350801000119105, 688381000119106, 84821000119100, 1259430009, 1240372009, 1260467009, 1268531004

#### 3.30 Valvular Heart Disease

ICD10CM:
I09.89, I34.9, I05.1, T82.09XD, Q22.0, I06.8, I07.9, I34.81, A52.03, T82.03XD, I05.2, I37.9, I37.1, I08.1, I08.8, Q22.3, I34.1, Q22.8, I36.8, I34.2, I35.8, I06.9, I51.1, I05.8, I07.0, I37.2, I05.0, I07.2, I39, I08.2, Q22.4, T82.6XXA, I08.0, Q25.3, Q22.5, T82.6XXS, I06.0, Q24.8, T82.01XD, I23.5, I06.1, Q23.0, I07.8, Q22.9, Q22.2, I35.1, Q23.9, I06.2, T82.6XXD, Q21.3, I35.0, T82.02XD, T82.09XS, Q22.1, T82.01XA, I02.0, I36.1, I07.1, I51.2, I36.2, I23.4, I08.3, I34.89, T82.03XA, Q23.3, I38, Q23.2, Q23.4, T82.02XA, T82.03XS, I01.1, I34.8, I08.9, I36.9, I35.2, T82.02XS, I34.0, I37.8, Q22.6, I05.9, I37.0, I36.0, T82.09XA, T82.01XS, Q23.8, I09.1, I35.9, Q23.1

ICD9CM:
746.01, 424.99, 746.3, 746.5, 397.1, 429.5, 395.9, 396.0, 395.1, 395.0, 996.61, 396.3, 093.22, 429.6, 397.0, 424.3, 093.24, 394.0, 996.02, 394.2, 396.2, 746.2, 746.09, 746.02, 396.8, 745.2, 397.9, 746.4,

746.6, 746.1, 391.1, 395.2, 394.1, 746.89, 424.0, 425.3, 424.2, 429.81, 424.90, 093.21, 394.9, 093.20, 396.9, 396.1, 747.22, 093.23, 746.00, 424.1

SNOMED CT:
28284007, 29899005, 63042009, 194843003, 233819005, 233838001, 233846000, 250975000, 253399009, 253409001, 253527000, 373092008, 413439005, 449440009, 459182005, 871613001, 35411000087108, 43176009, 51789008, 128405007, 194989009, 233868005, 248686007, 447695000, 461429009, 763778003, 838333005, 1162507002, 1254666000, 49699002, 58056005, 61612001, 71799002, 250976004, 253374008, 253419007, 266238009, 447690005, 472765003, 703253007, 762541006, 80387009, 253284006, 253335001, 253352002, 253479002, 370141003, 405550001, 449479003, 703223000, 721014007, 871639003, 1260142000, 35501000087105, 13453009, 22653005, 33618002, 40521006, 94703000, 253438003, 472769009, 472793004, 768552007, 838535004, 10629351000119108, 32574007, 880011001, 890206005, 250973007, 253428008, 253523001, 300995000, 371814000, 442421004, 449429001, 715655000, 763280005, 880048005, 1187212004, 1255265004, 29889000, 94702005, 194809007, 447941008, 448612003, 472786003, 472834000, 703213009, 840609007, 15712921000119103, 880010000, 253447006, 447874007, 719678003, 54160000, 53741008, 194732001, 253586007, 447824005, 461112001, 471301006, 880040003, 1208976003, 488007, 251001002, 251006007, 253420001, 253440008, 427372006, 703211006, 762254000, 782699008, 783738002, 46696008, 63183009, 85081000, 253599005, 373131000, 472748007, 763835004, 840304005, 1187113001, 1255319004, 194991001, 194995005, 253371000, 253484008, 301184001, 448308000, 448684008, 472326004, 472767006, 720639008, 15960781000119108, 1149066008, 1254658009, 1254664002, 26117009, 61012002, 76534005, 123642008, 195025008, 195033009, 457727004, 472325000, 705069006, 723448007, 838449006, 871636005, 871659004, 421929001, 880050002, 1141882007, 6022005, 253393005, 253450009, 371809004, 373135009, 398716006, 447283008, 448357002, 448809003, 448982005, 449133000, 459061001, 725909004, 460620006, 16891191000119108, 253531006, 408546009, 447665008, 449031001, 449190000, 461435009, 472749004, 472848000, 764955006, 422348008, 879955009, 41334000, 51442005, 63553008, 253342001, 253357008, 253542002, 398995000, 428163005, 461414007, 471279005, 703175001, 716661006, 721976003, 735579001, 31080005, 253496001, 36233006, 83799000, 83978005, 85284003, 253294001, 735768008, 766959002, 871603009, 7991000119102, 1755008, 54329005, 55087008, 69589006, 253411005, 253650001, 405554005, 448083003, 714180007, 732230001, 15960541000119107, 63247009, 420938005, 139971000119106, 39693003, 195000004, 204379001, 253280002, 447915002, 449529003, 449538001, 472806007, 473455001, 771513008, 871622000, 1204222000, 1255258007, 54073003, 64420008, 93271008, 109426009, 111284004, 194997002, 253425006, 301106001, 311831009, 445898001, 448001004, 448280008, 723863003, 204319006, 1187540008, 92087008, 204318003, 253458002, 264162009, 449140004, 449352003, 457747007, 457767003, 41371000119100, 15960021000119107, 880000009, 10818008, 37104009, 111286002, 195005009, 210052005, 253370004, 373094009, 448844001, 449587004, 460913001, 461331004, 471288001, 472783006, 472842004, 721013001, 15960061000119102, 1204155000, 44841000087109, 1954004, 92976003, 194857001, 253356004, 445310002, 448182006, 448920004, 702424003, 703168002, 703356002, 15963181000119104, 1003919001, 36133000, 72011007, 74034002, 89141000, 91604001, 92969008, 253287004, 448476001, 457587000, 457722005, 703292000, 762539005, 871666003, 15712841000119100, 10273003, 64043005, 70422006, 253281003, 253295000, 253541009, 373138006, 447933008, 449315003, 457672001, 461424004, 703297006, 715535009, 771509001, 233834004, 414795007, 446628002, 447845003, 449596004, 457612009, 459068007, 461431000, 766976003, 1208872002, 16567006, 54830008, 56786000, 62185008, 194727002, 253412003, 253573003, 253647004, 373093003, 427919004, 715403006, 771478008, 838450006, 871579003, 1204468000, 35401000087106, 34126003, 65340007, 233865008, 253408009, 253491006, 253493009, 253602000, 286954005, 426087004, 432992007, 445270008, 445539001, 460358000, 720612000, 780842009, 840469002, 451241000124108, 1268523009, 46165000, 75270000, 123656005, 250993004, 371804009, 450301003,

703185000, 1204127004, 27097002, 84733009, 126732009, 233825009, 253584005, 266250003, 443379009, 448081001, 448082008, 461104009, 770435005, 836480008, 871654009, 871672003, 455941000124104, 92083007, 94327000, 204354004, 213036006, 371803003, 405547004, 448634001, 449563001, 457757008, 735569003, 756003, 52048004, 63462008, 195137008, 204368006, 234180009, 250984000, 253283000, 253364005, 253373002, 253506005, 253528005, 253545000, 253717008, 401314000, 444851003, 447672009, 448184007, 459057007, 461382001, 718181001, 719395001, 763066009, 1197430005, 44051003, 48724000, 82458004, 89792004, 111285003, 253301004, 457682000, 472100003, 718681002, 720606005, 763279007, 868214006, 871615008, 63191000119107, 1204151009, 1230407004, 38230003, 40272001, 48872007, 128558004, 251038002, 253289001, 253336000, 253392000, 253464009, 253724009, 460510005, 460524009, 472801002, 473444001, 838537007, 55836009, 87648004, 123659003, 253299006, 373137001, 447875008, 457777001, 472807003, 836295007, 868222004, 234182001, 250983006, 253474007, 253537005, 413905004, 446909006, 449512001, 472853005, 703224006, 1255624007, 16573007, 42770003, 58911001, 194802003, 253719006, 401303003, 449014008, 472795006, 472844003, 17531000119105, 286001000119104, 1279844009, 12023003, 64072006, 73774007, 194944008, 250974001, 253462008, 253543007, 253711009, 264467005, 373136005, 445336009, 446633003, 448413002, 457662004, 471276003, 871668002, 50608004, 64627002, 73028002, 233844002, 234181008, 266249003, 472772002, 703196008, 896689003, 8209004, 81990004, 195002007, 253549006, 253552003, 302943003, 449110001, 449459006, 457742001, 840507004, 204311009, 880049002, 86100009, 128555001, 194867006, 253439006, 253608001, 274098004, 373095005, 448794008, 699668009, 775908005, 840517009, 142951000119106, 1163075005, 44851000087107, 56276002, 95440004, 233867000, 253587003, 445928005, 448104005, 470754002, 722206009, 120871000119108, 890207001, 1204153007, 1237076003, 16063004, 75398000, 194958005, 253485009, 449189009, 4374004, 123643003, 195020003, 253333008, 253593006, 448887003, 449599006, 457762009, 868215007, 52035003, 64862009, 460600005, 703190002, 723858002, 6996004, 59062007, 79152002, 195021004, 233851006, 426191007, 448777006, 459175006, 472849008, 703191003, 718556007, 11433004, 19057007, 109430007, 123658006, 204300001, 248684005, 268174004, 413844008, 472707009, 703328007, 713318009, 720858001, 871624004, 72181000119109, 16754391000119100, 880062003, 75473000, 253279000, 253429000, 253488006, 253575005, 253723003, 399046008, 447825006, 472787007, 703322008, 286011000119101, 1155719000, 15990001, 194734000, 194921005, 204342004, 253410006, 253532004, 253721001, 447696004, 457637004, 703330009, 737155005, 838455001, 838541006, 870301007, 871601006, 15964981000119104, 58629009, 76267008, 194926000, 251007003, 447938004, 449595000, 871604003, 1163259003, 44801000087106, 40445007, 89736004, 194948006, 253297008, 253486005, 253534003, 448305002, 448919005, 461428001, 713511002, 718713000, 767169001, 681061000124108, 282006, 39589002, 194947001, 234186003, 253604004, 447779007, 448099009, 448819009, 773139006, 838542004, 40964007, 68602002, 93237004, 251036003, 253304007, 253544001, 262941008, 266252006, 413995009, 449158005, 449520004, 457712004, 703188003, 14886009, 56819008, 78031003, 204378009, 253344000, 253402005, 871658007, 896697005, 2583009, 4082005, 93262004, 128563000, 253277003, 253381001, 253456003, 253567004, 253592001, 448064005, 471292008, 471293003, 726335002, 876858006, 11104006, 67696008, 88129004, 253512000, 253577002, 253603005, 253730009, 447774002, 448074008, 448744007, 449136008, 457707000, 473372009, 703169005, 762433009, 35551000087106, 1268734005, 36222008, 253403000, 253471004, 449544002, 461432007, 57373003, 67754003, 89323001, 253376005, 263944006, 448102009, 448475002, 448828005, 473360003, 698971006, 703189006, 719955006, 720609003, 721073008, 838358000, 1142308000, 13689005, 32615007, 59063002, 61959006, 95268002, 253278008, 253611000, 447998001, 457782008, 472831008, 473376007, 718216009, 766883006, 15712961000119108, 1204462004, 61048000, 78495000, 186877007, 194983005, 233849007, 253351009, 398274000, 446326008, 448113007, 448528000, 448836001, 449178002, 460581004, 460352004, 15713081000119108, 880047000, 890220003, 44781000087105, 92944008, 204330009, 250985004, 253407004, 253582009, 429710001, 447261009, 716773002, 15712881000119105,

1254662003, 45093008, 194978002, 309518001, 399617002, 459172009, 204357006, 890120009, 12563008, 43873004, 53043001, 109432004, 204448004, 233830008, 253487001, 445334007, 448827000, 472316006, 703293005, 722027009, 838452003, 5230009, 16855001, 26201005, 70211005, 88445007, 194992008, 253710005, 445294008, 447670001, 449532000, 459159008, 703360004, 840680009, 8239009, 234187007, 253470003, 315025001, 472703008, 472847005, 871602004, 35461000087105, 248679004, 253406008, 447286000, 448820003, 457702006, 712866001, 286291000119103, 2829000, 20529002, 30496006, 42531007, 46701001, 52029003, 77863005, 78320000, 204345002, 253350005, 253452001, 253610004, 413994008, 448072007, 448117008, 448604006, 448903003, 472800001, 788533006, 840679006, 871600007, 55827005, 204397009, 233863001, 253437008, 253461001, 447847006, 448155003, 471277007, 719380003, 723088005, 1153452008, 5919001, 64077000, 72352009, 81552002, 253396002, 302131003, 447830005, 472845002, 719839000, 37217002, 91357005, 253303001, 253404006, 253418004, 253511007, 373134008, 448100001, 448624005, 449005003, 457687006, 471885006, 473443007, 737156006, 871599009, 90539001, 237227006, 253353007, 253378006, 461384000, 838451005, 28248000, 31993003, 109429002, 204312002, 250988002, 253434001, 431261000, 447284002, 459181003, 724431008, 838356001, 840305006, 86299006, 35928006, 195142000, 234132006, 427443009, 447988007, 449539009, 471287006, 735585008, 785847000, 50270004, 3098007, 21430002, 55546004, 253489003, 253588008, 449382009, 762542004, 67741000119109, 286041000119102, 15096009, 21379009, 28574005, 87343002, 92963009, 253703004, 287272002, 429673002, 431238002, 447698003, 448842002, 715626008, 788860005, 868217004, 22462001, 66893008, 248680001, 253361002, 371805005, 446813000, 447919008, 448026008, 460583001, 473359008, 762251008, 1162915007, 74249003, 109425008, 111027004, 187041005, 194929007, 204398004, 233858000, 253349005, 253380000, 253708008, 266237004, 282729004, 447682005, 448181004, 449568005, 459055004, 461305001, 471278002, 1254661005, 17718000, 21981000, 35162007, 37639005, 186875004, 194957000, 253366007, 448330004, 448747000, 448780007, 461416009, 703178004, 15962541000119106, 890121008, 368009, 18484008, 253362009, 253363004, 253460000, 253536001, 253580001, 311830005, 457607008, 7169009, 248682009, 250941001, 253285007, 253590009, 253625004, 264086008, 371817007, 448080000, 448119006, 448328001, 473379000, 703210007, 721978002, 723089002, 733491005, 762252001, 880005004, 1255256006, 35481000087104, 39502009, 253291009, 253540005, 448356006, 460601009, 461345005, 472830009, 703319006, 838316003, 35431000087103, 681051000124106, 8676001, 48520006, 77696009, 123641001, 194949003, 204306007, 253436004, 253492004, 275282007, 370513009, 431189009, 445299003, 447681003, 448823001, 457647001, 461385004, 461427006, 471286002, 703259006, 703386009, 720989003, 781159007, 871627006, 9485100011907, 285961000119107, 35521000087104, 94278009, 94865004, 253324006, 253732001, 371806006, 448946000, 449436000, 449457008, 871616009, 12238151000119107, 194865003, 233854003, 253359006, 253482007, 253574009, 448643005, 449564007, 472785004, 868220007, 8186001, 68583001, 86856005, 233861004, 253483002, 405560005, 609389009, 697976003, 703359009, 720748007, 723304001, 15960341000119104, 1222708006, 18546004, 93778001, 194945009, 233843008, 253270001, 253305008, 371816003, 426856002, 457642007, 461383006, 783096008, 836481007, 35451000087107, 62703006, 194858006, 194987006, 236530006, 286950001, 449149003, 871606001, 139011000119104, 6210001, 94822000, 233821000, 253522006, 253581002, 444718001, 448277007, 699255005, 866118008, 120901000119108, 460618008, 1229876001, 8712002, 31085000, 39131004, 93076005, 253445003, 253530007, 270906004, 448066007, 457752002, 459056003, 460923005, 472782001, 703171005, 35541000087108, 84338002, 93234006, 248687003, 253473001, 253509003, 409712001, 445298006, 448633007, 460588005, 703252002, 784353002, 117051000119103, 890122001, 35421000087100, 8074002, 60514000, 84871007, 163093004, 405548009, 703159004, 871611004, 19830001000004106, 896696001, 248675005, 250989005, 253455004, 253596003, 425177009, 445284003, 447666009, 448682007, 459060000, 766881008, 840283003, 871638006, 1208867006, 1234750000, 1237512003, 250997003, 253718003, 447829000, 472841006, 735578009, 788887009, 827078006, 840608004, 871609008, 35491000087102, 93782004, 126733004, 233871002, 253508006,

405752007, 449428009, 703288007, 736978009, 460317007, 896690007, 30288003, 44241007, 253700001, 446659000, 449379004, 472747002, 472829004, 472850008, 720567008, 762253006, 421272004, 880041004, 23063005, 41049003, 85971001, 194956009, 253268005, 253375009, 275516004, 371815004, 459178008, 717812000, 840488009, 9909003, 34342000, 45227007, 83521008, 92974000, 248685006, 253442000, 301122000, 429245005, 448625006, 449228007, 703290008, 715865008, 871650000, 253274005, 309519009, 447913009, 448822006, 449523002, 459153009, 721010003, 723333000, 783011004, 840315000, 380001000004106, 16891151000119103, 15713001000119100, 19036004, 109431006, 204363002, 204395001, 253282005, 253414002, 253478005, 253500004, 445235006, 448159009, 448793002, 461312005, 471280008, 762538002, 782698000, 286021000119108, 1119300000, 876859003, 880054006, 1148765006, 11851006, 40532006, 253339007, 253397006, 268180007, 424045003, 449040002, 449492000, 459067002, 459161004, 19092004, 13213009, 49915006, 447285001, 449011000, 449086000, 840312002, 431921000124105, 204351007, 880051003, 30277009, 253507001, 276795005, 301110003, 301185000, 360481003, 405551002, 446670007, 447683000, 447943006, 449353008, 459174005, 461438006, 472799000, 723865005, 762610001, 60573004, 880014009, 1204130006, 67682002, 109427000, 194726006, 194750008, 253338004, 253347007, 459177003, 472828007, 719835006, 724066002, 846667001, 1222710008, 92808004, 205834002, 253448001, 253579004, 446786009, 52757001, 57803009, 93558006, 253379003, 457617003, 459162006, 868225002, 253273004, 253465005, 297285002, 447289007, 448103004, 460582006, 703289004, 15629641000119107, 279001, 27637000, 42069006, 93825008, 253430005, 253594000, 445540004, 449098005, 449354002, 449398002, 450302005, 471299004, 472855003, 703172003, 285821000119105, 2816000, 4851007, 26900001, 250994005, 253293007, 253712002, 276517000, 457677007, 703212004, 46109009, 187051006, 442224005, 447962009, 473373004, 703214003, 836293000, 1269257006, 204394002, 233831007, 253571001, 405553004, 448497000, 449037002, 459154003, 703287002, 460329003, 1141742009, 1254663008, 92238001, 111319002, 194821006, 233852004, 248678007, 253476009, 253564006, 399228007, 447669002, 449015009, 457622003, 735586009, 840309000, 871585005, 871623005, 460338001, 33700007, 82355002, 88318005, 128568009, 233842003, 253417009, 253728007, 444855007, 448085005, 461422000, 471298007, 472327008, 703291007, 186878002, 253569001, 373905003, 417094009, 443502000, 447838003, 449029005, 472833006, 714181006, 60732002, 60812006, 195012000, 233855002, 236529001, 253624000, 370477004, 449456004, 471297002, 473393007, 762609006, 75372006, 94150003, 194950003, 195003002, 253501000, 447780005, 449135007, 473458004, 703318003, 840313007, 285981000119103, 880009005, 1148763004, 62827000, 65457005, 253519009, 371807002, 449123008, 880002001, 36135007, 195126007, 253323000, 304914007, 399020009, 457657000, 461110009, 472797003, 871607005, 67391006, 72527006, 194733006, 253516002, 263961009, 276518005, 363435002, 447839006, 871621007, 1254665001, 7368005, 18687009, 233836002, 250995006, 253459005, 425548001, 448063004, 448208003, 449316002, 725416005, 53322007, 1208977007, 195147006, 205768003, 253576006, 473380002, 719379001, 722461004, 35471000087101, 35511000087107, 44791000087107, 90154003, 194990000, 233832000, 233853009, 253423004, 253518001, 363437005, 448576004, 448729006, 16528621000119102, 41884003, 79619009, 91434003, 195141007, 204362007, 253555001, 444961009, 448414008, 457582006, 459180002, 838546001, 15960661000119107, 1141647009, 2024009, 59554006, 65547006, 92830003, 93069000, 128599005, 194868001, 448120000, 448745008, 449490008, 471890009, 703170006, 762520000, 233869002, 253498000, 253704005, 445268004, 445373007, 448098001, 871649000, 1187149000, 204399007, 253553008, 253606002, 35441000087109, 44630002, 86995005, 271984008, 446781004, 449351005, 449537006, 459158000, 461420008, 472802009, 10971000087107, 1141646000, 1208857001, 66595008, 86466006, 123660008, 194828000, 253386006, 253467002, 274097009, 703194006, 15960381000119109, 1163089000, 35561000087109, 3759007, 30328003, 35577008, 126731002, 253475008, 253524007, 445131007, 447999009, 457652006, 472318007, 846668006, 874782009, 10809641000119100, 876856005, 11176009, 21470009, 60232001, 84648007, 195004008, 253394004, 445330003, 460437005, 718135001, 871620008, 1208998007, 73795002, 78046005, 234189005, 253416000, 253499008, 253729004,

473374005, 473375006, 473383000, 721009008, 871610003, 10743121000119108, 15960741000119103, 44761000087104, 233828006, 253431009, 447852001, 448062009, 448575000, 457692008, 726704006, 871618005, 16837681000119104, 60234000, 253538000, 445440001, 448975006, 449111002, 461439003, 762540007, 285151000119108, 253443005, 880012008, 1237074000, 43121002, 45058001, 186318004, 233817007, 262782002, 370551009, 459183000, 838538002, 1089931000119105, 1255264000, 10365005, 63739005, 253547008, 253585006, 457737009, 703187008, 703192005, 723993005, 735580003, 735771000, 91025000, 253441007, 420006002, 443414005, 448619007, 461111008, 792842004, 8722008, 27277001, 41514002, 58612006, 74918002, 234184000, 253533009, 253559007, 253722008, 445454009, 447932003, 448075009, 449032008, 471290000, 868221006, 871665004, 15629741000119102, 29375001, 36471000, 251003004, 253562005, 270510008, 297197008, 461105005, 471285003, 703119002, 703209002, 717859007, 721015008, 726083008, 868226001, 871583003, 459761000124100, 1596098100019105, 876846006, 1264404009, 44821000087100, 37925008, 61490001, 86348002, 194735004, 253383003, 253422009, 373096006, 398754006, 445453003, 470752003, 715987000, 876857001, 880013003, 1187147003, 94433008, 194823009, 297160003, 447671002, 447937009, 449455000, 457592003, 719824001, 764697003, 838543009, 840497008, 880044007, 35581000087103, 17759006, 111287006, 233826005, 253269002, 253271002, 253566008, 387842002, 445002005, 460944005, 471841009, 472324001, 718128009, 719400000, 720605009, 735571003, 880043001, 1255187002, 2439002, 19249002, 253446002, 253510008, 264571006, 457772007, 473377003, 716193004, 868223009, 880007007, 890393009, 1204152002, 64190005, 195136004, 253505009, 253535002, 253539008, 253554002, 315348000, 427515002, 446916007, 450300002, 461107002, 94706008, 123801008, 429347004, 445003000, 447667000, 448790004, 708121009, 35719004, 55482007, 233835003, 233860003, 253600008, 301105002, 449385006, 703296002, 838536003, 871581001, 1186718008, 1208897003, 1254657004, 50920009, 71932004, 194984004, 233837006, 253345004, 253481000, 360473004, 470751005, 473362006, 703179007, 703327002, 35571000087100, 15459006, 76257003, 311793000, 311796008, 448723007, 460538002, 460599007, 461298006, 461413001, 703173008, 735565009, 78741000119103, 16440002, 89456004, 371811008, 414545008, 447661004, 447673004, 448331000, 703323003, 838454002, 286391000119108, 890119003, 1255189004, 57054005, 70998009, 94818005, 253433007, 253435000, 253607006, 449270002, 472813007, 871625003, 871645006, 871652008, 1230303001, 210078001, 251000001, 253341008, 253715000, 449087009, 457717005, 461352007, 723336008, 863954001, 47292005, 74218008, 233866009, 253326008, 253348002, 253502007, 274947007, 445285002, 724173009, 770432008, 871597006, 871660009, 880037003, 194946005, 233856001, 253288009, 445636003, 448415009, 449112009, 449441008, 703222005, 703321001, 871594004, 880045008, 1142043006, 1255621004, 55510008, 194943002, 204346001, 253354001, 431262007, 445001003, 445607003, 448061002, 448310003, 460937001, 461417000, 703181009, 868224003, 871612006, 15713161000119100, 42402006, 253514004, 1119298000, 1197364005, 59464004, 92923008, 128567004, 250982001, 253427003, 703251009, 836294006, 8916007, 41964001, 92960007, 194863005, 233840006, 250942008, 250999000, 253463003, 253472006, 253503002, 412787009, 446630000, 460531008, 460602002, 471275004, 472851007, 838545002, 846684007, 871671005, 12238111000119106, 880046009, 1208873007, 1131009, 1255623001, 28880005, 233847009, 253302006, 253369000, 253504008, 253521004, 253563000, 253570000, 417081007, 445650008, 446699002, 449600009, 459059005, 763834000, 27007008, 78250005, 253343006, 253444004, 253449009, 297286001, 448116004, 460930004, 470753008, 713405002, 762255004, 726499301000119105, 890204008, 890402009, 787001, 79096004, 88782004, 233827001, 251024009, 253595004, 253707003, 457732003, 703320000, 703324009, 451041000124103, 12367281000119109, 91529004, 233930009, 233970002, 250979006, 253497005, 405552009, 449124002, 1255188007, 56309007, 109428005, 194709000, 194842008, 204391005, 204402008, 253300003, 371810009, 443762000, 449442001, 460614005, 719272007, 93078006, 253337009, 253591008, 448320008, 458039003, 460585008, 871587002, 871605002, 111000119104, 15713201000119105, 880003006, 194931003, 276790000, 449008001, 449383004, 449567000, 460438000, 786040006, 286031000119106, 1255257002, 681041000124109,

75403004, 76593002, 92132009, 253413008, 253648009, 363436001, 448278002, 472796007, 703167007, 703255000, 721841001, 840495000, 4557003, 19833008, 59494005, 83898004, 194741006, 251004005, 253642005, 371812001, 448183001, 460586009, 871630004, 1204154001, 67278007, 93914000, 253391007, 253556000, 278480000, 308687000, 447689001, 448007000, 700065003, 719456001, 723862008, 35304003, 60106004, 62067003, 78485007, 89222008, 109379000, 204296002, 233823002, 233841005, 253275006, 445286001, 448618004, 448786001, 763747002, 871647003, 15960141000119102, 18192007, 65718001, 123657001, 445264002, 471291001, 716740009, 44811000087108, 34845009, 72252007, 79009004, 204315000, 233873004, 234185004, 253388007, 253453006, 253601007, 430402003, 448923002, 448947009, 449186002, 459151006, 472854004, 703165004, 15960581000119102, 15713121000119105, 1144420008, 12770006, 25106000, 29934004, 40023000, 210064002, 253398001, 253454000, 428196007, 446358003, 446667008, 448743001, 459152004, 703225007, 722051004, 1204194004, 48121000, 94954008, 210060006, 233859008, 448477005, 460490009, 473454002, 703157002, 703200003, 880038008, 1149109001, 1255266003, 72972005, 253328009, 253395003, 253550006, 253551005, 253706007, 301183007, 459176007, 838544003, 1254660006, 48228003, 73699003, 253267000, 253415001, 253494003, 444856008, 462174007, 735769000, 770433003, 788529004, 285141000119106, 22298006, 36665001, 194845005, 204352000, 251002009, 253286008, 253426007, 448004007, 461109004, 472852000, 719451006, 773749003, 73660006, 195032004, 204339005, 253583004, 311792005, 429559004, 449534004, 735566005, 838448003, 108411000119109, 15964781000119101, 93445007, 210051003, 233829003, 234188002, 253546004, 253716004, 371808007, 448162007, 448871001, 471289009, 866080006, 871646007, 871648008, 253515003, 28869005, 59021001, 70320004, 253495002, 253720000, 413838009, 430616003, 447697008, 448309008, 459058002, 459062008, 703325005, 703358001, 762460002, 838539005, 12367321000119104, 1155721005, 1204210003, 4313003, 36079008, 64749001, 86175003, 93247001, 250978003, 250987007, 253382008, 418044006, 445435009, 460471001, 703160009, 880006003, 253327004, 253568009, 448073002, 449593007, 461359003, 461436005, 470750006, 721575005, 735581004, 1240531000000103, 1254659001, 1255569006, 17414004, 163092009, 204343009, 253360001, 373116009, 446657003, 460587000, 703164000, 703326006, 1119299008, 1234910007, 70142008, 78196008, 93075009, 128557009, 204317008, 233839009, 253390008, 253400002, 253405007, 430065009, 448778001, 449511008, 449555002, 449601008, 460584007, 472798008, 723864009, 846683001, 44771000087108, 20721001, 86194008, 204370002, 233850007, 253490007, 314116003, 394659003, 394710008, 449118008, 449139001, 461425003, 735570002, 26146002, 896691006, 91468009, 253298003, 253612007, 301109008, 458038006, 459066006, 460517008, 472846001, 704241002, 868216008, 1177176009, 1255622006, 129574000, 250996007, 253421002, 253529002, 253714001, 276516009, 448095003, 448516008, 448905005, 838453008, 28656008, 41893002, 91442002, 194942007, 253358003, 406428006, 448746009, 449605004, 705070007, 1137687006, 399216004, 448028009, 448821004, 449138009, 449458003, 457697002, 462171004, 717943008, 838540007, 871655005, 108421000119102, 15713041000119103, 880004000, 44831000087103, 16253001, 82608003, 95890006, 187195003, 253377001, 253451008, 253477000, 253598002, 307140009, 315615007, 371813006, 448115000, 457632005, 771075004, 783773000, 838456000, 91335003, 94720001, 248688008, 250990001, 253401003, 253565007, 428164004, 461108007, 725145002, 764452004, 791000119109, 880052005, 233862006, 233864007, 253346003, 253432002, 448843007, 459065005, 720610008, 31701000119107, 1186709006, 194866002, 253290005, 253469004, 253609009, 253725005, 264503005, 286947004, 314207007, 448724001, 461381008, 703186004, 880042006, 7484005, 194856005, 233929004, 251061000, 268185002, 445512009, 448626007, 448966009, 472820000, 472832001, 871643004, 451361000124102, 1003863006, 28931004, 52987001, 92517006, 93053007, 253641003, 278928000, 449116007, 703177009, 840310005, 17394001, 83119008, 128556000, 248691008, 253384009, 253385005, 447822009, 448478000, 703236000, 840316004, 23311000119105, 1052287002, 45503006, 57181007, 82522008, 186755002, 253578007, 373133002, 448948004, 460545002, 461421007, 472835004, 725027004, 29928006, 54072008, 62695002, 234183006, 253276007, 253387002, 253480004, 253597007,

285606005, 360465008, 413933004, 421327009, 699257002, 868219001, 253468007, 880008002, 1163440003, 1268520007, 93287006, 420395004, 442240008, 460345001, 472843009, 836482000, 840303004, 53874003, 90202001, 91634006, 92888002, 233833005, 253340009, 253513005, 253525008, 448876006, 449016005, 15960461000119105, 44993000, 93051009, 253272009, 423718003, 447876009, 448118003, 449099002, 11018701000119109, 1230344000, 42099002, 253355000, 253520003, 413444003, 417191003, 461423005, 472840007, 762519006, 67751000119106, 16025471000119107, 1179293006, 50570003, 129582000, 194736003, 234010000, 253548003, 448574001, 449594001, 457667005, 871640001, 1204192000, 42866003, 92941000, 94282006, 253264007, 253389004, 253457007, 448161000, 448681000, 449388008, 472794005, 1141645001, 1204460007

#### 3.31 Weight Loss and Malnutrition

ICD10CM:
E45, R63.4, E40, E44.0, R64, E44, E43, E44.1, E46, E41, E42

ICD9CM:
799.4, 263.8, 263.0, 783.22, 260, 783.2, 263.1, 263.2, 263, 263.9, 783.21

SNOMED CT:
762493006, 273491005, 238107002, 422868009, 199741009, 238109004, 281583001, 13365009, 1260096003, 360554000, 238108007, 267024001, 717925009, 77702009, 762495004, 58262005, 788876001, 360549009, 762494000, 422003001, 74257000, 421695000, 190606006, 10812131000119108, 29740003, 419261006, 284529003, 448765001, 190603003, 77091003, 717926005, 717927001, 272588001, 717928006, 190602008, 430237002
